# Mechanism of response to FHD-286 and decitabine combination in patients with advanced myeloid malignancies

**DOI:** 10.64898/2026.07.17.26358055

**Authors:** Michael P. Collins, David L. Lahr, Salih Topal, Alexis P. Khalil, Denice Hickman, Nicholas Spidale, Nikhil Pandit, Sarah A. Reilly, Kelly Lyons, Kim Horrigan, Tina Zhao, Joelle Batonga, Mia Bosinger, Katie D’Aco, Brian Ball, Ashwin Kishtagari, Courtney D. DiNardo, Eytan M. Stein, Alfonso Quintás-Cardama, Gromoslaw A. Smolen

## Abstract

Impaired cellular differentiation is a defining characteristic of myeloid malignancies and remains a major therapeutic challenge. The BRG1/Brahma-associated factor (BAF) chromatin remodeling complex, through the ATPases SMARCA4 and SMARCA2, maintains the stemness of leukemic blasts and thus represents a promising target for novel differentiation-based therapies. In a phase 1 study in advanced myeloid malignancies, the first-in-class dual SMARCA4/2 inhibitor FHD-286 combined with decitabine (DAC) was tolerated and produced an objective response rate of 12.8% (6/47) compared with no responses with FHD-286 monotherapy. To understand the basis of this activity, we integrated high-dimensional flow cytometry and single-cell genomic analyses of longitudinal bone marrow samples from responders and nonresponders. While FHD-286 monotherapy was predominantly associated with myeloid differentiation, responders to FHD-286+DAC combination therapy exhibited a range of myeloid and erythroid differentiation trajectories. FHD-286 potentiated the transcriptional impact of DAC, driving tumor clones to fully differentiate out of the immunophenotypically and transcriptionally defined blast compartment. Responders had a baseline transcriptional profile similar to that of *CEBPA*-mutant acute myeloid leukemia and showed further downregulation of *CEBPA* upon treatment. These findings reinforce tumor cell differentiation as a mechanism of response to pharmacologic SMARCA4/2 inhibition and support further evaluation of FHD-286+DAC in molecularly defined patient subsets.

**ONE SENTENCE SUMMARY:** In a phase 1 R/R AML study, FHD-286 potentiated the effects of decitabine to promote responses via tumor cell differentiation and *CEBPA* perturbation.

## Introduction

Chromatin remodelers are crucial for the regulation of gene transcription, DNA replication, and DNA repair (*1, 2*). The BAF (BRG1/Brahma-associated factor; also known as mammalian SWI/SNF) complex is a large multisubunit ATP-dependent chromatin remodeler that repositions nucleosomes to enable active transcription. The BAF complex has frequently been implicated as a potential therapeutic target in cancer due to its high mutation rates across various tumor types and its established roles in the regulation of lineage genes essential to the dedifferentiated state of many cancers (*3*).

Each BAF complex contains one of 2 ATPase subunits: either SMARCA4 (also known as BRG1) or SMARCA2 (also known as BRM). BAF complexes govern the critical differentiation and proliferation processes that are altered in acute myeloid leukemia (AML) and have been shown to be important in maintaining leukemic progenitor cells. Specifically, the SMARCA4 subunit, expression of which is upregulated in AML, contributes to leukemia maintenance in part by enhancing expression of the *MYC* oncogene, resulting in clonal expansion of undifferentiated myeloid precursors and impaired hematopoiesis (*4, 5*). In vitro studies showed that inhibition of both SMARCA4 and SMARCA2 was required to modulate leukemic transcriptional programs and induce differentiation and/or apoptosis in AML cells (*6*).

Study FHD-286-C-002 was an open-label, nonrandomized, phase 1 dose escalation study of the dual SMARCA4/2 inhibitor FHD-286 in patients with relapsed or refractory (R/R) AML, myelodysplastic syndrome (MDS), or chronic myelomonocytic leukemia (CMML) (NCT04891757). In the first part of the study, FHD-286 was administered once daily (QD) as monotherapy (*7*). No objective responses were reported, although 6 of 18 (33.3%) evaluable patients experienced over 50% reduction in bone marrow (BM) blasts with evidence of myeloid differentiation, and differentiation syndrome (DS) was observed in a fraction of patients (*7*).

Based on nonclinical results demonstrating enhanced cell killing, tumor growth inhibition, and prolonged survival in murine models of AML treated with the combination of FHD-286 and decitabine (DAC) (*8, 9*), the second part of Study FHD-286-C-002 evaluated this doublet. The combination induced objective responses, and patient samples collected longitudinally from responders and nonresponders in both parts of the study were extensively characterized to understand the molecular mechanisms involved in the antileukemic activity. Using a variety of orthogonal methods, we show here the impact of FHD-286+DAC on cellular differentiation and demonstrate that responses to combination therapy were associated with myeloid and erythroid differentiation, in contrast to FHD-286 monotherapy, which predominantly resulted in myeloid differentiation. Furthermore, our integrative analyses highlight the role of *CEBPA*, a key transcription factor regulating erythroid and myeloid differentiation, and suggest that responses are clustered in AML patients with *CEBPA* functional perturbations. Our results suggest a path for further clinical development of FHD-286+DAC in molecularly defined patient subpopulations.

## Results

### Clinical study description

In the combination therapy portion of Study FHD-286-C-002, 47 patients were enrolled at 5 sites in the United States between August 2023 and November 2024. All patients were included in analyses of safety, pharmacokinetics (PK), and clinical activity (fig. S1). Patients who were, at baseline, not receiving a triazole antifungal agent classified as a strong cytochrome P450 (CYP)3A inhibitor (Group B1) received FHD-286 at dose levels of 2.5 mg, 5 mg, 7.5 mg, 7.5/5 mg (interval dosing, with doses alternating every 2 weeks), and 7.5/10 mg (interval dosing) QD (fig. S2). Patients who were, at baseline, receiving a triazole antifungal agent classified as a strong CYP3A inhibitor (Group B2) received FHD-286 at the following dose levels: 1.5 mg, 1.5/2.5 mg (interval dosing), and 2.5 mg QD. Decitabine was administered intravenously for the first 5 days of each 28-day cycle at a dose of 20 mg/m^2^.

The median duration of treatment was 30 days (range: 1, 170), was slightly higher in Group B1 than in Group B2, and tended to decrease as the dose level increased. The most common reasons for treatment discontinuation were an adverse event (AE) (17 [36.2%] patients), disease progression/treatment failure (12 [25.5%]), and withdrawal of consent or investigator decision (8 [17.0%] each).

### Patient and disease characteristics

Forty-two (89.4%) patients enrolled to the combination portion of the study had a diagnosis of R/R AML, 4 (8.5%) had R/R MDS, and 1 (2.1%) had R/R CMML (table S1). The 87 total patients in this study were representative of the broader population of patients with these advanced myeloid malignancies (table S2). Most of the 87 patients had adverse-risk disease (65 [74.7%]) according to the 2022 European LeukemiaNet genetic risk classification (*10*). Patients had been heavily pretreated for AML, MDS, or an antecedent hematologic disorder. Eighty-two of 87 (94.3%) patients had experienced treatment failure with a hypomethylating agent (HMA; azacitidine or DAC). Eighty-one of 87 (93.1%) had failed a venetoclax-based regimen, 56 (64.4%) had failed a cytarabine-based regimen, and 25 (28.7%) had progressed after a hematopoietic stem cell transplant (HSCT). There were no substantial differences between the patients in the monotherapy and combination therapy parts of the study in terms of age, risk stratification by genetics, hematologic parameters, transfusion dependence, and prior therapy (table S3). Among the 47 patients treated with FHD-286+DAC, most had an abnormal karyotype (39 [82.9%]) and adverse-risk disease (32 [68.1%]). Patients had failed a median of 3 prior lines of therapy (range: 1, 9). Forty-five (95.7%) patients had experienced treatment failure with an HMA, 45 (95.7%) had failed a venetoclax-based regimen, 32 (68.1%) had failed a cytarabine-based regimen, and 13 (27.7%) had undergone HSCT (table S4).

### Safety, tolerability, and impact of strong CYP3A inhibitors

The safety and tolerability profile of FHD-286+DAC, including signs and symptoms associated with DS, was qualitatively similar to that of FHD-286 monotherapy (tables S5 and S6) (*7*). In the combination group, DS was reported in 9 (19.1%) patients and retrospectively adjudicated by an independent safety monitoring committee (ISMC) as having occurred in 10 (21.3%) patients. Importantly, the PK profile of FHD-286 administered in combination with DAC was similar to that of FHD-286 administered as monotherapy (*7*). In the combination part of the study, FHD-286 exposure was approximately 3-fold higher in patients who were concomitantly receiving a triazole antifungal classified as a strong CYP3A inhibitor (Group B2) compared with those who were not (Group B1) (fig. S3). FHD-286+DAC was better tolerated among patients in Group B1 than in Group B2, with fewer treatment discontinuations and fewer dose-limiting toxicities (DLTs), which allowed for the testing of higher doses. Seven DLTs were observed in 4 patients at the following doses: Group B1 7.5/10 mg QD (Grade 3 increased alanine aminotransferase, aspartate aminotransferase, conjugated bilirubin, and blood bilirubin [n = 1]), Group B2 1.5/2.5 mg QD (Grade 3 conduction disorder [n = 1]), and Group B2 2.5 mg QD (Grade 3 hyperglycemia [n = 1] and Grade 3 increased blood bilirubin [n = 1]). The maximum tolerated doses (MTDs) for Groups B1 and B2 were determined to be 7.5 mg QD and 1.5 mg QD, respectively.

### Efficacy

The objective response rate, per modified International Working Group (IWG) criteria (*11, 12*), was 12.8% (6 of 47 patients). Among patients with R/R AML, 2 had a best response of complete remission with incomplete platelet recovery (CRp) (Group B1 2.5 mg and 7.5 mg QD) and 3 had a best response of morphologic leukemia-free state (MLFS) (Group B1 5 mg QD [n = 1] and Group B2 2.5 mg QD [n = 2]). One patient with R/R MDS had a best response of partial remission (PR) (Group B1 2.5 mg QD) (fig. S4). The median time to best response was 1.93 months (range: 0.46, 4.04) and the median duration of response was 1.99 months (95% CI: 1.28, not estimable).

### Decitabine dampens FHD-286-induced myeloid differentiation

In the monotherapy portion of the study, marked decreases in the relative abundance of CD34^+^ cells and increases in the relative abundance of CD11b^+^ cells were observed in BM blasts by flow cytometry, suggesting a myeloid differentiation mechanism of action (*7*). We further investigated these effects by incorporating data from the combination therapy portion of the study. In patients with matched screening and Cycle 2 Day 1 (C2D1) samples (monotherapy, n = 12; combination, n = 14), dose-dependent decreases in CD34^+^ cells and increases in CD11b^+^ cells were observed within the blast compartment; however, the consistency and magnitude of these changes were generally weaker in the combination cohort (Fig. 1A). Changes in CD34^+^ and CD11b^+^ cell populations, as well in the populations of cells marked by other stem and maturation markers (CD117 and CD64, respectively), were also more strongly correlated with FHD-286 plasma concentration among patients treated with monotherapy (Fig. 1B). Among the 4 evaluable patients with an objective response (all receiving combination therapy), effects on CD34^+^ and CD11b^+^ populations were mixed, with 2 patients showing strong changes and 2 patients showing minimal effects (Fig. 1C). Overall, these results demonstrate that FHD-286+DAC induced weaker myeloid differentiation compared with FHD-286 monotherapy and, importantly, suggest that additional cellular mechanisms may be responsible for the increased clinical activity observed with FHD-286+DAC.

**Fig. 1.**
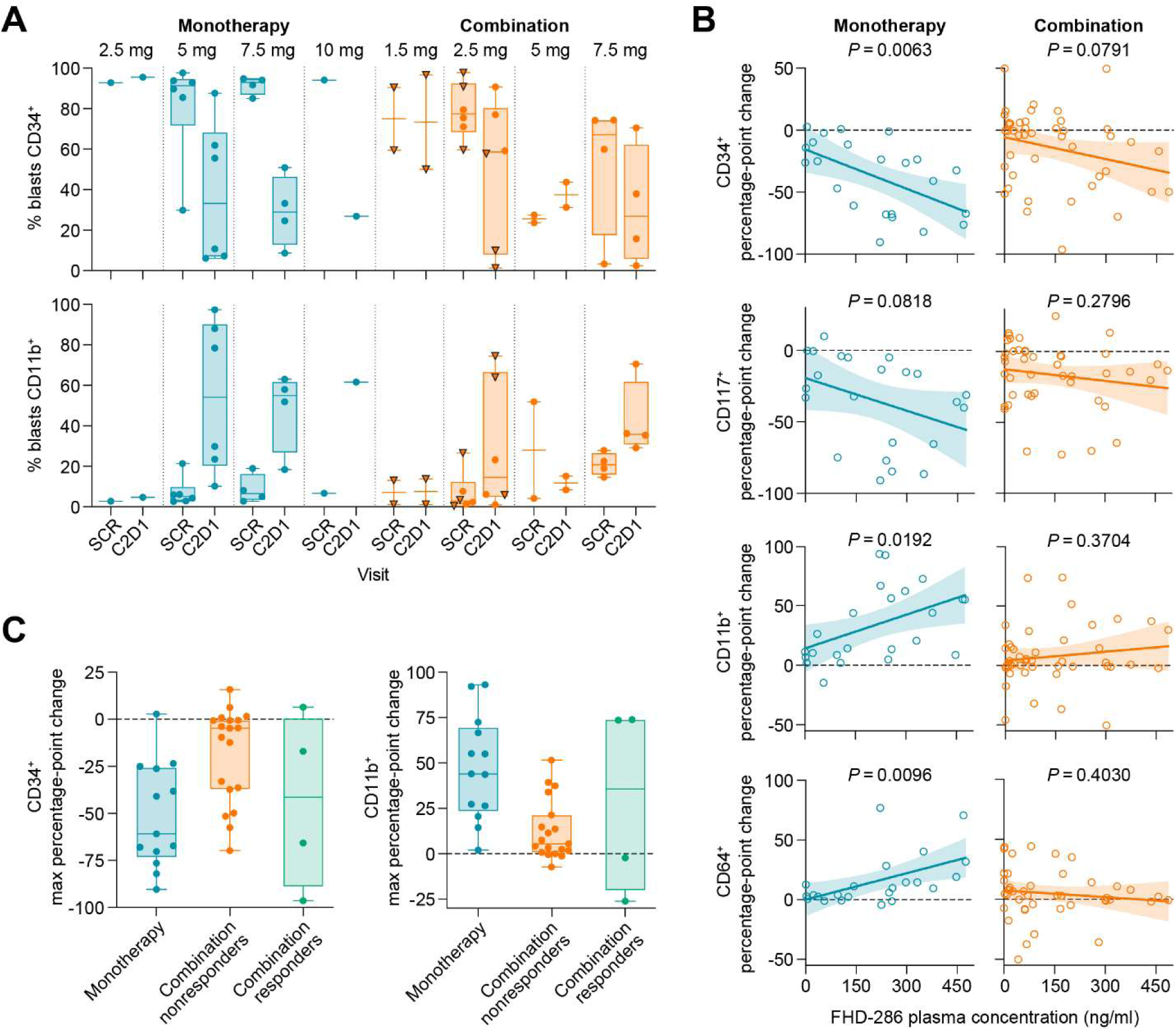
Myeloid differentiation was observed in blasts of patients treated with FHD-286. (**A**) Percentage of BM blasts positive for CD34 (top) or CD11b (bottom), at screening (SCR) and C2D1 (samples collected between study days 25 and 31) for all patients with matched SCR and on-treatment samples. In the combination groups, circles represent Group B1 samples and triangles represent Group B2 samples. Box plots represent median, interquartile range, and maximum/minimum values. (**B**) Percentage point change from SCR in BM blasts positive for indicated marker, plotted against FHD-286 plasma concentration at the corresponding visit. BMA samples obtained within 3 days of corresponding PK sample were included. Shaded areas represent 95% confidence interval of the best-fit line. (**C**) Maximum percentage point change from SCR in BM blasts positive for CD34 (left) or CD11b (right) for all patients with matched SCR and on-treatment (any time point) samples. Box plots represent median, interquartile range, and maximum/minimum values.

### FHD-286+DAC promotes expansion of the erythroid compartment

We next conducted a high-dimensional fluorescence-activated cell sorting (FACS) analysis of the BM aspirate (BMA) samples to more thoroughly characterize the immunophenotypic effects of FHD-286+DAC. This analysis was not solely restricted to the blast compartment and instead used multiparametric fluorescence data acquired for all viable BM mononuclear cells. We performed t-SNE (t-distributed stochastic neighbor embedding) dimensionality reduction and unsupervised clustering on 2,731,481 cells from 90 BMA samples from 36 patients (23 with paired screening and on-treatment samples; 13 with unpaired samples) and identified 11 clusters occupying distinct regions of the reduced dimensionality space. We annotated the cell identity of each cluster based on marker co-expression patterns (fig. S5A, B), and distinguished 3 blast, 2 myeloid, 2 erythroid, and 2 lymphoid clusters (Fig. 2A). The remaining 2 clusters, representing 0.7% of total cells, were not distinguishable based on the markers used, and were designated as unknown. To further validate the assigned cellular identities, we confirmed that cells from each cluster localized to the expected regions of conventional CD45/side scatter (SSC) plots (Fig. 2B).

**Fig. 2.**
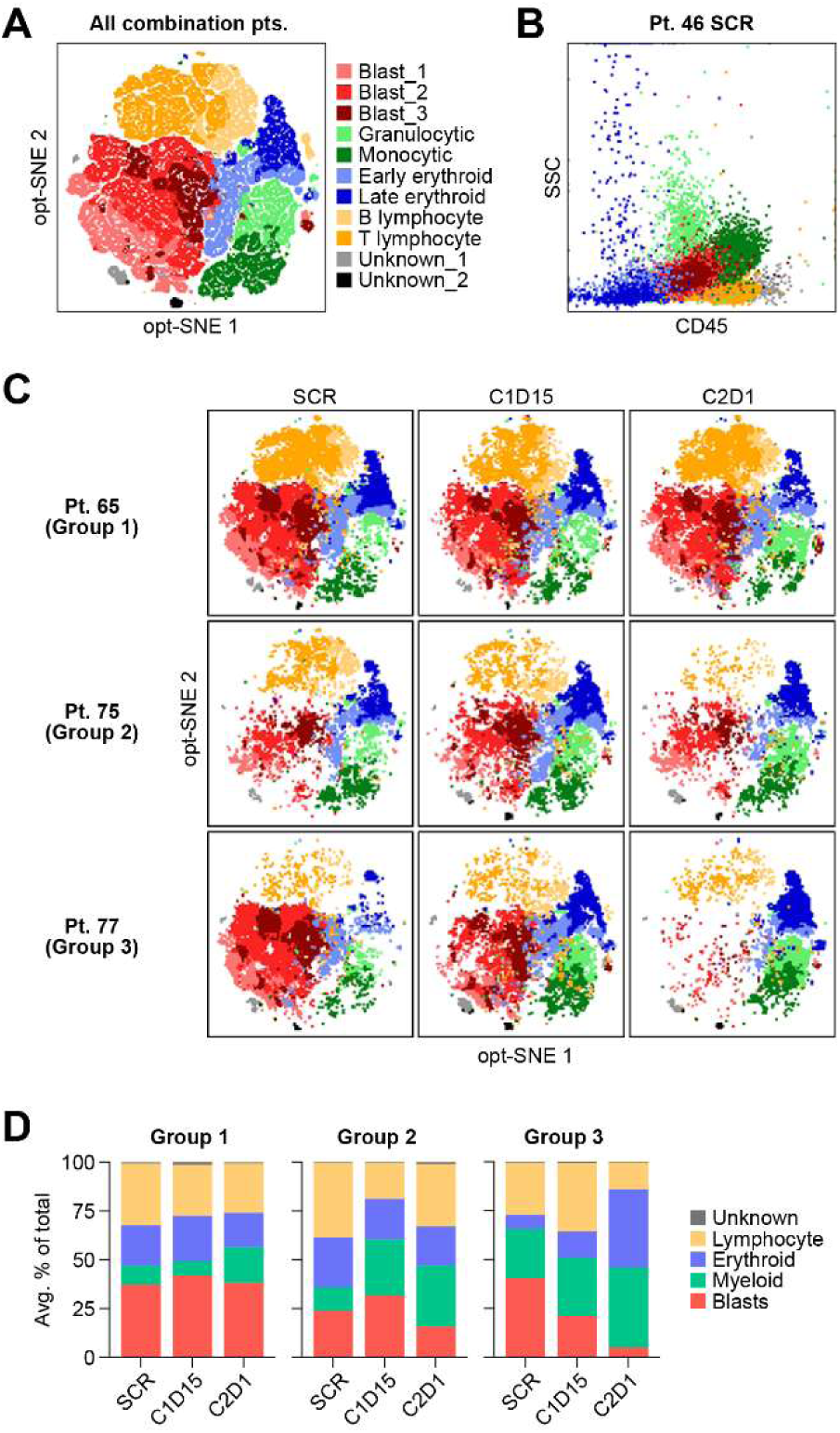
High-dimensional FACS analysis demonstrates increased erythroid populations in FHD-286+DAC responders. **(A)** Opt-SNE visualization of all cells from combination BMA samples, colored by cluster. **(B)** Mean fluorescence intensity of CD45 versus SSC for all cells from a representative sample, colored by cluster as in (A). **(C)** Individual opt-SNE plots at screening (SCR), C1D15, and C2D1, colored by cluster as in (A). Representative patients from Group 1 (nonresponders, <50% blast reduction), Group 2 (nonresponders, ≥50% blast reduction), and Group 3 (objective responders) are shown. **(D)** Mean compartment abundance for all samples at SCR, C1D15, and C2D1, stratified by group as described in (C). Compartments represent sums of similar clusters shown in (A) (e.g., myeloid = sum of granulocytic and monocytic clusters). Avg, average; Pt, patient.

We next investigated changes to the relative percent abundance of the identified cell compartments in response to FHD-286+DAC. The analysis population was subdivided by disease response: <50% blast reduction (n = 17; group 1), ≥50% blast reduction without objective response (n = 4; group 2), and objective response (n = 5, group 3). Representative t-SNE plots for screening and on-treatment samples for individual patients from these analysis groups are shown in Fig. 2C. After a full cycle of treatment with FHD-286+DAC, group 1 showed minimal changes to cell type abundance, while group 2 exhibited a modest reduction in the blast compartment and concomitant increases in the myeloid compartment (Fig. 2D). Group 3 had an extensive reduction in the blast compartment and was the only group to have substantial increases in the erythroid compartment, suggesting a potential mechanistic linkage of these two events that may result in an objective clinical response to FHD-286+DAC.

### Blast transcriptional effects are associated with disease response

To expand on these initial observations, we conducted single-cell genomics on BMA samples with sufficient cells for analysis. We performed single-cell RNA-sequencing (scRNA-seq) on 90 samples from 40 patients (36 paired, 4 unpaired), and identified 574,966 cells in 9 clusters, which were annotated based on well-established cell type–specific markers and gene expression signatures (Fig. 3A, fig. S6A). The dataset was successfully normalized and integrated, as evidenced by the fact that multiple samples and patients contributed to the annotated cell clusters (fig. S6B, C). Additionally, cells had qualitatively uniform distributions on the UMAP (uniform manifold approximation and projection), regardless of whether they were from screening or on-treatment visits, or from monotherapy or combination therapy cohorts (fig. S6D, E). Annotation of the blast compartment, comprising 286,655 cells, was conducted using orthogonal methods, including expression signatures and inferred genomic copy number analysis (fig. S7A-C). Importantly, the relative blast abundance determined by scRNA-seq was highly correlated with the blast abundance determined morphologically at clinical sites, indicating accurate identification of the blast compartment in the single-cell genomics dataset (fig. S7D).

**Fig. 3.**
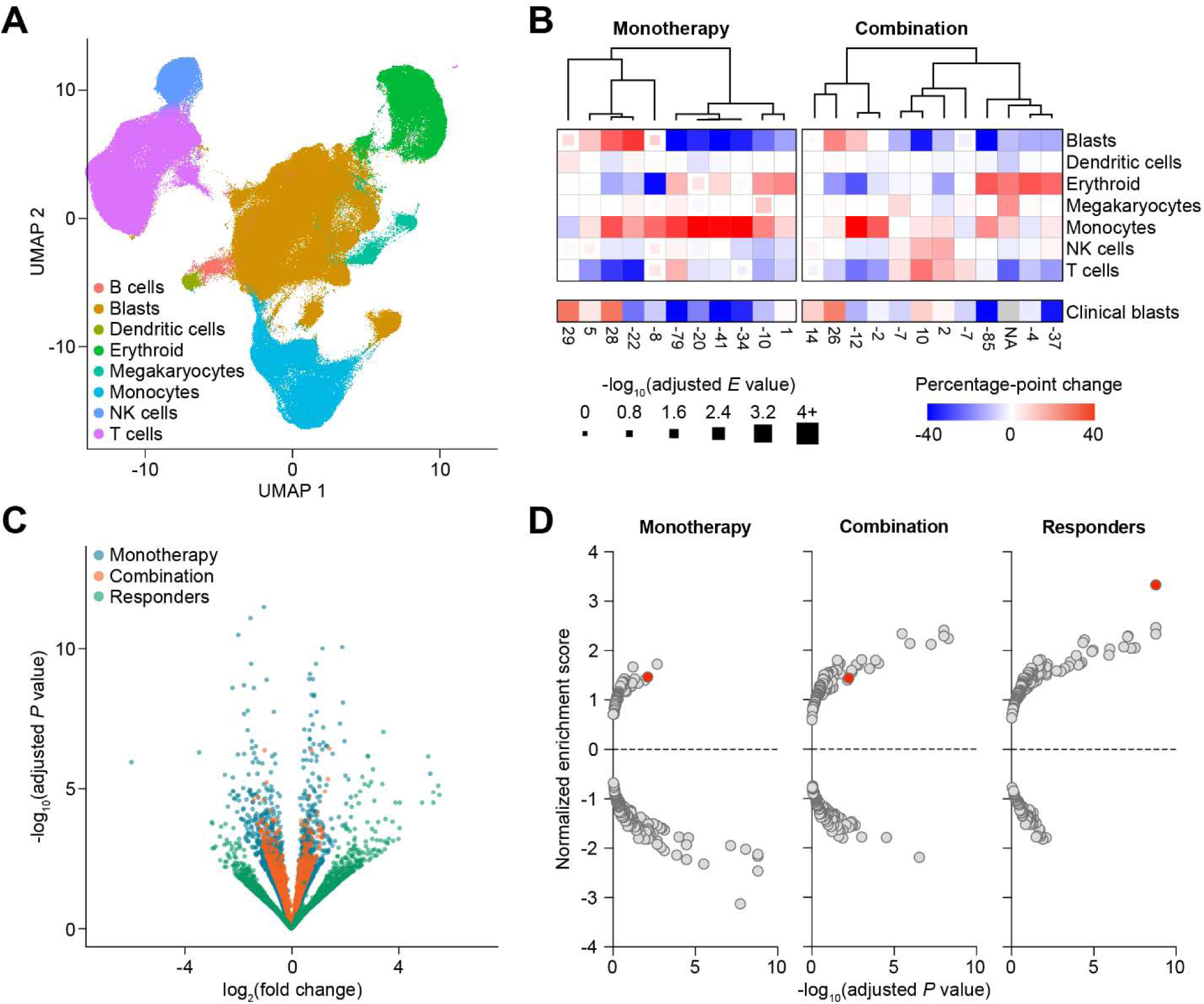
Blasts from FHD-286+DAC responders show upregulation of genes associated with erythroid differentiation. (**A**) UMAP visualization of all 574,966 cells in dataset. (**B**) Heatmap of percentage-point changes from screening to treatment (C1D15 or C2D1) in cell type abundance for patients with paired samples. (**C**) Volcano plot of DGE comparing screening and on-treatment (all time points) patient samples, stratified by monotherapy, combination therapy, and objective responders. (**D**) Pathway analysis of differentially expressed genes shown in (C). Red dot indicates Hallmark heme metabolism gene signature.

To broadly assess the impact of FHD-286+DAC, we compared the relative percent abundance changes in each cell type for each patient at C1D15 or C2D1 versus their matched screening sample. Substantial reductions in blast cluster abundance were observed in a subset of patients (Fig. 3B). Among patients in the FHD-286 monotherapy cohort who experienced substantial blast reductions, the most pronounced increases in percent abundance were in the monocyte cluster. In contrast, patients who experienced substantial blast reductions in the FHD-286+DAC combination cohort had the most pronounced increases in percent abundance in the erythroid cluster.

We then performed differential gene expression (DGE) analysis within the blast cluster, comparing screening and on-treatment samples for patients in the monotherapy portion of the study, in the combination portion of the study (regardless of response), and those who had an objective response. In the combination cohort, a weaker transcriptional impact was observed compared with monotherapy, in keeping with the weaker myeloid differentiation observed by flow cytometry (Fig. 3C). However, responders exhibited a markedly greater transcriptional impact than either the combination cohort or the monotherapy cohort. Gene set enrichment analysis (GSEA) was performed using the Hallmark and KEGG gene set collections, and Hallmark Heme Metabolism was the most significantly upregulated gene set among responders (Fig. 3D). Collectively, these results suggest that FHD-286+DAC promotes erythroid maturation of leukemic blasts and this transcriptional impact is associated with disease response.

### FHD-286 potentiates DAC transcriptional impact

The vast majority of patients in this trial had previously progressed after HMA therapy, including 5 of the 6 patients with an objective response, suggesting that DAC was unlikely to be the sole contributor to the responses observed. Therefore, we sought to confirm that the observed transcriptional effects were driven by the combined activities of FHD-286 and DAC. To our knowledge there is no broadly accepted clinically derived gene signature of the effect of DAC on AML blasts. We therefore generated a DAC gene expression signature using published scRNA-seq data from an analysis of BMA samples from 14 patients with AML or MDS (2 untreated, 12 R/R) before and after treatment with a single cycle of DAC monotherapy administered using the same dose schedule as in our study (*13*). To ensure consistency, we reanalyzed the published primary data using methodology identical to that used to analyze our scRNA-seq dataset (fig. S8A-D). Based on DGE analysis of screening versus on-treatment samples for the pseudobulked blast cluster, we constructed a transcriptional response signature containing the 144 genes significantly modulated by DAC (134 genes were induced and 10 genes were repressed) (fig. S9A). Consistent with previously described effects of DAC (*14–16*), we found that induced genes were highly enriched for signatures associated with erythroid differentiation (fig. S9B).

We next investigated whether the identified DAC transcriptional signature was modulated in response to FHD-286 and FHD-286+DAC. Interestingly, while effects on individual genes in the signature were relatively modest in the monotherapy and combination therapy cohorts, a substantial fraction of genes was significantly modulated among responders (Fig. 4A). Consistent with this observation, GSEA demonstrated that there was significantly greater enrichment of the DAC-induced and DAC-repressed gene sets in responders (fig. S9C). To directly investigate the impact of FHD-286 on DAC-modulated genes, we analyzed the DAC signature as a function of FHD-286 plasma concentration. Modulation of a substantial fraction of the DAC signature genes was significantly correlated with FHD-286 plasma concentration in patients treated with FHD-286+DAC (Fig. 4B). This observation was further supported by the expected directionality of GSEA with DAC-induced and DAC-repressed gene sets, with respect to FHD-286 plasma concentration (Fig. 4C). To further investigate these effects, we conducted gene set variation analysis (GSVA), which allows pathway analysis on individual samples. The expression of DAC-induced genes was strongly positively correlated with FHD-286 plasma concentration, while the DAC-repressed genes showed strong negative correlation with FHD-286 plasma concentration (Fig. 4D). DAC-induced and -repressed gene signatures were not significantly modulated by FHD-286 alone in the monotherapy cohort (Fig. 4D, fig. S9C). Taken together, these results suggest that FHD-286 potentiates the transcriptional impact of DAC to promote upregulation of key erythroid differentiation genes.

**Fig. 4.**
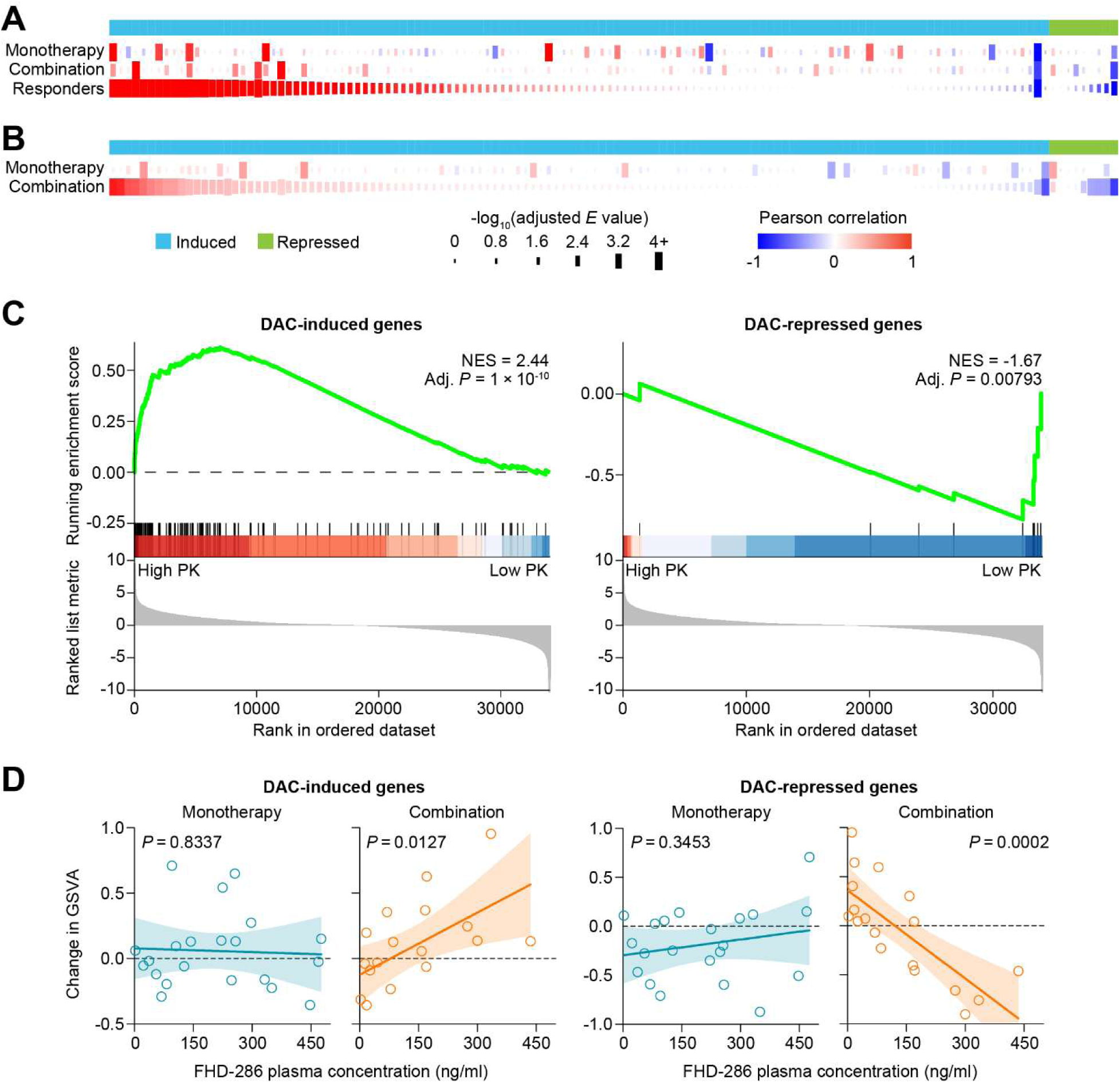
FHD-286 potentiates transcriptional impact of DAC in blasts. (**A**) Heatmap of logFC in expression from screening (SCR) to treatment (any time point) of genes in DAC response signature. Individual genes are displayed in columns. Patient cohorts and categories (monotherapy, combination therapy, objective responders) are displayed in rows. (**B**) Heatmap of logFC in gene expression with respect to FHD-286 plasma concentration for genes in DAC response signature. Individual genes are displayed in columns. Patient cohorts (monotherapy and combination therapy) are displayed in rows. FHD-286 plasma concentrations are multiplied by a factor of 100 for visualization. (**C**) GSEA of DAC response signature applied to differentially expressed genes with respect to FHD-286 plasma concentration in combination therapy patients. (**D**) Scatter plots of change (with respect to SCR) in GSVA of DAC response signature applied to individual patient samples versus FHD-286 plasma concentration. Shaded areas represent 95% confidence interval of the best-fit line. NES, normalized enrichment score; Adj., adjusted.

### Clonal analysis traces tumor cell differentiation into erythroid and monocyte compartments

Given the strong induction of erythroid maturation genes and the increased erythroid cell abundance by flow cytometry, we hypothesized that FHD-286+DAC may cause leukemic cells to differentiate into more mature erythroid precursors. Testing this hypothesis required a method to identify tumor cells that had potentially differentiated out of the transcriptionally defined blast compartment. With standard scRNA-seq methods, once a blast cell has differentiated into another compartment (eg, erythroid), it clusters transcriptionally with the cells in that compartment, making it indistinguishable from non-tumor cells. To overcome this limitation, we employed the computational tool Numbat, which integrates expression analysis of contiguous genomic regions with haplotype phasing of single nucleotide polymorphisms to infer copy number variants in scRNA-seq data (*17*). The amplifications and deletions identified by Numbat had statistically significant concordance (*P* = 1 × 10^-30^) with those identified by standard clinical cytogenetic testing (G-banding and fluorescence in situ hybridization [FISH]) (Fig. 5A, fig. S10A). Copy number variant profiles enabled discrimination between malignant and normal cells and provided clonal “fingerprints” that can be used to locate tumor cells within different cell clusters (Fig. 5B). Various patterns of differentiation were observed by longitudinal analysis, with tumor cells shifting out of the blast compartment and into the monocyte and erythroid compartments during treatment (Fig. 5C). In some cases, multiple tumor subclones were identified in a single patient, which could also be tracked longitudinally (fig. S10B, Fig. 5D). Remarkably, in all responding patients with paired samples for analysis (n = 3), the relative abundance of tumor clones decreased in the blast compartment and increased in either the monocyte or erythroid compartment during treatment (Fig. 5E). These results highlight the extent of differentiation in the responders, with a significant percentage of tumor cells exiting the transcriptionally defined blast compartment.

**Fig. 5.**
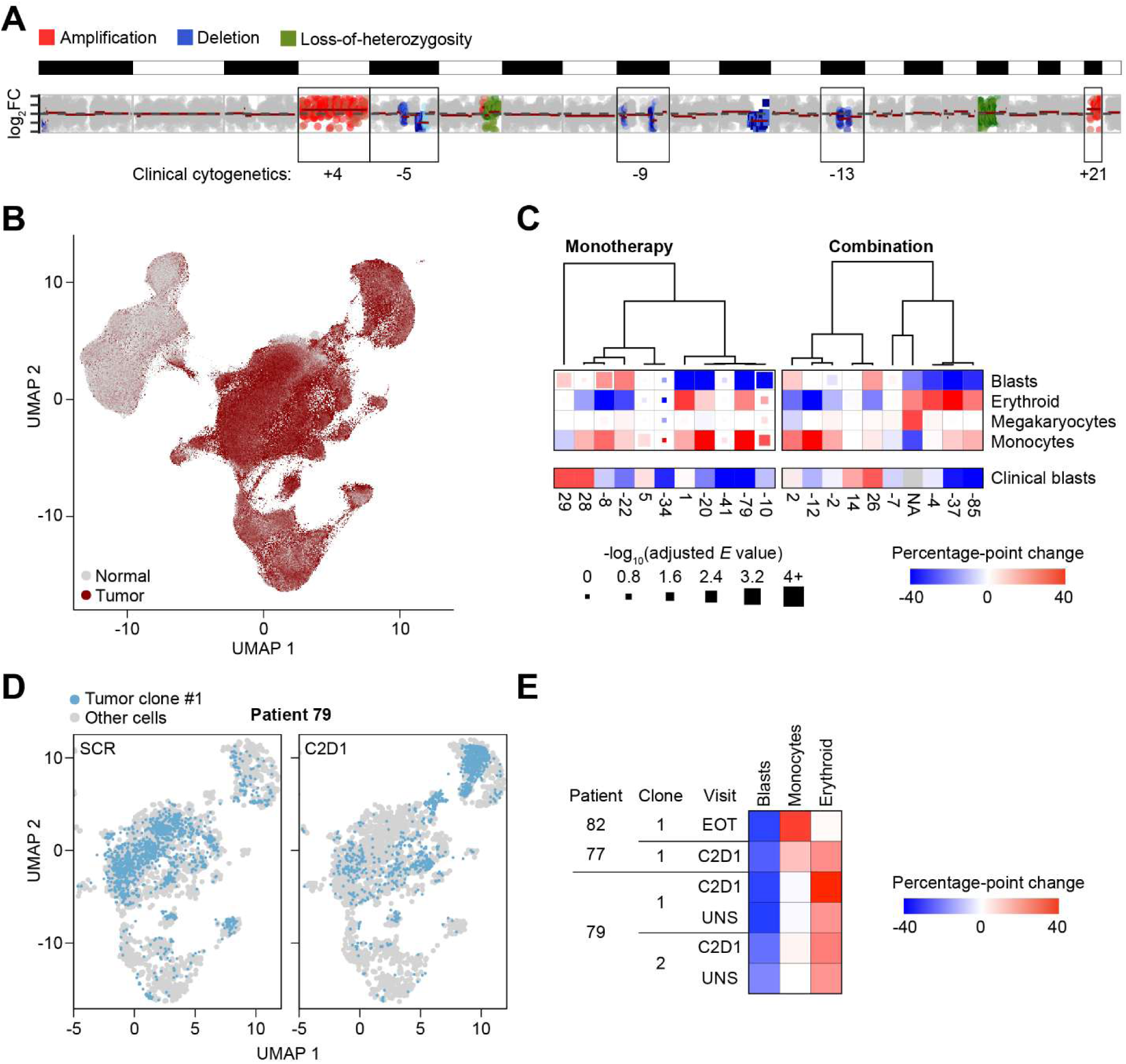
Tumor cell annotation enables analyses across multiple cell clusters. (**A**) Representative cytogenetic profile derived by Numbat analysis and comparison with cytogenetic alterations assessed clinically. Alternating black and white boxes above cytogenetic profile indicate positions of individual chromosomes, starting with chromosome 1 at left. Concordance of clinically measured cytogenetic aberrations is highlighted by individual boxes using chromosome-level resolution. (**B**) UMAP visualization of all tumor cells identified by Numbat. (**C**) Heatmap of percentage-point abundance changes from screening (SCR) to treatment (C1D15 or C2D1) in Numbat-annotated tumor cells within each cluster for patients with paired samples. (**D**) UMAP visualization of BM cells from Patient 79 (a responder) at SCR and C2D1 within the blast, monocyte, and erythroid compartments. (**E**) Heatmap of percentage-point abundance changes from SCR to treatment in individual tumor clones for responders with paired samples. UNS, unscheduled (Day 77).

### Responders display transcriptional similarity to *CEBPA*-mutant AML

To further understand the potential mechanisms underlying the clinical responses and identify associated biomarkers, we conducted an extensive correlation analysis of patient cytogenetic and gene mutation data collected at screening. Perhaps due to the small number of responders, none of these baseline characteristics were statistically significant predictors of response to FHD-286+DAC. Next, we investigated whether commonalities in transcriptional signatures present at screening could identify responders. We compared gene expression values from pseudobulked blast clusters between responders (n = 4) and nonresponders (n = 14) and identified a potential predictive signature of response to FHD-286+DAC. To overcome the limitations of the small number of patients in this trial, we mapped the signature onto large publicly available AML patient datasets by calculating Pearson correlation coefficients for patients in the BEAT-AML, TARGET-AML, and TCGA-AML datasets. Hierarchical clustering of the correlation coefficients revealed clear separation of patients according to well-established AML subtypes (Fig. 6A, fig. S11). Notably, the 4 responders were most closely related to patients with the *CEBPA*-mutant AML subtype, which is characterized by biallelic *CEBPA* mutations. There was strong transcriptional similarity between the objective responders and this AML subtype in all three large AML datasets (Fig. 6B, fig. S11A-D). While the potential FHD-286+DAC responder signature also showed some correlation with patients harboring mono-allelic *CEBPA* deletions, this correlation was weaker than with biallelic *CEBPA* mutations.

**Fig. 6.**
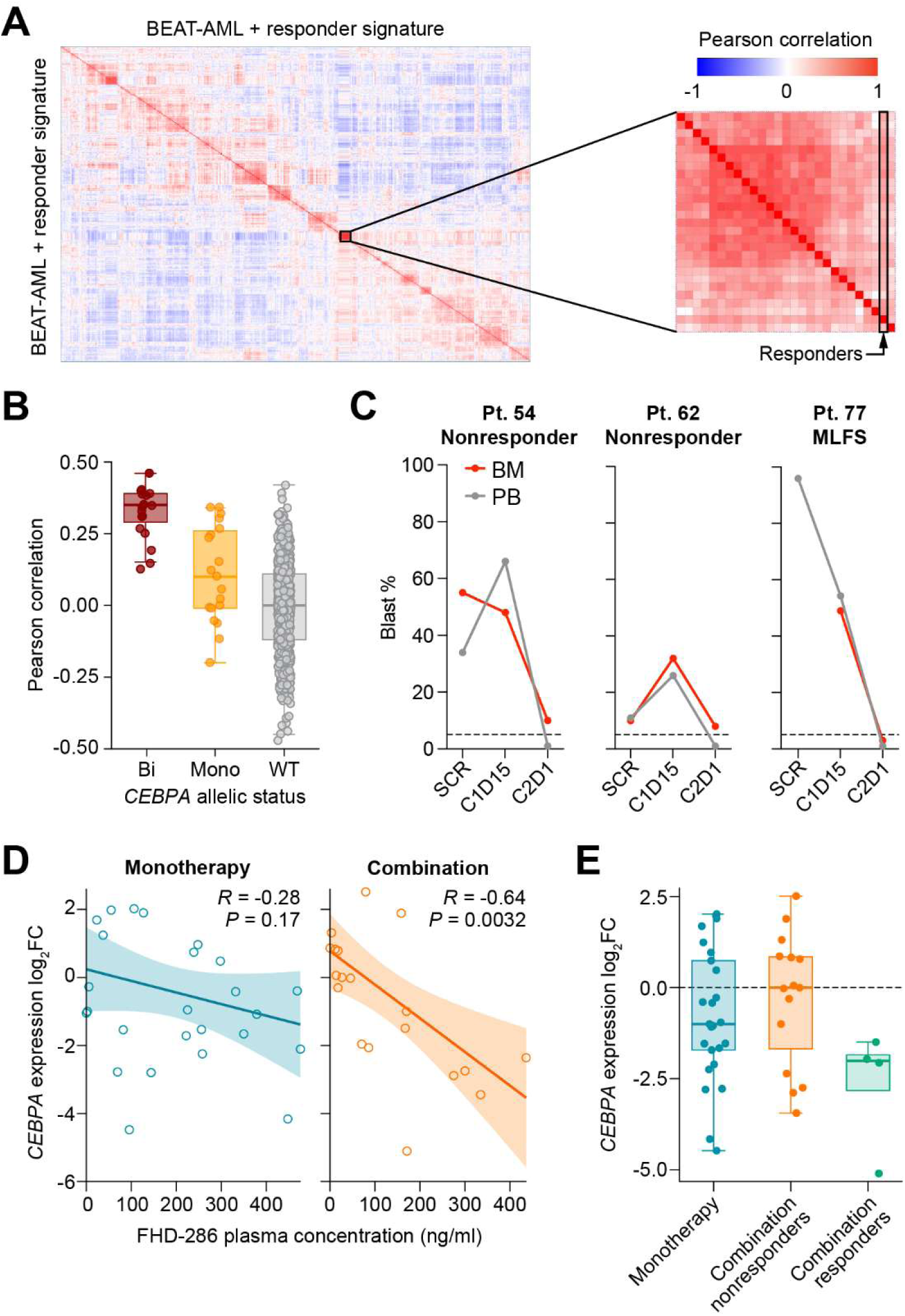
Responders display transcriptional similarity to patients with *CEBPA*-mutant AML. (**A**) Heatmap of pairwise Pearson correlation coefficients for comparison of transcriptional profiles of BEAT-AML patients and FHD286+DAC responders. BEAT-AML patients (n = 707) are represented individually in rows and columns. In addition, one sample represents a composite profile generated for all FHD286+DAC combination responders (n = 4). The subcluster representing *CEBPA*-mutant AML is indicated with a rectangle. The position of combination responders relative to this subcluster is indicated in zoomed-in heatmap. (**B**) Distribution of pairwise Pearson correlation coefficients from (A). Each point represents Pearson correlation coefficient of comparison between combination responders and all patients from BEAT-AML, stratified by *CEBPA* mutational status (biallelic [bi], monoallelic [mono], or wild-type [WT]). Box plots represent IQR, median, and 1.5 × IQR. (**C**) BM and peripheral blood (PB) blast percentages determined morphologically at clinical sites in all patients with *CEBPA* mutations in the FHD286+DAC combination cohort. (**D**) Changes (versus SCR) in *CEBPA* gene expression in individual patient samples vs FHD-286 plasma concentration at the corresponding visit. Shaded areas represent 95% confidence interval of the best-fit line. (**E**) Distribution of changes (versus SCR) in *CEBPA* gene expression in individual patient samples. Box plots represent IQR, median, and 1.5 × IQR.

Of the 4 patients contributing to the potential responder signature, 1 had a *CEBPA* mutation noted in the clinical data, 1 had a historical sample with a *CEBPA* mutation that was not detected at screening, and 2 did not have *CEBPA* mutations. Interestingly, 2 nonresponders treated with FHD-286+DAC also had *CEBPA* mutations, and both had substantial blast reductions on treatment (Fig. 6C). Thus, in this cohort, all 3 patients with *CEBPA* mutations at screening experienced substantial reductions in leukemic burden upon treatment with FHD-286+DAC.

To specifically interrogate the role of *CEBPA*, we analyzed the effect of treatment on *CEBPA* levels in blasts. While *CEBPA* expression was not significantly modulated by DAC monotherapy (data file S1) or FHD-286 monotherapy, it decreased as a function of FHD-286 plasma concentration in patients treated with FHD-286+DAC (Fig. 6D). Interestingly, when comparing paired screening and on-treatment samples, *CEBPA* appeared to be more dramatically downregulated in responders (Fig. 6E). While *CEBPA* mutation was only observed in 1 responder, the potent downregulation of *CEBPA* observed in most responders suggests potentially convergent mechanisms affecting *CEBPA* functional output.t.

### Diverse patterns of differentiation in objective responders

Using multiple orthogonal approaches, primarily multiparametric FACS and single-cell genomics, we characterized patterns of blast differentiation in objective responders. Our results revealed a diversity of differentiation patterns, either predominantly myeloid, predominantly erythroid, or mixed (Table 1). Collectively, these findings demonstrate that tumor cell differentiation is a key mechanism of action of FHD-286+DAC and highlight that various patterns of lineage maturation can enable response to this combination treatment.

**Table 1.** Patterns of cellular differentiation observed in responders. Increased (↑)/decreased (↓): >10 percentage-point change in the indicated measure relative to screening (SCR). No change (↔): <10 percentage-point change in the indicated measure relative to SCR. a. No SCR sample available; C1D15 sample used. b. No SCR or C1D15 samples available. NA, not available; BOR, best overall response; WT, wild type.

| Patient | Type of cancer | CEBPA status | BOR | Differentiation |  |  |  |
| --- | --- | --- | --- | --- | --- | --- | --- |
|  |  |  |  | Measure |  |  | Predominant overall pattern |
|  |  |  |  | % CD11b<sup>+</sup> blasts (FACS) | % Cells in erythroid or myeloid clusters (FACS) | % Tumor cells in erythroid or myeloid clusters (Numbat) |  |
| 46 | AML | WT | MLFS | ↔ | Erythroid ↑ | NA | Erythroid |
| 50<sup>a</sup> | AML | WT | CRp | ↑ | Erythroid ↑<br>Myeloid ↓ | NA | Mixed |
| 60<sup>b</sup> | AML | WT | CRp | NA | NA | NA | NA |
| 77 | AML | Mutated | MLFS | ↑ | Erythroid ↑<br>Myeloid ↑ | Erythroid ↑<br>Monocyte ↑ | Mixed |
| 79 | AML | History of mutation | MLFS | ↓ | Erythroid ↑<br>Myeloid ↓ | Erythroid ↑ | Erythroid |
| 82 | MDS | WT | PR | ↑ | ↔ | Monocyte ↑ | Myeloid |

## Discussion

Myeloid malignancies are characterized by the clonal expansion of hematopoietic progenitor cells that fail to properly differentiate into mature blood cells. While cytotoxic agents are the backbone of current treatment regimens, they do not address this fundamental aspect of disease biology. The therapeutic potential of differentiation-based approaches is exemplified by all-*trans* retinoic acid/arsenic trioxide, which has been transformative for the treatment of acute promyelocytic leukemia with *PML::RARA,* but has limited activity in other AML subtypes (*18*). More recently, several targeted agents have been developed with the ability to overcome the leukemic differentiation block in specific genetic contexts. These include inhibitors of IDH1/2 (ivosidenib, olutasidenib, enasidenib), FLT3 (midostaurin, gilteritinib, quizartinib), and menin (revumenib, ziftomenib) (*19*). Responses to IDH inhibitors have been associated with morphologic evidence of myeloid differentiation, and the emergence of functional *IDH2*-mutant neutrophils was observed in patients responding to enasidenib (*20–22*). Terminal blast differentiation has also been observed with *FLT3* inhibitors, with some patients experiencing increases in mature neutrophils harboring *FLT3* mutations (*23*). Finally, increased expression of myeloid maturation markers and morphologic evidence of myeloid differentiation was observed with revumenib in *KMT2A*-rearranged or *NPM1*-mutant AML (*24, 25*).

In our study, inhibition of SMARCA4/2 with FHD-286 promoted myeloid differentiation, consistent with the role of BAF in leukemic stemness maintenance. While this pro-differentiation effect did not translate into objective responses with FHD-286 monotherapy, 6 of 47 (12.8%) patients treated with FHD-286+DAC achieved an objective response (CRp, PR, or MLFS). We therefore considered whether the improved clinical activity of the combination was due solely to DAC. While impossible to conclude with certainty due to the lack of a control arm, several lines of evidence strongly point to a role for FHD-286 in contributing to the efficacy of the combination. First, the patients in our study had almost universally failed prior therapy with an HMA and were thus unlikely to respond to rechallenge with an HMA alone (*26*), which indirectly suggests that FHD-286 may resensitize some patients to DAC. Second, the median time to best response with FHD-286+DAC (1.9 months) was notably shorter than what is typically observed with the equivalent dosage of DAC monotherapy (4.3 months in patients with newly diagnosed AML) (*27*), indicating that the addition of FHD-286 may result in enhanced therapeutic activity over single-agent DAC. Finally, modulation of DAC-induced and -repressed genes correlated well with FHD-286 plasma concentrations, suggesting FHD-286 potentiates the transcriptional impacts of DAC.

The improved clinical activity imparted by the FHD-286+DAC combination was transcriptionally and phenotypically underpinned by marked erythroid differentiation, a mechanism of action only sporadically reported in the context of targeted therapies for AML. Anecdotal cases of erythroid differentiation have been reported with gilteritinib treatment (*28*), while enasidenib has been shown to promote high rates of red blood cell (RBC) transfusion independence, suggesting a role for IDH2 inhibition in stimulating erythroid differentiation (*29*). Several pro-erythroid therapies are approved for the treatment of anemia in patients with lower-risk MDS, including erythropoietin, lenalidomide, the transforming growth factor-beta ligand trap luspatercept, and the telomerase inhibitor imetelstat (*30–32*). In AML and high-risk MDS, the HMAs azacitidine and DAC have shown clinical evidence of erythropoietic stimulation in the form of RBC count recovery and reduced transfusion burden (*15, 16*). The active metabolite of DAC is a cytidine analog that incorporates into DNA during replication and inhibits DNA methyltransferase activity, leading to global DNA hypomethylation (*33*). While higher concentrations of DAC are primarily cytotoxic, lower concentrations promote cellular differentiation, perhaps through promoter demethylation and subsequent activation of myeloid lineage transcription factors (*34*). In a study in patients with AML (primarily newly diagnosed), DAC induced both myeloid and erythroid differentiation, with evidence that patients experiencing erythroid differentiation were more likely to achieve a response (*14*). Collectively, these results suggest that therapeutic strategies aimed at promoting erythroid differentiation in the context of AML warrant further investigation.

In our scRNA-seq cohort, objective responders had a baseline expression profile most closely related to that of *CEBPA*-mutant AML. *CEBPA* is a master myeloid transcription factor that is upregulated in granulocyte-monocyte progenitors and downregulated in megakaryocyte-erythroid progenitors during normal hematopoiesis (*35*). Experimentally, overexpression of *CEBPA* in hematopoietic stem cells induced myeloid differentiation and inhibited erythroid differentiation, while conditional knockout of *CEBPA* in mice led to increased erythropoiesis (*36*). Somatic mutations in *CEBPA*, which typically confer loss of function, occur in approximately 10% of AML (*37–40*). Consistent with the role of *CEBPA* in myeloid lineage specification, *CEBPA*-mutant AML has been associated with a more erythroid gene expression profile and immunophenotype (*41, 42*). Our results suggest erythroid differentiation and objective responses observed in this trial could be driven by transcriptional similarity to *CEBPA-*mutant AML and further reductions of *CEBPA* levels upon FHD-286+DAC treatment.

This study has several limitations. The number of objective responders was very small (n = 6), and the lack of a DAC control arm prevented a more balanced direct comparison. However, the use of a historical dataset, reanalyzed using our methodology, allowed us to understand the transcriptional changes underpinning the DAC response mechanism in a similar patient population (*13*). Furthermore, we have taken multiple steps to validate our analysis of tumor cells in cellular compartments other than blasts. However, we have not confirmed these observations using independent methods, such as conducting sequencing or FISH analyses on specific FACS-sorted differentiated cell populations.

In summary, we have analyzed samples from a first-in-human FHD-286+DAC combination trial and identified diverse patterns of tumor cell differentiation. Erythroid differentiation was a striking feature among responders, who displayed a transcriptional profile similar to that of *CEBPA*-mutant AML. These results provide guidance for further clinical development of FHD-286+DAC in genetically defined patient subpopulations.

## Materials and Methods

### Study design and methodology

FHD-286-C-002 was an open-label, non-randomized, phase 1, 3+3 dose escalation study of FHD-286 in patients with R/R myeloid malignancies designed to assess safety, tolerability, PK, pharmacodynamics, and preliminary clinical activity, and to determine the MTD and/or recommended phase 2 dose(s). Primary study endpoints included AEs and DLTs. Secondary study endpoints included PK and preliminary clinical activity. Exploratory endpoints included pharmacodynamics and exploratory biomarker analysis. The study design and methodology of the monotherapy portion of this study are described elsewhere (*7*). The combination portion of this study was conducted at 5 sites in the United States. FHD-286 was administered orally QD in 28-day cycles in combination with DAC 20 mg/m^2^ on days 1 through 5 of each cycle. A low-dose cytarabine arm was also included in the study; however, no patients were enrolled to that arm. Patients with a confirmed diagnosis of R/R AML, R/R MDS, or R/R CMML not in blast crisis who had received ≤4 prior lines of systemic anticancer therapy for their disease under study and were appropriate candidates for the combination therapy were assigned to groups based on whether they were receiving a triazole antifungal agent classified as a strong CYP3A inhibitor at the start of study treatment:

- Group B1: Subjects *not* receiving such an agent
- Group B2: Subjects receiving such an agent

Patients continued treatment until confirmed disease progression per modified IWG criteria for AML or MDS (used for CMML as well) (*11, 12*), treatment failure, death, unacceptable toxicity, start of alternative anticancer therapy, or study withdrawal.

The mechanism of action of FHD-286 includes promotion of differentiation of leukemic cells, which carries a risk of clinical DS. Guidelines for monitoring and management of suspected DS were provided to investigators in the protocol. An ISMC reviewed cumulative and patient-level safety data and retrospectively identified and graded cases of possible DS among all treated patients.

Treatment response was evaluated by BM biopsy and/or aspirate, along with complete blood counts and peripheral blood blast counts, on C1D15, C2D1, and day 1 of every cycle thereafter, based on the modified IWG criteria for AML or MDS (also used for CMML), as appropriate (*11, 12*), with stable disease in AML defined per protocol. Bone marrow biopsies and/or aspirates were also used for exploratory analyses. Serial blood samples for pharmacodynamics and/or exploratory biomarker analyses were obtained at predose and 8 hours postdose on C1D11; at predose on C1D2, 8, and 15; at predose on C2D1 and C3D1; and at predose on day 1 of every other cycle thereafter.

The study was conducted in accordance with the principles in the Declaration of Helsinki and Good Clinical Practice guidelines and was approved by the institutional review board at each site. Written informed consent was obtained before patient participation.

Full details are available in the protocol **(Supplementary Materials)**.

### Statistical analysis for clinical data

The safety and full analysis populations included all patients who had received ≥1 dose of FHD-286. The PK analysis population included all patients who received ≥1 dose of FHD-286 and had ≥1 blood sample providing evaluable PK data. Nominal dates and time points, and assigned doses, were used for the PK analysis. Descriptive statistics were used for clinical, laboratory, PK, pharmacodynamic, and exploratory variables. This study was not designed for formal hypothesis testing, and no power analyses were done.

### Pharmacokinetics

Serial blood samples for analysis of FHD-286 plasma concentrations and calculation of PK parameters were collected, processed, and analyzed as previously described (*7*).

### Flow cytometry

Bone marrow aspirates were collected at study sites and sent to a central laboratory (PPD, Inc.) for processing. Bone marrow mononuclear cells (BMMCs) were isolated by density gradient centrifugation, cryopreserved, and sent to Discovery Life Sciences for analysis. Monotherapy samples were stained with viability dye (DAPI, BioLegend #422801), blocked with an Fc receptor blocking solution (Human TruStain FcX, Biolegend #422302), and incubated with a cell surface antibody panel (table S7). Samples from the combination portion of the study were analyzed using a modified set of antibodies to detect both cell surface and intracellular antigens. Following viability dye staining (LIVE/DEAD Fixable Aqua Dead Cell Stain, ThermoFisher #L34957), blocking (Biolegend #422302), and cell surface antibody incubation, combination therapy samples were fixed/permeabilized (eBioscience Foxp3/Transcription Factor staining kit, Invitrogen #00-5523-00) and incubated with an intracellular antibody panel (table S7). All samples were run on a Miltenyi Bioscience MACSQuant Analyzer 16 flow cytometer (RRID:SCR_020263) (Miltenyi Biotec) and data were analyzed using FlowJo software (RRID:SCR_008520) (Becton, Dickinson & Company). Fluorescence compensation was performed using anti-mouse Igκ (Miltenyi #130-097-900) and ArC amine reactive beads (Invitrogen #A10346), as appropriate. Samples were gated to exclude debris, doublets, and dead cells, and blasts were identified as CD45 dim and SSC intermediate.

### High-dimensional FACS analysis

High-dimensional analysis was performed on 90 BM samples from patients in the combination portion of the study. All compensated parameters within the live cell gate were exported from FlowJo as new .fcs files and imported into OMIQ (RRID:SCR_027879; Dotmatics, Boston, MA, USA). The fluorescence data were scaled using the arcsinh function with a cofactor of 400. Dimensionality reduction was performed with optimized t-SNE (opt-SNE) (*43*) using the default settings in OMIQ (perplexity = 30). A total of 505 iterations were performed, with 45 as early exaggeration, and the final Kullback-Leibler divergence was 5.952976. Clustering was performed with FlowSOM (RRID:SCR_016899) (*44*) using an 8×8 grid, 10 training iterations, and Euclidean distance metric. The elbow metaclustering method was used to identify the optimal number of metaclusters.

FACS clusters were identified primarily with the following immunophenotypes:

- Blasts: SSC^int^/CD34^hi/^CD117^hi^
- Monocytic: SSC^hi^/CD11b^hi^/CD64^hi^
- Granulocytic: SSC^hi^/CD11b^hi^/CD15^hi^
- Early erythroid: CD45^lo^/Lin^lo^/BRG1^hi^
- Late erythroid: CD45^lo^/Lin^lo^/BRG1^int^
- B lymphocytic: SSC^lo^/CD45^hi^/CD123^int^
- T lymphocytic SSC^lo^/CD45^hi^/CD15^int^

### Determining association of cytogenetic alterations and genetic mutations with clinical response status

Cytogenetic alterations and genetic mutations reported at screening were tested to determine whether they predicted clinical response using Fisher’s exact test. Each alteration (present/not present) was compared with responder status (yes/no).

### Single-cell RNA-sequencing

For monotherapy samples, RNA was prepared, sequenced, and analyzed as previously described (*7*). For combination therapy samples, viably frozen BMMCs were analyzed at Discovery Life Sciences using a 10x Genomics kit (v3.1) on a Chromium Connect or Chromium Controller per the manufacturer’s protocol. Sequencing of prepared libraries was done using an Illumina NovaSeq 6000 Sequencing System (RRID:SCR_016387) (Illumina) using an S2 or S4 flow cell.

Single-cell sequencing data were aligned to the human reference genome (GRCh38) and processed using 10x Genomics Cell Ranger (RRID:SCR_017344; v3.1.0) to generate unique molecular identifier counts. The raw gene expression matrices were imported into R (“R Project for Statistical Computing;” RRID:SCR_001905) and further processed by the Seurat package (RRID:SCR_016341; v3.2.2). Low-quality cells were excluded from further analysis if the total gene count was <150 or the mitochondrial gene count was >10%. In a procedure analogous to EmptyDrops (*45*), further quality control was performed by evaluating the distribution of reads per droplet to identify 2 major modes in the “kneeplot.” The first mode contains a higher number of reads per droplet and corresponds to cell-containing droplets, while the lower mode with fewer reads corresponds to non–cell-containing droplets. Droplets in the lower mode were removed from further analysis. Doublets were removed using scDblFinder (RRID:SCR_022700) (*46*). The dataset was integrated using Harmony (RRID:SCR_022206) (*47*) and cell clusters were visualized using UMAP (RRID:SCR_018217). Cells were clustered using Seurat and the gene markers for each cluster were identified using the Seurat function “FindAllMarkers.” The cell type of each cluster was then identified using a combination of SingleR (RRID:SCR_023120), a published pediatric AML blast gene expression signature (*48*), Azimuth (RRID:SCR_021084), BoneMarrowMap (RRID:SCR_028324) (*49*), and manual curation using canonical marker genes. Scoring of the AML blast gene expression signature was calculated on a per-cell basis using the UCell algorithm (*50*). Copy number analysis to further support the annotation of the blast clusters was conducted using CopyKAT (RRID:SCR_024512) (*50*).

To identify differentially expressed genes (DEGs) while accounting for interindividual variability, a pseudobulk approach was employed. Single-cell RNA-seq counts were aggregated to the sample level using the AggregateExpression function in Seurat (v5.2.1) in R (v4.4.2). Specifically, raw counts were summed for each cell type within each sample.

Gene expression analysis was done for 3 major contrasts: treated versus screening, gene expression versus FHD-286 plasma concentration, and objective responders versus nonresponders. For the first 2 contrasts, a multifactor design was used to control for patient-specific effects:

- Design = patient + time point
- Design = patient + plasma concentration

Statistical analysis was performed using DESeq2 (RRID:SCR_015687, v1.43.5) in R (v4.4.0). Before formal testing, low-abundance genes were filtered to retain only those with a total count >1 across all samples (rowSums >1).

The screening time point was defined as the reference (baseline) level for all comparisons. Differential expression was tested using the Wald test. To provide more accurate estimates for effect size, log_2_ fold change (log_2_FC) values were processed using the ashr shrinkage estimator (*51*).

Significant DEGs were defined based on an adjusted *P* value (Benjamini-Hochberg) ≤0.05, an absolute shrunken log_2_FC ≥0, and average normalized counts greater than 0. Analysis was conducted on the blast cell type only.

Gene set enrichment analysis was calculated using the GSEA function within clusterProfiler (RRID:SCR_016884, v4.12.0). Gene set variation analysis was calculated using the GSVA package in v1.52.3 in R (*52*).

### Numbat analysis

Numbat (RRID:SCR_028323, v1.4.2) was used to analyze scRNA-seq data to distinguish tumor cells from normal cells (*17*). Bone marrow aspirate samples from patients from our study who had clinically measured blast percentages <10% were used as the normal reference. Default settings were used, except for samples for which the entropy parameter was increased to allow Numbat to run to completion. The effect of this change was tested by varying the entropy parameter on several samples that ran successfully with the default value. The results were highly concordant, indicating that the use of a different entropy parameter setting did not affect the results. Samples were pooled for each patient, and Numbat was run on a per-patient basis, allowing the tracking of clones that recurred across patient samples and the analysis of longitudinal increases or decreases in the proportion of clones.

The cytogenetic alterations identified in Numbat were manually compared with the cytogenetic alterations reported at screening, establishing concordance. Statistical significance was calculated using the Fisher exact test.

### Calculation of change in clone and cell type proportions within scRNA-seq

Changes in clone and/or cell type proportions within scRNA-seq (cell types, Numbat-identified tumor cells within cell types, Numbat-identified clones within cell types, or clone overall proportion) were calculated as the percentage value on treatment minus the percentage value at screening. Using the assumption that the count of cells in each category approximately follows a Poisson distribution, an E-test (*53*) was used via scipy.stats.poisson_means_test to estimate the statistical significance of the changes observed.

### Derivation of decitabine gene signature

To establish a DAC-induced gene signature, a publicly available scRNA-seq dataset (Gene Expression Omnibus accession number GSE223844) collected as part of a phase 1 clinical trial in patients with R/R MDS or AML treated with ipilimumab and DAC was used (*13*). Samples collected at screening and at the end of the priming cycle of DAC monotherapy (EOLN) were used. Samples from the subsequent cycles of ipilimumab and DAC administration were not included. A total of 18 patient samples were collected at both screening and EOLN. After quality control, 14 met the predefined criteria and the corresponding raw gene expression matrices were further analyzed, as described in the **Single-cell RNA-sequencing** section. Genes that were statistically differentially regulated (adjusted *P* value <0.05 and abs(log_2_FC) ≥1) were included in the DAC-responsive gene signature.

### Public AML datasets

To identify a potential predictive signature of response to FHD-286+DAC, the t-statistic from the DGE for the comparison of responders (n = 4) vs nonresponders in combination therapy (n = 14) at screening was used, with the modification that all genes with a *P* value ≤0.05 were retained.

Normalized gene expression tpm (transcripts per million) values and clinical and mutational data were downloaded from the BeatAML2, TARGET-AML, and TCGA-LAML portals (https://biodev.github.io/BeatAML2/, https://portal.gdc.cancer.gov/projects/TARGET-AML, and https://portal.gdc.cancer.gov/projects/TCGA-LAML). To ensure comparability across patients, expression values were z-score normalized before correlation analysis for each cohort. Subsequently, Pearson correlation coefficients were computed to assess transcriptional similarity between the objective responders and patients in the 3 publicly available AML datasets.

### Mutational and cytogenetic analysis

Analysis of gene mutations and cytogenetics was conducted in BM samples as part of the clinical trial (n = 34 for monotherapy and 40 for combination therapy). The method of molecular testing was determined by the study site and included next-generation sequencing (frequently MSK-IMPACT), polymerase chain reaction, and FISH.

## Supporting information

Protocol FHD-286-C-002

## Data availability

All data needed to evaluate the conclusions in the paper are present in the paper and/or the Supplementary Materials. The clinical data, including the human sequence data, are not publicly available due to patient privacy requirements, but are available upon reasonable request to the corresponding authors.

## SUPPLEMENTARY MATERIALS

Protocol FHD-286-C-002

Data file S1: Penter DAC Monotherapy Pseudobulked Blast Deseq2

Fig. S1 to S11

Table S1 to S7

References cited only in supplementary materials: 54-56

## ACKNOWLEDGMENTS

We acknowledge and thank the participants in this clinical trial and their families.

## Author contributions

Conceptualization: AQ-C, DLL, GAS, SAR

Methodology: DLL, GAS, KH, MPC, SAR, TZ

Software: DLL, KD, MB, ST

Formal analysis: AK, AQ-C, BB, CDD, DH, DLL, EMS, GAS, JB, KD, MB, MPC, NS, ST, TZ

Investigation: AK, BB, CDD, DLL, EMS, MPC

Resources: AK, BB, CDD, EMS

Data curation: DLL, JB, KH, MPC, ST, TZ

Writing – original draft: APK, AQ-C, DLL, GAS, MPC

Writing – review & editing: all authors

Visualization: APK, DLL, KD, MPC, ST, TZ

Supervision: AK, AQ-C, BB, CDD, DH, DLL, EMS, GAS

Project administration: KH, KL, NP

## Competing interests

AK declares consulting relationships with Sobi, Geron Corporation, MorphoSys AG, Rigel Pharmaceuticals, Servier Pharmaceuticals, Syndax Pharmaceuticals, Blueprint Medicines, Ascentage Pharma, Takeda Oncology, Cogent Biosciences, and Novartis. The author serves as co-host of Blood Cancer Talks podcast and has received honoraria and consulting fees from the aforementioned companies within the past 36 months.

APK, DH, GAS, and NP declare employment by and stock options in Foghorn Therapeutics Inc.

AQ-C, NS, and ST declare employment by Foghorn Therapeutics Inc.

CDD declares consultancy for AbbVie, AstraZeneca, Astellas, BeOne, BMS, Kura, Rigel, Servier, Solu, and Systimmune. CDD is supported by the LLS Scholar in Clinical Research Award.

DLL declares employment by and stock in Foghorn Therapeutics Inc., inventor on patent for use of FHD-286 in treating cancer: US Patent # 12383555, “Methods of treating cancers.”

EMS declares institutional research support from and consulting for Foghorn Therapeutics Inc.

JB declares employment by Certara.

KD declares employment by Diamond Age Data Science.

KH and KL declare former employment by Foghorn Therapeutics Inc.

MB declares employment by Diamond Age Data Science and the Worcester Polytechnic Institute.

MPC declares employment by and stock options in Foghorn Therapeutics Inc. and patent applications WO2025080767A1 and WO2026085172A1.

SAR declares employment by Akebia Therapeutics, former employment by Foghorn Therapeutics Inc., and stock in Adaptimmune Therapeutics and Foghorn Therapeutics Inc.

The remaining authors declare that they have no competing interests.

