## Supplementary material for "Mechanism of response to FHD-286 and decitabine combination in patients with advanced myeloid malignancies": Protocol FHD-286-C-002

### CLINICAL STUDY PROTOCOL
### FHD-286-C-002
### (EUDRACT NUMBER 2020-005567-32)

#### A Phase 1, Multicenter, Open-Label, Dose Escalation Study to Assess the Safety, Tolerability, Pharmacokinetics, Pharmacodynamics, and Clinical Activity of Orally Administered FHD-286, as Monotherapy or Combination Therapy, in Subjects With Advanced Hematologic Malignancies

| | |
|---|---|
| Study Sponsor: | Foghorn Therapeutics Inc.<br>500 Technology Square<br>Cambridge, MA 02139 USA<br>Phone: +1 617-586-3100<br>Fax: +1 617-649-8418 |
| Responsible Medical Officer: | [redacted]<br>500 Technology Square<br>Cambridge, MA 02139 USA<br>Office Phone: [redacted]<br>Mobile Phone: [redacted]<br>Email: [redacted] |
| Study Medical Monitor | Name and contact information are provided in the Investigator Site File |
| Document Version (Date): | Version 1.0 (02 December 2020)<br>Version 2.0 (06 January 2021); Amendment 1<br>Version 3.0 (08 March 2022); Amendment 2<br>Version 4.0 (25 May 2023); Amendment 3<br>Version 5.0 (30 November 2023); Amendment 4<br>Version 6.0 (26 February 2024); Amendment 5<br>Version 7.0 (14 May 2024); Amendment 6 |

This study will be conducted according to the protocol and in compliance with Good Clinical Practice, the ethical principles stated in the Declaration of Helsinki, and other applicable regulatory requirements.

**CONFIDENTIALITY NOTE:**

The information contained in this document is confidential and proprietary to Foghorn Therapeutics Inc. Any distribution, copying, or disclosure is strictly prohibited unless such disclosure is required by federal regulations or state law. Persons to whom the information is disclosed must know that it is confidential and that it may not be further disclosed by them.

### INVESTIGATOR'S AGREEMENT

I understand that all documentation provided to me by Foghorn Therapeutics Inc. or its designated representative(s) concerning this study that has not been published previously will be kept in strict confidence. This documentation includes the study protocol, Investigator's Brochure (IB), case report forms, and other scientific data.

This study will not commence without the prior written approval of a properly constituted Institutional Review Board (IRB)/Independent Ethics Committee (IEC). No changes will be made to the study protocol without the prior written approval of Foghorn Therapeutics Inc. and the IRB/IEC, except where necessary to eliminate an immediate hazard to the subject.

I have read, understood, and agree to conduct this study as outlined in the protocol and in accordance with the guidelines and all applicable government regulations.

| | | |
|---|---|---|
| Investigator Name (printed) | Investigator Signature | Date |

Investigational site or name of institution and location (printed)

### 2. TABLE OF CONTENTS, LIST OF TABLES, AND LIST OF FIGURES

### 3. LIST OF ABBREVIATIONS AND DEFINITIONS OF TERMS

| Abbreviation | Explanation |
|---|---|
| ADME | absorption, distribution, metabolism, and excretion |
| AE | adverse event |
| AESI | adverse event of special interest |
| AIDS | acquired immunodeficiency syndrome |
| ALP | alkaline phosphatase |
| ALT | alanine aminotransferase |
| AML | acute myeloid leukemia |
| ANC | absolute neutrophil count |
| aPTT | activated partial thromboplastin time |
| ARA | acid-reducing agent |
| AST | aspartate aminotransferase |
| ATP | adenosine triphosphate |
| AUC | area under the plasma concentration-time curve |
| BAF | BRG1/BRM-associated factors (The common forms are canonical BAF, or BAF; non-canonical BAF, or ncBAF; and polybromo BAF, or pBAF) |
| BCRP | breast cancer resistance protein |
| BTM | Bayesian Toxicity Monitoring |
| C1D1 | Cycle 1, Day 1 (The same format is used to designate other cycles and days within the specified cycle) |
| CL | clearance |
| $CL_{int}$ | intrinsic clearance |
| $C_{max}$ | maximum plasma concentration |
| CMML | chronic myelomonocytic leukemia |
| CNS | central nervous system |
| CR | complete remission |
| CRC | chromatin remodeling complex |
| CSR | clinical study report |
| CST | Clinical Study Team |
| CTCAE | Common Terminology Criteria for Adverse Events |
| CYP | cytochrome p450 |
| DDI | drug-drug interaction(s) |
| DLT | dose-limiting toxicity |
| DNA | deoxyribonucleic acid |
| ECG | electrocardiogram |

| Abbreviation | Explanation |
|---|---|
| ECHO | echocardiogram |
| ECOG | Eastern Cooperative Oncology Group |
| eCRF | electronic case report form |
| EFS | event-free survival |
| EOT | End of Treatment |
| F | bioavailability |
| FAS | Full Analysis Set |
| FDA | Food and Drug Administration |
| GCP | Good Clinical Practice |
| GFR | glomerular filtration rate |
| GI | gastrointestinal |
| GLP | Good Laboratory Practice |
| GVHD | graft-versus-host disease |
| H2 blocker | histamine H2-receptor antagonists |
| HBV | hepatitis B virus |
| HCV | hepatitis C virus |
| HIV | human immunodeficiency virus |
| HSCT | hematopoietic stem cell transplantation |
| IB | Investigator's Brochure |
| $IC_{50}$ | half-maximal inhibitory concentration |
| ICH | International Council for Harmonisation |
| IDH | isocitrate dehydrogenase |
| IEC | Independent Ethics Committee |
| INR | international normalized ratio |
| IRB | Institutional Review Board |
| IUD | intrauterine device |
| IUS | intrauterine hormone-releasing system |
| IV | intravenous(ly) |
| IWG | International Working Group |
| LDAC | low-dose cytarabine |
| LVEF | left ventricular ejection fraction |
| MATE | multidrug and toxin extrusion transporter |
| MDS | myelodysplastic syndromes |
| MTD | maximum tolerated dose |

| Abbreviation | Explanation |
|---|---|
| MRD | minimal residual disease |
| NCCN | National Comprehensive Cancer Network |
| NCI CTCAE | National Cancer Institute Common Terminology Criteria for Adverse Events |
| NOAEL | no-observed-adverse-effect level |
| NYHA | New York Heart Association |
| OAT | organic anion transporter |
| OCT | organic cation transporter |
| OS | overall survival |
| PBMC | peripheral blood mononuclear cell |
| PD | pharmacodynamic(s) |
| P-gp | permeability glycoprotein |
| PK | pharmacokinetic(s) |
| PPI | proton pump inhibitor |
| PR | partial remission |
| pRBC | packed red blood cell |
| PS | performance status |
| PT | prothrombin time |
| QD | once daily |
| QTc | corrected QT interval |
| QTcF | corrected QT interval using Fridericia's formula |
| RNA | ribonucleic acid |
| RP2D | recommended phase 2 dose |
| R/R | relapsed or refractory |
| SAE | serious adverse event |
| SAP | statistical analysis plan |
| SC | subcutaneously |
| SmPC | summary of product characteristics |
| $t_{1/2}$ | terminal elimination half-life |
| TEAE | treatment-emergent adverse event |
| TGI | tumor growth inhibition |
| TI | therapeutic index |
| $T_{max}$ | observed time to reach maximum plasma concentration |
| TPN | total parenteral nutrition |
| ULN | upper limit of normal |

| Abbreviation | Explanation |
|---|---|
| UM | uveal melanoma |
| USPI | United States prescribing information |
| UV | ultraviolet |
| WBC | white blood cell |

### 4. SYNOPSIS

| |
|---|
| **Name of Sponsor/Company:**<br>Foghorn Therapeutics Inc. |
| **Name of Investigational Product:**<br>FHD-286 |
| **Name of Active Ingredient:**<br>FHD-286 |
| **Title of Study:**<br>A Phase 1, Multicenter, Open-Label, Dose Escalation Study to Assess the Safety, Tolerability, Pharmacokinetics, Pharmacodynamics, and Clinical Activity of Orally Administered FHD-286, as Monotherapy or Combination Therapy, in Subjects With Advanced Hematologic Malignancies |
| **Study center(s):**<br>Up to approximately 13 study centers in the United States and Europe will participate in this study. |
| **Phase of development:** 1 |
| **Objectives:**<br>**Primary:**<br>• Parts I and II:<br> – To determine the safety and tolerability of FHD-286 when administered as an oral monotherapy in subjects with advanced hematologic malignancies<br> – To identify the recommended phase 2 dose(s) (RP2D[s]) of FHD-286 when administered as monotherapy in subjects with advanced hematologic malignancies<br>• Part III<br> – To determine the safety and tolerability of FHD-286 when administered in combination with either low-dose cytarabine (LDAC) or decitabine (with or without concomitant triazole antifungal agents classified as strong cytochrome P450 [CYP]3A4 inhibitors) in subjects with advanced hematologic malignancies<br> – To identify the RP2D(s) of FHD-286 when administered in combination with either LDAC or decitabine (with or without concomitant triazole antifungal agents classified as strong CYP3A4 inhibitors) in subjects with advanced hematologic malignancies<br>**Secondary:**<br>• Parts I and II:<br> – To determine the pharmacokinetics (PK) of FHD-286 when administered as an oral monotherapy in subjects with advanced hematologic malignancies<br> – To characterize the preliminary clinical activity associated with FHD-286 when administered as monotherapy in subjects with advanced hematologic malignancies<br>• Part III:<br> – To determine the PK of FHD-286 when administered in combination with either LDAC or decitabine in subjects with advanced hematologic malignancies who are *not* receiving triazole antifungal agents classified as strong CYP3A4 inhibitors |

  - To determine the PK of FHD-286 when administered in combination with either LDAC or decitabine in subjects with advanced hematologic malignancies who are receiving triazole antifungal agents classified as strong CYP3A4 inhibitors
  - To characterize the preliminary clinical activity associated with FHD-286 when administered in combination with either LDAC or decitabine in subjects with advanced hematologic malignancies

**Exploratory:**

- Parts I and II:
  - To evaluate the PK/pharmacodynamic (PD) relationship of FHD-286 when administered as monotherapy and changes in peripheral tissue (blood) and bone marrow biomarkers
  - To assess the associations of FHD-286 (when administered as monotherapy) exposure, clinical activity, and safety with PD and response markers in bone marrow and peripheral tissue (blood)
  - To investigate potential predictive and downstream markers of tumor response and/or resistance in bone marrow and peripheral tissue (blood)
  - To evaluate minimal residual disease (MRD)
- Part III:
  - To evaluate the PK/PD relationship of FHD-286 when administered in combination with either LDAC or decitabine and changes in peripheral tissue (blood) and bone marrow biomarkers
  - To assess the associations of FHD-286 (when administered in combination with either LDAC or decitabine) exposure, clinical activity, and safety with PD and response markers in bone marrow and peripheral tissue (blood)
  - To evaluate effects on PK, PD, safety, and clinical activity in subjects who began concomitant treatment with a triazole antifungal agent classified as a strong CYP3A4 inhibitor while on study
  - To investigate potential predictive and downstream markers of tumor response and/or resistance in bone marrow and peripheral tissue (blood)
  - To evaluate MRD

**Methodology:**

*Parts I and II: FHD-286 Monotherapy (Closed to Enrollment)*

This Phase 1, multicenter, open-label, dose escalation study (Parts I and II) is designed to assess the safety, tolerability, PK, PD, and preliminary clinical activity of FHD-286 oral monotherapy in subjects with advanced hematologic malignancies, specifically relapsed or refractory (R/R) acute myeloid leukemia (AML) or R/R myelodysplastic syndromes (MDS). The study aims to determine the RP2D of FHD-286 in subjects with advanced hematologic malignancies. This study will also provide a preliminary assessment of the clinical activity of FHD-286 in subjects with advanced hematologic malignancies.

A daily dosing regimen in 28-day cycles will be investigated during this study. Dose escalation will occur in two parts (Part I and Part II), as described below. Part I will enroll single-subject (n=1) cohorts. Dose escalations of up to 100% will be considered until any 1 (or more) Transition Criteria (Table 1) have been met, then dose escalation will transition to Part II which will enroll 3-subject cohorts in a "3+3" design. This allows for the optimization of dose escalation so that fewer subjects with rapidly progressive advanced hematologic malignancies are treated at potentially subtherapeutic dose levels of FHD-286.

Dose escalation will include approximately 25 to 50 subjects. The recommended starting dose was 5 mg once daily, as determined by the results of the FHD-286 Good Laboratory Practice (GLP)-compliant toxicology studies.

The dose escalation scheme for Part II will follow a modified Fibonacci design (ie, 100%, 67%, 50%, 40%, 33% increments from the previous dose in consecutive escalation cohorts). The safety of dosing will be evaluated by the Clinical Study Team (CST), comprising, at minimum, the Sponsor (Responsible Medical Officer) and Investigators. Real-time assessments of safety, tolerability, PK, and clinical activity will be conducted for each cohort evaluation throughout the study. Additionally, target engagement at each dose level will be evaluated by conducting ribonucleic acid (RNA)- and protein-based assessments on serial blood collections (peripheral blood mononuclear cells [PBMCs]) that measure changes in BRG1/BRM downstream targets in AML blasts. The CST will review the available safety, tolerability, PK, and PD data from Cycle 1 of treatment of each dose cohort to determine whether it is safe to proceed to the next dose level or whether the RP2D and/or maximum tolerated dose (MTD) has been identified. The RP2D is determined by the CST and is the dose regimen identified for continued study based on observed safety, tolerability, PK, PD, and clinical activity data; the RP2D will be further evaluated if and when expansion occurs. The identification of RP2D may or may not include the identification of the MTD (defined as the highest dose that causes dose-limiting toxicities [DLTs] in <2 of 6 subjects); RP2D and MTD may be equivalent. In the absence of observing an MTD, FHD-286 will only be escalated to the minimum or optimal safe and biologically effective dose.

Alternative dose levels and regimens (including, but not limited to, administration of the same total daily dose using different dosing schedules in concurrent groups, intermittent dosing, etc.) will be considered by the CST based on emerging data. Dosing cohorts may be added based on the emerging data. Additional subjects may be enrolled for the replacement of subjects who are not evaluable for the assessment of dose escalation, for evaluation of alternative dosing regimens, or for further exploring safety, PK, PK/PD, or preliminary clinical activity used to guide the selection of the RP2D.

*Part III: Combination Therapy*

This Phase 1, multicenter, open-label, dose escalation study (Part III) is designed to assess the safety, tolerability, PK, PD, and preliminary clinical activity of FHD-286 administered orally in combination with either LDAC or decitabine in subjects with R/R AML, R/R MDS, or R/R chronic myelomonocytic leukemia (CMML) not in blast crisis who have received ≤4 prior lines of systemic anticancer therapy for their disease under study and are appropriate candidates for either LDAC or decitabine. The study aims to determine the RP2D(s), the dose(s) and regimen(s) identified for continued study based on observed safety, tolerability, PK, PD, and preliminary clinical activity, and exposure-response analyses, of FHD-286 in combination with either LDAC or decitabine in subjects with R/R AML, R/R MDS, or R/R CMML not in blast crisis.

FHD-286 will be administered with either LDAC or decitabine:

- Arm A: FHD-286 administered orally once daily (QD) on Days 1 through 28 of all cycles + LDAC 20 mg/m$^2$ administered subcutaneously (SC) QD on Days 1 through 10 of all cycles, in 28-day cycles
  - Group A1: subjects *not* receiving a triazole antifungal agent classified as a strong CYP3A4 inhibitor at the start of study treatment
  - Group A2: subjects receiving a triazole antifungal agent classified as a strong CYP3A4 inhibitor at the start of study treatment
- Arm B: FHD-286 administered orally QD on Days 1 through 28 of all cycles + decitabine 20 mg/m$^2$ administered intravenously (IV) QD on Days 1 through 5 of all cycles, in 28-day cycles

- Group B1: subjects *not* receiving a triazole antifungal agent classified as a strong CYP3A4 inhibitor at the start of study treatment
- Group B2: subjects receiving a triazole antifungal agent classified as a strong CYP3A4 inhibitor at the start of study treatment

Up to approximately 72 subjects will be enrolled in Arm A (FHD-286 + LDAC 20 mg/$m^2$) and up to approximately 72 subjects will be enrolled in Arm B (FHD-286 + decitabine 20 mg/$m^2$) (up to approximately 144 subjects total). Subjects will be assigned to treatment arms at the Investigator's discretion. Dose escalation and RP2D(s) selection will be conducted independently for each of the 4 groups. The FHD-286 dose will be increased in up to 100% increments from the previous dose level ("dose level" refers to dose and regimen) in consecutive escalation cohorts. The proposed FHD-286 dose range is 2.5 mg QD to 10 mg QD for Groups A1 and B1, and 1.5 mg QD to 7.5 mg QD for Groups A2 and B2. For the FHD-286 5 mg QD and 7.5 mg QD dose groups in Groups A2 and B2, interval dosing will be implemented, with a 2.5/5 mg QD dose given on Days 1 through 14 of each cycle, and the 5/7.5 mg QD doses given on Days 15 through 28 of each cycle. Starting at C2D1, participants may continue at the higher dose (ie, transition to a consistent dose) if it is tolerated in the judgment of the Investigator, upon agreement from the Sponsor. Interval dosing may also be implemented for Groups A1 and B1 if agreed upon by the CST. Alternative interval dosing regimens may also be considered.

*Toxicity Grading & DLT Definition*

Toxicity severity will be graded according to the National Cancer Institute Common Terminology Criteria for Adverse Events (CTCAE) version 5.0.

A DLT is defined as any adverse event (AE) that:

- occurs during Cycle 1 (28 days) of treatment (or during the first 42 days of treatment, for participants in Part III who transition from interval dosing during Cycle 1 to a consistent dose at C2D1 employing the highest of the 2 doses administered during Cycle 1 of interval dosing)

  AND
- meets any 1 of the following criteria as determined by the CST:

  **Non-hematologic:**
  - Any Grade 4 Differentiation syndrome toxicity
  - Any Grade ≥3 toxicities not clearly resulting from the underlying advanced hematologic malignancy EXCEPT:
    - Grade 3 fatigue, asthenia, fever, or anorexia (Grade 3 anorexia can be excluded only if it does not result in hospitalization, tube-feeding or use of total parenteral nutrition [TPN])
    - Grade 3 or 4 infection responding to appropriate antimicrobial therapy within 7 days of treatment initiation
    - Grade 3 constipation, nausea, vomiting, or diarrhea not requiring tube feeding, TPN, hospitalization, or prolongation of current hospitalization
    - Grade 3 or 4 tumor lysis syndrome if it is successfully managed and resolves clinically within 7 days without end-organ damage
    - Grade 3 or 4 toxicity associated with lysis or elimination of extramedullary leukemic tissue if it is successfully managed clinically, resolves/improves, and does not result in chronic or permanent end-organ damage while the subject is on study
    - Grade 3 or 4 isolated electrolyte abnormalities resolving to Grade ≤2 within 72 hours with appropriate management
  - Any Hy's law cases

**Hematologic:**

- Any Grade 4 absolute neutrophil count (ANC), febrile neutropenia, or platelets lasting >28 days, in the absence of active disease

In addition, the following will be considered a DLT:

**Other:**

- Any toxicities related to FHD-286 that lead to permanent discontinuation of FHD-286

NOTE: Toxicities meeting any of the above criteria with a clear-cut alternative explanation (eg, due to disease progression) will not be considered DLTs.

All AEs that cannot clearly be determined to be unrelated to FHD-286 or the combination of FHD-286 with LDAC or decitabine will be considered relevant to determining DLTs and will be reviewed by the CST.

The CST also will review any other emergent toxicities that are not explicitly defined by the DLT criteria to determine if any warrant a DLT designation.

***Parts I and II: FHD-286 Monotherapy (Closed to Enrollment)***

*Part I: Single-Subject Dosing Cohorts*

One subject will be enrolled in the first dosing cohort and will be administered 5 mg of FHD-286 once daily for 28 days. The determination of the next dose level will be based on review of all available safety, tolerability, PK, and PD data from Cycle 1 (28 days) by the CST. One new subject will then be enrolled at the next dose level. Dose escalation will continue in this manner until any 1 (or more) of the Transition Criteria are met.

*Transition Criteria*

| | Criteria | Transition Action |
|---|---|---|
| 1 | One subject exhibits any DLT(s) during Cycle 1 of treatment | Cohort will enroll new subjects to reach n=6 |
| 2 | Two subjects exhibit any FHD-286-related non-DLT toxicities that are Grade ≥2 during Cycle 1 of treatment, regardless of dose level, in the opinion of the Investigator | Cohort at the highest dose level at the time Criterion 2 is met will enroll new subject(s) to reach n=3 |

As soon as any 1 (or more) of the above criteria are met, dose escalation will transition to a “3+3” design with multiple ascending doses.

- Part II: “3+3” Dosing Cohorts
- During Part II, each dosing cohort will enroll 3 subjects. As in Part I, subjects in Part II will receive a daily dosing regimen of FHD-286 in 28-day cycle(s).

If there are no DLTs observed after the third evaluable subject in a cohort completes Cycle 1 (ie, the 28-day DLT evaluation period), the study will proceed with dose escalation to the next cohort following review of all available data by the CST. If any 1 of the subjects in a cohort experiences a DLT during Cycle 1 of treatment, then additional subjects will be enrolled in that cohort until that cohort contains 6 subjects. If none of the added subjects experience a DLT during the Cycle 1 DLT-evaluation period (28 days), dose escalation may continue to the next cohort following review of all available data by the CST. If 2 or more subjects in a cohort experience a DLT during Cycle 1, dose escalation will be halted, and the next lower dose level will be declared the MTD. If the MTD cohort included <6 subjects, the cohort will be expanded to include at least 6 total subjects to confirm that no more than 1 out of 6 subjects experience a DLT at that dose. If that is the case, that dose level will be declared the MTD. Alternatively, a dose level in between the dose level exceeding MTD and the previous dose level may be explored and declared MTD if <2 out of 6 subjects experience a DLT at that dose.

Percentage increases for the dose escalations will be determined by the CST based on the available emerging safety, PK, PD, and clinical activity data and will never exceed 100%.

In Part II, if there are multiple subjects in the screening process at the time the third subject within a dose cohort begins treatment, up to 2 additional subjects may be enrolled in that cohort, with approval of the Sponsor.

Once the RP2D has been identified, an additional 6 to 12 subjects may be enrolled at that dose level(s) to confirm the observed safety and tolerability of FHD-286. Enrollment in this cohort will be paused for review for toxicities observed in 2 out of 6, 3 out of 10, or 4 out of 12 subjects.

***Part III: Combination Therapy***

Each dosing cohort will enroll a minimum of 3 evaluable subjects. Dose escalation will proceed according to a standard 3+3 design. Dose escalation will proceed independently for each of the 4 groups in this study: A1, A2, B1, and B2. If there are no DLTs observed after the third evaluable subject in a cohort completes the DLT evaluation period, then the dose of FHD-286 may be escalated and enrollment of the next dose level may begin, after approval by the CST. If 1 of the 3 subjects at a dose level experiences a DLT during the DLT evaluation period, then additional subjects will be enrolled at that dose level until the dose group contains 6 evaluable subjects, or until a second subject experiences a DLT, whichever occurs first. If none of the additional subjects has a DLT (ie, no more than 1 of 6 subjects has a DLT), then the FHD-286 dose may be escalated and enrollment of the next dose level may begin, after approval by the CST. If ≥2 of the evaluable subjects in a dose group experience a DLT, then the previous dose level will be declared the MTD, as long as <2 of 6 subjects experience a DLT at that dose level. Alternatively, a dose level between the non-tolerated dose level and the previous tolerated dose level may be explored and declared the MTD, as long as <2 of 6 subjects experience a DLT at that dose. The decision to escalate the FHD-286 dose will be governed by the CST and based on all available safety, tolerability, PK, and PD data from the DLT evaluation period for each dose cohort, in addition to other relevant data.

The RP2D(s) in each group (A1, A2, B1, and B2) will be determined by the Sponsor with endorsement from the CST, and may or may not be equivalent to the MTD. In the absence of observing an MTD, FHD-286 will only be escalated until the minimum and/or optimal safe and biologically effective dose has been determined. Once the presumptive RP2D(s) has been identified in a given group, an additional 6 to 14 subjects may be enrolled at that FHD-286 dose level(s) in that group to confirm the safety and tolerability of FHD-286 at the RP2D(s) in combination with either LDAC or decitabine. Enrollment of these additional subjects will be paused for review if DLTs are observed in 2 of 6, 3 of 10, or 4 of 14 subjects. Additional stopping criteria will also be considered.

An independent safety monitoring committee will periodically review safety data.

**Intra-subject Dose Escalation:**

*Parts I and II: FHD-286 Monotherapy (Closed to Enrollment)*

Intra-subject dose escalation may be permitted after discussion with the Sponsor, as described further in the protocol.

*Part III: Combination Therapy*

Intra-subject dose escalation will not be allowed.

**General Study Conduct:**

Study subjects will be assessed according to the Schedules of Assessments (Section 10.1). Assessments will support the primary, secondary, and exploratory endpoints of the study.

*Screening:*

Following informed consent (and assent, for subjects <18 years of age), subjects will undergo screening procedures within approximately 28 days prior to the first dose of study treatment to determine eligibility for the study. Screening procedures include medical, surgical, and medication history; complete physical examination including vital signs, height, and weight; echocardiogram (ECHO) (other methods of evaluating left ventricular ejection fraction [LVEF] may be performed according to institutional practice); 12-lead electrocardiograms (ECGs) in triplicate; Eastern Cooperative Oncology Group (ECOG) performance status (PS); and clinical laboratory assessments (hematology, chemistry, coagulation, inflammatory markers, cytokine panel, serum pregnancy test in females of childbearing potential, hepatitis panel, and human immunodeficiency virus [HIV] testing). A bone marrow biopsy sample to determine baseline disease burden is required within approximately 28 days of the administration of the first dose of study treatment (Cycle 1, Day 1 [C1D1]).

*Treatment Period:*

Required clinic visits during the treatment period are outlined in the Schedules of Assessments. The first dose of study treatment will be given on C1D1; these subjects will be required to remain in clinic for 8 hours after the first dose for clinical observation and PK/PD sampling. If the half-life of FHD-286 for any dose group differs significantly from what has been predicted, the duration of time required for subjects to remain in clinic may be adjusted.

For Part III, Arm A, on Days 1 through 5 of Cycle 1, LDAC will be administered at the study site. After the first 5 doses (ie, beginning on C1D6), at the Investigator's discretion and where consistent with institutional, state, and local guidelines and regulations, subjects may self-administer LDAC at home, except on scheduled visit days. For Arm B, decitabine will be administered at the study site on Days 1 through 5 of each cycle.

Subjects may withdraw or be withdrawn from study treatment under the following conditions: disease progression or treatment failure (based on disease response assessments), development of unacceptable toxicity, start of alternative anticancer therapy, or withdrawal of consent/assent (see also "Duration of Treatment," below). All subjects are to undergo an End of Treatment (EOT) assessment within 5 days after the date on which the Investigator decides to discontinue FHD-286 treatment (this date is not required to be equivalent to the date of the last FHD-286 dose).

Safety assessments conducted during the treatment period include physical examination, vital signs, ECOG PS, 12-lead ECGs in triplicate, ECHO (or other means of assessing LVEF), and clinical laboratory assessments (hematology, chemistry, coagulation, inflammatory markers, cytokine panel, pregnancy testing for females of childbearing potential).

All subjects will undergo sampling for PK/PD assessments during the treatment period as outlined in the Schedules of Assessments.

Disease response to treatment will be determined by the Investigators based on modified International Working Group (IWG) criteria, detailed in Section 10.6.

**Parts I and II (FHD-286 Monotherapy)** ***(Closed to Enrollment):*** Subjects will have the extent of their disease assessed by bone marrow biopsies and/or aspirates at Screening (within 28 days of C1D1), approximately every 4 weeks for the first 24 weeks, then approximately every 8 weeks for the next 48 weeks of treatment, as clinically indicated thereafter, and at EOT, independent of dose delays and/or dose interruptions, and/or at any time when a response or progression of disease is suspected.

**Part III (Combination Therapy):** Independent of study treatment delays and/or interruptions, subjects will have the extent of their disease assessed by bone marrow biopsies and/or aspirates at Screening (within 28 days of C1D1); on Cycle 1 Day 15; on Day 1 of Cycles 2 through 6; then on Day 1 of Cycles 8, 10, and 12; then on Day 1 of every third cycle thereafter (Cycle 15, 18, 21, etc); at the EOT Visit; and as clinically indicated.

*Safety Follow-up Visit:*

A Safety Follow-up Visit will occur 28 (±7) days after the EOT Visit. Every effort must be made to perform protocol-specified evaluations unless consent/assent to participate in the study is withdrawn. If a subject's FHD-286 dose is interrupted for 28 days and the subject then discontinues FHD-286 without restarting treatment, the EOT Visit can serve as the Safety Follow-up Visit.

**Part III (Combination Therapy):** For subjects who discontinue FHD-286 for reasons other than disease progression or treatment failure and who continue to be treated with LDAC or decitabine as monotherapy outside the context of this study, a bone marrow biopsy and/or aspirate should be collected at the Safety Follow-up Visit to assess disease response.

*Long-term Follow-up:*

After subjects have discontinued FHD-286, unless consent/assent to participate is withdrawn, they will be contacted by telephone approximately every 2 months to document disease status (unless subject discontinued FHD-286 due to disease progression/treatment failure, and until subject experiences disease progression/treatment failure or starts a new anticancer therapy), receipt and type of subsequent anticancer therapy, and survival status. Long-term follow-up will continue until all subjects have died, withdrawn consent/assent, or are lost to follow-up, or for 2 years after the last subject discontinues treatment with FHD-286, whichever occurs first.

*End of Study:*

End of Study is defined as:

- The time point at which all subjects in all countries have discontinued study treatment and have been followed for survival and disease status assessment for at least 2 years, or have died, been lost to follow-up, or withdrawn consent/assent.

**Number of subjects (planned):**

*Parts I and II: FHD-286 Monotherapy (Closed to Enrollment)*

Approximately 25 to 50 subjects are expected to be treated in Parts I and II of this study.

*Part III: Combination Therapy*

Up to approximately 72 subjects are expected to be enrolled in Arm A and up to approximately 72 subjects are expected to be enrolled in Arm B (up to approximately 144 subjects total).

**Inclusion criteria:**

*Parts I and II: FHD-286 Monotherapy (Closed to Enrollment)*

Subjects must meet all of the following criteria to be enrolled in Parts I and II of the study:

1. Subject must be ≥16 years of age.
2. Subject must have a confirmed diagnosis of an advanced hematologic malignancy, specified as follows:
   - R/R AML (subjects who relapse after transplantation; subjects in second or later relapse; subjects who are refractory to initial induction or reinduction treatment; subjects who relapse within 1 year of initial treatment; subjects who are otherwise considered relapsed or refractory in the opinion of the Investigator). Subjects with AML must have previously failed all prior therapies known to be active for treatment of their diagnosed hematologic disease.
   - R/R MDS. Subjects with MDS must have previously failed treatment with at least 4 cycles of a hypomethylating agent, known to be active for treatment of their diagnosed hematologic disease.
   - Other R/R advanced hematologic malignancies (for example, CMML). Subjects must have no other reasonable therapeutic option in the opinion of the Investigator. Subjects who

fulfill the inclusion/exclusion criteria may be considered on a case-by-case basis, with approval of the Sponsor.

3. Subject or his/her legal guardian (when applicable) must be able to understand and be willing to sign an informed consent and, when applicable, subject must sign an assent form.
4. Subject must be willing and able to comply with scheduled study visits and treatment plans.
5. Subject must be willing to undergo all study procedures (fresh bone marrow biopsy at baseline within 28 days of first dose plus bone marrow biopsies every 4 weeks for the first 24 weeks then every 8 weeks for the next 48 weeks of treatment, as clinically indicated thereafter, and 1 EOT bone marrow biopsy (unless contraindicated due to medical risk; other exceptions to this are at the discretion of the Sponsor), peripheral blood and tissue sampling, and urine sampling during the study.
6. Subject must have an ECOG PS of ≤2.
7. Subject must have a life expectancy of ≥3 months.
8. Subject must have adequate hepatic function as evidenced by:
   - Serum total bilirubin ≤1.5 × upper limit of normal (ULN), unless considered due to leukemic involvement following approval by the study Sponsor.
   - Aspartate aminotransferase (AST), alanine aminotransferase (ALT), and alkaline phosphatase (ALP) ≤3.0×ULN, unless considered due to leukemic involvement following approval by the study Sponsor.
   - Prothrombin time (PT) ≤1.5×ULN or international normalized ratio (INR) ≤1.4
   - Activated partial thromboplastin time (aPTT) ≤1.5×ULN

     Note: Anticoagulation therapy is permitted as long as coagulation parameters are within therapeutic range.
   - No known portal vein thrombosis
9. Subject must have adequate renal function as evidenced by:
   - Creatinine clearance >60 mL/min based on the Cockcroft-Gault glomerular filtration rate (GFR) estimation
10. Subject must have an adequate platelet level, defined as:
    - Platelets $>50\times10^9$/L (transfusions to achieve this level are allowed). Subjects with a baseline platelet count of $\leq50\times10^9$/L due to underlying malignancy are eligible.
11. Subject must have adequate cardiovascular, respiratory, and immune system function as evidenced by the below criteria and in the opinion of the Investigator:
    - LVEF of ≥40% by ECHO (or other means)
12. Subjects must agree to abide by dietary and other considerations required during the study.
13. Timing requirements with respect to prior therapy and surgery are as follows:
    - At least 2 weeks or at least 5 half-lives, whichever is shorter, must have elapsed since administration of the last dose of any prior systemic anticancer therapy. Hydroxyurea is allowed prior to enrollment and after the start of FHD-286 for the control of peripheral leukemic blasts in subjects with leukocytosis (eg, white blood cell [WBC] counts $>30\times10^9$/L). (Subjects must be intolerant to and/or have experienced disease progression on their prior therapy in the opinion of the treating physician.)
    - At least 4 weeks must have elapsed since the last dose of post-transplant calcineurin inhibitors. Exceptions may be made with Sponsor approval, eg, if waiting 4 weeks to elapse before beginning study drug introduces undue risk of disease progression.

- Subjects must be recovered from any clinically relevant effects of any prior surgery.
- At least 2 weeks must have elapsed since the last radiotherapy. Exceptions may be made at the discretion of the Sponsor.

14. Toxicity related to prior therapy must have returned to Grade ≤2 by CTCAE by approximately 14 days prior to study start or be deemed irreversible by the Investigator. Exceptions include alopecia, neuropathy, appropriately controlled endocrine toxicities, and other well controlled/stable toxicities with discussion with the Sponsor.
15. Female subjects must be:
    - postmenopausal, defined as at least 12 months post-cessation of menses (without an alternative medical cause); or
    - permanently sterile following documented hysterectomy, bilateral salpingectomy, bilateral oophorectomy, or tubal ligation or having a male partner with vasectomy as affirmed by the subject; or
    - nonpregnant, nonlactating, and if sexually active having agreed to use a highly effective method of contraception (ie, hormonal contraceptives associated with inhibition of ovulation or intrauterine device [IUD], or intrauterine hormone-releasing system [IUS], or sexual abstinence) from Screening Visit until 90 days after final dose of study drug.

    Note: The potential risk to female fertility posed by FHD-286 is unknown; it is recommended that subjects discuss options for fertility preservation with their doctor prior to study start.
16. Male subjects must have documented vasectomy or if sexually active must agree to use a highly effective method of contraception with their partners of childbearing potential (ie, hormonal contraceptives associated with the inhibition of ovulation or IUD, or IUS, or sexual abstinence) from Screening until 90 days after final dose of study drug. Male subjects must agree to refrain from donating sperm during this time period.

    Note: The potential risk to male fertility posed by FHD-286 is unknown; it is recommended that subjects discuss options for fertility preservation with their doctor prior to study start.

*Part III: Combination Therapy*

Subjects must meet all of the following criteria to be enrolled in Part III of the study:

1. Subject must be ≥16 years of age.
2. Subject must:
    - Have a confirmed diagnosis of R/R AML, R/R MDS, or R/R CMML not in blast crisis

      AND
    - Have received ≤4 prior lines of systemic anticancer therapy for their disease under study; subjects who have received >4 prior lines of systemic anticancer therapy for their disease under study must receive Sponsor approval.

      AND
    - Be an appropriate candidate for treatment with LDAC (Arm A) or decitabine (Arm B)
3. Subject or their legal guardian (when applicable) must be able to understand and be willing to sign an informed consent and, when applicable, subject must sign an assent form.
4. Subject must be willing and able to comply with scheduled study visits and treatment plans.
5. Subject must be willing to undergo all study procedures (including bone marrow biopsies, peripheral blood sampling, and urine sampling), unless contraindicated due to medical risk.
6. Subject must have an ECOG PS of ≤2.

7. Subject must have a life expectancy of ≥3 months.
8. Subject must have adequate hepatic function as evidenced by:
   - Serum total bilirubin ≤1.5× ULN, unless considered due to advanced hematologic malignancy involvement or Gilbert syndrome, following approval by the study Sponsor
   - AST, ALT, and ALP ≤3.0×ULN, unless considered due to advanced hematologic malignancy involvement, following approval by the study Sponsor
   - PT ≤1.5×ULN or INR ≤1.4
   - aPTT ≤1.5×ULN

     Note: Sponsor approval is required for a subject receiving anticoagulation therapy for treatment of a stable medical condition whose coagulation parameters are within therapeutic range.
   - No known portal vein thrombosis
   - No history of azole-induced liver dysfunction (if being considered for participation in Group A2 or B2 [subjects receiving a triazole antifungal agent classified as a strong CYP3A4 inhibitor])
9. Subject must have adequate renal function as evidenced by:
   - GFR ≥60 mL/min (based on a contemporary, widely accepted, and clinically applicable equation that estimates glomerular filtration rate or a measure of glomerular filtration rate)
10. Subject must have a WBC count ≤$20 \times 10^9$/L; treatment with a stable dose of hydroxyurea or other cytoreductive agent (eg, cytarabine) to achieve this count is allowed.
11. Subject must have adequate cardiovascular, respiratory, and immune system function as evidenced by the below criteria and in the opinion of the Investigator:
    - LVEF of ≥40% by ECHO (or other means)
12. Subject must agree to abide by dietary and other considerations required during the study (Section 8.4).
13. Timing requirements with respect to prior therapy and surgery are as follows:
    - Prior systemic anticancer therapy (including investigational drugs):
      - Not a small molecule inhibitor: At least 2 weeks or at least 5 half-lives, whichever is shorter, must have elapsed since administration of the last dose.
      - Small molecule inhibitor (eg, FLT3 kinase inhibitor, isocitrate dehydrogenase [IDH]1/IDH2 inhibitor), *except venetoclax*: At least 72 hours must have elapsed since administration of the last dose.
      - Venetoclax: At least 2 weeks must have elapsed since administration of the last dose.
      - Treatment with hydroxyurea (or other cytoreductive agent) is allowed during the washout period for blast control (as per inclusion criterion 10).
    - Post-transplant calcineurin inhibitors: At least 4 weeks must have elapsed since administration of the last dose.
    - Radiotherapy: At least 2 weeks must have elapsed since the last radiotherapy.
    - Surgery: Subject must be recovered from any clinically relevant effects of any prior surgery.
14. Toxicity related to prior therapy must have returned to Grade ≤2 by CTCAE by approximately 14 days before the start of study treatment or be deemed irreversible and stable by the Investigator. Exceptions include alopecia, neuropathy, appropriately controlled endocrine toxicities, and other well controlled/stable toxicities with discussion with the Sponsor.

15. Female subjects must be:
    - postmenopausal, defined as at least 12 months post-cessation of menses (without an alternative medical cause); or
    - permanently sterile following documented hysterectomy, bilateral salpingectomy, bilateral oophorectomy, or tubal ligation, or, if sexually active with male partners, these partners must be azoospermic (vasectomized or due to a medical cause), as affirmed by the subject; or
    - nonpregnant, nonlactating, and if sexually active with fertile male partners, having agreed to use a highly effective method of contraception (ie, hormonal contraceptives associated with inhibition of ovulation or IUD, or IUS, or sexual abstinence) from Screening Visit until the latest of the following:
        - All subjects: 90 days after the final dose of FHD-286
        - Subjects in Arm A (FHD-286 + LDAC): 6 months after the final dose of LDAC
        - Subjects in Arm B (FHD-286 + decitabine): 6 months after the final dose of decitabine

    Note: The potential risk to female fertility posed by FHD-286 is unknown; it is recommended that subjects discuss options for fertility preservation with their doctor before the start of study treatment.

    Note: Subjects in Arm B: Because of the possibility of infertility as a consequence of decitabine therapy, subjects should seek consultation regarding oocyte cryopreservation before the initiation of treatment.
16. Male subjects must have documented azoospermia (vasectomized or due to a medical cause), or, if fertile and sexually active, must agree to use a highly effective method of contraception with their partners of childbearing potential (ie, hormonal contraceptives associated with the inhibition of ovulation or IUD, or IUS, or sexual abstinence) from Screening until the latest of the following:
    - All subjects: 90 days after the final dose of FHD-286
    - Subjects in Arm A (FHD-286 + LDAC): 6 months after the final dose of LDAC
    - Subjects in Arm B (FHD-286 + decitabine): 90 days after the final dose of decitabine

    Male subjects must agree to refrain from donating sperm during this time period.

    Note: The potential risk to male fertility posed by FHD-286 is unknown; it is recommended that subjects discuss options for fertility preservation with their doctor before the start of study treatment.

    Note: Subjects in Arm B: Because of the possibility of infertility as a consequence of decitabine therapy, subjects should seek consultation regarding conservation of sperm before the initiation of treatment.

**Exclusion criteria:**

*Parts I and II: FHD-286 Monotherapy (Closed to Enrollment)*

Subjects who meet any of the following criteria will not be enrolled in Parts I and II of the study:

1. Subject (or parent or legal guardian, when applicable) is unable to provide informed consent (or assent, when applicable) and/or to follow protocol requirements.
2. Subject has undergone hematopoietic stem cell transplant (HSCT) within 60 days of the first dose of FHD-286, or subject has clinically significant graft-versus-host disease (GVHD). The use of a stable dose of oral steroids and/or immunosuppressive therapy post-HSCT is permitted with Sponsor approval.

3. Subject has clinical symptoms suggesting active central nervous system (CNS) leukemia or known CNS leukemia. Evaluation of the cerebrospinal fluid is only required if there is a clinical suspicion of CNS involvement by leukemia during screening.
4. Subject has an immediately life-threatening, severe complications of leukemia, such as uncontrolled bleeding, pneumonia with hypoxia or shock, and/or disseminated intravascular coagulation.
5. Subject has other malignancy which may interfere with the diagnosis and/or treatment of advanced hematologic malignancies.
6. Subject has active hepatitis B virus (HBV) or hepatitis C virus (HCV) infections; subjects with a sustained viral response to HCV treatment or immunity to prior HBV infection will be permitted. Subject has known positive HIV antibody results, or acquired immunodeficiency syndrome (AIDS)-related illness; subjects with CD4+ T-cell counts ≥350 cells/µL will be permitted, as will subjects who have not had an AIDS-related illness within the past 12 months.
7. Subject has an active severe infection that requires anti-infective therapy or has an unexplained temperature of >38.5°C during screening visits or on their first day of study drug administration (at the discretion of the Investigator, subjects with tumor fever may be enrolled).
8. Subject has an uncontrolled intercurrent illness.
9. Subjects with corrected QT interval (QTc) using Fridericia's formula (QTcF) >470 msec or other factors that increase the risk of QTc prolongation or arrhythmic events (eg, heart failure, hypokalemia, family history of long QT interval syndrome) including heart failure that meets New York Heart Association (NYHA) class III and IV definitions (see Appendix 15.3) are excluded. Subjects with bundle branch block and a prolonged QTc should be reviewed by the Sponsor for potential inclusion.
10. Subject has any other medical or psychological condition, deemed by the Investigator to be likely to interfere with a subject's ability to sign informed consent/assent, cooperate, or participate in the study.
11. Subject has known allergies or hypersensitivities to components of the FHD-286 formulation.
12. Subject is unable to tolerate the administration of oral medication or has gastrointestinal (GI) dysfunction that could interfere with absorption of FHD-286 (eg, ulcerative disease, uncontrolled nausea, vomiting, diarrhea, malabsorption syndrome, partial bowel resections).
13. Subject is receiving any other investigational agents.
14. At least 2 weeks or 5 half-lives, whichever is shorter, must have elapsed since last administration of a prior investigational drug. Exceptions include participation in any observational or nontherapeutic clinical trials.
15. Subject is on medications that are strong cytochrome P450 (CYP) 3A inhibitors, are strong CYP3A inducers, or are sensitive CYP3A substrates with narrow therapeutic indices (TIs; see Appendix 15.4). Exceptions may be made for therapy in the case of life-threatening infections, for example a triazole anti-fungal agent to reduce the risk of invasive fungal infections, at the discretion of the Sponsor.
16. Subject is on medications with narrow TIs that are sensitive permeability glycoprotein (P-gp) or breast cancer resistance protein (BCRP) substrates and are administered orally such as digoxin (see Appendix 15.5), or on medications that are strong inhibitors of P-gp or BCRP (see Appendix 15.6)
17. Administration of proton pump inhibitors (PPI) should be stopped or switched to another acid-reducing agent (ARA; eg, antacids or histamine H2-receptor antagonists [H2 blockers]) 7 days

before administration of study drug. In the event that it is medically necessary to dose PPIs concomitantly with FHD-286, this may be permitted with Sponsor approval. See Section 9.12.2 for further detail.

18. Subject is requiring clinically significant or increasing doses of systemic steroid therapy or any other systemic immunosuppressive medication. The use of a stable dose of systemic steroids and/or immunosuppressive medication is permitted with Sponsor approval. Local or targeted steroid and immunosuppressive therapies (eg, inhaled or topical steroids) are acceptable. Appropriate steroid replacement to manage endocrine toxicities resulting from prior anticancer systemic therapy is permitted.
19. Subject has undergone any prior treatment with a BRG1/BRM inhibitor.
20. Subject is pregnant or breastfeeding or is planning to become pregnant within 1 year of study start. Subject is a woman or man of childbearing capabilities who is unwilling to use effective contraception.

*Part III: Combination Therapy*

Subjects who meet any of the following criteria will not be enrolled in Part III of the study:

1. Subject (or parent or legal guardian, when applicable) is unable to provide informed consent (or assent, when applicable) and/or to follow protocol requirements.
2. Subject:
   - Has undergone chimeric antigen receptor T cell therapy or HSCT within 60 days of the first dose of study treatment

     OR
   - Has clinically significant GVHD
     - Clinically significant GVHD is defined as signs or symptoms of acute or chronic GVHD Grade >0, per Mount Sinai Acute GvHD International Consortium or National Institutes of Health criteria, respectively, within 4 weeks before the first dose of study treatment.
3. Subject has evidence (or suspicion) of extramedullary involvement, unless approved by Sponsor.
4. Subject has an immediately life-threatening, severe complication(s) of advanced myeloid malignancy, such as uncontrolled bleeding, pneumonia with hypoxia or shock, and/or disseminated intravascular coagulation.
5. Subject has other malignancy that may interfere with the diagnosis and/or treatment of advanced hematologic malignancies.
6. Subject has active HBV or HCV infections; subjects with a sustained viral response to HCV treatment or immunity to prior HBV infection will be permitted. Subject has known positive HIV antibody results, or AIDS-related illness; subjects with CD4+ T-cell counts ≥350 cells/µL will be permitted, as will subjects who have not had an AIDS-related illness within the past 12 months.
7. Subject has an active severe infection that requires anti-infective therapy or has an unexplained temperature of >38.5°C during screening visits or on their first day of study treatment (at the discretion of the Investigator, subjects with tumor fever may be enrolled).
8. Subject has an uncontrolled intercurrent illness.
9. Subject has QTcF >470 msec or other factors that increase the risk of QTc prolongation or arrhythmic events (eg, heart failure, hypokalemia, family history of long QT interval syndrome) including heart failure that meets NYHA class III and IV definitions (see

Appendix 15.3) are excluded. Subjects with QTcF >470 msec and bundle branch block and/or pacemaker rhythm may be enrolled after approval by the Sponsor.

10. Subject has any other medical or psychological condition, deemed by the Investigator to be likely to interfere with a subject's ability to sign informed consent/assent, cooperate, or participate in the study.
11. Subject has known allergies or hypersensitivities to:
    - All subjects: components of the FHD-286 formulation (refer to the FHD-286 Investigator's Brochure [IB])
    - Arm A (FHD-286 + LDAC): cytarabine or any of the excipients (refer to the approved drug label [eg, United States prescribing information (USPI), summary of product characteristics (SmPC)])
    - Arm B (FHD-286 + decitabine): decitabine or any of the excipients (refer to the approved drug label [eg, USPI, SmPC])
12. Subject is unable to tolerate the administration of oral medication or has GI dysfunction that would preclude adequate absorption, distribution, metabolism, or excretion of study drug.
13. Subject is receiving any other anticancer investigational agents. Investigational agents to treat non-cancer indications may be permitted with Sponsor approval.
14. Removed in protocol version 5.0.
15. Subject is on medications classified as:
    - Strong CYP3A4 inhibitors; see also Section 9.12.2 and Appendix 15.4
        - Exception: Triazole antifungal agents, including those classified as strong CYP3A4 inhibitors, are permitted. Subjects receiving triazole antifungal agents classified as strong CYP3A4 inhibitors at the start of study treatment will be assigned to Group A2 or B2. See Section 9.12.5.
    - Strong CYP3A inducers; see also Section 9.12.2 and Appendix 15.4
    - Sensitive CYP3A substrates with narrow TIs; see also Section 9.12.2 and Appendix 15.4
        - Stable doses of immunosuppressant medications that are sensitive CYP3A substrates may be permitted with Sponsor approval.
16. Subject is on medications with narrow TIs that are sensitive P-gp or BCRP substrates and are administered orally such as digoxin (see Appendix 15.5), or on medications classified as strong inhibitors of P-gp or BCRP (see Appendix 15.6).
17. Administration of PPI should be stopped or switched to another ARA (eg, antacids or H2 blockers) 7 days before administration of FHD-286. In the event that it is medically necessary to dose PPIs concomitantly with FHD-286, this may be permitted with Sponsor approval. See Section 9.12.2 for further detail.
18. Subject is requiring clinically significant or increasing doses of systemic steroid therapy or any other systemic immunosuppressive medication. The use of a stable dose of systemic steroids and/or immunosuppressive medication is permitted with Sponsor approval. Local or targeted steroid and immunosuppressive therapies (eg, inhaled or topical steroids) are acceptable. Appropriate steroid replacement to manage endocrine toxicities resulting from prior anticancer systemic therapy is permitted. See exclusion criterion 15 for exclusions regarding medications classified as strong CYP3A inducers or sensitive CYP3A substrates with narrow TIs.
19. Subject has undergone any prior treatment with a BRG1/BRM inhibitor.
20. Subject is pregnant or breastfeeding or is planning to become pregnant within 1 year of the start of study treatment.

**Study intervention, dosage, and mode of administration:**

FHD-286 will be supplied as 1.5 mg, 2.5 mg, and 5 mg strength capsules to be administered orally, fasted (as defined in Section 9.5.1).

*Part III: Combination Therapy*

Treatment will be administered on an outpatient basis; treatment may be administered in the inpatient setting as clinically indicated.

All arms: On days when FHD-286 is administered in combination with LDAC or decitabine, FHD-286 must be administered before the combination agent.

Arm A: LDAC (cytarabine 20 $mg/m^2$) will be administered SC QD on Days 1 through 10 of all cycles. On Days 1 through 5 of Cycle 1, LDAC will be administered at the study site. After the first 5 doses (ie, beginning on C1D6), at the Investigator's discretion and where consistent with institutional, state, and local guidelines and regulations, subjects may self-administer LDAC at home, except on scheduled visit days.

Arm B: Decitabine 20 $mg/m^2$ will be administered IV on Days 1 through 5 of all cycles.

**Duration of treatment:**

Subjects have the right to withdraw from study treatment at any time for any reason. Subjects may withdraw or be withdrawn from study treatment under the following conditions:

- Withdraws consent/assent for study treatment
- Experiences unacceptable toxicity. Such subjects will discontinue treatment though will remain in the study for long-term follow-up.
- The Investigator removes the subject from the study in the best interests of the subject for any reason, including any medical condition that, in the opinion of the Investigator, would put the subject at risk for continuing treatment.
- Development of disease progression or treatment failure, based on disease response assessments (subjects who are, in the opinion of the Investigator, benefitting from treatment may be allowed to continue on study treatment with the approval of the Sponsor)
  - In Part III (combination therapy), subjects who discontinue LDAC or decitabine for reasons other than treatment failure or progressive disease may continue FHD-286 monotherapy until any of the withdrawal criteria are met.
- Development of an intercurrent medical condition or need for a concomitant medication that precludes further participation in the study or that, in the opinion of the Investigator, would pose a safety risk to the subject
- Start of alternative anticancer therapy
- Noncompliance with protocol requirements
- Becomes pregnant
- Lack of efficacy (failure to achieve a partial remission [Section 10.6] or better by 6 months on treatment)

Subjects have the right to withdraw from the study at any time for any reason. Subjects must be withdrawn from the study if the subject:

- Withdraws consent/assent
- Is lost to follow-up
- Dies

**Reference therapy, dosage, and mode of administration:**

Not applicable

**Criteria for evaluation:**

**Safety:**

Monitoring of AEs, including determination of DLTs, serious AEs (SAEs), adverse events of special interest (AESIs), and AEs leading to discontinuation; clinical laboratory parameters; physical examination findings; ECGs; ECHO (or other means of assessing LVEF); vital signs; and ECOG PS

**Pharmacokinetics and Pharmacodynamics:**

Serial blood sampling for determination of concentration-time profiles of FHD-286

Blood and bone marrow for PD sampling will be collected according to the time points in the Schedules of Assessments to assess target engagement in peripheral tissues.

**Clinical Activity:**

Serial blood and bone marrow sampling to determine response to treatment based on modified IWG response criteria in AML and MDS, or other response criteria appropriate for the subject's advanced hematologic malignancy (Section 10.6)

Long-term follow-up for 2 years after the last subject has discontinued treatment with FHD-286

**Statistical methods:**

**Sample size considerations:**

*Parts I and II: Monotherapy (Closed to Enrollment)*

The sample size for the study will depend on the number of dose cohorts required and the incidence of DLTs required to determine the RP2D. It is estimated that approximately 25 to 50 subjects will be enrolled, assuming 1 subject (Part I) and 3 subjects (Part II) per cohort and a starting dose of 5 mg daily. Once the RP2D has been identified, an additional 6 to 12 subjects may be enrolled at that dose level(s) to confirm the observed safety and tolerability of FHD-286. Enrollment in this cohort will be paused for review for toxicities observed in 2 out of 6, 3 out of 10, or 4 out of 12 subjects.

*Part III: Combination Therapy*

The sample size will depend on the number of dose levels required and the incidence of DLTs required to determine the RP2D(s). It is estimated that up to approximately 72 subjects will be enrolled in Arm A (FHD-286 + LDAC) and up to approximately 72 subjects will be enrolled in in Arm B (FHD-286 + decitabine) (up to approximately 144 subjects total).

Once the RP2D(s) has been identified for a group (A1, A2, B1, or B2), an additional 6 to 14 subjects may be enrolled at that dose level(s) in that group to confirm the observed safety and tolerability of FHD-286 in combination with either LDAC or decitabine. Enrollment of these subjects will be paused for review if DLTs are observed in 2 of 6, 3 of 10, or 4 of 14 subjects. Additional stopping criteria will also be considered.

**Analysis populations:**

The Safety Analysis Set/Full Analysis Set (FAS) is defined as all subjects who were enrolled and received at least 1 dose of FHD-286. Subjects will be classified according to dose levels initially assigned. The Safety Analysis Set will be the primary set for the analysis of safety data. The FAS will be used for the summary of demographics, baseline characteristics, subject disposition, and analyses of all efficacy data.

PK analyses will be performed using the PK Analysis Set, defined as all subjects who have at least 1 blood sample providing evaluable PK data for FHD-286.

**Independent safety monitoring committee analyses:**

Preplanned analyses of safety will be provided to the independent safety monitoring committee as specified in the committee charter.

**Data presentation/descriptive statistics:**

Statistical analyses will be primarily descriptive in nature. Data will be summarized by FHD-286 dose level initially assigned. FHD-286 monotherapy (Parts I and II) and combination therapy (Part III) will be summarized separately. For Part III, all groups (A1, A2, B1, and B2) will be summarized separately. For analysis of clinical activity, subjects with different indications will be summarized separately. Tabulations will be produced for appropriate disposition, demographic, baseline, exposure, safety, PK, PD, and disease response parameters and will be presented by dose level and overall. Categorical variables will be summarized by frequency distributions (number and percentages of subjects) and continuous variables will be summarized by descriptive statistics (mean, standard deviation, median, minimum, and maximum).

| |
|---|
| AEs will be summarized by Medical Dictionary for Regulatory Activities System Organ Class and Preferred Term. All AEs will be listed. Only treatment-emergent adverse events will be summarized (and will be referred to as AEs). Separate tabulations will be produced for all AEs, treatment-related AEs (those considered by the Investigator to be at least possibly related to study treatment), SAEs, dose modifications due to AEs, and AEs of at least Grade 3 severity. By-subject listings will be provided for AEs leading to death, SAEs, DLTs, and AEs leading to discontinuation of treatment. AESIs will be summarized.<br>Descriptive statistics will be provided for clinical laboratory parameters, ECG intervals, ECHOs (or other means) for LVEF, and vital signs data, presented as both actual values and changes from baseline relative to each on-study evaluation and to the last evaluation on study. Shift analyses will be conducted for laboratory parameters and ECOG PS.<br>PK data will be summarized and listed in tabular format by each FHD-286 dose level and, where appropriate, for the entire PK Analysis Set, using descriptive statistics. PK data will also be displayed graphically as appropriate. The potential relationship between PK and safety, efficacy, and PD will be explored with descriptive and graphical methods. FHD-286 exposure (area under the concentration versus time curve and maximum concentration) will be descriptively compared between Groups A1 and A2, and between Groups B1 and B2.<br>Response to treatment as assessed by the site Investigators using applicable response criteria will be tabulated. Point estimates and 2-sided 90% Cis for response rates will be summarized and overall response will be summarized by best overall response categories.<br>Time-to-event endpoints will be estimated using Kaplan-Meier methods, if appropriate. |

### 5. INTRODUCTION

#### 5.1. Chromatin Regulatory System and Cancer

##### 5.1.1. Chromatin Remodeling Complexes

The entire human genome is composed of approximately $3 \times 10^9$ base pairs of deoxyribonucleic acid (DNA), which needs to fit in an orderly way into the nucleus of each human cell. To accomplish this, the DNA winds around a core of proteins called histones to form what is known as a nucleosome; multiple nucleosomes then cluster further to form even more densely packed chromatin, which is organized into chromosomes. Before DNA can be transcribed into ribonucleic acid (RNA) and then translated into protein, chromatin must be “unpacked” to allow the transcription machinery access to the DNA. To handle this unpacking, cells have evolved molecular machines known as adenosine triphosphate (ATP)-dependent chromatin remodeling complexes (CRCs). These complexes are required to locate and unpack specific regions of chromatin to orchestrate and allow for appropriate gene expression. A major family of CRCs with relevance to human cancer as well as other diseases is the mSWI/SNF (mammalian Switch/Sucrose-Nonfermentable), also referred to as the BAF (Brg/Brahma-associated factors) family of complexes (Ho and Crabtree 2010).

##### 5.1.2. Relevance of BAF Complexes in Cancer

BAF protein complexes are large, multi-subunit molecular machines that utilize the energy generated by ATP hydrolysis to catalyze the remodeling of nucleosomes to regulate gene transcription and DNA repair (Hodges, et al 2016). The products of 29 genes are modularly assembled into 3 distinct BAF complexes: canonical BAF (cBAF), polybromo-associated BAF (PBAF), and non-canonical BAF (ncBAF). Collectively, the genes encoding BAF complex subunits are mutated in approximately 20% of cancers, rendering BAF complexes among the most frequently mutated targets in cancer (Kadoch, et al 2013). At their catalytic core, each of these 3 complexes contains 1 of 2 closely related DNA-dependent ATPases belonging to the SNF2 superfamily: BRG1 and BRM (also called SMARCA4 and SMARCA2, respectively; see Figure 1).

**Figure 1: Biochemical Subunit Composition of Mammalian cBAF, PBAF, and ncBAF Complexes**

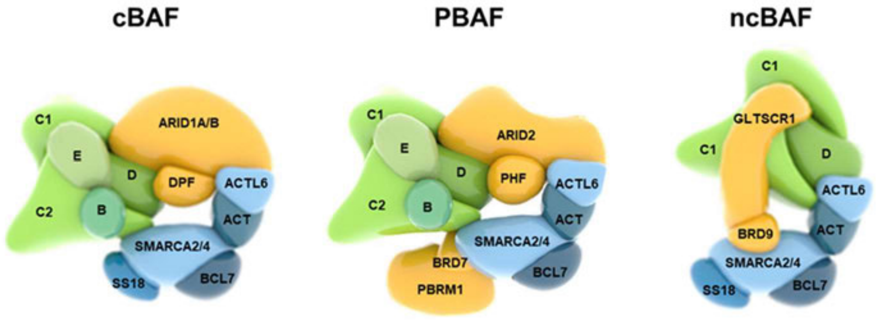

Source: Centore, et al (2020).
Legend for Subunit Paralog Families:

| | | | | |
|---|---|---|---|---|
| B: SMARCB1 | C1: SMARCC1 | C2: SMARCC2 | D: SMARCD1/D2/D3 | E: SMARCE1 |
| DPF: DPF1/2/3 | PHF: PHF10 | ACT: B-Actin | ACTL6: ACTL6A/B | BCL7: BCL7A/B/C |
| GLITSCR1/1L | SS18/18L1 | | | |

The enzymatic activity of BRG1 or BRM is essential for the mechanical movement of nucleosomes by BAF complexes. Such chromatin remodeling activity is critical for the establishment and maintenance of chromatin landscapes conducive to promoting lineage- as well as disease-specific transcriptional programs. The various BAF complexes collaborate with transcription factors to establish cell-type specific gene expression programs (Lessard and Crabtree 2010). Thus, as evidenced by functional genomics screening campaigns, many cancer cell lines of various lineages depend on BRG1 or BRM for proliferation and survival (DepMap 2020; Meyers, et al 2017; Tsherniak, et al 2017). Such dependency on BRG1 or BRM in many types of cancer renders these ATPases potential targets for therapeutic intervention.

The BAF complex has frequently been implicated as a potential therapeutic target in cancer due to its high mutation rates across various tumor types and its established roles in DNA damage repair and in the regulation of lineage genes essential to the dedifferentiated state of many cancers (Alfert, et al 2019; Alver, et al 2017). Additionally, BRG1 expression is widely upregulated across many tumors relative to their normal tissue counterparts and is particularly elevated in indications such as breast cancer, prostate cancer, acute myeloid leukemia (AML), and uveal melanoma (UM). This suggests that BAF may play a central role in the development and survival of these tumors and that targeting the BAF complex may have therapeutic potential (Buscarlet, et al 2014; Muthuswami, et al 2019; Wu, et al 2016). Recent publications have demonstrated that BAF regulates key genes required by UM and AML tumor models (Rago, et al 2020; Shi, et al 2013) and that transcriptional modulation of these genes through BAF disruption has a significant impact on tumor survival, proliferation, and stem cell-like potential. Knockdown of BRG1, or modulation of its key transcriptional targets, has been shown to induce differentiation in leukemic stem cells and to reduce disease burden and blast-like cells in AML in vivo models (McKenzie, et al 2019; Shi, et al 2013). This further suggests that a BAF inhibitor may have therapeutic potential in these indications.

#### 5.2. Advanced Myeloid Malignancies

AML is a heterogeneous group of cancers that arise from blood cell precursors in the bone marrow. In AML, malignant transformation of myeloid precursor cells leads to their uncontrolled proliferation and inability to differentiate from immature blast cells into mature blood cells. As a result, the bone marrow becomes overcrowded and is unable to produce adequate amounts of essential mature blood cells. This leads to the classic clinical manifestations of AML: bleeding and bruising (due to low platelets), infections (due to low neutrophils), and anemia (due to low red blood cell count), among others.

Though AML can occur in patients of all ages (from infants to the elderly), the median age at diagnosis is approximately 68 years and the risk of developing the disease increases with age. In the United States, there are approximately 20,000 new cases of AML diagnosed each year and the disease accounts for 1% to 2% of adult cancer deaths (SEER 2020).

Myelodysplastic syndromes (MDS) are a group of clonal disorders of hematopoiesis. Approximately one-third of patients with MDS will develop AML, known as secondary AML.

Chronic myelomonocytic leukemia (CMML) is a heterogenous clonal disorder of bone marrow stem cells, in which monocytosis is a major defining feature (SEER). The incidence of CMML is 12.8 per 100,000 persons per year. Transformation to AML occurs in approximately 15% to 30% of cases. Prognosis is generally poor and there is a limited availability of effective therapies.

For decades, the standard of care for AML has been intensive cytotoxic chemotherapy (namely, cytarabine plus an anthracycline), with or without hematopoietic stem cell transplantation (HSCT). Cure of disease is achieved in only approximately one-third of patients, 5-year overall survival (OS) is approximately 28%, and the drugs used are highly toxic and difficult to tolerate (SEER 2020). Furthermore, due to the toxicity of intensive chemotherapy, many AML patients are not eligible for the standard treatment regimens due to factors such as older age and comorbidities.

More recently, molecularly targeted therapeutic agents have become available for specific subsets of AML patients, largely as supplements to chemotherapy. These agents have not achieved significant increases in cure rates, but they have demonstrated improved survival and quality of life in the AML patient segments that they target (Lai, et al 2019).

Venetoclax, a BCL-2 inhibitor, was approved in 2020 in combination with azacitidine, or decitabine, or low-dose cytarabine for the treatment of newly diagnosed AML in adults aged 75 years or older, or who have comorbidities that preclude use of intensive induction chemotherapy, and is associated with improved response rates and better OS, compared with earlier therapies (Kayser and Levis 2022; Venclexta (venetoclax tablets) for oral use June 2022).

In patients who are older and/or those who are not candidates for or who decline intensive chemotherapy, low-intensity decitabine or low-dose cytarabine may be an option (NCCN 2023). In a study that included patients with AML and high-risk MDS who were primarily >60 years old and unfit for intensive chemotherapy, LDAC resulted in an 18% complete remission (CR) rate (Burnett, et al 2007). In a Phase 2 study in patients with previously untreated AML who were >60 years old, the overall CR rate with decitabine was 24% (Cashen, et al 2010). LDAC or decitabine may also be given as a less intensive treatment option in patients with relapsed or refractory (R/R) AML (NCCN 2023).

The treatment of CMML varies and is determined based on subtype. First-line treatment options include supportive therapy aimed at correcting cytopenias, hypomethylating agents (38% to 70% response rate), cytoreductive agents such as hydroxyurea (60% response rate), chemotherapy (treatment with cytarabine and topotecan achieved a 44% CR with a median OS of 41 weeks), ruxolitinib (a recent Phase 2 trial demonstrated a 17% overall response rate, a 66% clinical benefit rate, and a median OS of 24 months), and allogeneic HCT (an option in only a limited subset of patients and the only therapy that may lead to cure) (NCCN 2022; Onida, et al 2013; Padron, et al 2022). In the second line, supportive therapy and enrollment in clinical trials are recommended.

Given the low cure rate and poor overall survival and quality of life in the AML, MDS, and CMML population, as well as the highly toxic nature of the standard of care for the majority of patients, there remains a high degree of unmet need. There is a clear demand for continued efforts to develop drugs with lower toxicity profiles that can achieve improved survival, improved quality of life, and higher cure rates.

#### 5.3. Study Drug

##### 5.3.1. FHD-286

###### 5.3.1.1. Overview

The investigational drug FHD-286 is a potent, selective, allosteric, and orally available small molecule inhibitor of the enzymatic activity of both BRG1 and BRM.

FHD-286 was evaluated in a series of nonclinical in vitro and in vivo pharmacology studies to characterize the potency and selectivity of FHD-286 and its primary pharmacodynamics (PD). In these studies, FHD-286 demonstrated tumor inhibition in AML cell proliferation assays and xenograft model studies in mice. FHD-286 in combination with either cytarabine or decitabine resulted in enhanced antitumor effects in vitro and in xenograft model studies in mice.

The results of the nonclinical pharmacokinetic (PK) studies, including both single-dose and multiple dose evaluations in BALB/c nude mice, Sprague Dawley rats, Wistar Han rats, beagle dogs, and cynomolgus monkeys showed that FHD-286 has an acceptable absorption, distribution, metabolism, and excretion (ADME) profile for continued evaluation and development in humans.

The toxicity profile was evaluated in vitro in the bacterial reverse mutation assay and in vivo in Wistar Han rats and beagle dogs, including Good Laboratory Practice (GLP)-compliant, 28- and 91-day repeat-dose studies. Based on the results of the nonclinical toxicology program, FHD-286 has an acceptable safety profile for development in humans.

###### 5.3.1.2. Summary of Nonclinical Information

Details of the nonclinical data for FHD-286 are provided in the FHD-286 Investigator's Brochure (IB). A summary of key information is provided below.

**Nonclinical Pharmacology**

FHD-286 is a potent and selective BRG1 and BRM inhibitor. Cross-species sequence conservation assessments demonstrated that the ATPase domains for BRG1 and BRM are 100% identical across human, monkey, dog, rat, and mouse. FHD-286 is therefore expected to inhibit BRG1 and BRM ATPase activity across these 5 mammalian species. In vitro studies to assess potency and selectivity demonstrate that FHD-286 inhibits BRG1 and BRM enzyme ATPase activity with geometric mean half-maximal inhibitory concentration ($IC_{50}$) values of 6.5 nM and 4.5 nM, respectively. In contrast, FHD-286 did not inhibit the ATPase activity of the highly related chromodomain helicase DNA binding protein 4 enzyme (CHD4; the CHD4 ATPase domain is homologous to that of BRG1 and BRM) at concentrations up to 200 μM. Further profiling against 249 human ATPases detected little off-target impact of FHD-286. Other in vitro cellular studies show that FHD-286 potently inhibits BRG1 and BRM-dependent transcription at nanomolar concentrations, while having no significant effect on general BRG1/BRM-independent transcription. In a study conducted using the research tool compound FHT-1015, which is also a selective inhibitor of BRG1 and BRM, the sensitivity of a broad panel of various cancer cell subtypes to BRG1/BRM inhibition was determined. This cell panel screen demonstrated that while BAF inhibition via BRG1 and BRM has broad activity across multiple cancer cell types, AML cell lines were among the most sensitive cells. FHD-286 was further profiled in select AML cell lines and demonstrated similar activity.

In in vitro cell proliferation assays, the average $IC_{50}$ of FHD-286 in MV-4-11 AML cells was 8.8 nM.

Mechanistic studies in AML cells showed FHD-286 concentration- and time-dependent upregulation of the myeloid maturation marker CD11b, along with decreases in markers associated with highly proliferative, undifferentiated, and robust blast-like phenotypes (Ki67, BRG1, BCL-2) at clinically relevant concentrations. The phenotypic changes were reversible upon washout of FHD-286 after 7 days of exposure. Preliminary results from a separate study in HL-60 cells suggest that reducing the FHD-286 concentration, rather than removing FHD-286 entirely, may maintain the percentage of CD11b-positive cells. In addition, a longer duration of initial exposure to FHD-286 (10 days vs 7 days) may increase the durability of the differentiation effect after washout. Collectively, these results suggest that FHD-286 may overcome the differentiation block in AML blasts, that CD11b may be a useful clinical biomarker to monitor this, and that exposure to higher concentrations of FHD-286 may result in a faster shift to a differentiated phenotype. Similar in vitro studies using FHD-286 in combination with either cytarabine or decitabine suggest that the combinations may result in enhanced cytoreductive effects and increased antitumor activity in vivo.

After oral doses of FHD-286 in MV-4-11 xenografts, significant tumor growth inhibition (TGI) with a clear dose-response was observed (37% and 76% TGI at 0.5 and 1.5 mg/kg once daily [QD], respectively). Levels of MYC and CTTN mRNA, targets of BRG1/BRM in AML, correlated with plasma concentrations of FHD-286 in both single and repeated dosing studies.

In vivo studies of FHD-286 administered in combination with cytarabine or decitabine showed increased antitumor activity with the combination versus FHD-286, cytarabine, or decitabine as a single agent. In the MV-4-11 AML xenograft model, FHD-286 QD + cytarabine 5-days-on/2-days-off ×2 resulted in substantially greater TGI (70.6%) than either single agent (FHD-286: 39.4%; cytarabine: 44.5%). In another study in this model when FHD-286 was administered QD for 5 days, then FHD-286 and cytarabine were administered concomitantly on a 5-days-on/2-days-off regimen, the TGI with the combination (84.9%) was greater than with either single agent (FHD-286: 77.4%; cytarabine: 58.1%). In mice bearing patient-derived xenografts expressing MLL-AF9 and FLT3-TKD mutations, FHD-286 5-days-on/2-days off + decitabine Days 1 through 5 significantly reduced tumor burden versus single-agent FHD-286 ($p \leq 0.05$) or decitabine ($p \leq 0.001$), and significantly improved median survival versus single-agent FHD-286 or decitabine ($p \leq 0.05$ for both).

In AML xenograft models, 4 days of dosing with FHD-286 in combination with either cytarabine or decitabine resulted in increases in cleaved caspase-3, a marker of apoptosis, and γ-H2AX, a marker for the detection and quantification of DNA damage.

Safety pharmacology studies support that FHD-286 is not expected to have an impact on cardiovascular or respiratory systems. Oral administration of FHD-286 at single doses up to 1.0 mg/kg was not associated with effects on cardiovascular or respiratory function in dogs. In an hERG study, inhibition at 10 μM was statistically significant ($p < 0.05$) when compared to vehicle control values. The $IC_{50}$ for the inhibitory effect of FHD-286 on hERG potassium current was not calculated but was greater than 10 μM.

There were no effects in safety pharmacology studies that would preclude the safe administration of FHD-286 according to the proposed clinical protocol.

In summary, FHD-286 shows profound growth inhibition of AML cell lines, along with in vivo antitumor activity in relevant xenograft models as monotherapy and in combination with cytarabine or decitabine. Taken together, these data support plans to evaluate FHD-286 in AML and associated hematologic malignancies in humans.

**Nonclinical Drug Metabolism and Pharmacokinetics**

FHD-286 showed low clearance (CL) and low to moderate volume of distribution at steady state across nonclinical species following single intravenous (IV) administration. Terminal half-life ($t_{1/2}$) ranged from 1.5 hours in monkeys to 6.9 hours in dogs. Following oral administration in a solution formulation, oral bioavailability (F) ranged from 44% in monkeys to >100% in rats and dogs. When administered in a suspension formulation, FHD-286 showed good oral bioavailability (F ~75%) in rodents, and lower oral bioavailability (F ~11%) in dogs. When FHD-286 was administered by capsule, the proposed form to be used for first-in-human studies, to fasted and fed dogs, F was 47% and 31%, respectively. Mean maximum observed plasma concentration ($C_{max}$) and areas under the plasma concentration-time curves (AUCs) from time 0 to the last time point ($AUC_{0-t}$) were 2.7- and 1.5-fold higher in fasted dogs than in fed dogs, respectively. The differences were statistically insignificant. In the 28-day toxicity studies, FHD-286 was administered orally in a suspension formulation. Plasma exposure of FHD-286 increased with dose in rats (0.3 to 3 mg/kg/day) and dogs (0.1 to 1 mg/kg/day). Unbound exposure of FHD-286 on Day 28 (sex combined) was 5.4 and 11.1 ng*h/mL for the dose of 3 mg/kg/day in rats and 1 mg/kg/day in dogs, respectively. There was no sex-based difference observed in the plasma exposures of FHD-286. Following daily oral administration for 28 days, FHD-286 accumulation was minimal (accumulation ratios ≤2.2, sex combined) in both rats and dogs. The toxicokinetics of FHD-286 in the 91-day toxicity studies was similar.

FHD-286 was identified as a substrate for both human permeability glycoprotein (P-gp) and breast cancer resistance protein (BCRP) with passive permeability of $>15\times10^{-6}$ cm/s in Madin-Darby canine kidney cells. FHD-286 was highly bound to plasma proteins across species, with higher binding in rodents than non-rodents, and did not partition into blood cells extensively when tested in vitro in rat, dog, monkey, and human whole blood.

In rats, FHD-286 distributed readily into tissues. Bone marrow $C_{max}$ was observed by 1 hour postdose. The highest degree of distribution was observed in the walls of the large and small intestine, followed by the liver.

In metabolism studies using liver microsomes or cryopreserved hepatocytes, FHD-286 showed low to high intrinsic clearance ($CL_{int}$). The scaled $CL_{int}$ followed the rank order of monkey > human > rat > dog. In the metabolite profiling study in rat, dog, monkey, and human cryopreserved hepatocytes, FHD-286 was relatively stable with no metabolite greater than 5% of total drug-related material formed in any of the species. In plasma samples from subjects with metastatic uveal melanoma treated with FHD-286 10 mg QD in Study FHD-286-C-001, unchanged FHD-286 accounted for >85% of the total drug-related material. The metabolic pathways identified were amide hydrolysis, dealkylation, dehydrogenation, hydroxylation, hydrogenation, and di-oxidation.

Cytochrome P450 (CYP) 3A was identified as the major CYP enzyme responsible for FHD-286 in vitro metabolism. In preliminary excretion studies following single IV administration, excretion of FHD-286 as unchanged form was a minor elimination pathway in rats and monkeys.

In pooled human liver microsomes, the $IC_{50}$ value of direct inhibition was 20 µM for CYP2C8 and >30 µM for CYP1A2, CYP2B6, CYP2C, CYP2C19, CYP2D6, and CYP3A4/5. Preincubation with FHD-286 in human liver microsomes did not enhance inhibition for the CYP isoforms tested except CYP3A4/5. FHD-286 was a weak time- and nicotinamide adenine dinucleotide phosphate-dependent inhibitor of CYP3A4/5; time-dependent CYP3A inhibition was apparent at 30 µM, but not at concentrations ≤10 µM. In cells over-expressing specific transporters, FHD-286 inhibited organic anion transporter (OAT) 3, organic anion transporting peptide (OATP) 1B1, OATP1B3, multidrug and toxin extrusion transporter (MATE) 1, MATE2-K, BCRP and P-gp, but not OAT1 and organic cation transporter (OCT) 2. Estimated $IC_{50}$ ranged from 0.318 µM for MATE1 and 5.79 µM for OAT3 after being corrected for non-specific binding. Based on the observed clinical $C_{max}$, FHD-286 has low to no potential of drug-drug interactions (DDI) as an inhibitor of OATP1B1, OATP1B3, OAT1, OAT3, OCT2, MATE1 and MATE2-K, a moderate potential of DDI as an inhibitor of CYP1A2, CYP2C8, CYP2C9 and CYP3A4, and a high potential of DDI as an inhibitor of P-gp and BCRP. P-gp and BCRP inhibition in the gut may be clinically relevant for sensitive P-gp and BCRP substrates that are orally administered and have a narrow therapeutic index (TI).

In cultured human hepatocytes, FHD-286 treatment decreased mRNA levels of CYP1A2, CYP3A4, CYP2C8, and CYP2C9 in a concentration-dependent manner, whereas CYP2B6 and CYP2C19 expressions were not affected. More than 50% decrease in mRNA expressions were observed at FHD-286 concentrations ≥1 nM for CYP3A4, and at concentrations ≥10 nM for CYP1A2, CYP2C8, and CYP2C9. Though CYP3A4 mRNA levels decreased at concentrations that ranged from 1 to 50 nM, CYP3A activity was minimally affected (≤32% decrease). CYP1A2 enzyme activity was decreased by >50% at FHD-286 concentrations ≥10 nM. The in vivo relevance of these findings remains to be determined in the clinic as in vitro to in vivo extrapolation of CYP down-regulation has not been established (FDA 2020; Hariparsad, et al 2017).

In conclusion, the nonclinical PK and ADME studies support the clinical evaluation of FHD-286 in accordance with the proposed clinical protocol.

**Toxicology**

Based on the non-GLP-compliant, 14-day oral repeat dose study in Wistar Han rats, 3 mg/kg/day of FHD-286 was deemed the maximum tolerated dose (MTD) and was selected as the high dose for the 28-day GLP-compliant toxicity study in rats.

The administration of FHD-286 by oral gavage QD to Wistar Han rats for up to 28 days at doses of 0, 0.3, 1, or 3 mg/kg/day was well tolerated. FHD-286 induced reduction in mean food consumption and body weight gain in the male and female rats at 3 mg/kg/day throughout the dosing period. FHD-286-related changes in hematology, coagulation, and clinical chemistry parameters were noted at 3 mg/kg/day and a dose-related lower urine pH in all groups at the end of the dosing period. Administration of FHD-286 also resulted in organ weight decreases in the thymus and spleen at ≥1 mg/kg/day, liver at ≥0.3 mg/kg/day, and testis and epididymis in males at 3 mg/kg/day. Macroscopic changes were observed in the thymus, mesenteric lymph node and testes at 3 mg/kg/day. Microscopic changes were seen in the thymus and mesenteric lymph nodes at ≥1 mg/kg/day and the bone marrow, femoral/tibial bone, pancreas, testis, and epididymis at 3 mg/kg/day. At the end of the recovery period, organ weight decreases were still observed in the testis, epididymis, and thymus in males. Microscopic findings were observed in the testes and epididymis in the 3 mg/kg/day recovery males, exhibiting similar relative incidence, histologic characteristics, and severity to males in the 3 mg/kg/day main study group, indicating no resolution of changes by the end of the recovery period. No other treatment-related microscopic findings seen in the main study animals were noted in the recovery animals, indicating complete recovery with cessation of dosing. The no-observed-adverse-effect level (NOAEL) in the 28-day rat study was considered to be 1 mg/kg/day (sex combined $C_{max}$ and area under the concentration versus time curve from time 0 to the last quantifiable concentration [$AUC_{tlast}$] values were 312 ng/mL and 3790 ng*h/mL, respectively, on Day 28).

Based on the results of the non-GLP-compliant, 14-day oral repeat dose toxicity study in beagle dogs, 1 mg/kg/day of FHD-286 was determined to be the MTD and was selected as the high dose for the 28-day GLP-compliant toxicity study in dogs.

In the 28-day repeat dose toxicity study in beagle dogs at doses of 0.1, 0.3, and 1 mg/kg/day, administration of FHD-286 by QD oral gavage was tolerated at dose levels up to 1 mg/kg/day. FHD-286-related clinical observations included lameness/abnormal gait with no obvious injury or wound at ≥0.3 mg/kg/day in males and at 1 mg/kg/day in females. Repeat dose Carprofen at 2.2 mg/kg once a day resulted in normal ambulation within 1 to 2 days; additionally, upon cessation of the treatment during the recovery period, normal ambulation was observed. FHD-286-related reduced appetite or low body (condition) score treated with wet food enrichment occurred for several animals, and resolved upon cessation of treatment during recovery period. FHD-286-related changes were observed in hematology parameters at ≥0.1 mg/kg/day, clinical chemistry parameters at ≥0.1 mg/kg/day (males) and at 1 mg/kg/day (females), and coagulation at 1 mg/kg/day. Target organ effects included macroscopic abnormalities (dark or pale foci/discoloration) in the gastrointestinal (GI) tract, lymph nodes, and thymus (small). One or a combination of these macroscopic findings were noted in males and females at ≥0.1 mg/kg/day and all resolved by the end of the recovery period. Organ weight changes included decreased splenic weights (males at ≥0.1 mg/kg/day) and decreased thymic weights (males and females at ≥0.1 mg/kg/day) on Day 29. By Day 22 of recovery, all organ weight changes had resolved, except the decreased thymic weights in males at 1 mg/kg/day. FHD-286 administration was associated with dose-dependent minimal to moderate degeneration/atrophy of the seminiferous tubules of the testis at ≥0.3 mg/kg/day. In addition, FHD-286 administration was associated with histologic lesions in the epididymides (at 1 mg/kg/day), GI tract (at ≥0.1 mg/kg/day), bone marrow (at 1 mg/kg/day), spleen (at ≥0.1 mg/kg/day), thymus (at ≥0.1 mg/kg/day), mesenteric lymph node (at ≥0.1 mg/kg/day), and vascular lesions in the lung or urinary bladder (at ≥0.1 mg/kg/day) on Day 29. By Day 22 of recovery, histologic lesions present in the spleen and thymus at 1 mg/kg/day partially resolved, while the splenic extramedullary hematopoiesis was compensatory to the decreased bone marrow cellularity described on Day 29. All other histologic lesions resolved by this time point. In the 28-day dog study, the NOAEL was considered to be 1 mg/kg/day for females and 0.3 mg/kg/day for males due to FHD-286-related findings in testes.

Findings from the 3-month, repeat-dose, GLP-compliant toxicology studies (8- to 9-week recovery period) in rats and dogs were consistent with the findings from the 28-day repeat-dose GLP-compliant studies, and indicated reductions in body weight gain and modest changes in hematology parameters consistent with the observed histological changes in thymus and other lymphoid tissues (depletion) and bone marrow (reduced hematopoiesis). The major organ toxicity was testicular atrophy/degeneration and the presence of reduced or degenerating sperm in the epididymis. Prostatic atrophy also occurred, albeit less commonly. These male reproductive system changes occurred over a dose range of 0.1 mg/kg/day (minimal severity) to 1 mg/kg/day (marked) in the 3-month study in dogs, and were shown to be reversed (0.1 mg/kg/day) or reduced in severity (0.3 and 1.0 mg/kg/day) after 8 to 9 weeks of recovery. Similar changes were produced in the 3-month rat study at the high dose of 3 mg/kg/day, but not at lower doses; this effect was reduced in incidence or severity after 8 to 9 weeks of recovery, but not fully reversed in the rats.

In GLP-compliant genetic toxicity studies, FHD-286 was negative for potential mutagenicity in a bacterial mutation test under all test conditions. FHD-286 was considered positive for inducing structural aberrations in the Chinese hamster ovary cell line CHO-WBL with and without S9 under the specific study conditions. Additionally, there were significant test article-related increases in numerical aberrations (endoreduplication) in FHD-286-treated cultures in the short treatments with and without S9.

FHD-286 showed potential for phototoxicity in BALB/c 3T3 mouse fibroblasts. The $IC_{50}$ (µg/mL) in the absence of ultraviolet (UV) radiation (-UVR) was not achieved and the $IC_{50}$ (µg/mL) in the presence of UV radiation (+UVR) was 1.265 and 1.340 for definitive assays 1 and 2, respectively.

In conclusion, the nonclinical data support the conduct of the proposed clinical protocol with appropriate monitoring in place.

###### 5.3.1.3. Summary of Clinical Information

Refer to the FHD-286 IB for updated clinical data.

##### 5.3.2. Low-Dose Cytarabine

Cytarabine is a cytotoxic agent that inhibits DNA synthesis (Cytarabine Injection December 2022). At low doses, cytarabine, either alone or in combination with other anticancer agents, is used as a less intensive treatment option for advanced hematologic malignancies such as AML and MDS in patients not suitable to receive intensive chemotherapy (Burnett, et al 2007; Kantarjian, et al 2021). Current guidelines recommend LDAC monotherapy (20 mg/m$^2$/day subcutaneously [SC] for 10 consecutive days every 28 days) as a less intensive treatment option for R/R AML (NCCN 2023), and LDAC in combination with venetoclax for treatment of AML in patients not suitable for intensive chemotherapy (Döhner, et al 2022).

In a randomized clinical study in patients with AML and high-risk MDS who were primarily >60 years old and unfit for intensive chemotherapy, LDAC resulted in an 18% CR rate (Burnett, et al 2007).

Warnings and precautions for cytarabine relevant to the dose level, route, and regimen to be used in this study include myelosuppression (Cytarabine Injection December 2022).

##### 5.3.3. Decitabine

Decitabine is a nucleoside metabolic inhibitor that causes cellular differentiation or apoptosis, depending on dose (Dacogen (decitabine) for injection for intravenous use June 2020). Decitabine is indicated in the US for treatment of MDS (newly diagnosed and previously treated) and in the EU for treatment of AML, and is commonly used in the US and the EU to treat AML in patients who are older or not suitable to receive intensive chemotherapy (Dacogen 50 mg powder for concentrate for solution for infusion February 2022; Dacogen (decitabine) for injection for intravenous use June 2020; Kantarjian, et al 2021). Current guidelines recommend decitabine as a less intensive treatment option for R/R AML (NCCN 2023), and decitabine in combination with venetoclax for patients with AML not suitable for intensive chemotherapy (Döhner, et al 2022). Hypomethylating agents such as decitabine are recommended in the treatment paradigms for R/R MDS and for CMML (NCCN 2022).

In a Phase 2 study in patients with previously untreated AML who were >60 years old, the overall CR rate with decitabine 20 mg/m$^2$ IV QD on Days 1 through 5, every 4 weeks, was 24% (Cashen, et al 2010). In a Phase 3 clinical study in subjects with AML treated with decitabine at the same dose and on the same regimen, the CR + CRp (CR with incomplete platelet recovery) rate was 17.8% and the median overall survival was 7.7 months (Kantarjian, et al 2012).

Warnings and precautions for decitabine include myelosuppression (Dacogen (decitabine) for injection for intravenous use June 2020).

Decitabine may be administered intravenously on a 5-day regimen of 20 mg/m$^2$ over 1 hour daily for 5 days every 4 weeks (Dacogen (decitabine) for injection for intravenous use June 2020).

#### 5.4. Study Rationale

##### 5.4.1. Parts I and II: FHD-286 Monotherapy *(Closed to Enrollment)*

FHD-286 is a potent, selective, allosteric, and orally available small molecule inhibitor of the enzymatic activity of both BRG1 and BRM. In a MV-4-11 AML mouse xenograft model, significant tumor growth inhibition was observed.

This study is an ascending multiple dose clinical trial. It is primarily intended to evaluate the safety and tolerability of FHD-286 when administered orally to subjects with advanced hematologic malignancies, specifically R/R AML and R/R MDS. This study will allow for the determination of the recommended phase 2 dose (RP2D) in subjects with advanced hematologic malignancies. This study will also evaluate the PK/PD profiles of multiple dose administration of FHD-286 and a preliminary assessment of antitumor activity in subjects with AML as well as other associated hematologic malignancies.

The data from this study in subjects with advanced hematologic malignancies, including safety, tolerability, PK/PD findings, and antitumor activity, will form the basis for subsequent clinical development of FHD-286.

##### 5.4.2. Part III: Combination Therapy

FHD-286 is a potent, selective, allosteric, and orally available small molecule inhibitor of the enzymatic activity of both BRG1 and BRM. Enhanced antitumor activity has been observed in mouse AML xenograft models when FHD-286 was administered in combination with cytarabine or decitabine, compared with that of the single agents. Based on these results, LDAC and decitabine, standard of care agents for subjects with advanced hematologic malignancies not suitable to receive intensive chemotherapy, were selected for separate evaluation in the 2 combination therapy arms of this dose escalation study. In addition, the cytoreductive properties of cytarabine and decitabine may mitigate the risk for differentiation syndrome with FHD-286 (Fathi, et al 2021).

This study is an ascending multiple dose clinical trial. It is primarily intended to evaluate the safety and tolerability of FHD-286 when administered orally in combination with either LDAC or decitabine to subjects with R/R AML, R/R MDS, and R/R CMML not in blast crisis. This study will allow for the determination of the RP2D(s) in this population. This study will also evaluate the PK/PD profiles of multiple dose administration of FHD-286 when administered in combination with LDAC or decitabine, and a preliminary assessment of antitumor activity in subjects with R/R AML, R/R MDS, and R/R CMML not in blast crisis. In addition, this study will evaluate the PK and safety of FHD-286 when administered in combination with LDAC or decitabine in subjects who are, and are not, receiving concomitant triazole antifungal agents classified as strong CYP3A4 inhibitors.

The data from this study in subjects with advanced hematologic malignancies, including safety, tolerability, PK/PD findings, and antitumor activity, will form the basis for subsequent clinical development of FHD-286.

#### 5.5. Risk-Benefit Assessment and Risk Mitigation

##### 5.5.1. Parts I and II: FHD-286 Monotherapy *(Closed to Enrollment)*

This study design (two-part, accelerated titration design beginning with single-subject cohorts and transitioning to a 3+3 design) minimizes subtherapeutic dosing in early cohorts in subjects with rapidly progressing advanced hematologic malignancies while allowing for the evaluation of multiple dose levels of FHD-286 (Section 7.1).

FHD-286 shows profound growth inhibition AML cell lines and robust efficacy was observed in a series of in vivo studies conducted in a MV-4-11 xenograft mouse model. These data suggest FHD-286 demonstrates potential utility in treating AML in humans.

The nonclinical toxicology program demonstrated that FHD-286 has an acceptable safety profile to proceed with investigation in a clinical trial. The principal safety findings observed in GLP-compliant nonclinical settings that may be potential risks for humans include effects on the gastrointestinal tract (gastrointestinal bleeding, vomiting, and diarrhea); inappetence (reduced food consumption and weight loss); decreased bone marrow cellularity and effects in hematologic parameters, including thrombocytopenia due to an on-target effect on megakaryocytes slowing the production of new platelets; effects in mesenteric lymph nodes; testicular degeneration; effects in femoral/tibial bone; lameness/abnormal gait; and phototoxicity.

These potential risks have been considered when designing this clinical study, minimizing the number of subjects exposed at any one time at each dose level, following strict dose escalation criteria, and setting conservative stopping rules. A comprehensive series of safety evaluations, including monitoring for adverse events (AEs), laboratory parameters, physical examinations, vital signs, electrocardiograms (ECGs), echocardiograms (ECHOs) (or other means of assessing left ventricular ejection fraction [LVEF]), and Eastern Cooperative Oncology Group (ECOG) performance status (PS) assessment will be conducted to evaluate and monitor the safety profile of FHD-286. The Clinical Study Team (CST), comprised of the Sponsor (Responsible Medical Officer), Study Medical Monitor, and Investigators, will evaluate the safety of dosing by reviewing the emerging data from each dose cohort to determine the next dose level and regimen or whether the RP2D and/or MTD has been identified.

The results of the nonclinical PK studies showed that FHD-286 has an acceptable ADME profile to proceed with investigation in clinical trials and that FHD-286 has the potential for DDI (Section 5.3.1.2). Due to this potential for drug interaction, concomitant administration of strong CYP3A inhibitors, strong CYP3A inducers, sensitive CYP3A substrates with a narrow TI, and orally administered sensitive P-gp and BCRP substrates with narrow TIs will be avoided. Exceptions may be made for therapy in the case of life-threatening infections, for example a triazole anti-fungal agent to reduce the risk of invasive fungal infections, at the discretion of the Sponsor. Appropriate eligibility criteria and concomitant medication guidance regarding the potential for drug interactions are provided in Section 8.3 and Section 9.12, respectively.

Differentiation syndrome is an identified risk with FHD-286, as described in the IB; identification and management of differentiation syndrome is described in Section 9.7.4.

Overall, the potential benefits of treatment with FHD-286, a BRG1 and BRM inhibitor acting through a novel mechanism, outweigh the potential risks to subjects with advanced hematologic malignancies, conditions that are rapidly progressive and with significant unmet medical needs.

##### 5.5.2. Part III: Combination Therapy

The design of Part III (3+3 dose escalation in 2 combination therapy arms) allows the FHD-286 dose to be escalated efficiently and safely while coadministering standard doses and regimens of either LDAC (20 mg/m$^2$ SC QD, Days 1 through 10 of each cycle) or decitabine (20 mg/m$^2$ IV QD, Days 1 through 5 of each cycle). This study design enables close subject monitoring while evaluating the safety and potential clinical activity of FHD-286 in combination with LDAC or decitabine. Each group (A1, A2, B1, and B2) will be evaluated independently.

The potential risks with FHD-286 were identified based on GLP-compliant nonclinical toxicology studies and continue to be monitored in the FHD-286 clinical development program. These include effects on the gastrointestinal tract (gastrointestinal bleeding, vomiting, and diarrhea); inappetence (reduced food consumption and weight loss); decreased bone marrow cellularity and effects in hematologic parameters, including thrombocytopenia due to an on-target effect on megakaryocytes slowing the production of new platelets; effects in mesenteric lymph nodes; testicular degeneration; effects in femoral/tibial bone; lameness/abnormal gait; and phototoxicity.

Differentiation syndrome, which has only been observed in subjects with advanced hematologic malignancies (Study FHD-286-C-002), is an identified risk with FHD-286. Identification and management of differentiation syndrome is described in Section 9.7.4.

Exploratory analyses of clinical PD data suggest dose-dependent target engagement and effects on the biology of AML in response to FHD-286. In peripheral blood samples from subjects treated with FHD-286 monotherapy in this study, increased levels of markers of myeloid and monocyte maturation (CD11b and CD64) and decreased levels of markers of hematopoietic stem cell identity and apoptosis (CD34, BCL2, and BRG1) have been observed, compared with baseline. A similar pattern was observed in bone marrow. This suggests that FHD-286 has a differentiation effect on blast cells, which supports its potential utility in advanced hematologic malignancies. However, while a subset of subjects with advanced hematologic malignancies treated with FHD-286 monotherapy achieved stable or decreasing peripheral and/or bone marrow blast counts, with or without absolute neutrophil count (ANC) recovery, overall preliminary clinical activity with FHD-286 monotherapy in this population has thus far been limited.

Use of a standard of care agent offers subjects the opportunity to receive clinical benefit while the safety and potential clinical activity of FHD-286 in combination with these agents is explored. Enhanced antitumor activity, compared with that of the individual agents (FHD-286, cytarabine, decitabine) as monotherapy, has been observed in mouse AML xenograft models when FHD-286 was administered in combination with cytarabine or decitabine, suggesting that the combination of FHD-286 with these standard of care agents may result in improved clinical activity in patients with advanced hematologic malignancies. In addition, the cytoreductive properties of cytarabine and decitabine may mitigate the risk for differentiation syndrome with FHD-286 (Fathi, et al 2021).

The potential and identified risks of FHD-286 have been considered when designing this clinical study. The study design minimizes the number of subjects exposed at any one time at each dose level, follows strict dose escalation criteria, sets conservative stopping rules, and incorporates comprehensive monitoring and management guidelines for differentiation syndrome and other toxicities. A comprehensive series of safety evaluations, including monitoring for AEs, laboratory parameters, physical examinations, vital signs, ECGs, and assessments of LVEF, will be conducted to evaluate and monitor the safety profile of FHD-286 in combination with LDAC or decitabine. The CST, comprising, at minimum, the Sponsor (Responsible Medical Officer) and Investigators, will evaluate the safety of dosing FHD-286 in combination with LDAC or decitabine by reviewing all relevant emerging data from each dose group to determine the next dose level or whether the RP2D(s) and/or MTD has been identified. An independent safety monitoring committee will periodically review safety data.

The results of the nonclinical PK studies showed that FHD-286 has an acceptable ADME profile to proceed with investigation in clinical trials and that FHD-286 has the potential for DDIs. Due to this potential for drug interaction, concomitant administration of strong CYP3A inhibitors, strong CYP3A inducers, sensitive CYP3A substrates with a narrow TI, orally administered sensitive P-gp and BCRP substrates with narrow TIs, strong P-gp and BCRP inhibitors, and proton pump inhibitors (PPIs) is prohibited. Triazole antifungal agents, including those that are strong CYP3A4 inhibitors, are permitted. The PK and safety of FHD-286 when administered in combination with LDAC or decitabine will be evaluated in subjects who are, and are not, receiving concomitant triazole antifungal agents classified as strong CYP3A4 inhibitors. Appropriate eligibility criteria and concomitant medication guidance regarding the potential for drug interactions are provided in Section 8.3 and Section 9.12, respectively.

The study population to be enrolled includes subjects with R/R AML, R/R MDS, or R/R CMML not in blast crisis who have received ≤4 prior lines of systemic anticancer therapy for their disease under study and who are appropriate candidates for LDAC or decitabine treatment. Given the poor prognosis and lack of substantially effective treatments for these patients, the potential benefits of treatment with FHD-286 in combination with standard of care LDAC or decitabine outweigh the potential risks.

### 6. STUDY OBJECTIVES AND ENDPOINTS

#### 6.1. Primary Objectives

The primary objectives of the study are:

- Parts I and II:
  - To determine the safety and tolerability of FHD-286 when administered as an oral monotherapy in subjects with advanced hematologic malignancies
  - To identify the RP2D(s) of FHD-286 when administered as monotherapy in subjects with advanced hematologic malignancies
- Part III:
  - To determine the safety and tolerability of FHD-286 when administered in combination with either LDAC or decitabine (with or without concomitant triazole antifungal agents classified as strong CYP3A4 inhibitors) in subjects with advanced hematologic malignancies
  - To identify the RP2D(s) of FHD-286 when administered in combination with either LDAC or decitabine (with or without concomitant triazole antifungal agents classified as strong CYP3A4 inhibitors) in subjects with advanced hematologic malignancies

#### 6.2. Secondary Objectives

The secondary objectives of the study are:

- Parts I and II:
  - To determine the PK of FHD-286 when administered as an oral monotherapy in subjects with advanced hematologic malignancies
  - To characterize the preliminary clinical activity associated with FHD-286 when administered as monotherapy in subjects with advanced hematologic malignancies
- Part III:
  - To determine the PK of FHD-286 when administered in combination with either LDAC or decitabine in subjects with advanced hematologic malignancies who are *not* receiving triazole antifungal agents classified as strong CYP3A4 inhibitors
  - To determine the PK of FHD-286 when administered in combination with either LDAC or decitabine in subjects with advanced hematologic malignancies who are receiving triazole antifungal agents classified as strong CYP3A4 inhibitors
  - To characterize the preliminary clinical activity associated with FHD-286 when administered in combination with either LDAC or decitabine in subjects with advanced hematologic malignancies

#### 6.3. Exploratory Objectives

Exploratory objectives of the study are:

- Parts I and II:
  - To evaluate the PK/PD relationship of FHD-286 when administered as monotherapy and changes in peripheral tissue (blood) and bone marrow biomarkers
  - To assess the associations of FHD-286 (when administered as monotherapy) exposure, clinical activity, and safety with PD and response markers in bone marrow and peripheral tissue (blood)
  - To investigate potential predictive and downstream markers of tumor response and/or resistance in bone marrow and peripheral tissue (blood)
  - To evaluate minimal residual disease (MRD)
- Part III:
  - To evaluate the PK/PD relationship of FHD-286 when administered in combination with either LDAC or decitabine and changes in peripheral tissue (blood) and bone marrow biomarkers
  - To assess the associations of FHD-286 (when administered in combination with either LDAC or decitabine) exposure, clinical activity, and safety with PD and response markers in bone marrow and peripheral tissue (blood)
  - To evaluate effects on PK, PD, safety, and clinical activity in subjects who began concomitant treatment with a triazole antifungal agent classified as a strong CYP3A4 inhibitor while on study
  - To investigate potential predictive and downstream markers of tumor response and/or resistance in bone marrow and peripheral tissue (blood)
  - To evaluate MRD

#### 6.4. Study Endpoints

##### 6.4.1. Primary Endpoints

- Incidence of AEs, dose-limiting toxicities (DLTs), serious AEs (SAEs), AEs leading to discontinuation, and adverse events of special interest (AESIs); safety laboratory assessments

##### 6.4.2. Secondary Endpoints

Plasma concentrations of FHD-286 will be determined and the following PK parameters will be calculated:

- $AUC_{0\text{-last}}$: AUC from time 0 to the time of the last quantifiable concentration
- $AUC_{0\text{-}24}$: AUC from time 0 to the time of 24 hours

- $AUC_{0-72}$: AUC from time 0 to the time of 72 hours
- $AUC_{0-inf}$: AUC from time 0 extrapolated to infinity
- $C_{max}$
- $T_{max}$: observed time to reach $C_{max}$
- $t_{1/2}$
- Vd/F: apparent volume of distribution
- CL/F: apparent total body clearance

Preliminary clinical activity (see Section 10.6):

- AML
  - CR rate
  - Duration of CR
  - CR + CRh (CR with partial hematologic recovery) rate
  - Duration of CR + CRh
  - Transfusion independence rate
  - Event-free survival (EFS)
  - OS
- MDS and CMML
  - CR rate
  - Duration of CR
  - Partial remission (PR) rate
  - Duration of PR
  - CR + PR rate
  - Duration of CR + PR
  - Hematologic improvement rate
  - EFS
  - OS

##### 6.4.3. Exploratory Endpoints

Exploratory analysis will include, but not be limited to, the following:

- Plasma concentration of FHD-286 and/or derived PK parameters
- Amount of FHD-286 in urine and fraction excreted in urine (Part III only)

- Measurements of PD and response biomarkers, including markers of differentiation and apoptosis (eg, CD11b, CD64, BCL2) in bone marrow and peripheral tissue (blood) samples
- Baseline and change from baseline in molecular status of multiple cancer, immune, and target-related genes (mutation/copy number/expression, etc.) and proteins in blood and bone marrow samples
- Evaluation of MRD (see Section 10.6)

### 7. INVESTIGATIONAL PLAN

#### 7.1. Overall Study Design

This is a Phase 1, multicenter, open-label, dose escalation study of FHD-286 administered orally as monotherapy or in combination with either LDAC or decitabine in subjects with advanced hematologic malignancies, specifically R/R AML, R/R MDS, and R/R CMML not in blast crisis (Figure 2). The study aims to determine the RP2D(s), the dose(s) and regimen(s) identified for continued study based on observed safety, tolerability, PK, PD, and preliminary clinical activity, and exposure-response analyses, of FHD-286, as monotherapy or in combination with either LDAC or decitabine, in subjects with advanced hematologic malignancies.

In Parts I and II of this dose escalation study, which are closed to enrollment, FHD-286 was administered QD as monotherapy in 28-day cycles.

In Part III of this dose escalation study, FHD-286 will be administered with either LDAC or decitabine:

- Arm A: FHD-286 administered orally QD on Days 1 through 28 of all cycles + LDAC 20 mg/m$^2$ administered SC QD on Days 1 through 10 of all cycles, in 28-day cycles
- Arm B: FHD-286 administered orally QD on Days 1 through 28 of all cycles + decitabine 20 mg/m$^2$ administered IV QD on Days 1 through 5 of all cycles, in 28-day cycles

Subjects will be assigned to groups in each treatment arm based on whether they are receiving a triazole antifungal agent classified as a strong CYP3A4 inhibitor at the start of study treatment:

- Groups A1 and B1: subjects *not* receiving a triazole antifungal agent classified as a strong CYP3A4 inhibitor at the start of study treatment
- Groups A2 and B2: subjects receiving a triazole antifungal agent classified as a strong CYP3A4 inhibitor at the start of study treatment

**Figure 2: Study Schema**

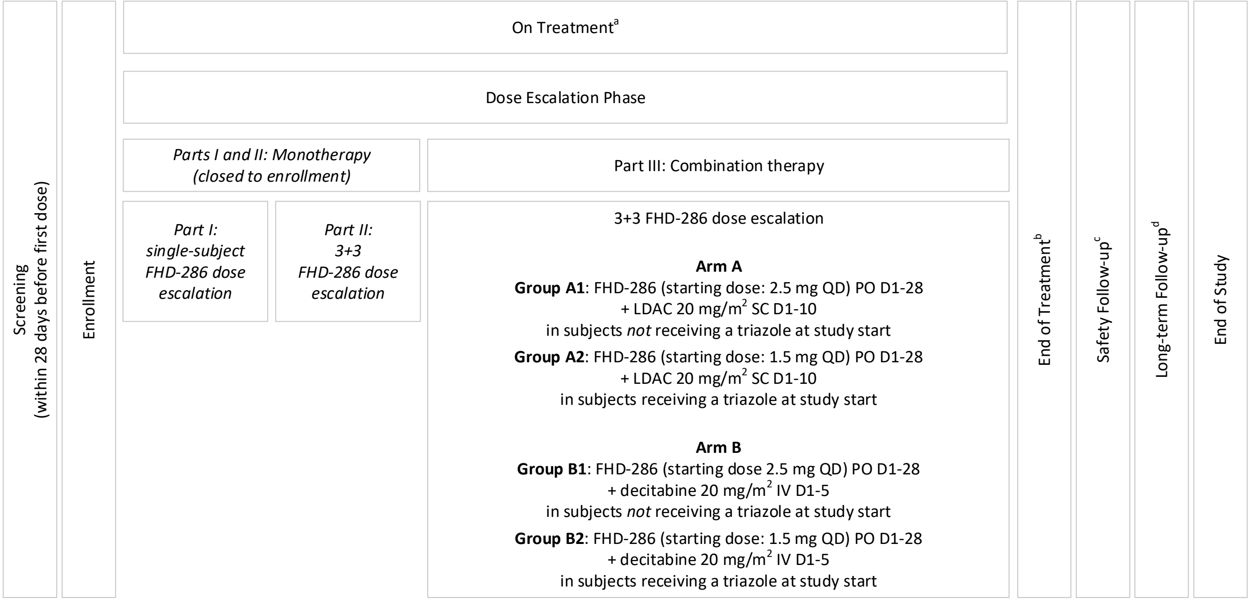

Abbreviations: CYP = cytochrome P450; D = Day; EOT = end of treatment; IV = intravenous(ly); LDAC = low-dose cytarabine; PO = oral(ly); QD = once daily; SC = subcutaneous(ly).
Note: All study drugs are administered on 28-day cycles. Triazole = triazole antifungal agent classified as a strong CYP3A4 inhibitor.

[a] Subjects may continue study treatment until 1 or more subject withdrawal criteria are met, or at the subject's discretion, or if the study is terminated.

[b] An EOT Visit will occur within 5 days after treatment end, the date on which the Investigator decides to discontinue FHD-286 treatment (this date is not required to be equivalent to the date of the last FHD-286 dose). If a subject's FHD-286 treatment is interrupted for 28 days and the subject then discontinues study participation, the EOT Visit will serve as the Safety Follow-up Visit.

[c] A Safety Follow-up Visit will occur approximately 28 days after the EOT Visit.

[d] After discontinuing treatment with FHD-286, subjects will be contacted by telephone approximately every 2 months for long-term follow-up, including assessment of disease status (unless subject discontinued FHD-286 treatment due to progressive disease/treatment failure, and until subject experiences progressive disease/treatment failure or starts a new anticancer therapy), survival status, and initiation of new anticancer therapies, until the subject has died, been lost to follow-up, withdrawn consent/assent, or for 2 years after the last subject discontinues FHD-286, whichever occurs first.

##### 7.1.1. Dose Escalation

###### 7.1.1.1. Parts I and II: FHD-286 Monotherapy *(Closed to Enrollment)*

A daily dosing regimen in 28-day cycles will be investigated during this study. Dose escalation will occur in two parts (Part I and Part II), as described below. Part I will enroll single-subject (n=1) cohorts. Dose escalations of up to 100% will be considered until any 1 (or more) Transition Criteria (Table 1) have been met, then dose escalation will transition to Part II which will enroll 3-subject cohorts in a "3+3" design. This allows for the optimization of dose escalation so that fewer subjects with rapidly progressive advanced hematologic malignancies are treated at potentially subtherapeutic dose levels of FHD-286.

Dose escalation will include approximately 25 to 50 subjects. The recommended starting dose was 5 mg once daily, as determined by the results of the FHD-286 Good Laboratory Practice (GLP)-compliant toxicology studies.

The dose escalation scheme for Part II will follow a modified Fibonacci design (ie, 100%, 67%, 50%, 40%, 33% increments from the previous dose in consecutive escalation cohorts). The safety of dosing will be evaluated by the CST. Real-time assessments of safety, tolerability, PK, and clinical activity will be conducted for each cohort evaluation throughout the study. Additionally, target engagement at each dose level will be evaluated by conducting RNA- and protein-based assessments on serial blood collections (PBMCs) that measure changes in BRG1/BRM downstream targets in AML blasts. The CST will review the available safety, tolerability, PK, and PD data from Cycle 1 of treatment of each dose cohort to determine whether it is safe to proceed to the next dose level or whether the RP2D and/or MTD has been identified. The RP2D is determined by the CST and is the dose regimen identified for continued study based on observed safety, tolerability, PK, PD, and clinical activity data; the RP2D will be further evaluated if and when expansion occurs. The identification of RP2D may or may not include the identification of the MTD (defined as the highest dose that causes DLTs in <2 of 6 subjects); RP2D and MTD may be equivalent. In the absence of observing an MTD, FHD-286 will only be escalated to the minimum or optimal safe and biologically effective dose.

Alternative dose levels and regimens (including, but not limited to, administration of the same total daily dose using different dosing schedules in concurrent groups, intermittent dosing, etc.) will be considered by the CST based on emerging data. Dosing cohorts may be added based on the emerging data. Additional subjects may be enrolled, for the replacement of subjects who are not evaluable for the assessment of dose escalation, for evaluation of alternative dosing regimens, or for further exploring safety, PK, PK/PD, or preliminary clinical activity used to guide the selection of the RP2D.

Toxicities that meet DLT criteria will be evaluated throughout the duration of treatment. Toxicities will be graded and documented according to the National Cancer Institute Common Terminology Criteria for Adverse Events (NCI CTCAE), version 5.0 (see Appendix 15.1). A DLT is defined as any AE that occurs during Cycle 1 (28 days) of treatment and that meets the specific criteria outlined in Section 9.6.1. All AEs that cannot clearly be determined to be unrelated to FHD-286 will be considered relevant to determining DLTs and will be reviewed by the CST.

**Part I: Single-Subject Dosing Cohorts**

One subject will be enrolled in the first dosing cohort and will be administered 5 mg of FHD-286 once daily for 28 days. The determination of the next dose level will be based on review of all available safety, tolerability, PK, and PD data from Cycle 1 (28 days) by the CST. One new subject will then be enrolled at the next dose level. Dose escalation will continue in this manner until any 1 (or more) of the Transition Criteria below (Table 1) are met.

**Table 1: Transition Criteria**

| Criteria | | Transition Action |
|---|---|---|
| 1 | One subject exhibits any DLT(s) during Cycle 1 of treatment | Cohort will enroll new subjects to reach n=6 |
| 2 | Two subjects exhibit any FHD-286-related non-DLT toxicities that are Grade ≥2 during Cycle 1 of treatment, regardless of dose level, in the opinion of the Investigator | Cohort at the highest dose level at the time Criterion 2 is met will enroll new subject(s) to reach n=3 |

As soon as any 1 (or more) of the above Transition Criteria are met, the dose escalation will transition to a "3+3" design with multiple ascending doses.

**Part II: "3+3" Dosing Cohorts**

During Part II of dose escalation, each dosing cohort will enroll 3 subjects. As in Part I, subjects in Part II will receive a daily dosing regimen of FHD-286 in 28-day cycle(s). If there are no DLTs observed after the third evaluable subject in a dose cohort completes Cycle 1 (ie, the 28-day DLT evaluation period), the study will proceed with dose escalation to the next cohort following review of all data by the CST. If any 1 of the subjects in a cohort experiences a DLT during Cycle 1 of treatment, then additional subjects will be enrolled in that cohort until that cohort contains 6 subjects. If none of the added subjects experience a DLT during the Cycle 1 DLT-evaluation period (28 days), dose escalation may continue to the next group following review of all available data by the CST. If 2 or more subjects in a cohort experience a DLT during Cycle 1, dose escalation will be halted, and the next lower dose level will be declared the MTD. Alternatively, a dose level in between the dose level exceeding MTD and the previous dose level may be explored and declared MTD if <2 out of 6 subjects experience a DLT at that dose.

In Part II, if there are multiple subjects in the screening process at the time the third subject within a dose cohort begins treatment, up to 2 additional subjects may be enrolled in that cohort, with approval of the Sponsor. Enrollment in this cohort will be paused for review for toxicities observed in 2 out of 6, 3 out of 10, or 4 out of 12 subjects.

Once the RP2D has been identified, an additional 6 to 12 subjects may be enrolled at that dose level(s) to confirm the observed safety and tolerability of FHD-286. Enrollment in this cohort will be paused for review for toxicities observed in 2 out of 6, 3 out of 10, or 4 out of 12 subjects.

##### 7.1.1.2. Part III: Combination Therapy

Part III of dose escalation will evaluate up to approximately 72 subjects in Arm A (FHD-286 + LDAC 20 mg/m$^2$) and up to approximately 72 subjects in Arm B (FHD-286 + decitabine 20 mg/m$^2$) (up to approximately 144 subjects total) (Section 12.1). Subjects will be assigned to treatment arms at the Investigator's discretion. Dose escalation and RP2D(s) selection will be conducted independently for each of the 4 groups in this study: A1, A2, B1, and B2..

Each dose group will enroll a minimum of 3 evaluable subjects.

The FHD-286 dose will be increased in up to 100% increments from the previous dose level ("dose level" refers to dose and regimen) in consecutive escalation cohorts. The CST will review the available safety, tolerability, PK, and PD data from the DLT evaluation period (Section 9.6.1) for each dose group, in the context of and in addition to other relevant data, to determine whether it is safe to proceed to the next FHD-286 dose level or whether the RP2D(s) and/or MTD, which may be equivalent, has been identified. An independent safety monitoring committee will perform comprehensive safety data review (Section 13.6).

The RP2D(s), which is determined by the Sponsor with endorsement from the CST, is the dose and regimen identified for continued study based on observed safety, tolerability, PK, PD, preliminary clinical activity, and exposure-response analyses; the RP2D(s) will be further evaluated if and when an expansion phase of this study occurs. The identification of RP2D(s) may or may not include the identification of the MTD (defined as the highest FHD-286 dose that causes DLTs in <2 of 6 subjects); an RP2D and the MTD may be equivalent. In the absence of observing an MTD, FHD-286 will only be escalated until the minimum and/or optimal safe and biologically effective dose(s) has been determined.

If there are no DLTs observed after the third evaluable subject at a dose level completes the DLT evaluation period, the study will proceed with dose escalation to the next dose level after the CST has reviewed all available data and approved the FHD-286 dose escalation. If any 1 of the subjects in a dose group experiences a DLT during the DLT evaluation period, then additional subjects will be enrolled in that dose group until that dose group contains 6 evaluable subjects, or a second DLT occurs, whichever occurs first. If none of the added subjects experience a DLT during the DLT evaluation period, dose escalation may continue to the next FHD-286 dose level after the CST has reviewed all available data and approved the FHD-286 dose escalation. If 2 or more subjects in a dose group experience a DLT during the DLT evaluation period, that dose level will be considered as exceeding the MTD. Dose escalation will be halted, and the next lower dose level will be declared the MTD, as long as <2 of 6 subjects experience a DLT at that dose level. Alternatively, a dose level lower than the dose level exceeding the MTD and higher than the previous dose level may be explored and declared the MTD if <2 of 6 subjects experience a DLT at that dose.

RP2Ds will be identified separately for each of the 4 groups in this study: A1, A2, B1, and B2. Once the presumptive RP2D(s) has been identified for a given group, an additional 6 to 14 subjects may be enrolled at that FHD-286 dose level(s) in that group to confirm the observed safety and tolerability of FHD-286 at the RP2D(s) in combination with LDAC or decitabine. Enrollment of these additional subjects will be paused for review if DLTs are observed in 2 of 6, 3 of 10, or 4 of 14 subjects. Additional stopping criteria will also be considered, as described in Section 9.8.

Interval dosing (Section 9.5.1) will be implemented for the FHD-286 5 mg QD and 7.5 mg QD doses for Groups A2 and B2 and may be implemented for Groups A1 and B1 if agreed upon by the CST. Alternative interval dosing regimens may be considered.

Dosing cohorts may be added based on the emerging data.

Additional subjects may be enrolled to replace subjects who are not evaluable for the assessment of dose escalation.

Toxicities that meet DLT criteria will be evaluated throughout the duration of treatment. Toxicities will be graded and documented according to the NCI CTCAE, version 5.0 (see Appendix 15.1). A DLT is defined as any AE that occurs during Cycle 1 (28 days) of treatment (or during the first 6 weeks of treatment, for participants who transition from interval dosing during Cycle 1 to a consistent dose at C2D1 employing the highest of the 2 doses administered during Cycle 1 of interval dosing [Section 9.5.1]) and that meets the specific criteria outlined in Section 9.6.1. All AEs that cannot clearly be determined to be unrelated to FHD-286 or the combination of FHD-286 with LDAC or decitabine, and for which there is no clear-cut alternative explanation (eg, due to disease progression), will be considered relevant to determining DLTs and will be reviewed by the CST.

###### 7.1.1.3. Intra-subject Dose Escalation

**Parts I and II: FHD-286 Monotherapy *(Closed to Enrollment)***

Intra-subject dose escalation of FHD-286 may be permitted; see Section 9.6.3. Data from subjects who have undergone intra-subject dose escalation will not be used by the CST to determine dose levels for subsequent cohorts.

**Part III: Combination Therapy**

Intra-subject dose escalation will not be permitted.

###### 7.1.1.4. Clinical Study Team Teleconferences and Documentation

Regularly scheduled teleconferences will serve as a forum for review of safety and other relevant data by the CST. Decisions to escalate (or maintain or de-escalate) the FHD-286 dose or to implement interval dosing for Part III, Groups A1 and B1, will be documented along with a summary of the information supporting the decision.

##### 7.1.2. General Study Conduct

It is anticipated that this study will be conducted at up to approximately 13 clinical sites in the United States and Europe.

**Parts I and II: FHD-286 Monotherapy *(Closed to Enrollment)***

Approximately 25 to 50 subjects are expected to be treated in Parts I and II of this study.

Additional subjects may be enrolled for the replacement of subjects who are not evaluable for the assessment of dose escalation, for evaluation of alternative dosing regimens, or for further exploring safety, PK, PK/PD, or preliminary clinical activity used to guide the selection of the RP2D.

The Schedules of Assessments for the study are provided in Section 10 for subjects receiving FHD-286 daily and in Appendix 15.2 for subjects receiving FHD-286 on alternative dosing regimens (eg, intermittent dosing).

**Part III: Combination Therapy**

Up to approximately 72 subjects each in Arms A and B (up to approximately 144 subjects total) are expected to be treated (Section 12.1).

The Schedules of Assessments for the study are provided in Section 10.

###### 7.1.2.1. Screening

Following informed consent (and assent, for subjects <18 years of age), subjects will undergo screening procedures within 28 days prior to the first dose of study treatment to determine eligibility for the study. Screening procedures include the following:

- Medical, surgical, and medication history
- Complete physical examination including vital signs, height, and weight
- ECHO for LVEF measurement (other methods of evaluating LVEF may be performed according to institutional practice)
- 12-lead ECGs in triplicate (started after approximately 3 minutes of recumbency or semi-recumbency, with each ECG conducted approximately 2 minutes apart)
- ECOG PS assessment
- Clinical laboratory assessments (hematology, chemistry, coagulation, inflammatory markers, cytokine panel, serum pregnancy test in females of childbearing potential, hepatitis panel, and human immunodeficiency [HIV] testing)
- A bone marrow biopsy and/or aspirate sample to determine baseline disease burden is required within approximately 28 days of the administration of the first dose of study treatment (Cycle 1, Day 1 [C1D1]).

###### 7.1.2.2. Treatment Period

Required clinic visits during the treatment period are outlined in the Schedules of Assessments. Dosing with study treatment will begin on C1D1; on this day, subjects will be required to remain in clinic for 8 hours after the first dose for clinical observation and PK/PD sampling. If the half-life of FHD-286 for any dose group differs significantly from what was predicted, the duration of time required in the clinic may be adjusted.

For subjects enrolled in Part III, Arm A, on Days 1 through 5 of Cycle 1 only, LDAC will be administered at the study site. After the first 5 doses (ie, beginning on C1D6), at the Investigator’s discretion and where consistent with institutional, state, and local guidelines and regulations, subjects may self-administer LDAC at home, except on scheduled visit days.

For subjects enrolled in Part III, Arm B, decitabine will be administered at the study site on Days 1 through 5 of each cycle.

Treatment will be administered on an outpatient basis; treatment may be administered in the inpatient setting as clinically indicated.

Subjects may withdraw or be withdrawn from study treatment under the following conditions: disease progression or treatment failure, based on disease response assessments; development of unacceptable toxicity; start of alternative anticancer therapy; or withdrawal of consent/assent (see Section 8.6 for additional withdrawal criteria). All subjects are to undergo an End of Treatment (EOT) assessment within 5 days after the date on which the Investigator decides to discontinue FHD-286 treatment (this date is not required to be equivalent to the date of the last FHD-286 dose).

Safety assessments conducted during the treatment period include the following:

- Physical examination
- Vital signs
- ECHO (or other means) for LVEF measurement
- 12-lead ECGs in triplicate (started after approximately 3 minutes of recumbency or semi-recumbency, with each ECG conducted approximately 2 minutes apart)
- ECOG PS assessment
- Clinical laboratory assessments (hematology, chemistry, coagulation, inflammatory markers, cytokine panel, pregnancy testing for females of childbearing potential)

All subjects will undergo sampling for PK/PD assessments during the treatment period as outlined in the Schedules of Assessments. Subjects must take their dose of study treatment in clinic on any PK and/or PD sampling days. Continuous urine collection will be performed starting on C1D15 for 24 hours for at least 3 subjects participating in Part III of this study. These subjects will be chosen in discussion with Investigator(s) based on considerations such as the dose at which the subject is to be initiated and the individual subject's ability to participate in the urine collection assessment.

**Parts I and II (FHD-286 Monotherapy) *(Closed to Enrollment):*** A bone marrow biopsy and/or aspirate will be performed approximately every 4 weeks for the first 24 weeks, then approximately every 8 weeks for the next 48 weeks of treatment, as clinically indicated thereafter, and once at EOT for evaluation of disease response by the Investigators based on modified International Working Group (IWG) criteria (Section 10.6).

**Part III (Combination Therapy):** A bone marrow biopsy and/or aspirate will be performed on Cycle 1 Day 15; on Day 1 of Cycles 2 through 6; on Day 1 of Cycles 8, 10, and 12; then on Day 1 of every third cycle (Cycle 15, 18, 21, etc); at the EOT Visit; and as clinically indicated. Disease response will be evaluated by the Investigators based on modified IWG criteria or other criteria appropriate for the subject's advanced hematologic malignancy (Section 10.6).

**All subjects:** Disease status may continue to be monitored as part of long-term follow-up as described in Section 7.1.2.4.

###### 7.1.2.3. Safety Follow-up

A Safety Follow-up Visit will occur 28 (±7) days after the EOT Visit. Every effort must be made to perform protocol-specified evaluations unless consent/assent to participate in the study is withdrawn. If a subject's FHD-286 dose is interrupted for 28 days and the subject then discontinues FHD-286 without restarting treatment, the EOT Visit can serve as the Safety Follow-up Visit.

**Part III (Combination Therapy):** If subjects who discontinue FHD-286 for reasons other than disease progression or treatment failure continue to be treated with LDAC or decitabine as monotherapy outside the context of this study, a bone marrow biopsy and/or aspirate should be collected at the Safety Follow-up Visit to assess disease response.

###### 7.1.2.4. Long-term Follow-up

After subjects have discontinued FHD-286, unless consent/assent to participate in the study is withdrawn, they will be contacted by telephone approximately every 2 months to document disease status (unless subject discontinued FHD-286 due to disease progression/treatment failure, and until subject experiences disease progression/treatment failure or starts new anticancer therapy), receipt and type of subsequent anticancer therapy, and survival status. Long-term follow-up will continue until all subjects have died, withdrawn consent/assent, or are lost to follow-up, or for 2 years after the last subject discontinues FHD-286, whichever occurs first.

#### 7.2. Justification of the Study Design

The primary objectives of this study are to evaluate the safety and tolerability of FHD-286, administered as either a monotherapy or in combination with either LDAC or decitabine, including the determination of the RP2D(s) to carry forward into further development.

**Parts I and II: FHD-286 Monotherapy *(Closed to Enrollment)***

The dose escalation in this study will begin with a daily dosing regimen of FHD-286 in cohorts of 1 subject each over a 28-day dosing cycle. This design minimizes exposure of subjects with rapidly progressive advanced hematologic malignancies to subtherapeutic doses of FHD-286.

Once any 1 (or more) of the safety Transition Criteria described (Table 1) are met, dose escalation will transition to a “3+3” design to assess ascending, multiple doses of FHD-286 with evaluation of the safety and tolerability of each dose cohort prior to dose escalation. This design was chosen to minimize risk to subjects while allowing evaluation of multiple dose levels of study drug.

A comprehensive series of safety evaluations, including monitoring for AEs, laboratory parameters, physical examinations, vital signs, ECGs, ECHOs (or other means of assessing LVEF), and ECOG PS will be conducted to evaluate the safety profile of FHD-286 and to aid in the determination of the RP2D(s).

This study includes serial blood sampling across multiple dose levels to assess its PK profile.

Consistent with the design of many Phase 1 oncology studies, preliminary evaluation of the potential clinical activity of FHD-286 is a secondary objective of this study. Secondary objectives include assessments of disease response based on modified IWG criteria or other criteria appropriate for the subject’s advanced hematologic malignancy (Section 10.6).

**Part III: Combination Therapy**

Dose escalation of FHD-286 in combination with either LDAC or decitabine will be conducted using a “3+3” design to assess multiple ascending doses of FHD-286, with evaluation of the safety, tolerability, PK, and PD of each dose group occurring before FHD-286 dose escalation. This design was chosen to minimize risk to subjects while allowing evaluation of multiple doses and regimens of FHD-286 in combination with either LDAC or decitabine.

Triazole antifungal agents are commonly used to treat or reduce the risk for invasive fungal infections in subjects with advanced hematologic malignancies. Some triazole antifungal agents are strong CYP3A4 inhibitors and are thus associated with risks of DDIs; preliminary data suggest that FHD-286 exposure is approximately 2- to 3-fold higher in subjects receiving concomitant moderate or strong CYP3A4 inhibitors. To allow evaluation of the effects of these concomitant therapies on the PK and safety of FHD-286, subjects in each of the 2 treatment arms will be assigned to 2 separate groups based on whether they are, at the start of study treatment, receiving therapy with a triazole antifungal agent that is a strong CYP3A4 inhibitor. Dose escalation will proceed independently in the 4 groups (A1, A2, B1, and B2).

A comprehensive series of safety evaluations, including monitoring for AEs, laboratory parameters, physical examinations, vital signs, ECGs, and assessment of LVEF will be conducted to evaluate the safety and tolerability profile of FHD-286 in combination with either LDAC or decitabine. The observed safety, tolerability, PK, PD, and preliminary clinical activity, and exposure-response analyses will inform the determination of the RP2D(s).

Serial blood sampling will be conducted across multiple FHD-286 dose levels to assess its PK profile.

Consistent with the design of many Phase 1 oncology studies, preliminary evaluation of the potential clinical activity of FHD-286 in combination with either LDAC or decitabine is a secondary objective of this study. Secondary objectives include assessments of disease response based on modified IWG criteria or other criteria appropriate for the subject's advanced hematologic malignancy (Section 10.6).

#### 7.3. Rationale for the Dose Selected

##### 7.3.1. Parts I and II: FHD-286 Monotherapy *(Closed to Enrollment)*

The starting dose for the study is based upon results of GLP-compliant toxicity studies performed in rodents and dogs. Daily oral doses of FHD-286 were administered to Wistar Han rats (0.3, 1, and 3 mg/kg/day) and beagle dogs (0.1, 0.3, and 1 mg/kg/day) for 28 days. The highest non-severely toxic dose was >3 mg/kg/day in rats and 1 mg/kg in dogs. Using the highest non-severely toxic dose in the most sensitive species (1 mg/kg in dogs), the equivalent highest non-severely toxic dose in humans was calculated to be 30 mg (using body surface area for a 60-kg human). One-sixth of the human equivalent highest non-severely toxic dose is 5 mg, which is recommended as the starting dose. After correcting for plasma protein binding, predicted human exposure at the starting dose of 5 mg will be 4- to 8-fold or 8- to 17-fold lower than the exposure observed on Day 28 in the 28-day rat (3 mg/kg/day) and dog (1 mg/kg/day) GLP toxicity study, respectively, suggesting that 5 mg is a safe starting dose for first-in-human studies.

##### 7.3.2. Part III: Combination Therapy

Based on changes in peripheral blast levels and markers of myeloid and monocyte maturation in subjects with advanced hematologic malignancies, FHD-286 has shown PD activity at dose levels of 2.5 mg QD, 5 mg QD, 7.5 mg QD, and 10 mg QD, with 2.5 mg QD associated with minimal PD activity. In nonclinical studies in mice, the combination of FHD-286 and either cytarabine or decitabine resulted in enhanced antitumor activity, compared with the single agents.

To date, the highest daily monotherapy FHD-286 dose evaluated and determined to be safe by the CST for Studies FHD-286-C-002 and FHD-286-C-001 is 7.5 mg QD. Taking into consideration the anticipated additive activity and potential for overlapping (eg, myelosuppression) or coinciding toxicities with FHD-286 in combination with LDAC or decitabine, a safety factor of 3 is applied to the highest monotherapy FHD-286 dose determined to be safe (7.5 mg QD), for a proposed FHD-286 starting dose of 2.5 mg QD in combination with LDAC or decitabine for subjects not receiving a triazole antifungal agent classified as a strong CYP3A4 inhibitor at the start of study treatment (Groups A1 and Group B1). Based on preliminary data, FHD-286 exposure is approximately 2- to 3-fold higher in subjects receiving concomitant moderate or strong CYP3A4 inhibitors. Therefore, the proposed FHD-286 starting dose for subjects receiving a triazole antifungal agent classified as a strong CYP3A4 inhibitor at the start of study treatment (Groups A2 and B2) is 1.5 mg QD in combination with LDAC or decitabine.

Based on emerging clinical and nonclinical data, interval dosing of FHD-286, with a higher dose given during the last 14 days of each cycle, may allow subjects to achieve exposures that may result in clinical benefit while improving tolerability. An interval dosing regimen will initially be evaluated in subjects receiving a triazole antifungal agent classified as a strong CYP3A4 inhibitor at the start of study treatment (Groups A2 and B2), and may also be evaluated for subjects not receiving such agents (Groups A1 and B1).

Based on the preliminary safety and dose intensity data for FHD-286 10 mg QD monotherapy in subjects with advanced hematologic malignancies and metastatic uveal melanoma, it is expected that the highest FHD-286 dose to be evaluated in Groups A2 and B2 is 7.5 mg QD. However, if supported by the emerging clinical data and agreed upon by the CST, an FHD-286 dose of 10 mg QD may be evaluated in Groups A1 and B1.

LDAC (cytarabine 20 mg/m$^2$) will be administered SC QD on Days 1 through 10 of each 28-day cycle, a standard route and regimen.

Decitabine 20 mg/m$^2$ IV will be administered QD on Days 1 through 5 of each 28-day cycle, a standard route and regimen.

#### 7.4. Criteria for Study Termination

This study may be prematurely terminated if, in the opinion of the Sponsor, there is sufficiently reasonable cause. In the event of such action, written notification documenting the reason for study termination will be provided to each Investigator.

Circumstances that may warrant termination include, but are not limited to:

- Determination of unexpected, significant, or unacceptable risk to subjects
- Failure to enroll subjects at an acceptable rate
- Insufficient adherence to protocol requirements
- Plans to modify, suspend, or discontinue the development of FHD-286

- Other administrative reasons

Should the study be closed prematurely, all study materials must be returned to the Sponsor or Sponsor's designee.

### 8. STUDY POPULATION

#### 8.1. Number of Subjects

##### 8.1.1. Parts I and II: FHD-286 Monotherapy *(Closed to Enrollment)*

Approximately 25 to 50 subjects will be enrolled in Parts I and II of this study. The sample size will depend on the number of dose groups required and the incidence of DLTs required to determine the RP2D. It is estimated that approximately 25 to 50 subjects will be enrolled in this study assuming 1 subject (Part I) and 3 subjects (Part II) per cohort (6 for the MTD level) and a starting dose of 5 mg daily. Once the RP2D has been identified, an additional 6 to 12 subjects may be enrolled at that dose level(s) to confirm the observed safety and tolerability of FHD-286. Enrollment in this cohort will be paused for review for toxicities observed in 2 out of 6, 3 out of 10, or 4 out of 12 subjects.

##### 8.1.2. Part III: Combination Therapy

The sample size will depend on the incidence of DLTs observed across dose levels and the number of dose levels required to determine the RP2D(s) (Section 12.1). It is estimated that up to approximately 72 subjects in Arm A and up to approximately 72 subjects in Arm B (up to approximately 144 subjects total) will be enrolled. Once the RP2D(s) has been identified for a group, an additional 6 to 14 subjects may be enrolled at that dose level(s) to confirm the observed safety and tolerability of FHD-286 in combination with LDAC or decitabine. Enrollment of these subjects will be paused for review if DLTs are observed in 2 of 6, 3 of 10, or 4 of 14 subjects. Additional stopping criteria will also be considered, as described in Section 9.8.

#### 8.2. Inclusion Criteria

##### 8.2.1. Parts I and II: FHD-286 Monotherapy *(Closed to Enrollment)*

Subjects must meet all of the following criteria to be enrolled in Part I or II of the study:

1. Subject must be ≥16 years of age.
2. Subject must have a confirmed diagnosis of an advanced hematologic malignancy, specified as follows:
    - R/R AML (subjects who relapse after transplantation; subjects in second or later relapse; subjects who are refractory to initial induction or reinduction treatment; subjects who relapse within 1 year of initial treatment; subjects who are otherwise considered relapsed or refractory in the opinion of the Investigator). Subjects with AML must have previously failed all prior therapies known to be active for treatment of their diagnosed hematologic disease.
    - R/R MDS. Subjects with MDS must have previously failed treatment with at least 4 cycles of a hypomethylating agent, known to be active for treatment of their diagnosed hematologic disease.
    - Other R/R advanced hematologic malignancies (for example, CMML). Subjects must have no other reasonable therapeutic option in the opinion of the Investigator.

Subjects who fulfill the inclusion/exclusion criteria may be considered on a case-by-case basis, with approval of the Sponsor.

3. Subject or his/her parent or legal guardian (when applicable) must be able to understand and be willing to sign an informed consent and, when applicable, subject must sign an assent form.
4. Subject must be willing and able to comply with scheduled study visits and treatment plans.
5. Subject must be willing to undergo all study procedures (fresh bone marrow biopsy and/or aspirate at baseline within 28 days of first dose plus bone marrow evaluations every 4 weeks for the first 24 weeks, then every 8 weeks for the next 48 weeks of treatment, as clinically indicated thereafter, and 1 EOT bone marrow evaluation (unless contraindicated due to medical risk; other exceptions to this are at the discretion of the Sponsor), peripheral blood and tissue sampling, and urine sampling during the study.
6. Subject must have an ECOG PS of ≤2.
7. Subject must have a life expectancy of ≥3 months.
8. Subject must have adequate hepatic function as evidenced by:
   - Serum total bilirubin ≤1.5× upper limit of normal (ULN), unless considered due to leukemic involvement following approval by the study Sponsor
   - AST, alanine aminotransferase (ALT), and alkaline phosphatase (ALP) ≤3.0×ULN, unless considered due to leukemic involvement following approval by the study Sponsor
   - Prothrombin time (PT) ≤1.5×ULN or international normalized ratio (INR) ≤1.4
   - Activated partial thromboplastin time (aPTT) ≤1.5×ULN

     Note: Anticoagulation therapy is permitted as long as coagulation parameters are within therapeutic range.
   - No known portal vein thrombosis
9. Subject must have adequate renal function as evidenced by:
   - Creatinine clearance >60 mL/min based on the Cockcroft-Gault glomerular filtration rate (GFR) estimation
10. Subject must have an adequate platelet level, defined as:
    - Platelets $>50\times10^9$/L (transfusions to achieve this level are allowed). Subjects with a baseline platelet count of $\leq50\times10^9$/L due to underlying malignancy are eligible.
11. Subject must have adequate cardiovascular, respiratory, and immune system function as evidenced by the below criteria and in the opinion of the Investigator:
    - LVEF of ≥40% by ECHO (or other means)
12. Subjects must agree to abide by dietary and other considerations required during the study (Section 8.4).

13. Timing requirements with respect to prior therapy and surgery are as follows:
    - At least 2 weeks or at least 5 half-lives, whichever is shorter, must have elapsed since administration of the last dose of any prior systemic anticancer therapy. Hydroxyurea is allowed prior to enrollment and after the start of FHD-286 for the control of peripheral leukemic blasts in subjects with leukocytosis (eg, white blood cell [WBC] counts $>30\times10^9$/L). (Subjects must be intolerant to and/or have experienced disease progression on their prior therapy in the opinion of the treating physician.)
    - At least 4 weeks must have elapsed since the last dose of post-transplant calcineurin inhibitors. Exceptions may be made with Sponsor approval, eg, if waiting 4 weeks to elapse before beginning study drug introduces undue risk of disease progression.
    - Subjects must be recovered from any clinically relevant effects of any prior surgery.
    - At least 2 weeks must have elapsed since the last radiotherapy. Exceptions may be made at the discretion of the Sponsor.
14. Toxicity related to prior therapy must have returned to Grade ≤2 by CTCAE by approximately 14 days prior to study start or be deemed irreversible by the Investigator. Exceptions include alopecia, neuropathy, appropriately controlled endocrine toxicities, and other well controlled/stable toxicities with discussion with the Sponsor.
15. Female subjects must be:
    - postmenopausal, defined as at least 12 months post-cessation of menses (without an alternative medical cause), or
    - permanently sterile following documented hysterectomy, bilateral salpingectomy, bilateral oophorectomy, or tubal ligation or having a male partner with vasectomy as affirmed by the subject, or
    - nonpregnant, nonlactating, and if sexually active having agreed to use a highly effective method of contraception (ie, hormonal contraceptives associated with inhibition of ovulation or intrauterine device [IUD], or intrauterine hormone-releasing system [IUS], or sexual abstinence) from Screening Visit until 90 days after the final dose of the study drug.

    Note: The potential risk to female fertility posed by FHD-286 is unknown; it is recommended that subjects discuss options for fertility preservation with their doctor prior to study start.
16. Male subjects must have documented vasectomy or if sexually active must agree to use a highly effective method of contraception with their partners of childbearing potential (ie, hormonal contraceptives associated with the inhibition of ovulation or IUD, or IUS, or sexual abstinence) from Screening until 90 days after the final dose of the study drug. Male subjects must agree to refrain from donating sperm during this time period.

    Note: The potential risk to male fertility posed by FHD-286 is unknown; it is recommended that subjects discuss options for fertility preservation with their doctor prior to study start.

##### 8.2.2. Part III: Combination Therapy

Subjects must meet all of the following criteria to be enrolled in Part III of the study:

1. Subject must be ≥16 years of age.
2. Subject must:
   - Have a confirmed diagnosis of R/R AML, R/R MDS, or R/R CMML not in blast crisis

     AND
   - Have received ≤4 prior lines of systemic anticancer therapy for their disease under study; subjects who have received >4 prior lines of systemic anticancer therapy for their disease under study must receive Sponsor approval.

     AND
   - Be an appropriate candidate for treatment with LDAC (Arm A) or decitabine (Arm B)
3. Subject or their parent or legal guardian (when applicable) must be able to understand and be willing to sign an informed consent and, when applicable, subject must sign an assent form.
4. Subject must be willing and able to comply with scheduled study visits and treatment plans.
5. Subject must be willing to undergo all study procedures (including bone marrow biopsies, peripheral blood sampling, and urine sampling) unless contraindicated due to medical risk.
6. Subject must have an ECOG PS of ≤2.
7. Subject must have a life expectancy of ≥3 months.
8. Subject must have adequate hepatic function as evidenced by:
   - Serum total bilirubin ≤1.5× ULN, unless considered due to advanced hematologic malignancy involvement or Gilbert syndrome, following approval by the study Sponsor
   - AST, ALT, and ALP ≤3.0×ULN, unless considered due to advanced hematologic malignancy involvement, following approval by the study Sponsor
   - Prothrombin time (PT) ≤1.5×ULN or INR ≤1.4
   - aPTT ≤1.5×ULN

     Note: Sponsor approval is required for a subject receiving anticoagulation therapy for treatment of a stable medical condition whose coagulation parameters are within therapeutic range.
   - No known portal vein thrombosis

- No history of azole-induced liver dysfunction (if being considered for participation in Group A2 or B2 [subjects receiving a triazole antifungal agent classified as a strong CYP3A4 inhibitor])

9. Subject must have adequate renal function as evidenced by:
   - GFR ≥60 mL/min (based on a contemporary, widely accepted, and clinically applicable equation that estimates glomerular filtration rate or a measure of glomerular filtration rate)
10. Subject must have a WBC count ≤$20\times10^9$/L; treatment with a stable dose of hydroxyurea or other cytoreductive agent (eg, cytarabine) to achieve this count is allowed.
11. Subject must have adequate cardiovascular, respiratory, and immune system function as evidenced by the below criteria and in the opinion of the Investigator:
    - LVEF of ≥40% by ECHO (or other means)
12. Subject must agree to abide by dietary and other considerations required during the study (Section 8.4).
13. Timing requirements with respect to prior therapy and surgery are as follows:
    - Prior systemic anticancer therapy (including investigational drugs):
      - Not a small molecule inhibitor: At least 2 weeks or at least 5 half-lives, whichever is shorter, must have elapsed since administration of the last dose.
      - Small molecule inhibitor (eg, FLT3 kinase inhibitor, isocitrate dehydrogenase [IDH]1/IDH2 inhibitor), *except venetoclax*: At least 72 hours must have elapsed since administration of the last dose.
      - Venetoclax: At least 2 weeks must have elapsed since administration of the last dose.
      - Treatment with hydroxyurea (or other cytoreductive agent) is allowed during the washout period for blast control (as per inclusion criterion 10).
    - Post-transplant calcineurin inhibitors: At least 4 weeks must have elapsed since administration of the last dose.
    - Radiotherapy: At least 2 weeks must have elapsed since the last radiotherapy.
    - Surgery: Subject must be recovered from any clinically relevant effects of any prior surgery.
14. Toxicity related to prior therapy must have returned to Grade ≤2 by CTCAE by approximately 14 days before the start of study treatment or be deemed irreversible and stable by the Investigator. Exceptions include alopecia, neuropathy, appropriately controlled endocrine toxicities, and other well-controlled/stable toxicities with discussion with the Sponsor.
15. Female subjects must be:
    - postmenopausal, defined as at least 12 months post-cessation of menses (without an alternative medical cause), or

- permanently sterile following documented hysterectomy, bilateral salpingectomy, bilateral oophorectomy, or tubal ligation, or, if sexually active with male partners, these partners must be azoospermic (vasectomized or due to a medical cause), as affirmed by the subject, or
- nonpregnant, nonlactating, and, if sexually active with fertile male partners, having agreed to use a highly effective method of contraception (ie, hormonal contraceptives associated with inhibition of ovulation or IUD, or IUS, or sexual abstinence) from Screening Visit until the latest of the following:
  - All subjects: 90 days after the final dose of FHD-286
  - Subjects in Arm A (FHD-286 + LDAC): 6 months after the final dose of LDAC
  - Subjects in Arm B (FHD-286 + decitabine): 6 months after the final dose of decitabine

Note: The potential risk to female fertility posed by FHD-286 is unknown; it is recommended that subjects discuss options for fertility preservation with their doctor before the start of study treatment.

Note: Subjects in Arm B: Because of the possibility of infertility as a consequence of decitabine therapy, subjects should seek consultation regarding oocyte cryopreservation before the initiation of treatment.

16. Male subjects must have documented azoospermia (vasectomized or due to a medical cause) or, if fertile and sexually active, must agree to use a highly effective method of contraception with their partners of childbearing potential (ie, hormonal contraceptives associated with the inhibition of ovulation or IUD, or IUS, or sexual abstinence) from Screening until the latest of the following:
    - All subjects: 90 days after the final dose of FHD-286
    - Subjects in Arm A (FHD-286 + LDAC): 6 months after the final dose of LDAC
    - Subjects in Arm B (FHD-286 + decitabine): 90 days after the final dose of decitabine

    Male subjects must agree to refrain from donating sperm during this time period.

    Note: The potential risk to male fertility posed by FHD-286 is unknown; it is recommended that subjects discuss options for fertility preservation with their doctor before the start of study treatment.

    Note: Subjects in Arm B: Because of the possibility of infertility as a consequence of decitabine therapy, subjects should seek consultation regarding conservation of sperm before the initiation of treatment.

#### 8.3. Exclusion Criteria

##### 8.3.1. Parts I and II: FHD-286 Monotherapy *(Closed to Enrollment)*

Subjects who meet any of the following criteria will not be enrolled in Parts I and II of the study:

1. Subject (or parent or legal guardian, when applicable) is unable to provide informed consent (or assent, when applicable) and/or to follow protocol requirements.
2. Subject has undergone HSCT within 60 days of the first dose of FHD-286, or subject has clinically significant graft-versus-host disease (GVHD). The use of a stable dose of oral steroids and/or immunosuppressive therapy post-HSCT is permitted with Sponsor approval.
3. Subject has clinical symptoms suggesting active central nervous system (CNS) leukemia or known CNS leukemia. Evaluation of the cerebrospinal fluid is only required if there is a clinical suspicion of CNS involvement by leukemia during screening.
4. Subject has an immediately life-threatening, severe complications of leukemia, such as uncontrolled bleeding, pneumonia with hypoxia or shock, and/or disseminated intravascular coagulation.
5. Subject has other malignancy which may interfere with the diagnosis and/or treatment of advanced hematologic malignancies.
6. Subject has active hepatitis B virus (HBV) or hepatitis C virus (HCV) infections; subjects with a sustained viral response to HCV treatment or immunity to prior HBV infection will be permitted. Subject has known positive HIV antibody results, or acquired immunodeficiency syndrome (AIDS)-related illness; subjects with CD4+ T-cell counts ≥350 cells/µL will be permitted, as will subjects who have not had an AIDS-related illness within the past 12 months.
7. Subject has an active severe infection that requires anti-infective therapy or has an unexplained temperature of >38.5°C during screening visits or on their first day of study drug administration (at the discretion of the Investigator, subjects with tumor fever may be enrolled).
8. Subject has an uncontrolled intercurrent illness.
9. Subjects with corrected QT interval (QTc) using Fridericia's formula (QTcF) >470 msec or other factors that increase the risk of QTc prolongation or arrhythmic events (eg, heart failure, hypokalemia, family history of long QT interval syndrome) including heart failure that meets New York Heart Association (NYHA) class III and IV definitions (see Appendix 15.3) are excluded. Subjects with bundle branch block and a prolonged QTc should be reviewed by the Sponsor for potential inclusion.
10. Subject has any other medical or psychological condition, deemed by the Investigator to be likely to interfere with a subject's ability to sign informed consent/assent, cooperate, or participate in the study.
11. Subject has known allergies or hypersensitivities to components of the FHD-286 formulation.
12. Subject is unable to tolerate the administration of oral medication or has GI dysfunction that could interfere with absorption of FHD-286 (eg, ulcerative disease, uncontrolled nausea, vomiting, diarrhea, malabsorption syndrome, partial bowel resections).
13. Subject is receiving any other investigational agents.

14. At least 2 weeks or 5 half-lives, whichever is shorter, must have elapsed since last administration of a prior investigational drug at the start of study treatment. Exceptions include participation in any observational or nontherapeutic clinical trials.
15. Subject is on medications that are strong CYP3A inhibitors, are strong CYP3A inducers, or are sensitive CYP3A substrates with narrow TIs (see Appendix 15.4). Exceptions may be made for therapy in the case of life-threatening infections, for example a triazole anti-fungal agent to reduce the risk of invasive fungal infections, at the discretion of the Sponsor. See Section 9.12.2 for further detail.
16. Subject is on medications with narrow TIs that are sensitive P-gp or BCRP substrates and are administered orally, such as digoxin (see Appendix 15.5) or on medications that are strong inhibitors of P-gp or BCRP (see Appendix 15.6).
17. Administration of PPIs should be stopped or switched to another acid-reducing agent (ARA; eg, antacids or histamine H2-receptor antagonists [H2] blockers) 7 days before administration of study drug. In the event that it is medically necessary to dose PPIs concomitantly with FHD-286, this may be permitted with Sponsor approval. See Section 9.12.2 for further detail.
18. Subject is requiring clinically significant or increasing doses of systemic steroid therapy or any other systemic immunosuppressive medication. The use of a stable dose of systemic steroids and/or immunosuppressive medication is permitted with Sponsor approval. Local or targeted steroid and immunosuppressive therapies (eg, inhaled or topical steroids) are acceptable. Appropriate steroid replacement to manage endocrine toxicities resulting from prior anticancer systemic therapy is permitted.
19. Subject has undergone any prior treatment with a BRG1/BRM inhibitor.
20. Subject is pregnant or breastfeeding or is planning to become pregnant within 1 year of study start. Subject is a woman or man of childbearing capabilities who is unwilling to use effective contraception.

##### 8.3.2. Part III: Combination Therapy

Subjects who meet any of the following criteria will not be enrolled in Part III of the study:

1. Subject (or parent or legal guardian, when applicable) is unable to provide informed consent (or assent, when applicable) and/or to follow protocol requirements.
2. Subject:
   - Has undergone chimeric antigen receptor T cell therapy or HSCT within 60 days of the first dose of study treatment

     OR
   - Has clinically significant GVHD
     – Clinically significant GVHD is defined as signs or symptoms of acute or chronic GVHD Grade >0, per Mount Sinai Acute GvHD International Consortium or National Institutes of Health criteria, respectively, within 4 weeks before the first dose of study treatment.

3. Subject has evidence (or suspicion) of extramedullary involvement, unless approved by Sponsor.
4. Subject has an immediately life-threatening, severe complication(s) of advanced myeloid malignancy, such as uncontrolled bleeding, pneumonia with hypoxia or shock, and/or disseminated intravascular coagulation.
5. Subject has other malignancy that may interfere with the diagnosis and/or treatment of advanced hematologic malignancies.
6. Subject has active HBV or HCV infections; subjects with a sustained viral response to HCV treatment or immunity to prior HBV infection will be permitted. Subject has known positive HIV antibody results, or AIDS-related illness; subjects with CD4+ T-cell counts ≥350 cells/µL will be permitted, as will subjects who have not had an AIDS-related illness within the past 12 months.
7. Subject has an active severe infection that requires anti-infective therapy or has an unexplained temperature of >38.5°C during screening visits or on their first day of study treatment (at the discretion of the Investigator, subjects with tumor fever may be enrolled).
8. Subject has an uncontrolled intercurrent illness.
9. Subject has QTcF >470 msec or other factors that increase the risk of QTc prolongation or arrhythmic events (eg, heart failure, hypokalemia, family history of long QT interval syndrome) including heart failure that meets NYHA class III and IV definitions (see Appendix 15.3). Subjects with QTcF >470 msec and bundle branch block and/or pacemaker rhythm may be enrolled after approval by the Sponsor.
10. Subject has any other medical or psychological condition, deemed by the Investigator to be likely to interfere with a subject's ability to sign informed consent/assent, cooperate, or participate in the study.
11. Subject has known allergies or hypersensitivities to:
    - All subjects: components of the FHD-286 formulation (refer to the FHD-286 IB)
    - Arm A: cytarabine or any of the excipients (refer to the approved drug label [eg, United States prescribing information (USPI), summary of product characteristics (SmPC)])
    - Arm B: decitabine or any of the excipients (refer to the approved drug label [eg, USPI, SmPC])
12. Subject is unable to tolerate the administration of oral medication or has GI dysfunction that would preclude adequate absorption, distribution, metabolism, or excretion of FHD-286.
13. Subject is receiving any other anticancer investigational agents. Investigational agents to treat non-cancer indications may be permitted with Sponsor approval.
14. Removed in protocol version 5.0.
15. Subject is on medications classified as:

- Strong CYP3A inhibitors; see also Section 9.12.2 and Appendix 15.4
  - Exception: Triazole antifungal agents, including those classified as strong CYP3A inhibitors, are permitted. Subjects receiving triazole antifungal agents classified as strong CYP3A inhibitors at start of study treatment will be assigned to Group A2 or B2. See Section 9.12.5.
- Strong CYP3A inducers; see also Section 9.12.2 and Appendix 15.4
- Sensitive CYP3A substrates with narrow TIs; see also Section 9.12.2 and Appendix 15.4
  - Stable doses of immunosuppressant medications that are sensitive CYP3A substrates may be permitted with Sponsor approval.

16. Subject is on medications with narrow TIs that are sensitive P-gp or BCRP substrates and are administered orally, such as digoxin (see Appendix 15.5), or on medications classified as strong inhibitors of P-gp or BCRP (see Appendix 15.6).
17. Administration of PPIs should be stopped or switched to another ARA (eg, antacids or H2 blockers) 7 days before administration of FHD-286. In the event that it is medically necessary to dose PPIs concomitantly with FHD-286, this may be permitted with Sponsor approval. See Section 9.12.2 for further detail.
18. Subject is requiring clinically significant or increasing doses of systemic steroid therapy or any other systemic immunosuppressive medication. The use of a stable dose of systemic steroids and/or immunosuppressive medication is permitted with Sponsor approval. Local or targeted steroid and immunosuppressive therapies (eg, inhaled or topical steroids) are acceptable. Appropriate steroid replacement to manage endocrine toxicities resulting from prior anticancer systemic therapy is permitted. See exclusion criterion 15 for exclusions regarding medications classified as strong CYP3A inducers or sensitive CYP3A substrates with narrow TIs.
19. Subject has undergone any prior treatment with a BRG1/BRM inhibitor.
20. Subject is pregnant or breastfeeding or is planning to become pregnant within 1 year of the start of study treatment.

#### 8.4. Lifestyle Considerations

Subjects must discontinue intake of beverages, herbal supplements, or food known to inhibit or induce CYP3A, including grapefruit, grapefruit products, Seville oranges, starfruit, St John's wort, echinacea, and goldenseal, from 72 hours before the first FHD-286 dose until 72 hours after the last FHD-286 dose.

FHD-286 may have phototoxic potential; as such, subjects should be warned to avoid direct sun exposure or exposure to UV light. When exposure to sunlight is anticipated for longer than 15 minutes, the subject should be instructed to apply factor 30 or higher sunscreen to exposed areas and wear protective clothing and sunglasses.

#### 8.5. Subject Identification and Registration

Subjects who are candidates for enrollment into the study will be evaluated for eligibility by the Investigator to ensure that the inclusion and exclusion criteria (see Section 8.2 and Section 8.3, respectively) have been satisfied and that the subject is eligible for participation in this clinical study. Subjects will be assigned a subject number upon consent/assent.

#### 8.6. Discontinuation of Study Treatment, Subject Withdrawal Criteria, and Replacement of Subjects

Subjects have the right to withdraw from study treatment at any time for any reason. Subjects may withdraw or be withdrawn from study treatment under the following conditions:

- Withdraws consent/assent for study treatment
- Experiences unacceptable toxicity. Such subjects will discontinue treatment though will remain in the study for long-term follow-up.
- The Investigator removes the subject from the study in the best interests of the subject for any reason, including any medical condition that, in the opinion of the Investigator, would put the subject at risk for continuing treatment.
- Development of disease progression or treatment failure, based on disease response assessments (subjects who are, in the opinion of the Investigator, benefitting from treatment may be allowed to continue on study treatment with the approval of the Sponsor)
    - In Part III (combination therapy), subjects who discontinue LDAC or decitabine for reasons other than treatment failure or progressive disease may continue FHD-286 monotherapy until any of the withdrawal criteria in Section 8.6 are met.
- Development of an intercurrent medical condition or need for a concomitant medication that precludes further participation in the study or that, in the opinion of the Investigator, would pose a safety risk to the subject (Section 9.12)
- Start of alternative anticancer therapy
- Noncompliance with protocol requirements
- Becomes pregnant
- Lack of efficacy (failure to achieve a partial remission [Section 10.6] or better by 6 months on treatment)

Should a subject decide to withdraw from treatment, all efforts will be made to determine the reason for withdrawal.

Subjects have the right to withdraw from the study at any time for any reason. Subjects must be withdrawn from the study if the subject:

- Withdraws consent/assent
- Is lost to follow-up
- Dies

In the event a subject is withdrawn from the study, the Sponsor must be informed.

When a subject is withdrawn from study treatment, but has not withdrawn consent/assent to participate in the study, the Investigator should make every effort to have the subject complete all end-of-treatment assessments and all scheduled follow-up visits (Safety Follow-up Visit, Long-Term Follow-up).

When a subject is withdrawn from study treatment due to unacceptable toxicity, the subject should be followed to monitor for resolution or stabilization of AEs. Refer to Section 11 for further details on AE collection, reporting, and monitoring procedures.

When a subject withdraws from the study, the primary reason for discontinuation must be recorded in the appropriate section of the electronic case report form (eCRF) and all efforts will be made to complete and report final study observations as thoroughly as possible.

Subjects who do not meet the minimum treatment and safety evaluation requirements for evaluation of DLT (Section 9.6.1) and who do not experience DLT will be replaced if the minimum of 3 evaluable subjects within a dose cohort has not been met.

### 9. STUDY TREATMENT

Study treatment is defined as any individual investigational intervention(s) or marketed product(s) intended to be administered to a study subject according to this study.

The combination agents to be administered in this study are considered standard of care for the advanced hematologic malignancies under study and therefore will be obtained commercially by the site.

#### 9.1. Description of Study Drug

FHD-286 will be supplied as 1.5 mg, 2.5 mg, and 5 mg strength capsules. FHD-286 is for investigational use only and is to be used only within the context of this study. All FHD-286 drug product will be supplied by the Sponsor. See the FHD-286 IB for further details.

#### 9.2. Study Drug Packaging and Labeling

FHD-286 capsules will be supplied in appropriate containers with child-resistant closures and will be labeled appropriately as investigational product for this study. Packaging and labeling will be prepared to meet all regulatory requirements.

#### 9.3. Study Drug Storage

Bottles of FHD-286 capsules must be stored according to the package label.

All study drug product must be stored in a secure, limited-access location and may be dispensed only be the Investigator or by a member of the staff specifically authorized by the Investigator.

#### 9.4. Randomization and Blinding

This is a nonrandomized open-label study.

#### 9.5. Study Drug Preparation, Dispensation, and Administration

##### 9.5.1. FHD-286

Treatment with FHD-286 will be administered as a single agent, or in combination with either LDAC or decitabine, dosed orally on Days 1 through 28 of a 28-day cycle. On days when FHD-286 is administered in combination with LDAC or decitabine, FHD-286 must be administered before the combination agent.

The initial FHD-286 dosing regimen will be QD (approximately every 24 hours) without planned drug holidays.

Table 2 lists the proposed dose levels for Part III. For the FHD-286 5 mg QD and 7.5 mg QD dose groups in Groups A2 and B2, interval dosing will be implemented, with a 2.5/5 mg QD dose given on Days 1 through 14 of each cycle, and the 5/7.5 mg QD doses given on Days 15 through 28 of each cycle. Starting at C2D1, participants may continue at the higher dose (ie, transition to a consistent dose) if it is tolerated in the judgment of the Investigator, upon agreement from the Sponsor. Interval dosing may also be implemented for Groups A1 and B1 if agreed upon by the CST. Alternative interval dosing regimens may also be considered, including

"1 week low/1 week high" (FHD-286 1.5 mg QD×7 days, FHD-286 5/7.5 mg QD×7 days). Dose escalation decisions will be made by the CST as described in Section 9.6.2.2, and it is possible that not all dose levels listed in the table will be evaluated, and that intermediate dose levels not listed may be evaluated. It is expected that the highest FHD-286 dose to be evaluated in Groups A2 and B2 is 7.5 mg QD; however, if supported by the emerging clinical data and agreed upon by the CST, an FHD-286 dose of 10 mg QD may be evaluated in Groups A1 and B1.

**Table 2: Part III: Proposed FHD-286 Dose Levels**

| Groups A1 and B1 | Groups A2 and B2 |
|---|---|
| 2.5 mg QD | 1.5 mg QD |
| 5 mg QD | 2.5 mg QD |
| 7.5 mg QD | 5 mg QD |
| 10 mg QD | 7.5 mg QD |

Abbreviations: C = Cycle; CST = Clinical Study Team; D = Day; QD = once daily.
Note: Groups A1 and B1: subjects not receiving a triazole antifungal agent classified as a strong CYP3A4 inhibitor at the start of study treatment. Groups A2 and B2: subjects receiving a triazole antifungal agent classified as a strong CYP3A4 inhibitor at the start of study treatment.
For the FHD-286 5 mg QD and 7.5 mg QD dose groups in Groups A2 and B2, interval dosing will be implemented, with a 2.5/5 mg QD dose given on Days 1 through 14 of each cycle, and the 5/7.5 mg QD doses given on Days 15 through 28 of each cycle. Starting at C2D1, participants may continue at the higher dose if it is tolerated in the judgment of the Investigator, upon agreement from the Sponsor. Interval dosing may also be implemented for Groups A1 and B1 if agreed upon by the CST. Alternative interval dosing regimens may also be considered, including "1 week low/1 week high" (FHD-286 1.5 mg QD×7 days, FHD-286 5/7.5 mg QD×7 days).

All doses of FHD-286 should be taken under fasted conditions (Section 9.5.1.1). Subjects should take their daily dose at approximately the same time each morning. Each dose should be taken with a glass of water and consumed over as short of a time as possible. Subjects should be instructed to swallow capsules whole and to not chew the capsules.

If the subject forgets to take the daily dose, then they should take FHD-286 within 12 hours after the missed dose. If more than 12 hours have elapsed, then that dose should be omitted, and the subject should resume treatment with the next scheduled dose. If a subject vomits after taking their dose, the subject should not retake their dose and should simply resume their dosing regimen at their next scheduled dose.

The appropriate number of Sponsor-packaged, labeled bottles will be dispensed to subjects to allow for dosing for a full cycle; alternatively, they may be dispensed appropriate bottle(s) until the next scheduled visit. Subjects are to return all unused capsules (or the empty bottles) on Day 1 of each treatment cycle or at the next scheduled visit.

###### 9.5.1.1. Fasted Conditions

After a fast of at least 2 hours, subjects should take their dose of FHD-286 with approximately 240 mL (8 fluid ounces) of water. Subjects may have a low-fat meal before the fast (Table 3). Additional water is permitted ad libitum. Subjects should not consume any food for at least 2 hours after the dose has been administered.

The time of fasting required before and after dose administration may change based on emerging data.

**Table 3: Meal Definitions**

| Meal Type[a] | Total Kcal | Fat | | |
|---|---|---|---|---|
| | | Kcal | Grams | Percent |
| Low-fat | 400-500 | 100-125 | 11-14 | 25 |

a Table derived from Appendix 2 of the FDA Draft Guidance: "Assessing the Effects of Foods on Drugs in INDs and NDAs – Clinical Pharmacology Considerations" (February 2019).

An example of a light (ie, low-fat) breakfast includes 8 ounces of 1% milk, 1 boiled egg, and 1 packet of flavored instant oatmeal made with water.

###### 9.5.2. LDAC

LDAC (cytarabine 20 mg/$m^2$) will be administered SC QD on Days 1 through 10 of all cycles. For Days 1 through 5 of Cycle 1, LDAC will be administered at the study site. After the first 5 doses (ie, beginning on C1D6), at the Investigator's discretion and where consistent with institutional, state, and local guidelines and regulations, subjects may self-administer LDAC at home, except on scheduled visit days. For subjects self-administering LDAC at home, prefilled syringes will be dispensed to subjects as per institutional practice.

LDAC will be commercially sourced by each site. All supply should be handled, prepared, stored, and administered in accordance with the approved drug label (eg, USPI, SmPC).

###### 9.5.3. Decitabine

Decitabine 20 mg/$m^2$ will be administered IV QD on Days 1 through 5 of all cycles.

Decitabine will be commercially sourced by each site. All supply should be handled, prepared, stored, and administered in accordance with the approved drug label (eg, USPI, SmPC).

###### 9.5.4. Study Subject Diary

Subjects will be given a dosing diary for each treatment cycle. They should record relevant information regarding FHD-286 and, if applicable, self-administered LDAC in the diary (eg, confirmation that each dose was taken, reasons for missed doses, confirmation of fasted condition [FHD-286 only], whether food was consumed within required fasting times before and after each dose [FHD-286 only], time of administration).

The dosing diary will be used to assess treatment compliance for FHD-286 and self-administered LDAC (Section 9.10).

##### 9.6. Dose Modification Criteria

###### 9.6.1. Dose-Limiting Toxicity

Dose-limiting toxicities will be evaluated throughout the duration of treatment. The primary DLT evaluation period is Cycle 1 (ie, 28 days) of treatment (or during the first 42 days of treatment for participants in Part III who transition from interval dosing during Cycle 1 to a consistent dose at C2D1 employing the highest of the 2 doses administered during Cycle 1 of interval dosing [Section 9.5.1]). However, if a subject experiences a toxicity starting after the DLT evaluation period that warrants assessment as to whether the event constitutes a DLT, this will be evaluated by the CST. Toxicities will be graded and documented according to the NCI CTCAE, version 5.0 (see Appendix 15.1). A DLT is defined as any AE that occurs during the DLT evaluation period that also meets any 1 of the following criteria as determined by the CST:

**Non-hematologic:**

- Any Grade 4 Differentiation syndrome toxicity
- Any Grade ≥3 toxicities not clearly resulting from the underlying advanced hematologic malignancy EXCEPT:
  - Grade 3 fatigue, asthenia, fever, or anorexia (Grade 3 anorexia can be excluded only if it does not result in hospitalization, tube-feeding or use of total parenteral nutrition [TPN])
  - Grade 3 or 4 infection responding to appropriate antimicrobial therapy within 7 days of treatment initiation
  - Grade 3 constipation, nausea, vomiting, or diarrhea not requiring tube feeding, TPN, hospitalization, or prolongation of current hospitalization
  - Grade 3 or 4 tumor lysis syndrome if it is successfully managed and resolves clinically within 7 days without end-organ damage
  - Grade 3 or 4 toxicity associated with lysis or elimination of extramedullary leukemic tissue if it is successfully managed clinically, resolves/improves, and does not result in chronic or permanent end-organ damage while the subject is on study
  - Grade 3 or 4 isolated electrolyte abnormalities resolving to Grade ≤2 within 72 hours with appropriate management
- Any Hy's law cases

**Hematologic:**

- Any Grade 4 ANC, febrile neutropenia, or platelets lasting >28 days, in the absence of active disease

In addition, the following will be considered a DLT:

**Other**:

- Any toxicities related to FHD-286 that lead to permanent discontinuation of FHD-286

NOTE: Toxicities meeting any of the above criteria with a clear-cut alternative explanation (eg, due to disease progression) will not be considered DLTs.

All AEs that cannot clearly be determined to be unrelated to FHD-286 or the combination of FHD-286 with LDAC or decitabine will be considered relevant to determining DLTs and will be reviewed by the CST.

The CST also will review any other emergent toxicities that are not explicitly defined by the DLT criteria to determine if any warrant a DLT designation.

##### 9.6.2. Dose Escalation Procedures

###### 9.6.2.1. Parts I and II: FHD-286 Monotherapy *(Closed to Enrollment)*

Dose escalation for this study will occur in two parts; Part I will utilize single-subject cohorts and Part II will utilize a "3+3" design (Section 7.1.1.1).

Study drug doses will be escalated sequentially following review of the safety data collected during Cycle 1 from subjects at the current dose level. Decisions to escalate to the next higher dose will be made by mutual agreement with the CST, comprised of the Sponsor (Responsible Medical Officer), the Medical Monitor, and the Investigators. Regularly scheduled teleconferences will serve as a forum for review of safety, tolerability, PK, and other relevant data (preliminary clinical and PD data, if available). Decisions to escalate the dose will be documented along with a rationale statement for the decision and summary of the information supporting the decision.

A minimum of 1 (Part I) or 3 (Part II) evaluable subjects in the dose group must have completed study drug dosing for Cycle 1 (28 days) to conduct the safety review for a decision to be made regarding escalation to the next dose level. Evaluable subjects are those who meet the minimum treatment and safety evaluation requirements of the study and/or who experience a DLT during Cycle 1. The minimum treatment and safety evaluation requirements are met if, in Cycle 1, the subject has received approximately 80% or more of the planned doses of FHD-286, and/or is considered by the CST to have sufficient safety data available to conclude that a DLT did not occur. Subjects who do not meet these minimum treatment and safety evaluation requirements and who do not experience a DLT will be replaced if the minimum of 3 evaluable subjects has not been satisfied.

The decision to dose-escalate may be made after the first (Part I) or third (Part II) evaluable subject enrolled to the dose level under review has completed the first cycle of treatment.

In summary:

In Part I of dose escalation:

- If after 1 subject has completed Cycle 1 dosing at a single dose level:
  - The subject does not experience a DLT, then enrollment of the next single-subject cohort may commence following safety review by the CST.
  - The subject experiences a DLT, then that cohort will enroll new subjects to reach n=6 and the Dose Escalation will transition to a "3+3" design.
- If after 2 (or more) subjects have completed Cycle 1 dosing, at various dose levels:
  - 2 subjects experience FHD-286-related non-DLT toxicities that are Grade ≥2, then the cohort at the highest dose level will enroll new subject(s) to reach n=3 and the Dose Escalation will transition to a "3+3" design.

In Part II of dose escalation, if after at least 3 evaluable subjects have completed Cycle 1 (28 days) dosing:

- None of the 3 subjects experience a DLT, then enrollment of the next dose cohort may commence following safety review by the CST.
- 1 of the 3 subjects within a dose cohort experiences a DLT, then additional subjects are to be enrolled at that dose level until the cohort has a total of 6 subjects. If none of the additional subjects has a DLT (ie, 1 of 6 subjects has a DLT), then enrollment at the next scheduled dose may commence following safety review by the CST.
- If ≥2 of the 3 subjects within a dose cohort experience a DLT, then the previous dose level will be declared the MTD, as long as <2 of 6 subjects experiences a DLT at that dose level. Alternatively, a dose level intermediate between the non-tolerated dose level and the previously tolerated dose level may be explored and declared the MTD if <2 out of 6 subjects experience a DLT at that dose.

Increases in the dose of FHD-286 for each escalation cohort will be guided by an accelerated titration design. The absolute percent increase in the daily dose will be determined by the CST based on the available safety, toxicity, clinical activity, PK, and/or PD data (but will never exceed 100%). Escalation will continue in this manner until the RP2D is determined. If warranted based on the emerging data, an alternative dosing schedule may be explored as agreed upon by the CST.

In Part II, if there are multiple subjects in the screening process at the time the third subject within a dose cohort begins treatment, up to 2 additional subjects may be enrolled in that cohort, with approval of the Sponsor.

Subjects initiated on doses of FHD-286 that have already been deemed safe by the CST will not count toward the DLT criteria for dose escalation.

###### 9.6.2.2. Part III: Combination Therapy

Part III will utilize a "3+3" design (Section 7.1.1.2). Dose escalation will proceed independently for each of the 4 groups in this study: A1, A2, B1, and B2.

FHD-286 doses will be escalated sequentially after review of the safety, tolerability, PK, and PD data collected during the DLT evaluation period from subjects at the current dose level, in the context of and in addition to other relevant data. Decisions to escalate to the next higher FHD-286 dose level will be made by mutual agreement with the CST, comprising, at minimum, the Sponsor (Responsible Medical Officer) and the Investigators. Regularly scheduled teleconferences will serve as a forum for review of safety and other relevant data by the CST. Decisions to escalate the FHD-286 dose level or to implement an interval dosing regimen for Groups A1 and B1 will be documented along with the rationale for the decision and a summary of the information supporting the decision.

At least 3 evaluable subjects at a given dose level must have completed FHD-286 dosing for the DLT evaluation period in order to conduct the safety review in support of a decision regarding escalation to the next dose level. Evaluable subjects are those who meet the minimum treatment and safety evaluation requirements of the study, as defined below, and/or who experience a DLT during the DLT evaluation period.

Minimum treatment and safety evaluation requirements:

- During the DLT evaluation period, the subject received:
  - Approximately 80% or more of each planned dose of FHD-286
    **and**
  - Approximately 80% or more of the planned doses of LDAC/decitabine
    **and**
  - For Groups A2 and B2 only, at least 14 days of treatment with a triazole antifungal agent classified as a strong CYP3A4 inhibitor

and/or

- Is considered by the CST to have sufficient safety data available to conclude that a DLT did not occur

Subjects who do not meet these minimum treatment and safety evaluation requirements and who do not experience a DLT will be replaced if the minimum of 3 evaluable subjects has not been met.

The decision to dose-escalate may be made after the third evaluable subject enrolled to the dose level under review has completed the DLT evaluation period.

In summary, if after at least 3 evaluable subjects in a dose group have completed the DLT evaluation period:

- None of the 3 subjects experience a DLT, then the FHD-286 dose may be escalated and enrollment of the next dose level may begin, after approval by the CST
- 1 of the 3 subjects experiences a DLT, then additional subjects are to be enrolled at that dose level until the dose group has a total of 6 evaluable subjects. If none of the additional subjects has a DLT (ie, 1 of 6 subjects has a DLT), then the FHD-286 dose may be escalated and enrollment of the next FHD-286 dose level may begin, after approval by the CST.
- ≥2 of the 3 subjects experience a DLT, then the previous dose level will be declared the MTD, as long as <2 of 6 subjects experience a DLT at that dose level. Alternatively, an FHD-286 dose level between the non-tolerated dose level and the previous tolerated dose level may be explored and declared the MTD if <2 of 6 subjects experience a DLT at that dose.

The absolute percent increase in the daily dose will be determined by the CST based on the available safety, toxicity, preliminary clinical activity, PK, and/or PD data (but will never exceed 100%). Escalation will continue in this manner until the presumptive RP2D(s) is determined; RP2D determination will occur independently for each of the 4 groups: A1, A2, B1, and B2. Interval dosing (Section 9.5.1) may be implemented for Groups A1 and B1 if agreed upon by the CST.

Additional subjects may be enrolled for evaluation of alternative dosing regimens, or for further exploring safety, PK, PK/PD, or preliminary clinical activity used to guide the selection of the RP2D.

It is expected that the highest FHD-286 dose to be evaluated in Groups A2 and B2 is 7.5 mg QD; however, if supported by the emerging clinical data and agreed upon by the CST, an FHD-286 dose of 10 mg QD may be evaluated in Groups A1 and B1.

The RP2D determination will consider dose intensity across the evaluated dose levels and regimens.

###### 9.6.3. Intra-subject Dose Escalation Criteria

###### 9.6.3.1. Parts I and II: FHD-286 Monotherapy *(Closed to Enrollment)*

Intra-subject dose escalation may be permitted following discussion with the Sponsor. Specifically, with Sponsor approval, subjects who are receiving a dose level (dose level is defined as dose and regimen) of FHD-286 that has been cleared as defined in Section 9.6.2 may be escalated to a higher dose level that has been cleared by the CST, and at which there have been no DLTs. Subjects who experienced Grade ≤2 toxicities at their current dose are eligible for intra-subject dose escalation. Subjects who experienced a DLT at their current dose level will not be eligible for intra-subject dose escalation. There is no limit to the number of times the dose level of FHD-286 may be increased for a given eligible subject following Sponsor approval as long as the next higher dose level has been cleared.

###### 9.6.3.2. Part III: Combination Therapy

Intra-subject dose escalation is not permitted.

###### 9.6.4. Dose Interruptions and Dose Reductions

Subjects who experience a DLT as defined in Section 9.6.1 will have dosing with study treatment at least temporarily interrupted, and potentially discontinued. Subjects who experience a DLT who are, in the opinion of the Investigator, benefiting from treatment may be allowed to restart FHD-286 at a reduced dose level with approval of the Sponsor. If the duration of dose interruption required for recovery from a DLT (ie, return to Grade ≤2 toxicity and/or at least baseline levels) is more than 28 days (1 cycle), a subject’s continuation in the study, based on the risks and benefits thereof, should be discussed with the Sponsor.

Dose reductions and/or interruption of dosing may be allowed in the event of non-DLTs that are assessed as related to treatment with FHD-286, after discussion with the Sponsor.

If the time required for recovery from toxicity (ie, return to at least baseline levels; exceptions include alopecia, neuropathy, appropriately controlled endocrine toxicities, and other toxicities similar to these at the discretion of the Sponsor) is more than 28 days (1 cycle), a subject’s continuation in the study, based on the benefits and risks thereof, will be discussed with the Sponsor.

Subjects who have had an interruption of study treatment due to intercurrent illness and/or toxicity and/or necessary medical intervention may restart in the event that the interruption has been less than approximately 28 days and in the opinion of the Investigator, the subject has returned to baseline health, with approval from the Sponsor (see Section 9.7 for further details).

If a subject experiences ≥2 occurrences of the same treatment-emergent AE (TEAE) for which attribution to FHD-286 or (Part III only) the combination of FHD-286 with LDAC or decitabine cannot be excluded (for example, because the TEAE is believed to be due to underlying disease), the Investigator should consult with the Sponsor before resuming or discontinuing study treatment.

Dose modifications may also be required with certain concomitant medications; see Section 9.12.2.

For subjects receiving FHD-286 in combination with LDAC or decitabine in Part III, the Investigator should consult with the Sponsor to determine how to proceed in the event of dose delays with LDAC or decitabine.

#### 9.7. Toxicity Management Guidelines

Guidance on management of toxicities, including dose modifications of FHD-286 and discontinuation of FHD-286 when warranted, is provided in the following sections. For guidance on dose modifications and discontinuation of LDAC/decitabine, refer to the approved label (eg, USPI, SmPC) and institutional guidelines. No dose reductions are allowed for LDAC/decitabine. Exceptions may be allowed with approval from the Sponsor.

##### 9.7.1. Myelosuppression With or Without Fever

Monitor complete blood counts at the time of study enrollment and throughout the treatment period. Supportive measures such as antimicrobials administered for prophylaxis or in the setting of prolonged myelosuppression, and/or at the first signs of infection, are recommended to reduce the risk of a serious or severe infection that may lead to a fatal outcome in the setting of neutropenia with or without fever. Treatment should be individualized based on the level of myelosuppression at the time of study enrollment and throughout the course of study treatment, with consideration for the subject’s medical history, inclusive of prior infections.

Myeloid growth factors may be used in the setting of severe or prolonged neutropenia. The use of blood products, including packed red blood cells (pRBCs), and platelet transfusions are permitted and are to be administered at the Investigator’s discretion. Recommended guidelines for transfusion support include a platelet threshold of $10\times10^9$/L for platelet transfusion and a hemoglobin threshold of 8.0 g/dL for pRBC transfusion, or as clinically indicated at the Investigator’s discretion.

Use of these supportive measures is allowed during the DLT period and throughout study enrollment.

##### 9.7.2. Absolute Neutrophil and/or Platelet Count Decreases: FHD-286 Dose Modification Guidelines

The toxicity management guidelines in Table 4 should be followed for any event that has not been clearly and solely attributed to the underlying disease.

**Table 4: Absolute Neutrophil and/or Platelet Count Decreases: Dose Modification Guidelines**

| Toxicity | Dose Modification Guidelines | | | Additional Guidance |
|---|---|---|---|---|
| | **FHD-286** | **LDAC** | **Decitabine** | |
| Treatment-emergent decrease in ANC to <0.1×$10^9$/L and/or treatment-emergent decrease in platelet count to <5×$10^9$/L in the absence of evidence of disease-related myelosuppression | Hold FHD-286 until ANC recovers to >0.5×$10^9$/L and/or platelet count recovers to >20×$10^9$/L.<br>Resumption of FHD-286 before recovery to these levels should be discussed with the Sponsor. | Follow dose modification guidelines in the approved label (eg, USPI, SmPC). | Follow dose modification guidelines in the approved label (eg, USPI, SmPC). | Treat promptly as clinically indicated. |
| Any treatment-emergent decrease in ANC and/or platelet count associated with clinically significant fever, infection, and/or bleeding | Hold FHD-286 until clinically significant fever, infection, and/or bleeding is resolving and ANC recovers to >0.5×$10^9$/L and/or platelet count recovers to >20×$10^9$/L.<br>Resumption of FHD-286 before recovery to these levels should be discussed with the Sponsor. | Follow dose modification guidelines in the approved label (eg, USPI, SmPC). | Follow dose modification guidelines in the approved label (eg, USPI, SmPC). | Treat promptly as clinically indicated. |

Abbreviations: ANC=absolute neutrophil count; LDAC=low-dose cytarabine; SmPC=Summary of Product Characteristics; USPI=United States prescribing information.

##### 9.7.3. New-Onset Grade 3 or 4 Non-Hematologic Toxicities: FHD-286 Dose Modification Guidelines

The toxicity management guidelines in Table 5 should be followed for any event that has not been clearly and solely attributed to the underlying disease. A second occurrence is defined as an event that is medically equivalent to the first occurrence event.

**Table 5: New-Onset Grade 3 or 4 Non-Hematologic Toxicities: Dose Modification Guidelines**

| Toxicity | Occurrence | Dose Modification Guidelines | | | Additional Guidance |
|---|---|---|---|---|---|
| | | **FHD-286** | **LDAC** | **Decitabine** | |
| New Grade 3 or 4 non-hematologic toxicity that does not improve with maximal medical intervention within 7 days | First | Hold all study drugs until toxicity returns to baseline or Grade $\leq 2$.<br>If toxicity resolves to Grade $\leq 2$ within 7 **days** from onset date, resume FHD-286 at same dose level.<br>Consider reducing FHD-286 by 1 dose level from starting dose for all future treatment cycles if toxicity does not resolve to Grade $\leq 2$ within **7 days** from onset date despite maximal medical intervention.<br>Consult with Medical Monitor. | Follow dose modification guidelines in the approved label (eg, USPI, SmPC). | Follow dose modification guidelines in the approved label (eg, USPI, SmPC). | Treat promptly as clinically indicated. |
| | Second or later | Hold all study drugs until toxicity returns to baseline or Grade $\leq 2$.<br>Consult with Medical Monitor before resuming, reducing, or discontinuing study drugs. Medical Monitor will guide dose modification actions to be taken with study drugs. | Follow dose modification guidelines in the approved label (eg, USPI, SmPC). | Follow dose modification guidelines in the approved label (eg, USPI, SmPC). | Treat promptly as clinically indicated. |

Abbreviations: LDAC=low-dose cytarabine; SmPC=Summary of Product Characteristics; USPI=United States prescribing information.

##### 9.7.4. Differentiation Syndrome: Guidelines for Monitoring and Management

Differentiation syndrome is an AESI with FHD-286.

Differentiation syndrome is caused by the rapid release of a high level of cytokines from leukemia cells and may occur with or without concomitant leukocytosis. Signs and symptoms of differentiation syndrome are also frequently observed with other relatively common complications of advanced hematologic malignancies. Because the onset of differentiation syndrome can be rapid and because the syndrome can be severe, treatment for differentiation syndrome must be initiated at the earliest suspicion of the syndrome. Guidelines for monitoring and management of differentiation syndrome in subjects treated with FHD-286 are presented in Figure 3. Note that if FHD-286 treatment is interrupted because of differentiation syndrome, consultation with the Sponsor is required before reinitiating treatment with FHD-286 to determine the appropriate dose.

Clinical and/or laboratory signs and symptoms potentially associated with differentiation syndrome may manifest at any time during treatment with FHD-286, and may recur with premature discontinuation of treatment (eg, corticosteroids, hydroxyurea).

If differentiation syndrome is suspected, Investigators must report the event as an important medical event.

**Figure 3: Differentiation Syndrome Monitoring and Management**

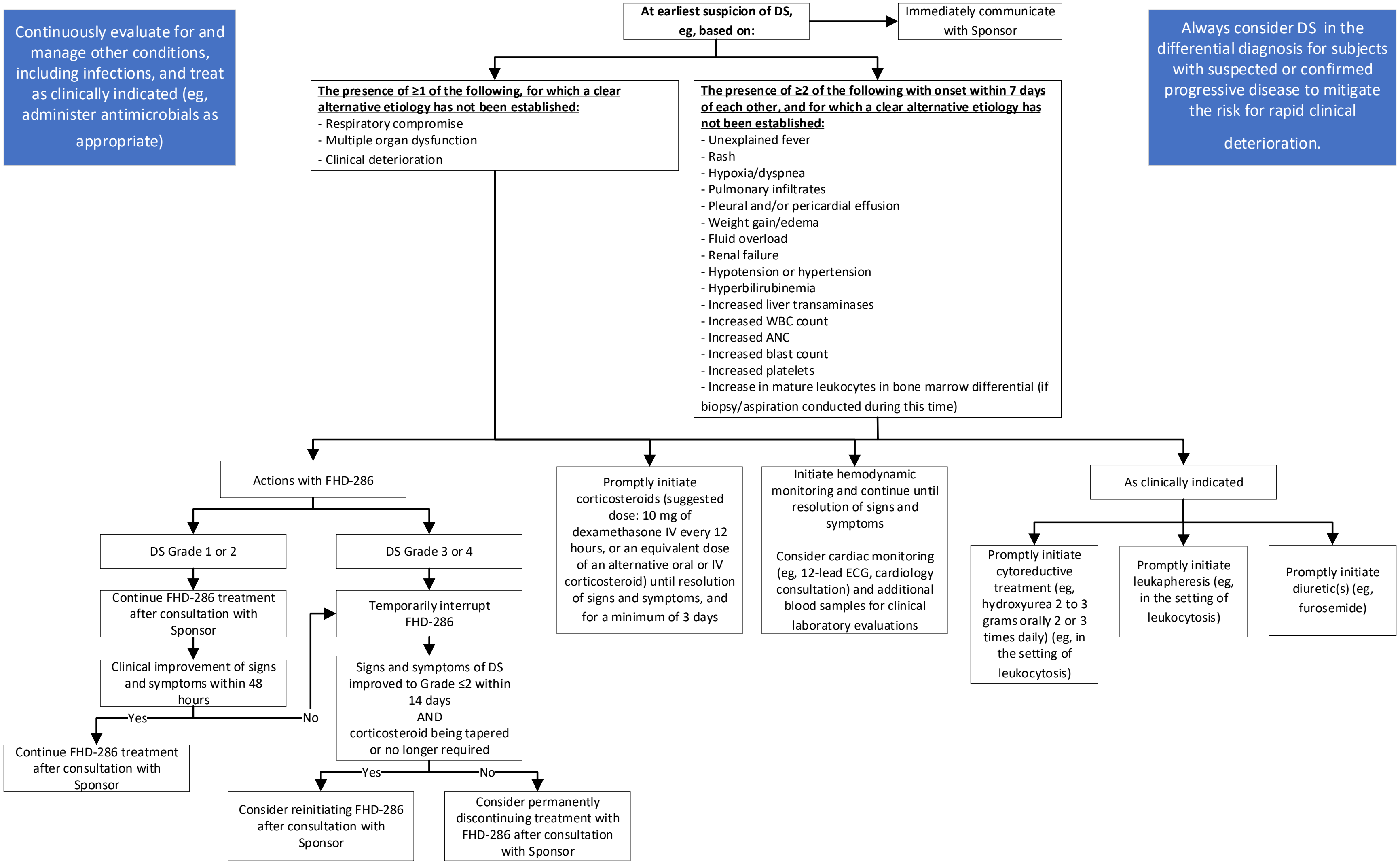

Abbreviations: ANC = absolute neutrophil count; DS = differentiation syndrome; ECG = electrocardiogram; IV = intravenous(ly); WBC = white blood cell.
Note: Additional guidance on potential signs and symptoms of differentiation syndrome is located in Appendix 15.8.

##### 9.7.5. Noninfectious Leukocytosis: Guidelines for Management

In Part III, subjects must have a WBC count $\leq 20\times10^9$/L to begin study treatment; treatment with a stable dose of hydroxyurea (or other cytoreductive agent, eg, cytarabine) to achieve and maintain this count is allowed.

Once treatment with FHD-286 has begun, it is recommended that cytoreductive treatment be undertaken at the earliest manifestations of leukocytosis. Leukapheresis should be initiated promptly if clinically indicated. Cytoreductive treatment should be tapered once noninfectious leukocytosis improves or resolves, as clinically indicated.

If the subject's WBC count is not decreasing within 24 to 48 hours with maximal medical intervention:

- Temporarily interrupt treatment with FHD-286
- Notify the Sponsor

Once the subject's WBC count improves to $\leq 20\times10^9$/L, FHD-286 may be reinitiated. The dose at which FHD-286 should be reinitiated should be discussed with the Sponsor. The subject should be monitored for rebound noninfectious leukocytosis after FHD-286 is reinitiated.

##### 9.7.6. QT Interval Prolongation: Management Guidelines

Dose modification criteria and monitoring guidelines for QTc prolongation are provided in Table 6.

**Table 6: QTc Prolongation: Dose Modification Criteria and Monitoring Guidelines**

| **Electrocardiogram QT corrected interval prolonged** | | | |
|---|---|---|---|
| **NCI CTCAE Grade:** | | | |
| **1**<br>Average QTc 450-480 ms | **2**<br>Average QTc 481-500 ms<br>For subjects with a baseline average QTc of 450-470 ms, follow Grade 2 management if QTc from baseline >30 ms | **3**<br>Average QTc ≥501 ms; >60 ms change from baseline | **4**<br>Torsades de pointes; polymorphic ventricular tachycardia; signs/symptoms of serious arrhythmia |
| **Management:** | | | |
| None required | Continue current dose or consider modifying dose level[a] of FHD-286.<br>Increase ECG monitoring to once weekly.<br>Obtain blood sample to assess electrolyte status (potassium, calcium, magnesium).<br>Administer appropriate electrolyte supplementation for correction of any values outside the normal range.<br>Review and adjust concomitant medications with known or suspected QTc-prolonging effects (Appendix 15.9).<br>Obtain blood sample to assess FHD-286 PK.<br>If current dose of FHD-286 is continued, collect ECGs once weekly until at least 2 consecutive results demonstrate stable QTc Grade ≤1 or baseline.<br>If dose of FHD-286 is modified, collect ECGs once weekly until at least 2 consecutive results demonstrate QTc Grade ≤1 or baseline.<br>Subject may return to the prior dose once at least 2 consecutive results demonstrate QTc Grade ≤1 or baseline. Once back at prior dose, once-weekly ECGs should be continued until 2 consecutive results demonstrate QTc Grade ≤1 or baseline. | Interrupt treatment with FHD-286.<br>Evaluate for underlying cause(s); evaluation may include cardiology consultation. Hospitalization may be required to medically manage the QTc prolongation, based on Investigator's assessment of appropriate level of medical care necessary to ensure subject's safety.<br>Monitor and supplement electrolyte levels (ie, potassium, magnesium, calcium) as clinically indicated.<br>Review and adjust concomitant medications with known or suspected QTc-prolonging effects (Appendix 15.9).<br>When QTc returns to the subject's baseline, subject may resume treatment with FHD-286, at the same or a reduced dose, after consultation with Sponsor.<br>Monitor ECGs at least weekly after return of QTc to subject's baseline, for a minimum of 2 weeks.<br>FHD-286 may be permanently discontinued in any subject who experiences a recurrence of Grade 3 toxicity of QTc prolongation after the restart of FHD-286 if underlying cause(s) cannot be medically managed. Consultation with Sponsor is required before resuming treatment with FHD-286 in any subject with more than 1 occurrence of a Grade 3 toxicity of QTc prolongation. | Stop dosing FHD-286.<br>Evaluate for underlying cause(s); evaluation may include cardiology consultation.<br>Admit to hospital for continuous cardiac monitoring and discharge only after review by a cardiologist.<br>Monitor and supplement electrolyte levels (ie, potassium, magnesium, calcium) as clinically indicated.<br>Review and adjust concomitant medications with known or suspected QTc-prolonging effects (Appendix 15.9). |

Abbreviations: ECG = electrocardiogram; ms = millisecond(s); NCI CTCAE = National Cancer Institute Common Terminology Criteria for Adverse Events; PK = pharmacokinetics; QTc = corrected QT interval; QTcF = QT interval corrected using Fridericia formula.

Note: This protocol uses the Fridericia formula (QTcF) for corrected QT interval values.

[a] Dose modification may include reduction of the individual dose and/or reduction of dosing frequency. All dose modifications should be discussed with the Sponsor.

#### 9.8. Stopping Criteria

The occurrence of either of the following will prompt temporary suspension of enrollment to the dose level at which the criterion was met:

- Any death during the DLT evaluation period that is related to study treatment
- Any Grade 3 or 4 AE of Differentiation syndrome that does not improve to Grade ≤2 within 14 days after maximal medical interventions

If a stopping criterion is met, enrollment to that dose level will be suspended until the CST has performed a prompt review of the cumulative safety data and of the circumstances of the event(s) in question, to determine whether dosing and/or the protocol should be modified. The independent safety monitoring committee may be consulted if the CST cannot reach consensus.

The Sponsor will determine whether a pause in enrollment to the study is warranted based on recommendations made by the CST and/or the independent safety monitoring committee.

#### 9.9. Duration of Subject Participation

Subjects may continue study treatment until 1 or more subject withdrawal criteria (Section 8.6) are met, or at the subject's discretion, or if the study is terminated.

After discontinuation of FHD-286, subjects are to attend a Safety Follow-up Visit 28 days after the EOT Visit. When FHD-286 is withheld from a subject to resolve a toxicity and the subject does not subsequently restart treatment, EOT is defined as the date on which the Investigator decides to discontinue FHD-286 and is not required to be equivalent to the date of the last FHD-286 dose. Subjects should proceed with EOT assessments, safety follow-up, and long-term follow-up. If the decision to not restart FHD-286 occurs outside of the 28-day safety follow-up window, the subject should proceed with EOT assessments and long-term follow-up.

After discontinuing FHD-286, subjects will be contacted by telephone approximately every 2 months for long-term follow-up. Long-term follow-up includes assessment of:

- Survival status
- Receipt and type of subsequent anticancer therapy
- Disease status, including date of most recent evaluation (unless subject discontinued FHD-286 due to progressive disease or treatment failure)
  - Collection of disease status will only continue until the subject experiences disease progression or treatment failure, or starts a new anticancer therapy.

Long-term follow-up will continue until all subjects have died, withdrawn consent/assent, or are lost to follow-up, or for 2 years after the last subject discontinues FHD-286, whichever occurs first.

##### 9.9.1. End of Study

End of study is defined as the time at which all subjects in all countries have discontinued study treatment and have been followed for survival and disease status assessment for at least 2 years, or have died, been lost to follow-up, or withdrawn consent/assent.

This study may be prematurely terminated, if in the opinion of the Sponsor, there is sufficiently reasonable cause. In the event of such action, written notification documenting the reason for study termination will be provided to each Investigator.

Circumstances that may warrant termination include, but are not limited to:

- Determination of unexpected, significant, or unacceptable risk to subjects.
- Failure to enroll subjects at an acceptable rate.
- Insufficient adherence to protocol requirements.
- Plans to modify, suspend, or discontinue the development of FHD-286.
- Other administrative reasons.

Should the study be closed prematurely, all study materials must be returned to the Sponsor or Sponsor's designee.

#### 9.10. Treatment Compliance

Subjects will be asked to return all unused study treatment on Day 1 of each cycle or at the next study visit.

Treatment compliance with FHD-286 and, if applicable, self-administered LDAC will be assessed based on return of unused drug and the dosing diary (see Section 9.5.4).

When subjects are dosed at the site, they will receive study treatment from the Investigator or designee, under medical supervision.

#### 9.11. Study Drug Accountability

Accountability for the study drug at the study site is the responsibility of the Investigator. The Investigator will ensure that the study drug is used only in accordance with this protocol. Where allowed, the Investigator may choose to assign drug accountability responsibilities to a pharmacist or other appropriate individual.

The Investigator or delegate will maintain accurate drug accountability records indicating the drug's delivery date to the site, inventory at the site, use by each subject, and return to Foghorn Therapeutics Inc. (hereafter Foghorn) or its designee (or disposal of the drug, if approved by Foghorn). These records will adequately document that the subjects were provided the doses as specified in the protocol and should reconcile all study drug received from Foghorn. Accountability records will include dates, quantities, batch/serial numbers, expiration dates (if applicable), and subject numbers. The Sponsor or its designee will review drug accountability on an ongoing basis during monitoring visits.

Study drug must not be used for any purpose other than the present study. Study drug which has been dispensed to a subject and returned unused must not be re-dispensed to a different subject.

Subjects will receive instructions for home administration of FHD-286 along with a diary to record the date and time of each dose, as well as the number and strength (mg) of capsules taken.

All unused and used study drug will be retained at the site until inventoried by the study monitor. All used, unused, or expired study drug will be returned to the Sponsor or its designee or if authorized, disposed of at the study site per the site's Standard Operating Procedures and must be documented. All material containing FHD-286 will be treated and disposed of as hazardous waste in accordance with governing regulations.

#### 9.12. Prior and Concomitant Medications and Treatments

##### 9.12.1. Prior Medications and Procedures

All medications administered and procedures conducted within 28 days prior to the first day of study treatment administration and through 28 days after the last dose of FHD-286 are to be recorded on the eCRF. In addition, all prior treatments for the underlying malignancy should be recorded.

##### 9.12.2. Prohibited Concomitant Therapy

Prohibited concomitant therapies include:

- Strong CYP3A inhibitors (Appendix 15.4), except triazole antifungal agents (Section 9.12.5)
- Strong CYP3A inducers (Appendix 15.4)
- Sensitive CYP3A substrates that have a narrow TI (Appendix 15.4)
- Sensitive P-gp and BCRP substrates that are orally administered and have a narrow TI (Appendix 15.5)
- Strong P-gp or BCRP inhibitors (Appendix 15.6)
- PPIs

If treatment with a strong CYP3A inhibitor or inducer, a sensitive CYP3A substrate with a narrow TI, an orally administered sensitive P-gp or BCRP substrate with a narrow TI, or a strong P-gp or BCRP inhibitor is medically necessary, discussion with the Sponsor is required before continuing FHD-286 dosing. Dose reduction, interruption, and/or discontinuation of FHD-286 may be necessary.

**Strong CYP3A4 inhibitors and inducers:** Strong CYP3A inhibitors and inducers may affect FHD-286 plasma exposure. If these are concomitantly administered, blood samples may be collected to determine FHD-286 concentrations in the plasma and to understand the PK behaviors of this compound. Available PK data will be reviewed by the CST and dose modifications may be made as needed. Monitoring of blood levels of concomitantly administered CYP3A4 inhibitors is recommended as per standard of care.

**PPIs:** When possible, administration of PPIs should be stopped or switched to another ARA (eg, antacids or H2 blockers; see Section 9.12.4) 7 days before administration of FHD-286. If it is medically necessary to dose PPIs concomitantly with FHD-286, this may be permitted with Sponsor approval. Proton pump inhibitors are acid-reducing agents and may affect FHD-286 absorption, decreasing its bioavailability. If these are concomitantly administered, blood samples may be collected to determine FHD-286 concentrations in the plasma and to understand the PK behaviors of this compound.

##### 9.12.3. Concomitant Therapy Requiring Careful Monitoring

Precautionary measures must be taken for:

- Moderate CYP3A inhibitors and inducers (Appendix 15.4)
- Substrates that are predominantly metabolized by CYP3A that do not have a narrow TI (Appendix 15.4; discuss with Sponsor before starting concomitant therapy)
- Substrates that are predominantly metabolized by CYP1A2, 2C8, and 2C9

**Moderate CYP3A inhibitors and inducers:** Moderate CYP3A inhibitors and inducers may affect FHD-286 plasma exposures. If these are concomitantly administered, blood samples may be collected to determine FHD-286 concentrations in the plasma and to understand the PK behaviors of this compound.

**Substrates of CYP3A, 1A2, 2C8, and 2C9:** Coadministration of FHD-286 and substrates that are predominantly metabolized by CYP3A that do not have a narrow TI, or by 1A2, 2C8, or 2C9, may increase the concentration of these drugs. Subjects should be monitored for either loss of therapeutic effects of these drugs or heightened safety profile. Dose modification of the concomitant medication may be required with certain drugs that are predominantly metabolized by CYP1A2, 2C8, or 2C9, or that are predominantly metabolized by CYP3A and do not have a narrow TI.

##### 9.12.4. Antacids and H2 Blockers

Administration of antacids and H2 blockers is permitted according to the following guidelines:

- Antacids: FHD-286 should be administered 2 hours before or 2 hours after administration of antacids.
- H2 blockers: FHD-286 should be administered at least 2 hours before or 10 to 12 hours after administration of H2 blockers.

Antacids and H2 blockers are acid-reducing agents and may affect FHD-286 absorption, decreasing its bioavailability. If these are concomitantly administered, blood samples may be collected to determine FHD-286 concentrations in the plasma and to understand the PK behaviors of this compound.

##### 9.12.5. Triazole Antifungal Agents

Triazole antifungal agents, including those classified as strong CYP3A4 inhibitors, are permitted with careful monitoring. Monitoring of blood levels for triazole antifungal agents classified as strong CYP3A4 inhibitors is recommended as per standard of care.

**Groups A1 and B1:** If a subject in Group A1 or B1 requires concomitant treatment with a triazole antifungal agent classified as a strong CYP3A4 inhibitor after their first dose of FHD-286:

- Discussion with the Sponsor is required before continuing FHD-286 dosing. Dose reduction, interruption, and/or discontinuation of FHD-286 may be necessary.
- In addition to the assessments of PK and safety outlined in the Schedules of Assessments (Section 10.1.2):

- It is recommended that clinical laboratory assessments (Table 14), ECGs, and a limited physical examination (Section 10.4.1) be performed at least weekly for the first 4 weeks of concomitant treatment with a triazole antifungal agent classified as a strong CYP3A4 inhibitor, and as clinically indicated thereafter.
- Additional serial FHD-286 PK and PD blood samples may be collected to determine FHD-286 concentrations and effects on PD biomarkers.

**Groups A2 and B2:** Assessments of PK and safety are outlined in the Schedules of Assessments (Section 10.1.2).

###### 9.12.6. Allowed Concomitant Therapy

Medications and treatments other than those specified above are permitted during the study.

All intercurrent medical conditions and complications of the underlying malignancy will be treated at the discretion of the Investigator according to acceptable local standards of medical care. Subjects should receive analgesics, antiemetics (Investigator-prescribed), antidiarrheals, anti-infectives, antipyretics, and blood products as necessary. All concomitant medications, including transfusions of blood products, will be recorded on the eCRF.

Palliative radiation is permitted at the discretion of the Sponsor.

### 10. STUDY ASSESSMENTS

#### 10.1. Schedules of Events

##### 10.1.1. Parts I and II: FHD-286 Monotherapy *(Closed to Enrollment)*

The Schedule of Assessments for subjects receiving study drug daily in this study is provided in Table 7. The timing of PK, PD, exploratory/translational tissue sample collection, and ECGs for subjects receiving study drug daily is provided in Table 8.

Schedules of Assessments and timing of PK, PD, exploratory/translational tissue sample collection, and ECGs for subjects receiving study drug on alternative dosing regimens (eg, intermittent dosing) are provided in Appendix 15.2.

In the event that the volume of blood to be collected from any given subject will total >7 mL/kg in any 8-week period and/or there is clinical concern regarding the amount of blood to be collected in a given period for a specific subject, please contact the Medical Monitor.

**Table 7: Parts I and II *(Closed to Enrollment)*: FHD-286 Monotherapy Schedule of Assessments**

| Visit/Cycle | Screening[a] | Cycle 1, 2, & 3 | | | | Cycle 4+ | | EOT Visit[b] | Safety Follow-up | Long-Term Follow-up[c] |
|---|---|---|---|---|---|---|---|---|---|---|
| Study Day | | 1[d] | 3 or 4 (Cycle 1 only) | 8 | 15 | 1 | 15[e] | | +28[f] | |
| Window (days) | | NA (Cycle 1) or ±2 (Cycles 2 & 3) | NA | ±2 | | | | +5 | ±7 | ±14 |
| Informed consent/assent | X | | | | | | | | | |
| Study Eligibility | X | | | | | | | | | |
| Demographics | X | | | | | | | | | |
| Medical & surgical history | X | | | | | | | | | |
| Physical examination[g] | X | X | | X | X | X | X | X | X | |
| Vital signs | X | X | | X | X | X | X | X | X | |
| ECOG PS | X | X | | X | X | X | X | X | X | |
| Clinical laboratory tests (local)[h] | See Table 12. | | | | | | | | | |
| ECHO (or other means) for LVEF | X | X (either C3D1 or C4D1 [±3 days]) | | | | | | X | | |
| Electrocardiogram[i] | See Table 12. | | | | | | | | | |
| PK/PD assessments[j] | See Table 12. | | | | | | | | | |
| Bone marrow biopsy and/or aspirate[k] | X | Starting on C2D1, bone marrow biopsy and/or aspirate will be collected approximately every 4 weeks for the first 24 weeks, then approximately every 8 weeks for the next 48 weeks of treatment, and as clinically indicated thereafter | | | | | | X | | |
| Evaluate extent of disease and response to treatment[l] | | Evaluation will occur in accordance with bone marrow biopsy and/or aspirate collection | | | | | | X | | |

**Table 7: Parts I and II *(Closed to Enrollment)*: FHD-286 Monotherapy Schedule of Assessments**

| Visit/Cycle | Screening[a] | Cycle 1, 2, & 3 | | | | Cycle 4+ | | EOT Visit[b] | Safety Follow-up | Long-Term Follow-up[c] |
|---|---|---|---|---|---|---|---|---|---|---|
| Study Day | | 1[d] | 3 or 4 (Cycle 1 only) | 8 | 15 | 1 | 15[e] | | +28[f] | |
| Window (days) | | NA (Cycle 1) or ±2 (Cycles 2 & 3) | NA | ±2 | | | | +5 | ±7 | ±14 |
| Hepatitis panel (A, B, C) | X | | | | | | | | | |
| Serum HIV | X | | | | | | | | | |
| Pregnancy testing[m] | X | X | | | | X | | | X | |
| Dispense study medication[n] | | X | | X | X | X | X | | | |
| Concomitant medications/ procedures | X | X | | X | X | X | X | X | X | |
| AE assessment | X | X | | X | X | X | X | X | X | |
| Survival status | | | | | | | | | X | X |

Abbreviations: AE = adverse event; C1D1 = Cycle 1, Day 1; C4D15 = Cycle 4, Day 15; ECG = electrocardiogram; ECHO = echocardiogram; ECOG = Eastern Cooperative Oncology Group; EOT = End of Treatment; hCG = human chorionic gonadotropin; HIV = human immunodeficiency virus; IWG = International Working Group; NA = not applicable; PD = pharmacodynamics; PK = pharmacokinetics; PS = Performance Status.

[a] Within 28 days prior to first dose of FHD-286 (C1D1).

[b] EOT Visit will take place within 5 days after treatment ends (treatment end is defined as the date on which the Investigator decided to discontinue FHD-286 treatment and is not required to be the date of the last FHD-286 dose). If a subject's dose is interrupted for 28 days and then the subject discontinues study participation, the EOT Visit will serve as the Safety Follow-up Visit.

[c] After subjects have either experienced documented disease progression/treatment failure or discontinued study treatment, whichever occurs later, they will be contacted by telephone approximately every 2 months to assess survival status, document receipt and type of subsequent anticancer therapy, and document disease status (until subject experiences disease progression/treatment failure or starts a new anticancer therapy).

[d] Assessments on C1D1 will be performed predose, except for PK/PD samples and ECGs which will be collected according to Table 12.

[e] At C4D15 and beyond, samples for laboratory assessments will be collected on site; other assessments may be made via telemedicine.

[f] Safety Follow-up Visit will occur approximately 28 days after the EOT Visit.

[g] A complete physical examination, including height and weight, will be performed during Screening. Physical examinations may be abbreviated (ie, partial) at all other protocol-specified visits. Weight should be collected on Day 1 of each cycle.

[h] Clinical laboratory tests include hematology, serum chemistry, and coagulation studies. Samples for analysis will be collected as specified in Table 12. Specific analytes are outlined in Table 14.

[I] 12-lead ECGs will be collected in triplicate prior to each PK time point as described in Table 12 (starting after approximately 3 minutes of recumbency or semi-recumbency, with each ECG performed approximately 2 minutes apart). There is no associated PK sample collection for the Screening ECG; as such, the 12-lead Screening ECG may be performed in triplicate at any time during the Screening window.
[j] PK/PD and translational/exploratory assessments will be collected as specified in Table 12. Drug should be administered on site on days that PK/PD assessments will take place.
[k] A bone marrow biopsy and/or aspirate should be conducted if progression of disease is suspected. If additional bone marrow biopsies and/or aspirates are performed during the study at the Investigator's discretion, study samples will be collected.
[l] Disease response will be based upon modified IWG criteria as detailed in Section 10.6.
[m] Female subjects of reproductive potential only. Serum hCG testing is to be conducted at the Screening Visit. Serum or urine hCG testing will be conducted predose on Day 1 of every cycle.
[n] Subjects will be dispensed the appropriate number of Sponsor-packaged, labeled bottles to allow for dosing for a full cycle or until the next scheduled visit.

Note: All doses of FHD-286 will be administered under fasted conditions as described in Section 9.5.1.1.

**Table 8: Parts I and II *(Closed to Enrollment)*: Timing of Safety, Pharmacokinetic, Pharmacodynamic, and Translational/Exploratory Sampling and Electrocardiograms**

| Cycle | Study Day | Hour[a] | Safety Blood | PK Blood | PD Blood | Exploratory Blood | ECG[b] |
|---|---|---|---|---|---|---|---|
| Screening | -28[c] | | X | | | X | X |
| 1 | 1 | 0 | X | X | X | | X |
| | | 0.5 | | X | | | |
| | | 1 | | X | | | |
| | | 2 | | X | | | X |
| | | 4 | | X | X | | X |
| | | 6 | | X | | | X |
| | | 8 | | X | X | | X |
| | 2 | 0 | | X | X | | X |
| | 3 or 4 | | X[d] | | | | |
| | 8 | 0 | X | X | X | | |
| | | 2 | | X | | | |
| | 15 | 0 | X | X | X | X | X |
| | | 0.5 | | X | | | |
| | | 1 | | X | | | |
| | | 2 | | X | | | X |
| | | 4 | | X | X | | X |
| | | 6 | | X | | | X |
| | | 8 | | X | X | | X |
| | 16 | 0 | | X | X | | X |
| 2 | 1 | 0 | X | X | X | | X |
| | | 2 | | X | | | X |
| | 8 | 0 | X | X | | | |
| | 15 | 0 | X | X | | | |
| 3 | 1 | 0 | X | X | X | X | X |
| | | 4 | | X | X | | X |
| | | 8 | | X | X | | |
| | 8 | 0 | X | X | | | |
| | 15 | 0 | X | X | | | |
| 4+ | 1 | 0 | X | X | X[e] | X[e] | |

**Table 8: Parts I and II *(Closed to Enrollment)*: Timing of Safety, Pharmacokinetic, Pharmacodynamic, and Translational/Exploratory Sampling and Electrocardiograms**

| Cycle | Study Day | Hour[a] | Safety Blood | PK Blood | PD Blood | Exploratory Blood | ECG[b] |
|---|---|---|---|---|---|---|---|
| EOT[f] | | | X | X | X | X | |

Abbreviations: ECG = electrocardiogram; EOT = End of Treatment; PD = pharmacodynamics; PK = pharmacokinetics.
[a] Hour 0 samples to be collected within 30 minutes before dose. Samples at all other hours to be collected within ±10 minutes of specified time. ECGs should be collected within approximately 10 minutes prior to PK sample collection.
[b] 12-lead ECGs will be collected in triplicate prior to PK sample collection (starting after approximately 3 minutes of recumbency or semi-recumbency; each ECG performed approximately 2 minutes apart). There is no associated PK sample collection for the Screening ECG; as such, the 12-lead Screening ECG may be performed in triplicate at any time during the Screening window.
[c] Within 28 days prior to first dose of FHD-286 (C1D1).
[d] In Cycle 1 only, serum chemistry will be collected on either Day 3 or Day 4.
[e] After Cycle 3, PD and translational/exploratory blood will be collected on D1 of every odd-numbered cycle (Cycle 5, 7, etc.) for the first 48 weeks.
[f] EOT Visit will take place within 5 days after treatment ends (treatment end is defined as the date on which the Investigator decided to discontinue FHD-286 treatment and is not required to be equal to the date of the last FHD-286 dose).

Notes:
1. Drug should be administered on site on days that PK and/or PD assessments will take place.
2. Samples for PK may also be used for other exploratory analysis, including but not limited to metabolite profiling and additional biomarker analysis.
3. Subjects may be requested to undergo unscheduled PK/PD/safety assessments and ECGs if a safety event occurs.

##### 10.1.2. Part III: Combination Therapy

The Schedules of Assessments for subjects receiving combination therapy with FHD-286 QD are provided in:

- Arm A (FHD-286 + LDAC): Table 9
- Arm B (FHD-286 + decitabine): Table 10

The treatment schedules for Arms A and B are provided in Table 11.

The timing of clinical laboratory testing, PK, PD, exploratory tissue sample collection, and ECGs for subjects receiving combination therapy with FHD-286 at a consistent dose is provided in Table 12. The timing of clinical laboratory testing, PK, PD, exploratory tissue sample collection, and ECGs for subjects receiving combination therapy with FHD-286 interval dosing is provided in Table 13.

If the volume of blood to be collected from any given subject will total >7 mL/kg in any 8-week period and/or there is clinical concern regarding the amount of blood to be collected in a given period for a specific subject, contact the Sponsor.

**Table 9: Part III Arm A: FHD-286 QD + LDAC Schedule of Assessments**

| Visit/Cycle: | SCRN[a] | 1 | | | | | | | | 2 and 3 | | | 4+ | | EOT Visit[b,c] | Safety Follow-up Visit[c,d] | Long-Term Follow-up[e] |
|---|---|---|---|---|---|---|---|---|---|---|---|---|---|---|---|---|---|
| **Study Day:** | **−28** | **1[f]** | **2** | **3** | **4** | **5** | **8** | **15** | **22** | **1** | **8** | **15** | **1** | **15[g]** | **+5** | **+28** | **N/A** |
| **Visit Window (days):** | **–** | **N/A** | **N/A** | **N/A** | **N/A** | **N/A** | **±2** | **±2** | **±2** | **±2** | **±2** | **±2** | **±2** | **±2** | **N/A** | **±7** | **±14** |
| Informed consent/assent | X | | | | | | | | | | | | | | | | |
| Study eligibility | X | | | | | | | | | | | | | | | | |
| Demographics | X | | | | | | | | | | | | | | | | |
| Medical & surgical history | X | | | | | | | | | | | | | | | | |
| Physical examination[h] | X | X | | | | | | | | X | | | X | | X | X | |
| Vital signs | X | X | X | X | X | X | X | X | X | X | X | X | X | X | X | X | |
| ECOG PS | X | X | | | X | | X | X | X | X | | | X | | X | | |
| Clinical laboratory tests (local) | Refer to Table 12. | | | | | | | | | | | | | | | | |
| ECHO (or other means) for LVEF | X | | | | | | | | | C3D1 or C4D1 ±3 days | | | | | X | | |
| Electrocardiogram | Refer to Table 12. | | | | | | | | | | | | | | | | |
| PK/PD assessments | Refer to Table 12. | | | | | | | | | | | | | | | | |
| Bone marrow biopsy and/or aspirate | X | | | | | | | X | | X | | | X[i] | | X | X[j] | |
| Evaluate extent of disease and response to treatment[k] | X | | | | | | | X | | X | | | X | | X | X[j] | |
| Hepatitis panel (A, B, C) | X | | | | | | | | | | | | | | | | |
| Serum HIV | X | | | | | | | | | | | | | | | | |
| Pregnancy testing[l] | X | X | | | | | | | | X | | | X | | | X | |
| FHD-286 dosing | | Refer to Table 11. | | | | | | | | | | | | | | | |

##### Table 9: Part III Arm A: FHD-286 QD + LDAC Schedule of Assessments

| **Visit/Cycle:** | **SCRN[a]** | **1** | | | | | | | | **2 and 3** | | | **4+** | | **EOT Visit[b,c]** | **Safety Follow-up Visit[c,d]** | **Long-Term Follow-up[e]** |
|---|---|---|---|---|---|---|---|---|---|---|---|---|---|---|---|---|---|
| **Study Day:** | **−28** | **1[f]** | **2** | **3** | **4** | **5** | **8** | **15** | **22** | **1** | **8** | **15** | **1** | **15[g]** | **+5** | **+28** | **N/A** |
| **Visit Window (days):** | **–** | **N/A** | **N/A** | **N/A** | **N/A** | **N/A** | **±2** | **±2** | **±2** | **±2** | **±2** | **±2** | **±2** | **±2** | **N/A** | **±7** | **±14** |
| LDAC dosing | | Refer to Table 11. | | | | | | | | | | | | | | | |
| Concomitant medications/procedures | X | <==========================================================> | | | | | | | | | | | | | X | X | |
| AE assessment | X | <==========================================================> | | | | | | | | | | | | | X | X | |
| Survival status | | | | | | | | | | | | | | | | X | X |

Abbreviations: AE = adverse event; LDAC = low-dose cytarabine; ECG = electrocardiogram; ECHO = echocardiogram; ECOG PS = Eastern Cooperative Oncology Group Performance Status; EOT = End of Treatment; hCG = human chorionic gonadotropin; HIV = human immunodeficiency virus; IWG = International Working Group; LVEF = left ventricular ejection fraction; N/A = not applicable; PD = pharmacodynamic(s); PK = pharmacokinetic(s); QD = once daily; SCRN = Screening.

Note: If LDAC is discontinued and treatment with FHD-286 monotherapy is continued, any protocol visits intended solely for the administration of LDAC may be omitted.

[a] Within 28 days before the first dose of study treatment (ie, Cycle 1 Day 1).

[b] EOT Visit will occur within 5 days after treatment end, which is defined as the date on which the Investigator decided to discontinue FHD-286 treatment. The treatment end date is not required to be the date of the last FHD-286 dose.

[c] If a subject's FHD-286 treatment is interrupted for 28 days and the subject then discontinues study participation, the EOT Visit will serve as the Safety Follow-up Visit.

[d] The Safety-Follow-up Visit will occur 28 days ±7 days after the EOT Visit.

[e] After a subject has discontinued FHD-286, unless consent/assent to participate is withdrawn, they will be contacted by telephone approximately every 2 months to document survival status, receipt and type of subsequent anticancer therapy, and disease status (unless subject discontinued FHD-286 due to disease progression/treatment failure, and until subject experiences disease progression/treatment failure or starts a new anticancer therapy).

[f] Cycle 1 Day 1 assessments will be performed predose, unless specified otherwise.

[g] Starting in Cycle 4, the Day 15 assessments may be performed at a clinic local to the subject. Day 1 assessments must be performed at the study site.

[h] During Screening and at the EOT Visit, a complete physical examination, including assessment of height and weight, should be performed. At all other protocol-specified visits, physical examinations may be abbreviated (limited physical examination). Weight should be collected, and body surface area calculated, on Day 1 of each cycle.

[i] Independent of study treatment delays and/or interruptions, bone marrow biopsy and/or aspirate will be collected on Day 1 of Cycles 4, 5, and 6; on Day 1 of Cycles 8, 10, and 12; then on Day 1 of every third cycle (Cycles 15, 18, 21, etc); at the End of Treatment visit; and as clinically indicated (study samples will be obtained from these collections).

[j] Only for subjects who discontinue FHD-286 for reasons other than disease progression or treatment failure and continue to be treated with LDAC monotherapy outside the context of this study: bone marrow biopsy and/or aspirate should be collected at the Safety Follow-up Visit to support assessment of disease response.

[k] Disease response will be assessed at the same time points as collection of bone marrow biopsy and/or aspirate, using the modified IWG criteria (Section 10.6) or other criteria appropriate for the subject's advanced hematologic malignancy.

[l] Female subjects of reproductive potential only. At Screening, pregnancy testing is to be done via serum hCG assessment. Thereafter, pregnancy testing may be done via either serum or urine hCG assessment. Pregnancy testing is to be performed predose.

**Table 10: Part III Arm B: FHD-286 QD + Decitabine Schedule of Assessments**

| **Visit/Cycle:** | **SCRN[a]** | **1** | | | | | **2 and 3** | | | | **4+** | | | **EOT Visit[b,c]** | **Safety Follow-up Visit[c,d]** | **Long-term Follow-up[e]** |
|---|---|---|---|---|---|---|---|---|---|---|---|---|---|---|---|---|
| **Study day:** | **−28** | **1[f]** | **2 to 5** | **8** | **15** | **22** | **1** | **2 to 5** | **8** | **15** | **1** | **2 to 5** | **15[g]** | **+5** | **+28** | **N/A** |
| **Visit window (days):** | **N/A** | **N/A** | **N/A** | **±2** | **±2** | **±2** | **±2** | **±2** | **±2** | **±2** | **±2** | **±2** | **±2** | **N/A** | **±7** | **±14** |
| Informed consent/assent | X | | | | | | | | | | | | | | | |
| Study eligibility | X | | | | | | | | | | | | | | | |
| Demographics | X | | | | | | | | | | | | | | | |
| Medical & surgical history | X | | | | | | | | | | | | | | | |
| Physical examination[h] | X | X | | | | | X | | | | X | | | X | X | |
| Vital signs | X | X | X | X | X | X | X | X | X | X | X | X | X | X | X | |
| ECOG PS | X | X | D4 only | X | X | X | X | | | | X | | | X | | |
| Clinical laboratory tests | Refer to Table 12. | | | | | | | | | | | | | | | |
| ECHO (or other means) for LVEF | X | | | | | | C3D1 or C4D1 ±3 days | | | | | | | X | | |
| Electrocardiograms | Refer to Table 12. | | | | | | | | | | | | | | | |
| PK/PD assessments | Refer to Table 12. | | | | | | | | | | | | | | | |
| Bone marrow biopsy and/or aspirate | X | | | | X | | X | | | | X[i] | | | X | X[j] | |
| Evaluate extent of disease and response to treatment[k] | X | | | | X | | X | | | | X | | | X | X[j] | |
| Hepatitis panel (A, B, C) | X | | | | | | | | | | | | | | | |
| Serum HIV | X | | | | | | | | | | | | | | | |
| Pregnancy testing[l] | X | X | | | | | X | | | | X | | | | X | |
| FHD-286 dosing | | Refer to Table 11. | | | | | | | | | | | | | | |

**Table 10: Part III Arm B: FHD-286 QD + Decitabine Schedule of Assessments**

| **Visit/Cycle:** | **SCRN**[a] | **1** | | | | | **2 and 3** | | | | **4+** | | | **EOT Visit**[b,c] | **Safety Follow-up Visit**[c,d] | **Long-term Follow-up**[e] |
|---|---|---|---|---|---|---|---|---|---|---|---|---|---|---|---|---|
| **Study day:** | **−28** | **1**[f] | **2 to 5** | **8** | **15** | **22** | **1** | **2 to 5** | **8** | **15** | **1** | **2 to 5** | **15**[g] | **+5** | **+28** | **N/A** |
| **Visit window (days):** | **N/A** | **N/A** | **N/A** | **±2** | **±2** | **±2** | **±2** | **±2** | **±2** | **±2** | **±2** | **±2** | **±2** | **N/A** | **±7** | **±14** |
| Decitabine dosing | | Refer to Table 11. | | | | | | | | | | | | | | |
| Concomitant medications/procedures | X | <===================================================> | | | | | | | | | | | | X | X | |
| AE assessment | X | <===================================================> | | | | | | | | | | | | X | X | |
| Survival status | | | | | | | | | | | | | | | X | X |

Abbreviations: AE = adverse event; ECG = electrocardiogram; ECHO = echocardiogram; ECOG PS = Eastern Cooperative Oncology Group Performance Status; EOT = End of Treatment; hCG = human chorionic gonadotropin; HIV = human immunodeficiency virus; IWG = International Working Group; LVEF = left ventricular ejection fraction; N/A = not applicable; PD = pharmacodynamic(s); PK = pharmacokinetic(s); QD = once daily; SCRN = Screening.

Note: If decitabine is discontinued and treatment with FHD-286 monotherapy is continued, any protocol visits intended solely for the administration of decitabine may be omitted.

[a] Within 28 days before the first dose of study treatment (ie, Cycle 1 Day 1).

[b] EOT Visit will occur within 5 days after treatment end, which is defined as the date on which the Investigator decided to discontinue FHD-286 treatment. The treatment end date is not required to be the date of the last FHD-286 dose.

[c] If a subject's FHD-286 treatment is interrupted for 28 days and the subject then discontinues study participation, the EOT Visit will serve as the Safety Follow-up Visit.

[d] The Safety-Follow-up Visit will occur 28 days ±7 days after the last dose of FHD-286.

[e] After a subject has discontinued FHD-286, they will be contacted by telephone approximately every 2 months to assess survival status, document receipt and type of subsequent anticancer therapy, and document disease status (unless subject discontinued FHD-286 due to disease progression/treatment failure, and until subject experiences disease progression/treatment failure or starts a new anticancer therapy).

[f] Cycle 1 Day 1 assessments will be performed predose, unless specified otherwise.

[g] Starting in Cycle 4, the Day 15 assessments may be performed at a clinic local to the subject. Day 1 through 5 assessments must be performed at the study site.

[h] During Screening and at the EOT Visit, a complete physical examination, including assessment of height and weight, should be performed. At all other protocol-specified visits, physical examinations may be abbreviated (limited physical examination). Weight should be collected, and body surface area calculated, on Day 1 of each cycle.

[i] Independent of study treatment delays and/or interruptions, bone marrow biopsy and/or aspirate will be collected on Day 1 of Cycles 4, 5, and 6; on Day 1 of Cycles 8, 10, and 12; then on Day 1 of every third cycle (Cycles 15, 18, 21, etc); at the End of Treatment visit; and as clinically indicated (study samples will be obtained from these collections).

[j] Only for subjects who discontinue FHD-286 for reasons other than disease progression or treatment failure and continue to be treated with decitabine monotherapy outside the context of this study: bone marrow biopsy and/or aspirate should be collected at the Safety Follow-up Visit, to support assessment of disease response.

[k] Disease response will be assessed at the same time points as collection of bone marrow biopsy and/or aspirate, using the modified IWG criteria (Section 10.6) or other criteria appropriate for the subject's advanced hematologic malignancy.

[l] Female subjects of reproductive potential only. At Screening, pregnancy testing is to be done via serum hCG assessment. Thereafter, pregnancy testing may be done via either serum or urine hCG assessment. Pregnancy testing is to be performed predose.

**Table 11: Part III Arms A and B: Treatment Schedule**

| Cycle: | All Cycles | | | | | | | | | | |
|---|---|---|---|---|---|---|---|---|---|---|---|
| **Study day:** | **1** | **2** | **3** | **4** | **5** | **6** | **7** | **8** | **9** | **10** | **11-28** |
| **Dosing window:** | **Dosing windows are not allowed** | | | | | | | | | | |
| FHD-286[a,b,c] PO QD | X | X | X | X | X | X | X | X | X | X | X |
| LDAC[d] 20 mg/m$^2$ SC QD **(Arm A only)** | X | X | X | X | X | X | X | X | X | X | |
| Decitabine 20 mg/m$^2$ IV QD **(Arm B only)** | X | X | X | X | X | | | | | | |

Abbreviations: IV = intravenous; LDAC = low-dose cytarabine; PD = pharmacodynamic(s); PK = pharmacokinetic(s); PO = oral; QD = once daily; SC = subcutaneous.
Note: Study treatment should be administered on site on scheduled visit days, including days when PK/PD assessments take place (see Table 9 [LDAC], Table 10 [decitabine], Table 12, and Table 13 for time points; note that windows are not allowed for dosing days). FHD-286 and LDAC (see footnote d) may otherwise be administered at home. The appropriate amount of FHD-286 capsules in bottles will be dispensed to the subject to allow dosing for a full cycle or until the next scheduled visit. For subjects self-administering LDAC at home, prefilled syringes will be dispensed to subjects as per institutional practice.

[a] If the combination agent is discontinued for reasons other than treatment failure or progressive disease (eg, toxicity), subject may continue FHD-286 monotherapy until any of the withdrawal criteria in Section 8.6 are met.

[b] All doses of FHD-286 must be administered under fasted conditions (Section 9.5.1.1).

[c] On combination dosing days, FHD-286 must be administered before the combination agent.

[d] For Days 1 through 5 of Cycle 1, LDAC will be administered at the study site. After the first 5 doses (ie, beginning on Cycle 1 Day 6), at the Investigator’s discretion and where consistent with institutional, state, and local guidelines and regulations, subjects may self-administer LDAC at home, except on scheduled visit days.

**Table 12: Part III Arms A and B, Consistent FHD-286 Dose: Timing of Clinical Laboratory, Pharmacokinetic, Pharmacodynamic, and Exploratory Sampling and Electrocardiograms**

| Cycle | Study Day | Hr[a] | Clinical Laboratory Testing[b] | | PK Blood | PD Blood | Exploratory Blood | PK Urine | ECG[c] |
|---|---|---|---|---|---|---|---|---|---|
| | | | Hematology/ Chemistry/ Coagulation | Inflammatory Markers/ Cytokines | | | | | |
| SCRN | −28[d] | | X | X | | X | X | | X |
| | −3 | | X[e] | | | | | | |
| 1 | 1 | 0 | X | X | X | X | | | X |
| | | 0.5 | | | X | | | | |
| | | 1 | | | X | | | | |
| | | 2 | | | X | | | | X |
| | | 4 | | | X | | | | X |
| | | 6 | | | X | | | | X |
| | | 8 | | | X | X | | | X |
| | 2 | 0 | X | | X | X | | | X |
| | 3 | 0 | X | | | | | | |
| | 4 | 0 | X | X | | | | | |
| | 5 | 0 | X | | | | | | |
| | 8 | 0 | X | X | X | X | | | X |
| | | 2 | | | X | | | | |
| | 11 | 0 | X | X | | | | | |
| | 15 | 0 | X | X | X | X | X | X[f] | X |
| | | 0.5 | | | X | | | | |
| | | 1 | | | X | | | | |
| | | 2 | | | X | | | | X |
| | | 4 | | | X | | | | X |
| | | 6 | | | X | | | | X |
| | | 8 | | | X | | | | X |
| | 18 | 0 | X | X | | | | | |
| | 22 | 0 | X | X | | | | | X |
| | 25 | 0 | X | X | | | | | |

**Table 12: Part III Arms A and B, Consistent FHD-286 Dose: Timing of Clinical Laboratory, Pharmacokinetic, Pharmacodynamic, and Exploratory Sampling and Electrocardiograms**

| Cycle | Study Day | Hr[a] | Clinical Laboratory Testing[b] | | PK Blood | PD Blood | Exploratory Blood | PK Urine | ECG[c] |
|---|---|---|---|---|---|---|---|---|---|
| | | | Hematology/ Chemistry/ Coagulation | Inflammatory Markers/ Cytokines | | | | | |
| 2 | 1 | 0 | X | X | X | X | | | X |
| | | 2 | | | X | | | | X |
| | 8 | 0 | X | X | X | | | | |
| | 15 | 0 | X | X | X | | | | |
| 3 | 1 | 0 | X | X | X | X | X | | X |
| | | 4 | | | X | | | | X |
| | | 8 | | | X | | | | |
| | 8 | 0 | X | X | X | | | | |
| | 15 | 0 | X | X | X | | | | |
| 4 | 1 | 0 | X | X | X | | | | X |
| | 15 | 0 | X | | | | | | |
| 5+ | 1 | 0 | X | X | X | X[g] | X[g] | | X |
| | 15 | 0 | X | | | | | | |
| EOT[h] | | | X | X | X | X | X | | |
| Safety Follow-up Visit[i] | | | X | | | | | | |

Abbreviations: ECG = electrocardiogram; EOT = End of Treatment; PD = pharmacodynamics; PK = pharmacokinetics.

[a] Relative to the FHD-286 dose. Hour 0 samples for clinical laboratory testing to be collected within 12 hours before dose. All other Hour 0 samples to be collected within 2 hours before dose. Samples at all other hours to be collected within ±10 minutes of specified time. ECGs should be collected within approximately 10 minutes prior to PK sample collection.

[b] Clinical laboratory parameters are defined in Table 14. All samples will be analyzed locally. In Cycle 1, assessments on Days 4, 11, 18, and 25 may be performed within ±1 day of the specified time point. In Cycle 1, assessments on Days 11, 18, and 25 may be performed at a healthcare provider local to the subject.

[c] 12-lead ECGs will be collected in triplicate prior to PK sample collection (starting after approximately 3 minutes of recumbency or semi-recumbency; each ECG performed approximately 2 minutes apart). There is no associated PK sample collection for the Screening ECG; as such, the 12-lead Screening ECG may be performed in triplicate at any time during the Screening window.

[d] Within 28 days before the first dose of study treatment (Cycle 1 Day 1).

[e] A white blood cell count assessment must be completed within 72 hours before the subject's first scheduled dose of study treatment to confirm eligibility.

[f] Continuous urine collection over 24 hours, beginning at predose on Cycle 1 Day 15, will be performed for at least 3 subjects in Part III, who will be chosen in discussion with the Investigator(s) based on considerations such as the dose at which the subject is to be initiated and the individual subject's ability to participate in the urine collection assessment.

[g] After Cycle 3, PD and exploratory blood will be collected on Day 1 of every odd-numbered cycle (Cycle 5, 7, 9, and 11) for the first 48 weeks.

[h] EOT Visit will take place within 5 days after treatment ends (treatment end is defined as the date on which the Investigator decided to discontinue FHD-286 treatment and is not required to be equal to the date of the last FHD-286 dose).

[i] Safety Follow-up Visit will occur 28 days ±7 days after the EOT Visit.

Notes:

1. Study treatment should be administered on site on days that PK and/or PD assessments will take place.

2. Samples for PK may also be used for other exploratory analysis, including but not limited to metabolite profiling and additional biomarker analysis.
3. Subjects may be requested to undergo unscheduled PK/PD/safety assessments and ECGs if a safety event occurs.

**Table 13: Part III Arms A and B, Interval Dosing of FHD-286: Timing of Clinical Laboratory, Pharmacokinetic, Pharmacodynamic, and Exploratory Sampling and Electrocardiograms**

| Cycle | Study Day | Hr[a] | Clinical Laboratory Testing[b] | | PK Blood | PD Blood | Exploratory Blood | PK Urine | ECG[c] |
|---|---|---|---|---|---|---|---|---|---|
| | | | Hematology/ Chemistry/ Coagulation | Inflammatory Markers/ Cytokines | | | | | |
| SCRN | −28[d] | | X | X | | X | X | | X |
| | −3 | | X[e] | | | | | | |
| 1 | 1 | 0 | X | X | X | X | | | X |
| | | 0.5 | | | X | | | | |
| | | 1 | | | X | | | | |
| | | 2 | | | X | | | | X |
| | | 4 | | | X | | | | X |
| | | 6 | | | X | | | | X |
| | | 8 | | | X | X | | | X |
| | 2 | 0 | X | | X | X | | | X |
| | 3 | 0 | X | | | | | | |
| | 4 | 0 | X | X | | | | | |
| | 5 | 0 | X | | | | | | |
| | 8 | 0 | X | X | X | X | | | X |
| | | 2 | | | X | | | | |
| | 11 | 0 | X | X | | | | | |
| | 15 | 0 | X | X | X | X | | X[f] | X |
| | 18 | 0 | X | X | | | | | |
| | 22 | 0 | X | X | X | X | | | X |
| | 25 | 0 | X | X | | | | | |

**Table 13: Part III Arms A and B, Interval Dosing of FHD-286: Timing of Clinical Laboratory, Pharmacokinetic, Pharmacodynamic, and Exploratory Sampling and Electrocardiograms**

| Cycle | Study Day | Hr[a] | Clinical Laboratory Testing[b] | | PK Blood | PD Blood | Exploratory Blood | PK Urine | ECG[c] |
|---|---|---|---|---|---|---|---|---|---|
| | | | Hematology/ Chemistry/ Coagulation | Inflammatory Markers/ Cytokines | | | | | |
| 2 | 1 | 0 | X | X | X | X | X | | X |
| | | 0.5 | | | X | | | | |
| | | 1 | | | X | | | | |
| | | 2 | | | X | | | | X |
| | | 4 | | | X | | | | X |
| | | 6 | | | X | | | | X |
| | | 8 | | | X | | | | X |
| | 8 | 0 | X | X | X | | | | |
| | 15 | 0 | X | X | X | | | | |
| 3 | 1 | 0 | X | X | X | X | X | | X |
| | | 4 | | | X | | | | X |
| | | 8 | | | X | | | | |
| | 8 | 0 | X | X | X | | | | |
| | 15 | 0 | X | X | X | | | | |
| 4 | 1 | 0 | X | X | X | | | | X |
| | 15 | 0 | X | | | | | | |
| 5+ | 1 | 0 | X | X | X | X[g] | X[g] | | X |
| | 15 | 0 | X | | | | | | |
| EOT[h] | | | X | X | X | X | X | | |
| Safety Follow-up Visit[i] | | | X | | | | | | |

Abbreviations: ECG = electrocardiogram; EOT = End of Treatment; PD = pharmacodynamics; PK = pharmacokinetics.

[a] Relative to the FHD-286 dose. Hour 0 samples for clinical laboratory testing to be collected within 12 hours before dose. All other Hour 0 samples to be collected within 2 hours before dose. Samples at all other hours to be collected within ±10 minutes of specified time. ECGs should be collected within approximately 10 minutes prior to PK sample collection.

[b] Clinical laboratory parameters are defined in Table 14. All samples will be analyzed locally. In Cycle 1, assessments on Days 4, 11, 18, and 25 may be performed within ±1 day of the specified time point. In Cycle 1, assessments on Days 11, 18, and 25 may be performed at a healthcare provider local to the subject.

[c] 12-lead ECGs will be collected in triplicate prior to PK sample collection (starting after approximately 3 minutes of recumbency or semi-recumbency; each ECG performed approximately 2 minutes apart). There is no associated PK sample collection for the Screening ECG; as such, the 12-lead Screening ECG may be performed in triplicate at any time during the Screening window.

[d] Within 28 days before the first dose of study treatment (Cycle 1 Day 1).

[e] A white blood cell count assessment must be completed within 72 hours before the subject's first scheduled dose of study treatment to confirm eligibility.

[f] Continuous urine collection over 24 hours, beginning at predose on Cycle 1 Day 15, will be performed for at least 3 subjects in Part III, who will be chosen in discussion with the Investigator(s) based on considerations such as the dose at which the subject is to be initiated and the individual subject's ability to participate in the urine collection assessment.
[g] After Cycle 3, PD and exploratory blood will be collected on Day 1 of every odd-numbered cycle (Cycles 5, 7, 9, and 11) for the first 48 weeks.
[h] EOT Visit will take place within 5 days after treatment ends (treatment end is defined as the date on which the Investigator decided to discontinue FHD-286 treatment and is not required to be equal to the date of the last FHD-286 dose).
[i] Safety Follow-up Visit will occur 28 days ±7 days after the EOT Visit.
Notes:
1. Study treatment should be administered on site on days that PK and/or PD assessments will take place.
2. Samples for PK may also be used for other exploratory analysis, including but not limited to metabolite profiling and additional biomarker analysis.
3. Subjects may be requested to undergo unscheduled PK/PD/safety assessments and ECGs if a safety event occurs.

#### 10.2. Informed Consent

A complete description of the study is to be presented to each potential subject and a signed and dated informed consent (and signed and dated informed assent for subjects <18 years of age) is to be obtained before any study-specific procedures are performed.

#### 10.3. Demographic Data and Medical, Surgical, and Medication History

According to applicable local regulations, subject demographic data, including gender, date of birth, age, race, and ethnicity, will be obtained during screening.

A complete medical and surgical history, including the type of underlying malignancy and the date of confirmation of the histologic diagnosis of the underlying malignancy, will be obtained during screening. The medical history is to include all relevant prior medical history, including prior diagnoses of differentiation syndrome, as well as all current medical conditions.

All medications or herbal supplements administered and all procedures conducted within 28 days prior to C1D1 should be reported in the eCRF. In addition, all prior treatment regimens for the underlying malignancy will be reported.

#### 10.4. Safety Assessments

Refer to the Schedules of Assessments (Section 10.1) for the time points at which safety assessments are to be conducted.

##### 10.4.1. Physical Examination and ECOG Performance Status

A complete physical examination, including assessment of height and weight, will be obtained at the Screening Visit and at the EOT Visit. The complete physical examination will include assessment of general appearance and a review of systems (dermatologic, head, eyes, ears, nose, mouth/throat/neck, thyroid, lymph nodes, respiratory, cardiovascular, gastrointestinal, extremities, musculoskeletal, and neurologic systems).

A limited physical examination will be completed at visits during the treatment period. The limited physical examination will include assessment of general appearance and a review of the dermatologic, respiratory, and cardiovascular systems. Additional body systems should be assessed based on reported AEs. Height does not need to be obtained as part of the limited physical examination.

For subjects in Part III, weight will be collected on Day 1 of each cycle; institutional policy regarding body surface area calculations should be followed to determine the LDAC or decitabine dose to be administered.

Determination of ECOG PS will be performed. See Appendix 15.7 for ECOG PS scoring.

##### 10.4.2. Vital Signs

Vital signs, including systolic and diastolic blood pressure, heart rate, respiratory rate, and temperature, will be obtained. Assessments should be conducted while the subject is seated or supine.

##### 10.4.3. Echocardiograms

Echocardiography (other methods of evaluating LVEF may be performed according to institutional practice) to assess left ventricular systolic function via LVEF measurement will be performed.

##### 10.4.4. Electrocardiograms

Standard safety 12-lead ECGs in triplicate are to be obtained at the specified time points and as clinically indicated. A cardiology consultation is recommended as clinically indicated for prompt management of treatment-emergent cardiac arrhythmias and/or other clinically significant cardiac findings. Refer to Section 9.7 for dose modification criteria and monitoring measures for QTc prolongation.

The 12-lead ECGs should be obtained after approximately 3 minutes of recumbency or semi-recumbency, and each ECG should be spaced approximately 2 minutes apart.

All ECGs will be collected by a central vendor. The central vendor will also provide ECG equipment and training to all study sites.

##### 10.4.5. Safety Laboratory Assessments

Clinical laboratory evaluations are to be performed by the site's local laboratory. Prior to starting the study, the Investigator will provide to the Sponsor (or its designee) copies of all laboratory certifications and normal ranges for all laboratory parameters to be performed by that laboratory.

Clinical laboratory evaluations are to be collected at the specified time points and as clinically indicated. In addition, all clinically significant laboratory abnormalities noted on testing will be followed by repeat testing and further investigated according to the judgment of the Investigator.

The clinical laboratory parameters to be determined are specified in Table 14.

**Note for Part III (Combination Therapy):** Subjects' WBC count will be assessed before the initiation of study treatment as part of the predose safety laboratory assessments on Cycle 1 Day 1. If the count exceeds $20\times10^9$/L, study treatment will not be initiated until the WBC count is $\leq 20\times10^9$/L.

**Table 14: Clinical Laboratory Tests**

| **Hematology** | **Coagulation** | **Serum Chemistry** |
|---|---|---|
| RBC | PT/INR | Sodium |
| WBC with differential | aPTT | Potassium |
| Blast count | Fibrinogen | Chloride |
| Platelets | **Inflammatory Markers** | Bicarbonate or $CO_2$ |
| Hemoglobin | C-reactive protein | Urea or blood urea nitrogen[a] |
| Hematocrit | Ferritin | Creatinine |
| MCV | **Cytokine Panel** | Creatine kinase |
| MCH | IL-6 | AST |
| MCHC | IFN-γ | Troponin (I or T)[a] |
| RDW | TNF-α | BNP or NT-proBNP[a] |
| Reticulocyte count | | ALT |
| | | ALP |
| | | Bilirubin (total and direct) |
| | | Glucose |
| | | LDH |
| | | Calcium |
| | | Albumin |
| | | Total protein |
| | | Uric acid |
| | | Magnesium |
| | | Phosphorous |

Abbreviations: ALT = alanine aminotransferase; ALP = alkaline phosphatase; aPTT = activated partial thromboplastin time; AST = aspartate aminotransferase; BNP = B-type natriuretic peptide; $CO_2$ = carbon dioxide; CST = Clinical Study Team; IFN-γ = interferon gamma; IL-6 = interleukin 6; INR = international normalized ratio; LDH = lactate dehydrogenase; MCH = mean corpuscular hemoglobin; MCHC = mean corpuscular hemoglobin concentration; MCV = mean corpuscular volume; NT-proBNP = N-terminal pro hormone BNP; PT = prothrombin time; RBC = red blood cell; RDW = red cell distribution width; TNF-α = tumor necrosis factor alpha; WBC = white blood cell.

Note: If a laboratory value is found to be clinically significantly abnormal, repeat blood work will be performed at time intervals determined by the CST. If the laboratory value is persistently abnormal, then additional testing, closer monitoring, and/or possible discontinuation of the study treatment will be considered.

[a] The same assessment to evaluate urea, troponin, and BNP should be used at all required time points, when possible.

##### 10.4.6. Adverse Events

Each subject must be carefully monitored for the development of any AEs throughout the study from signing of the informed consent/assent to 28 days after the last FHD-286 dose. In addition, SAEs that are assessed as related to study treatment that occur >28 days after the last FHD-286 dose are to be reported.

This information should be obtained in the form of non-leading questions (eg, "How are you feeling?"), and from signs and symptoms detected during each examination, from laboratory evaluation, observations by study personnel, and spontaneous reports from subjects.

All AEs will be graded using the NCI CTCAE version 5.0 grading system (Appendix 15.1).

Complete details on AE monitoring are provided in Section 11.

#### 10.5. Bone Marrow Samples and Peripheral Blood Leukemic Blast Cells

Bone marrow biopsies and/or aspirates are to be obtained at the time points noted in Section 10.1 and as clinically indicated, independent of study treatment delays and/or interruptions.

Bone marrow aspirates and core sampling should be performed according to standard of care and analyzed at the local site's laboratory in accordance with the International Council for Standardization in Hematology Guidelines (Lee, et al 2008).

The diagnosis and evaluation of AML, MDS, or CMML can be made by bone marrow aspiration when a core sample is unobtainable and/or is not a part of the standard of care. A bone marrow biopsy is required in case of dry tap or failure (mainly dilution) with the aspiration.

Standard assessments for bone marrow core biopsies and aspirates will include morphology, flow cytometry, and karyotype to assess potential clinical activity (see Section 10.6).

#### 10.6. Clinical Activity Assessments

The clinical activity of study treatment will be evaluated by assessing preliminary response to treatment according to modified IWG criteria for AML and MDS (Cheson, et al 2003; Cheson, et al 2006), as well as additional response criteria. The modified IWG criteria for MDS will be used for CMML as well.

Disease response to treatment will be assessed through the evaluation of bone marrow biopsies and/or aspirates, along with complete blood counts and examination of peripheral blood films.

Subjects will have the extent of their disease assessed and recorded at the time points indicated in the Schedules of Assessments (Section 10.1), independent of study treatment delays and/or dose interruptions, and as clinically indicated.

The criteria outlined in Table 15, Table 16, and Table 17 will be used to assess response to treatment in subjects with AML. The criteria outlined in Table 18 and Table 19 will be used to assess response to treatment in subjects with MDS and CMML.

**Table 15: Proposed Modified International Working Group Response Criteria for Acute Myeloid Leukemia**

| Category | Definition |
|---|---|
| Complete remission (CR)* | Bone marrow blasts <5%; absence of blasts with Auer rods; absence of extramedullary disease; absolute neutrophil count >$1.0\times10^9$/L (1000/µL); platelet count >$100\times10^9$/L (100,000/µL); independence of red cell transfusions |
| CR with incomplete platelet recovery (CRp) | All CR criteria except for residual thrombocytopenia (platelet counts <$100\times10^9$/L [100,000/µL]) |
| CR with incomplete blood count recovery (CRi)• | All CR criteria except for residual neutropenia (absolute neutrophil count <$1.0\times10^9$/L [1000/µL]) |
| Morphologic leukemia-free state (MLFS)∆ | Bone marrow blasts <5 percent; absence of blasts with Auer rods; absence of extramedullary disease; no hematologic recovery required |
| Partial remission (PR) | Relevant in the setting of Phase 1 and 2 clinical trials only; all hematologic criteria of CR; decrease of bone marrow blast percentage to 5 to 25 percent; and decrease of pretreatment bone marrow blast percentage by at least 50 percent |
| Cytogenetic CR (CRc)◊ | Reversion to a normal karyotype at the time of morphologic CR (or CRi) in cases with an abnormal karyotype at the time of diagnosis; based on the evaluation of 20 metaphase cells from bone marrow |
| Molecular CR (CRm)§ | No standard definition; depends on molecular target |
| **Treatment failure** | |
| Resistant disease (RD) | Failure to achieve CR or CRi (general practice; Phase 2/3 trials), or failure to achieve CR, CRi, or PR (Phase 1 trials); only includes subjects surviving ≥7 days following completion of initial treatment, with evidence of persistent leukemia by blood and/or bone marrow examination |
| Death in aplasia | Deaths occurring ≥7 days following completion of initial treatment while cytopenic; with an aplastic or hypoplastic bone marrow obtained within 7 days of death, without evidence of persistent leukemia |
| Death from indeterminate cause | Deaths occurring before completion of therapy, or <7 days following its completion; or deaths occurring ≥7 days following completion of initial therapy with no blasts in the blood, but no bone marrow examination available |
| Relapse (defined only for subjects who have previously attained CR, CRi, CRp, or MLFS)¥ | Bone marrow blasts ≥5 percent; or reappearance of blasts in the blood; or development of extramedullary disease |

##### Table 15: Proposed Modified International Working Group Response Criteria for Acute Myeloid Leukemia

Source: Cheson, et al (2003).
Abbreviations: AML = acute myeloid leukemia; eCRF = electronic case report form; MDS = myelodysplastic syndromes.
* All criteria needed to be fulfilled; marrow evaluation was to be based on a count of 200 nucleated cells in an aspirate with spicules; if ambiguous, repeat exam after 5 to 7 days was to be considered; flow cytometric evaluation could help to distinguish between persistent leukemia and regenerating normal marrow; a marrow biopsy was to be performed in cases of dry tap, or if no spicules are obtained; no minimum duration of response required.
• The criterion of CRi was of value in protocols using intensified induction or double induction strategies, in which hematologic recovery was not awaited, but intensive therapy was to be continued. In such protocols, CR could even not be achieved in the course of the entire treatment plan. In these instances, the overall remission rate was to include CR and CRi subjects. Some subjects may not have achieved complete hematologic recovery upon longer observation times.
Δ This category may have been useful in the clinical development of novel agents within Phase 1 clinical trials, in which a transient morphologic leukemia-free state could be achieved at the time of early response assessment.
◊ Four studies showed that failure to convert to a normal karyotype at the time of CR predicted inferior outcome.
§ As an example, in core-binding factor AML low-level polymerase chain reaction-positivity was detected in subjects even in long-term remission. Normalizing to $10^4$ copies of *ABL1* in accordance with standardized criteria, transcript levels below 10 to 12 copies appeared to be predictive for long-term remission.

¥ A repeat marrow was to be performed to confirm relapse with 2 consecutive assessments separated by at least a month. Appearance of new dysplastic changes were to be closely monitored for emerging relapse. In a subject who had been recently treated, dysplasia or a transient increase in blasts may have reflected a chemotherapy effect and recovery of hematopoiesis. Cytogenetics were to be tested to distinguish true relapse from therapy-related MDS/AML.

##### Table 16: Additional Response Criteria for Acute Myeloid Leukemia

| Category | Definition |
|---|---|
| CR with partial hematologic recovery (CRh) | For documentation of CRh, FDA has used the following definition:<br>− Marrow blasts <5% by morphological examination,<br>− ANC >0.5 Gi/L (>0.5×$10^9$/L (500/µL); and platelet count >50 Gi/L (>50×$10^9$/L (50,000/µL), but the count recovery criteria for CR are not met,<br>− Absence of leukemic blasts in the peripheral blood by morphological examination, and<br>− No evidence of extramedullary disease. |
| Transfusion independence | The absence of red blood cell and platelet transfusions for 28 days during continued treatment. For patients with active AML, transfusion dependence at baseline is based on the receipt of any red blood cell or platelet transfusions within at least 28 days prior to the start of study treatment. |

Abbreviations: AML = acute myeloid leukemia; ANC = absolute neutrophil count; CR = complete remission.

**Table 17: Criteria for Stable Disease and Progressive Disease for Acute Myeloid Leukemia**

| Category | Response Criteria |
|---|---|
| Stable disease | Failure to achieve a response and not meeting the criteria for disease progression |
| Disease progression (defined only for subjects who have previously attained PR or stable disease) | Progression was defined as the following:<br>For subjects with 5 to 67% bone marrow blasts at nadir:<br>a >50% increase in bone marrow blast count percentage from the nadir and that was ≥20%.<br>For subjects with ≥67% bone marrow blasts at nadir:<br>a doubling of the nadir absolute peripheral blood blast count and the final absolute peripheral blood blast count was $>10 \times 10^9$/L.<br>Progression should be confirmed by 2 consecutive assessments separated by at least 1 month.<br>Development of new biopsy-confirmed extramedullary disease since last disease evaluation<br>The date of progressive disease was defined as the first date of progression. |

**Table 18: Proposed Modified International Working Group Response Criteria for Altering Natural History of Myelodysplastic Syndromes**

| Category | Response Criteria (Responses Must Last At Least 4 Weeks) |
|---|---|
| CR | Bone marrow: ≤5% myeloblasts with normal maturation of all cell lines*<br>Persistent dysplasia will be noted*†<br>Peripheral blood‡<br>Hgb ≥11 g/dL<br>Platelets $\geq 100 \times 10^9$/L<br>Neutrophils $\geq 1.0 \times 10^9$/L†<br>Blasts = 0% |
| PR | All CR criteria if abnormal before treatment except:<br>Bone marrow blasts decreased by ≥50% over pretreatment but still >5%<br>Cellularity and morphology not relevant |
| Marrow CR | Bone marrow: ≤5% myeloblasts and decrease by ≥50% over pretreatment†<br>Peripheral blood: if HI responses, they will be noted in addition to marrow CR† |
| Stable disease | Failure to achieve at least PR, but no evidence of progression for >8 weeks |
| Failure | Death during treatment or disease progression characterized by worsening of cytopenias, increase in percentage of bone marrow blasts, or progression to a more advanced MDS FAB subtype than pretreatment |
| Relapse after CR or PR | At least 1 of the following:<br>Return to pretreatment bone marrow blast percentage<br>Decrement of ≥50% from maximum remission/response levels in granulocytes or platelets<br>Reduction in Hgb concentration by ≥1.5 g/dL or transfusion dependence |

##### Table 18: Proposed Modified International Working Group Response Criteria for Altering Natural History of Myelodysplastic Syndromes

| Category | Response Criteria (Responses Must Last At Least 4 Weeks) |
|---|---|
| Cytogenetic response | Complete: Disappearance of the chromosomal abnormality without appearance of new ones<br>Partial: At least 50% reduction of the chromosomal abnormality |
| Disease progression | For subjects with:<br>Less than 5% blasts: ≥50% increase in blasts to >5% blasts<br>5%-10% blasts: ≥50% increase to >10% blasts<br>10%-20% blasts: ≥50% increase to >20% blasts<br>20%-30% blasts: ≥50% increase to >30%<br>Any of the following:<br>At least 50% decrement from maximum remission/response in granulocytes or platelets<br>Reduction in Hgb by ≥2 g/dL<br>Transfusion dependence |

Source: Cheson, et al (2006).
Abbreviations: AML = acute myeloid leukemia; CR = complete remission; DFS = disease-free survival; FAB = French-American-British; Hgb = hemoglobin; HI = hematologic improvement; MDS = myelodysplastic syndromes; PFS = progression-free survival; PR = partial remission.
* Dysplastic changes should consider the normal range of dysplastic changes (modification).
† Modification to IWG response criteria (Cheson, et al 2003).
‡ In some circumstances, protocol therapy may require the initiation of further treatment (eg, consolidation, maintenance) before the 4-week period. Such subjects can be included in the response category into which they fit at the time the therapy is started. Transient cytopenias during repeated chemotherapy courses should not be considered as interrupting durability of response, as long as they recover to the improved counts of the previous course.

##### Table 19: Proposed Modified International Working Group Response Criteria for Hematologic Improvement

| Hematologic Improvement* | Response Criteria (Responses Must Last At Least 8 Weeks)† |
|---|---|
| Erythroid response (pretreatment, <11 g/dL) | Hgb increase by ≥1.5 g/dL<br>Relevant reduction of units of RBC transfusions by an absolute number of at least 4 RBC transfusions/8 wk compared with the pretreatment transfusion number in the previous 8 wk. Only RBC transfusions given for a Hgb of ≤9.0 g/dL pretreatment will count in the RBC transfusion response evaluation† |
| Platelet response (pretreatment, $<100\times10^9$/L) | Absolute increase of $\geq30\times10^9$/L for subjects starting with $>20\times10^9$/L platelets<br>Increase from $<20\times10^9$/L to $>20\times10^9$/L and by at least 100%† |
| Neutrophil response (pretreatment, $<1.0\times10^9$/L) | At least 100% increase and an absolute increase $>0.5\times10^9$/L† |
| Progression or relapse after HI‡ | At least 1 of the following:<br>At least 50% decrement from maximum response levels in granulocytes or platelets<br>Reduction in Hgb by >1.5 g/dL<br>Transfusion dependence |

Source: Cheson, et al (2006).
Abbreviations: Hgb indicates hemoglobin; RBC: red blood cell; HI: hematologic improvement.
Note: Deletions to the IWG response criteria are not shown.

**Table 19: Proposed Modified International Working Group Response Criteria for Hematologic Improvement**

| Hematologic Improvement* | Response Criteria (Responses Must Last At Least 8 Weeks)† |
|---|---|

Note: To convert hemoglobin from g/L to g/dL, divide g/L by 10.
* Pretreatment counts averages of at least 2 measurements (not influenced by transfusions) ≥1 week apart (modification).
† Modification to IWG response criteria (Cheson, et al 2003).
‡ In the absence of another explanation, such as acute infection, repeated courses of chemotherapy (modification), gastrointestinal bleeding, hemolysis, and so forth. It is recommended that the 2 kinds of erythroid and platelet responses be reported overall as well as by the individual response pattern.

After subjects have discontinued FHD-286, unless consent/assent to participate is withdrawn, they will be contacted every 2 months (±14 days) for long-term follow-up. Long-term follow-up includes assessment of:

- Survival status
- Receipt and type of subsequent anticancer therapy
- Disease status, including date of most recent evaluation (unless subject discontinued FHD-286 due to progressive disease or treatment failure)
  - Collection of disease status will only continue until the subject experiences disease progression or treatment failure, or starts a new anticancer therapy.

Long-term follow-up will continue until 2 years after the last subject discontinues FHD-286 or until all subjects have died, withdrawn consent/assent, or are lost to follow-up, whichever occurs first.

#### 10.7. Exploratory Tissue Samples

In this study, biomarker analyses from bone marrow aspirate and biopsies and peripheral tissue samples (eg, blood) will be used to investigate the effect of FHD-286 at the molecular and cellular level, as well as to determine how changes in the markers may relate to exposure and clinical outcomes, and to identify predictive response markers.

**Part III Only:** If a subject in Group A1 or B1 begins treatment with a triazole antifungal agent classified as a strong CYP3A4 inhibitor after their first dose of FHD-286, additional PD blood samples may be collected concurrently with additional serial PK blood samples.

Peripheral tissue samples (eg, blood) will be collected and utilized for the assessment and analysis of several molecular and cellular entities, including, but not limited to, PBMC subpopulations, leukemic blasts, circulating tumor cells, circulating tumor DNA, and cell-free RNA, as well as circulating immune cells, exosomes, cytokines, and secreted proteins.

To the extent feasible, tumor and peripheral samples will be analyzed using standard and exploratory techniques to monitor several molecular features, including genomic, immunophenotypic, immune cell function, transcriptomic, and proteomic features (including analysis of serum cytokines), to determine if these correlate with efficacy, or reveal novel prognostic or cancer-related biomarkers of clinical utility. These assessments will include, but are not limited to, the determination of the mutational and copy number status and expression levels of genes of interest, such as *SMARCA2*/*BRM*, *SMARCA4*/*BRG1*, and other SWI/SNF- and cancer-related genes. In addition, levels of BAF target genes, immune checkpoints, immune cell subpopulations, apoptotic markers, and other features may be assessed using immunohistochemistry, fluorescence-activated cell sorting analysis, immunofluorescence, or other relevant methods. Exploratory techniques, such as RNA sequencing, may be employed to measure baseline expression and expression level changes mediated by BAF inhibition and to confirm target engagement within tumor and normal cells.

The remaining biological samples may be stored for up to 10 years after the end of study and further analyzed to address scientific questions related to FHD-286 and/or cancer. This may include research to develop ways to detect, monitor, or treat cancer. A decision to perform such exploratory biomarker research studies would be based on outcome data from this study or from new scientific findings related to the drug class or disease, as well as reagent and assay availability.

While the goal of the biomarker assessments is to provide supportive data for the clinical study, there may be circumstances when a decision is made to stop a collection, or not perform or discontinue an analysis due to either practical or strategic reasons (eg, inadequate sample number, issues related to the quality of the sample or issues related to the assay that preclude analysis, impossibility to perform correlative analyses). Therefore, depending on the results obtained during the study, sample collection and/or analysis may be omitted at the discretion of the Sponsor.

If a decision is made by the Sponsor to stop collection of any biomarker or biopsy samples, notification will be provided to every Investigator in writing.

#### 10.8. Pharmacokinetic Assessments

PK blood samples will be collected at the time points specified in Section 10.1. Those samples will be used to determine FHD-286 concentrations in the plasma and to understand the PK behaviors of this compound. Other exploratory analysis, including but not limited to metabolite profiling and additional biomarker analysis, may be conducted using the residual plasma samples, as appropriate.

**Part III Only:** If a subject in Group A1 or B1 begins treatment with a triazole antifungal agent classified as a strong CYP3A4 inhibitor after their first dose of FHD-286, additional serial PK blood samples may be collected.

Continuous urine collection will be performed for 24 hours starting on C1D15 for at least 3 subjects participating in Part III of this study. These subjects will be chosen in discussion with Investigator(s) based on considerations such as the dose at which the subject is to be initiated and the individual subject's ability to participate in the urine collection assessment.

Subjects participating in urine collection are to empty their bladder just before the administration of study treatment on C1D15. Continuous urine collection will then be performed while the subject is on site (ie, through 8 hours postdose for subjects receiving FHD-286 QD). Subjects will be provided a urine collection vessel in which to collect urine continuously until their return to the study site the following day (C1D16), at which point they will be instructed to bring the urine collected at home to the study site.

#### 10.9. Pharmacodynamic and Exploratory Assessments

Bone marrow biopsy and/or aspirate and peripheral tissue samples of processed blood will be requested to assess PD biomarkers and to explore molecular, epigenetic, and proteomic changes at the time points specified in Section 10.1.

#### 10.10. Sample Processing, Storage, and Shipment

Instructions for the collection, processing, storage, and shipment of all study samples for central analysis will be provided in the Study Laboratory Manual.

### 11. ADVERSE EVENTS

Monitoring of AEs will be conducted throughout the study after subjects sign the informed consent form or informed assent form (for subjects <18 years of age).

#### 11.1. Definition of Adverse Events

##### 11.1.1. Adverse Event

An AE is any untoward medical occurrence associated with the use of a drug in humans, whether or not considered drug related. An AE (also referred to as an adverse experience) can:

- Be any unfavorable and unintended sign (eg, an abnormal laboratory finding), symptom, or disease temporally associated with the use of a drug, whether or not considered related to the drug
- Arise from any use of the drug (eg, use in combination with another drug) and from any route of administration, formulation, or dose, including an overdose
- Arise from development of a new disease or the worsening of an existing disease or recurrence of an intermittent medical condition not present at baseline
- Be any deterioration in laboratory value and/or other clinical test that is associated with symptoms or leads to a change in study treatment or concomitant treatment or discontinuation from study treatment
- Arise related to protocol procedures (including those occurring prior to initiating treatment with a study drug)

Disease progression or death due to progression of the underlying malignancy will not be considered an AE (or an SAE) in this study but will be collected as an outcome or reason for discontinuation, as appropriate. AEs (or SAEs) considered as complications of disease progression that occur during the AE reporting period should be reported.

##### 11.1.2. Adverse Events of Special Interest

Differentiation syndrome is an AESI with FHD-286. Differentiation syndrome includes a constellation of signs and symptoms caused by the rapid release of a high level of cytokines from leukemia cells. These signs and symptoms are also frequently observed with other relatively common complications of advanced hematologic malignancies. Any event of Grade ≥1 Differentiation syndrome should be reported, at minimum, as an important medical event to the Sponsor within 24 hours of awareness (Section 11.2).

##### 11.1.3. Suspected Adverse Reaction

A suspected adverse reaction is any AE for which there is a reasonable possibility that the drug caused the AE. For the purposes of expedited safety reporting, "reasonable possibility" means there is evidence to suggest a causal relationship between the drug and the AE.

##### 11.1.4. Unexpected Adverse Event

An unexpected AE is one for which the nature or severity of the event is not consistent with the applicable product information, eg, the IB or regional product label.

##### 11.1.5. Serious Adverse Event

An SAE is defined as any untoward medical occurrence that at any dose:

- Results in death;
- Is life-threatening, meaning that the subject was at risk of death at the time of the event but does not mean that the event hypothetically might have caused death if it were more severe;
- Requires hospitalization or prolongation of existing hospitalization;
- Results in persistent or significant disability or incapacitation;
- Is a congenital anomaly or birth defect; or
- Is another important medical event (see below).

Death is an outcome of an SAE and not an SAE in and of itself. When death is an outcome, the underlying medical diagnosis or suspected diagnosis that is deemed the primary condition leading to the fatal outcome should be reported as the SAE term (eg, "pulmonary embolism"). Generally, only 1 such event should be reported.

The term "sudden death" should only be used for the occurrence of an abrupt and unexpected death due to presumed cardiac causes in a subject with or without pre-existing heart disease, within 1 hour of the onset of acute symptoms or, in the case of an unwitnessed death, within 24 hours after the subject was last seen alive and stable. If the cause of death is unknown and cannot be ascertained at the time of reporting, "unexplained death" should be recorded on the Adverse Event eCRF. If the cause of death later becomes available (eg, after autopsy), "unexplained death" should be replaced by the established cause of death.

Hospitalization admissions and/or surgical operations scheduled to occur during the study period, but planned prior to study entry are not considered AEs if the illness or disease existed before the subject was enrolled in the study, provided that it did not deteriorate in an unexpected manner during the study (eg, surgery performed earlier than planned).

Important medical events that do not result in death, are not life-threatening, or do not require hospitalization may be considered SAEs when, based upon appropriate medical judgment, they may jeopardize the subject and may require medical or surgical intervention to prevent one of the outcomes listed above. Examples of such medical events include allergic bronchospasm requiring intensive treatment in an emergency room or at home, blood dyscrasias or convulsions that do not result in in-patient hospitalization, or the development of drug dependency or drug abuse.

#### 11.2. Procedures for Reporting Adverse Events and Serious Adverse Events

The Investigator is responsible for ensuring that each subject is carefully monitored for the development of any AEs. This information should be obtained in the form of non-leading questions (eg, "How are you feeling?") and from signs and symptoms detected during each examination, observations of study personnel, and spontaneous reports from subjects.

After informed consent/assent has/have been obtained but prior to initiation of study treatment, only SAEs caused by a protocol-mandated intervention should be reported (eg, SAEs related to invasive study procedures such as biopsies). After initiation of study treatment, all AEs and SAEs regardless of attribution should be reported until 28 days after the last administration of FHD-286, unless the subject has withdrawn consent/assent for study participation, or started a new anticancer treatment for the disease under study, whichever occurs first. Subjects will be assessed at the Safety Follow-up Visit to determine if any new AEs have occurred. Hospitalization to start a new anticancer treatment for the disease under study after study treatment has been permanently discontinued does not meet AE reporting criteria. After completion of the Safety Follow-up Visit, Investigators should report only SAEs that are considered to be related to study treatment.

All AEs (serious and non-serious) spontaneously reported by the subject and/or in response to an open question from study personnel or revealed by observation, physical examination, or other diagnostic procedures will be recorded on the Adverse Event eCRF. Any clinically relevant deterioration in laboratory assessments or other clinical findings is considered an AE and must be recorded on the Adverse Event eCRF. When possible, signs and symptoms indicating a common underlying pathology should be noted as 1 comprehensive event rather than as individual events.

All SAEs are to be reported immediately/no later than within 24 hours of awareness by the Investigator to the Foghorn Therapeutics Global Safety and Pharmacovigilance Contact (see table below). All SAEs must be reported whether or not they are considered causally related to FHD-286 or other study treatments (eg, LDAC or decitabine). An SAE form will be completed, and the information collected will include, at minimum, subject number, event term(s), an assessment by the Investigator as to the severity and relatedness to study treatment and a narrative description of the event. Follow-up information on the SAE may be requested by the Sponsor.

| **Foghorn Therapeutics Global Safety and Pharmacovigilance Contact Information:** |
|---|
| [redacted] |
| SAE Fax: [redacted] |
| E-mail: [redacted] |

If there are serious, unexpected adverse drug reactions associated with the use of FHD-286, LDAC, or decitabine, the Sponsor will notify the appropriate regulatory agency(ies) and all participating Investigators on an expedited basis. The local Institutional Review Board (IRB)/Independent Ethics Committee (IEC) will be promptly notified based on local regulations where required by the IRB/IEC of all serious, unexpected adverse drug reactions involving risk to human subjects.

All AEs, whether serious or not, will be described in the source documents and on the AE page of the eCRF. All new events, as well as those that worsen in severity or frequency relative to baseline, which occur after signing the informed consent/assent through 28 days after the last dose of FHD-286, must be recorded. AEs that are ongoing at the time of treatment discontinuation should be followed as described in Section 11.3. Serious AEs felt by the Investigator to be related to FHD-286 must be reported any time the Investigator becomes aware of such an event, even if this occurrence is more than 28 days after the last dose of FHD-286.

Information to be reported in the description of each AE includes:

- A medical diagnosis of the event (if a medical diagnosis cannot be determined, a description of each sign or symptom characterizing the event should be recorded)
- The date of onset of the event
- The date of resolution of the event
- Whether the event is serious or not
- Severity of the event (see below for definitions)
- Relationship of the event to study treatment (see below for definitions)
- Action taken
- Outcome

**Severity** of all AEs, including clinically significant treatment-emergent laboratory abnormalities, will be graded according to the NCI CTCAE version 5.0 (Appendix 15.1). AEs not listed by the NCI CTCAE will be graded as follows:

- Grade 1 – Mild: the event is noticeable to the subject but does not interfere with routine activity.
- Grade 2 – Moderate: the event interferes with routine activity but responds to symptomatic therapy or rest.
- Grade 3 – Severe: the event significantly limits the subject’s ability to perform routine activities despite symptomatic therapy.
- Grade 4 – Life-threatening: an event in which the subject was at risk of death at the time of the event
- Grade 5 – Fatal: an event that results in the death of the subject.

**Relationship** to study treatment will be determined by the Investigator based on clinical judgement and according to the following criteria:

- **Not Related:** An AE will be considered “not related” to the use of FHD-286, LDAC, decitabine, or the combination of FHD-286 with LDAC or decitabine if there is no plausible temporal or causal relationship between the study treatment and AE (eg, exposure to the study treatment did not occur, or the occurrence of the AE is not reasonably related in time, or the AE is considered unlikely to be related to the study treatment).

- **Related:** An AE will be considered "related" to the use of FHD-286, LDAC, decitabine, or the combination of FHD-286 with LDAC or decitabine if there is a reasonable possibility of a temporal and causal relationship between the study treatment and AE. "Reasonable possibility" means there is evidence to suggest a causal relationship between the study treatment and the AE.

For the purpose of safety analyses, all AEs that are classified as related will be considered treatment-related AEs.

Alternative causes, such as underlying disease(s), concomitant therapy, and other risk factors, as well as the temporal relationship of the event to the study treatment, will be considered and investigated.

The Investigator may change opinion of causality in light of follow-up information.

#### 11.3. Follow-up of Adverse Events and Serious Adverse Events

After the initial AE/SAE report, the Investigator is required to actively follow each subject at subsequent visits/contacts. All SAEs and AESIs (as defined in Section 11.1.2) will be followed until resolution (ie, returned to baseline state of health), stabilization (ie, Investigator does not expect any further improvement or worsening of subject's condition), the subject is lost to follow-up, the subject withdraws consent/assent for study participation, or the subject has died (see Section 8.6).

#### 11.4. Regulatory Reporting Requirements for Serious Adverse Events

Prompt notification by the Investigator to the Sponsor is essential so that legal obligations and ethical responsibilities towards the safety of subjects and the safety of a study intervention under clinical investigation are met.

The Sponsor has a legal responsibility to notify regulatory agencies about the safety of a study intervention under investigation. The Sponsor will comply with regulatory requirements relating to the regulatory authority, IRB/IEC, and Investigators.

Investigator safety reports must be prepared for suspected unexpected serious adverse reactions according to local regulatory requirements and Sponsor policy and forwarded to the Investigators, as necessary.

An Investigator who receives an investigator safety report describing an SAE or other specific safety information from the Sponsor will review and file it along with the IB, and will notify the IRB/IEC, as appropriate, according to local requirements.

#### 11.5. Pregnancy Reporting

Pregnancy is neither an AE nor an SAE, unless a complication relating to the pregnancy occurs (eg, spontaneous abortion, which may qualify as an SAE) and/or unless there is a suspicion that FHD-286 may have interfered with the effectiveness of a contraceptive medication. However, any pregnancy in a participating female subject or partner of a male subject that occurs during this study or within 90 days after the last dose of FHD-286 must be reported to the Sponsor within 24 hours of being notified of the pregnancy (see Foghorn Therapeutics Global Safety and Pharmacovigilance Contact Information in Section 11.2). If a female subject becomes pregnant while on-study, study treatment should be immediately discontinued. The Investigator must follow up and document the course and outcome of all pregnancies even if the subject was discontinued from the study or if the study has finished. The female subject or partner of a male subject should receive any necessary counseling regarding the risks of continuing the pregnancy and the possible effects on the fetus. Monitoring should continue until conclusion of the pregnancy.

All outcomes of pregnancy must be reported by the Investigator to the Sponsor on a Pregnancy Outcome Report form within 30 days after they have gained knowledge of the delivery or elective abortion.

Any SAE that occurs during pregnancy must be recorded on the SAE report form (eg, maternal serious complications, spontaneous or therapeutic abortion, ectopic pregnancy, stillbirth, neonatal death, congenital anomaly, or birth defect) and reported within 24 hours in accordance with the procedure for reporting SAEs (Section 11.2)

All subjects, male and female, must agree to use effective contraception during the entire study and for the longest of the following:

- All subjects: 90 days after the last dose of FHD-286
- Subjects in Part III, Arm A (FHD-286 + LDAC): 6 months after the last dose of LDAC
- Subjects in Part III, Arm B (FHD-286 + decitabine):
  - Female: 6 months after the last dose of decitabine
  - Male: 90 days after the last dose of decitabine

### 12. STATISTICAL METHODS

#### 12.1. Sample Size Estimation

##### 12.1.1. Parts I and II: FHD-286 Monotherapy *(Closed to Enrollment)*

The sample size for the study will be dependent on the number of dose groups required and the incidence of DLTs required to determine the RP2D. It is estimated that approximately 25 to 50 subjects will enroll in Part I and II of this study, assuming 1 subject (Part I) and 3 subjects (Part II) per cohort (6 subjects at the MTD level) and a starting dose of 5 mg daily. Once the RP2D has been identified, an additional 6 to 12 subjects may be enrolled at that dose level(s) to confirm the observed safety and tolerability of FHD-286. These newly added subjects will be monitored with Bayesian Toxicity Monitoring (BTM) (see https://biostatistics.mdanderson.org/shinyapps/BTOX/). The BTM assumes a maximum probability of excess toxicity 0.2, prior distribution of Beta (1,1), maximum subjects 12, minimum number of subjects before pausing 4, and posterior probability 0.8; enrollment in this cohort will be paused for review for toxicities observed in 2 out of 6, 3 out of 10, or 4 out of 12 subjects.

Additional subjects may be enrolled during dose escalation, for the replacement of subjects who are not evaluable for the assessment of dose escalation, for evaluation of alternative dosing regimens, or for further exploring safety, PK, PK/PD, or preliminary clinical activity used to guide the selection of the RP2D.

##### 12.1.2. Part III: Combination Therapy

The sample size will depend on the number of dose levels required and the incidence of DLTs required to determine the RP2D(s). It is estimated that up to approximately 72 subjects will be enrolled in in Arm A (FHD-286 + LDAC) and up to approximately 72 subjects will be enrolled in Arm B (FHD-286 + decitabine) (up to approximately 144 subjects total).

Once the RP2D(s) has been identified for a group (A1, A2, B1, or B2), an additional 6 to 14 subjects may be enrolled at that dose level(s) in that group to confirm the observed safety and tolerability of FHD-286 in combination with LDAC or decitabine. These newly added subjects will be monitored with BTM (see https://biostatistics.mdanderson.org/shinyapps/BTOX/). The BTM assumes a maximum probability of excess toxicity 0.2, prior distribution of Beta (1,1), maximum subjects 14, minimum number of subjects before pausing 4, and posterior probability 0.8. Enrollment of these subjects will be paused for review if DLTs are observed in 2 of 6, 3 of 10, or 4 of 14 subjects. Additional stopping criteria will also be considered, as described in Section 9.8.

##### 12.1.3. Probability of Dose Escalation

Figure 4 presents the probability of escalation from a lower dose to the next higher dose, for a range of true rates of DLT, in the standard 3+3 dose-escalation design. For example, if the true DLT rate was 0.20 (20%), then the chance of dose escalation would be approximately 0.70 (70%).

**Figure 4: Probability of Dose Escalation for the 3+3 Design**

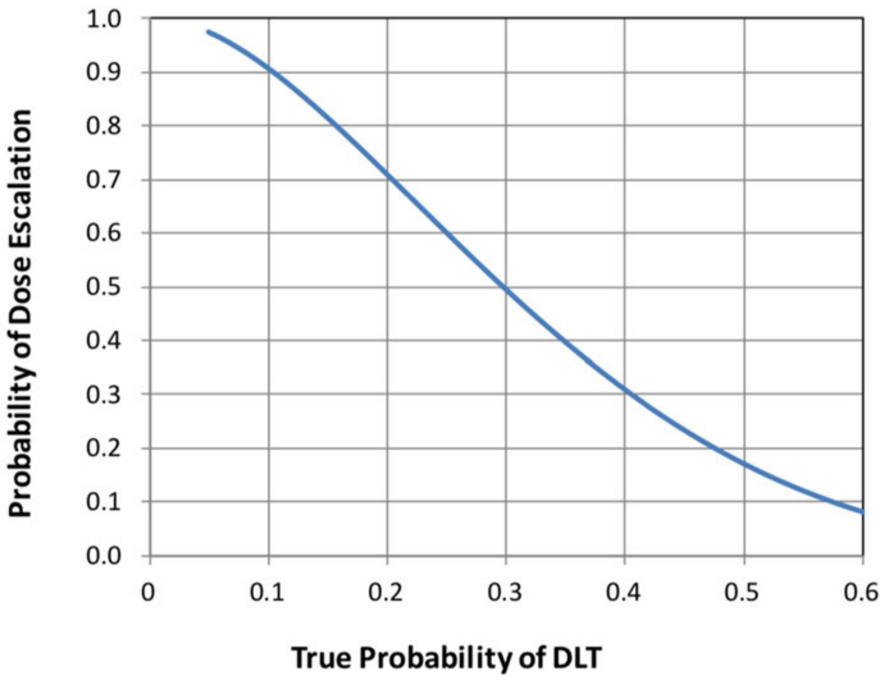

Abbreviation: DLT = dose-limiting toxicity.

#### 12.2. Populations for Analysis

The following subject populations (ie, analysis sets) will be evaluated and used for presentation of the data:

Safety Analysis Set/Full Analysis Set (FAS): All subjects who were enrolled and received at least 1 dose of study treatment. Subjects will be classified according to dose levels initially assigned. The Safety Analysis Set will be the primary set for the analysis of safety data. The FAS will be used for the summary of demographics, baseline characteristics, subject disposition, and analyses of all efficacy data.

Pharmacokinetic Analysis Set: All subjects who have at least 1 blood sample providing evaluable PK data for FHD-286.

#### 12.3. Procedures for Handling Missing, Unused, and Spurious Data

No imputation will be performed for missing data elements.

When tabulating AE data, partial dates will be handled as follows. If the day of the month is missing, the onset day will be set to the first day of the month unless it is the same month and year as first dose of study treatment. In this case, in order to conservatively report the event as treatment emergent, the onset date will be assumed to be the date of treatment. If the onset day and month are both missing, the day and month will be assumed to be 01 January, unless the event occurred in the same year as the first dose of study treatment. In this case, the event onset will be coded to the day of treatment in order to conservatively report the event as treatment emergent. A missing onset date will be coded as the day of treatment.

For the purposes of reporting, subjects who have not had an endpoint-defined event at the time of analyses will have time-to-event data (eg, duration of response and EFS) censored at the date of last documented disease assessment prior to the data cutoff date. Further details of censoring rules will be documented in the statistical analysis plan (SAP).

Within efficacy analysis, subjects with a best overall response of “Unknown” or “Not Evaluable” will be considered non-responders in estimating response rates.

#### 12.4. Interim Analyses

Not applicable as no formal interim analyses are planned.

#### 12.5. Independent Safety Monitoring Committee Analyses

Preplanned analyses of safety will be provided to the independent safety monitoring committee as specified in the committee charter.

#### 12.6. Statistical Methodology

##### 12.6.1. General Methods

Statistical analyses will be primarily descriptive. This will be achieved by the results of a deterministic algorithm; thus, statistical hypothesis testing is not intended for assessment of MTD or the RP2D(s).

Tabulations will be produced for disposition, demographic and baseline characteristics, safety, PK, PD, and clinical activity parameters. Data will be summarized by FHD-286 dose level initially assigned. FHD-286 monotherapy (Parts I and II) and combination therapy (Part III) will be summarized separately. For Part III, all groups (A1, A2, B1, and B2) will be summarized separately. For analyses of clinical activity, subjects with different indications will be summarized separately. Details of data handling and analysis methods will be described in an SAP for the study.

Categorical variables will be summarized by frequency distributions (number and percentages of subjects) and continuous variables will be summarized by descriptive statistics (mean, standard deviation, median, minimum, and maximum).

All data will be provided in by-subject listings.

The study data will be analyzed and reported in the primary clinical study report (CSR) based on all subjects’ data at the time when all subjects have completed at least 6 months of treatment with FHD-286 or discontinued FHD-286 earlier. Any additional data for subjects continuing to receive FHD-286 or who are in long-term follow-up past the data cutoff date for the primary CSR will be reported at the end of study.

##### 12.6.2. Disposition

A tabulation of the disposition of subjects will be presented, including the number treated, and the reasons for treatment and study discontinuation will be reported. Entry criteria and protocol deviations will be listed.

##### 12.6.3. Baseline Evaluations

Demographic and baseline disease characteristics data will be summarized using descriptive statistics. Data to be tabulated will include sex, age, and race and ethnicity, as well as disease-specific information.

##### 12.6.4. Exposure and Safety Analyses

Study treatment exposure, including number of doses administered, total dose received, duration of treatment, dose intensity, compliance, and the proportion of subjects with dose modifications, as appropriate, will be listed and summarized using descriptive statistics.

Treatment-emergent AEs are defined as any AEs that begin or worsen on or after the start of study treatment through 28 days after the last dose of FHD-286. All AEs will be listed. Only TEAEs will be summarized and will be referred to as AEs throughout. AEs will be summarized by Medical Dictionary for Regulatory Activities (version 23.1 or later) System Organ Class and Preferred Term. Separate tabulations will be produced for all AEs, treatment-related AEs (those considered by the Investigator as at least possibly related to study treatment), SAEs, dose modifications due to AEs, and AEs of at least Grade 3 severity. By-subject listings will be provided for AEs leading to death, SAEs, DLTs, and AEs leading to discontinuation of treatment.

AESIs (Section 11.1.2) will be summarized as detailed in the SAP.

Descriptive statistics will be provided for clinical laboratory parameters, ECG intervals, ECHOs (or other means) for LVEF, and vital signs data, presented as both actual values and changes from baseline relative to each on-study evaluation and to the last evaluation on study. Baseline is defined as the last non-missing result prior to the first dosing.

Shift tables of laboratory data from baseline to worst grade post-baseline will be presented based on NCI CTCAE version 5.0 (Appendix 15.1) grading. Shift tables also will be provided for ECOG PS from baseline to worst value on treatment.

##### 12.6.5. Pharmacokinetic Analyses

Descriptive statistics (ie, number of subjects, mean, standard deviation, geometric mean, coefficient of variation, median, minimum, and maximum) will be used to summarize PK parameters for each dose group and, where appropriate, for the entire PK Analysis Set. Such parameters will include (but are not limited to) $C_{max}$, $T_{max}$, AUC, $t_{1/2}$, and the fraction of drug excreted unchanged in the urine. The relationships between dose and both $C_{max}$ and AUC will be explored graphically for dose-proportionality. The potential relationship between PK and safety, efficacy, and PD will be explored with descriptive and graphical methods. FHD-286 exposure (AUC and $C_{max}$) will be descriptively compared between Groups A1 and A2, and between Groups B1 and B2. Full details of the PK analyses will be provided in a separate PK Analysis Plan.

##### 12.6.6. Pharmacodynamic Analyses

The potential relationship between plasma exposure of FHD-286 and PD markers will be described with descriptive and graphical methods.

##### 12.6.7. Clinical Activity Analyses

Response to treatment as assessed by the site Investigators using applicable response criteria (Section 10.6) will be tabulated.

Point estimates and 2-sided 90% CIs for response rates will be summarized and overall response will be summarized by best overall response categories.

Time-to-event endpoints will be estimated using Kaplan-Meier methods, if appropriate. Additional details of the planned statistical analyses will be included in the SAP.

##### 12.6.8. Exploratory Analyses

Details on evaluation of exploratory analyses, including evaluation of preliminary clinical activity and possible relationships with PK/PD and mutational status and gene and protein expression levels, will be described in a separate analysis plan.

#### 12.7. Procedures for Reporting Deviations to Original Statistical Analysis Plan

All deviations from the original SAP will be provided in the final CSR.

### 13. ADMINISTRATIVE REQUIREMENTS

#### 13.1. Good Clinical Practice

The study will be conducted in accordance with the International Council for Harmonisation (ICH) guideline for Good Clinical Practice (GCP) and the appropriate regulatory requirement(s). The Investigator will be thoroughly familiar with the appropriate use of the study drug as described in the protocol and IB. Essential clinical documents will be maintained to demonstrate the validity of the study and the integrity of the data collected. Master files should be established at the beginning of the study, maintained for the duration of the study, and retained according to the appropriate regulations.

#### 13.2. Ethical Considerations

The study will be conducted in accordance with ethical principles founded in the Declaration of Helsinki.

The Investigator must obtain IRB/IEC approval for the investigation and must submit written documentation of the approval to the Sponsor before they can enroll any subject into the study. The IRB/IEC will review all appropriate study documentation in order to safeguard the rights, safety, and well-being of the subjects. The study will only be conducted at sites where IRB/IEC approval has been obtained. The protocol, IB, informed consent/assent, advertisements (if applicable), written information given to the subjects (including diary cards), safety updates, annual progress reports, and any revisions to these documents will be provided to the IRB/IEC. The IRB/IEC is to be notified of any amendment to the protocol in accordance with local requirements. Progress reports and notifications of serious unexpected adverse drug reactions are to be provided to the IRB/IEC according to local regulations and guidelines.

#### 13.3. Subject Information and Informed Consent

The Investigator at each center will ensure that the subject is given full and adequate oral and written information about the nature, purpose, possible risk, and benefit of the study. Subjects must also be notified that they are free to discontinue from the study at any time. The subject should be given the opportunity to ask questions and allowed time to consider the information provided.

After the study has been fully explained, written informed consent will be obtained from either the subject or their guardian or legal representative prior to study participation. Written informed assent will be obtained from subjects <18 years of age.

The subject's signed and dated informed consent/assent must be obtained before conducting any study-related procedures. The Investigator must maintain the original, signed consent/assent form. A copy of the signed form must be given to the subject.

The method of obtaining and documenting the informed consent/assent and the contents of the consent/assent will comply with ICH GCP and all applicable regulatory requirement(s).

#### 13.4. Subject Confidentiality

In order to maintain subject privacy, all source documents/eCRFs, study drug accountability records, study reports and communications will identify the subject by initials and the assigned subject number. The Investigator will grant monitor(s) and auditor(s) from the Sponsor or its designee and regulatory authority(ies) direct access to the subject's original medical records for verification of data gathered on the source documents/eCRFs and to audit the data collection process. The subject's confidentiality will be maintained and will not be made publicly available to the extent permitted by the applicable laws and regulations.

#### 13.5. Protocol Compliance

The Investigator will conduct the study in compliance with the protocol. Modifications to the protocol should not be made without agreement of both the Investigator and the Sponsor. Changes to the protocol will require written IRB/IEC approval/favorable opinion prior to implementation, except when the modification is needed to eliminate an immediate hazard(s) to subjects. The IRB/IEC may provide, if applicable, where regulatory authority(ies) permit, expedited review and approval/favorable opinion for minor change(s) in ongoing studies that have the approval/favorable opinion of the IRB/IEC. The Sponsor or designee will submit all protocol modifications to the regulatory authority(ies) in accordance with the governing regulations.

When immediate deviation from the protocol is required to eliminate an immediate hazard(s) to subjects, the Investigator will contact the Sponsor, if circumstances permit, to discuss the planned course of action. Any departures from the protocol must be fully documented in the source documents/eCRF.

#### 13.6. Independent Safety Monitoring Committee

##### 13.6.1. Part III: Combination Therapy

An independent safety monitoring committee will review cumulative safety data on a regular basis and will adjudicate cases of differentiation syndrome as specified in the committee charter. Additionally, the committee may review subject-level data on an ad hoc basis (eg, in the case of an event that may meet the criteria for pausing study enrollment [Section 9.8]), or as otherwise specified and described in the committee charter.

#### 13.7. Data Management

All data for the subjects recruited for the study, including clinically relevant unscheduled data, will be entered into the eCRFs via an Electronic Data Capture system provided by the Sponsor or designee. Only authorized staff may enter data onto the eCRFs. If an entry error is made, the corrections to the eCRFs will be made according to eCRF guidelines by an authorized member of the site staff.

eCRFs will be checked for correctness against source document data by the Sponsor's monitor. If any entries into the eCRF are incorrect or incomplete, the monitor will ask the Investigator or the study site staff to make appropriate corrections, and the corrected eCRF will again be reviewed for completeness and consistency. Any discrepancies will be noted in the eCRF system by means of electronic data queries. Authorized site staff will be asked to respond to all electronic queries according to the eCRF guidelines.

#### 13.8. Source Document/Case Report Form Completion

Source documents/eCRFs will be completed for each study subject. It is the Investigator's responsibility to ensure the accuracy, completeness, and timeliness of the data reported in the subject's source document/eCRF. The source document/eCRF should indicate the subject's participation in the study and should document the dates and details of study procedures, AEs, and subject status.

The Investigator, or designated representative, should complete the source document/eCRF as soon as possible after information is collected, preferably on the same day that a study subject is seen for an examination, treatment, or any other study procedure. Any outstanding entries must be completed immediately after the final examination. An explanation should be given for all missing data.

The Investigator must sign and date the Investigator's Statement at the end of the source document/eCRF to endorse the recorded data.

The Investigator will retain all completed source documents.

#### 13.9. Direct Access to Source Data

The study will be monitored by the Sponsor or its designee. Monitoring will be done by personal and/or remote visits from a representative of the Sponsor (site monitor) and will include on-site and/or electronic review of the source documents/eCRFs for completeness and clarity, cross-checking with source documents, and clarification of administrative matters will be performed. The review of medical records will be performed in a manner to ensure that subject confidentiality is maintained.

The site monitor will ensure that the investigation is conducted according to protocol design and regulatory requirements by frequent communications (letter, telephone, e-mail, and fax).

All unused study drug and other study materials are to be returned to the Sponsor or designee and/or appropriately destroyed/disposed of after the study has been completed.

Regulatory authorities, the IEC/IRB, and/or the Sponsor's clinical quality assurance group or designee may request access to all source documents, eCRFs, and other study documentation for an on-site audit or inspection. Direct access to these documents must be guaranteed by the Investigator, who must provide support at all times for these activities.

#### 13.10. Record Retention

The Investigator will maintain all study records according to ICH-GCP and applicable regulatory requirement(s) in force at time of trial completion. ICH-GCP requires that records will be retained for at least 2 years after the last marketing application approval or 2 years after formal discontinuation of the clinical development of the investigational product The EU Clinical Trial Regulation (EU No 536/2014), requires clinical data will be stored at least 25 years after the end of the clinical trial unless other national law requires archiving for a longer period.

If the Investigator withdraws from the responsibility of keeping the study records, custody must be transferred to a person willing to accept the responsibility. The Sponsor must be notified in writing if a custodial change occurs.

#### 13.11. Liability and Insurance

The Sponsor has subscribed to an insurance policy covering, in its terms and provisions, its legal liability for injuries caused to participating persons and arising out of this research performed strictly in accordance with the scientific protocol as well as with applicable law and professional standards.

#### 13.12. Publication of Study Findings and Use of Information

All information regarding FHD-286 supplied by the Sponsor or designee to the Investigator is privileged and confidential information. The Investigator agrees to use this information to accomplish the study and will not use it for other purposes without consent from the Sponsor. It is understood that there is an obligation to provide the Sponsor with complete data obtained during the study. The information obtained from the clinical study will be used towards the development of FHD-286 and may be disclosed to regulatory authority(ies), other Investigators, corporate partners, or consultants as required.

Cytarabine Injection. Package Insert. Hospira; December 2022.

Dacogen 50 mg powder for concentrate for solution for infusion. Summary of Product Characteristics. Janssen-Cilag Ltd; February 2022.

Dacogen (decitabine) for injection for intravenous use. Package Insert. Otsuka US; June 2020.

DepMap. DepMap 19Q2 Public. Broad Institute. Accessed 11 November 2020. 10.6084/m9.figshare.8061398.v1

Döhner H, Wei AH, Appelbaum FR, et al. Diagnosis and management of AML in adults: 2022 recommendations from an international expert panel on behalf of the ELN. *Blood.* 2022;140(12):1345-1377. doi: 10.1182/blood.2022016867

Fathi AT, Stein EM, DiNardo CD, Levis MJ, Montesinos P, de Botton S. Differentiation syndrome with lower-intensity treatments for acute myeloid leukemia. *Am J Hematol.* 2021;96(6):735-746. doi: 10.1002/ajh.26142

FDA. Guidance for Industry: In Vitro Drug Interaction Studies - Cytochrome P450 Enzyme- and Transporter-Mediated Drug Intearctions. U.S. Department of Health and Human Services, Food and Drug Administration, Center for Drug Evaluation and Research (CDER); January 2020.

Hariparsad N, Ramsden D, Palamanda J, et al. Considerations from the IQ Induction Working Group in response to drug-drug interaction guidance from regulatory agencies: focus on downregulation, CYP2C induction, and CYP2B6 positive control. *Drug Metab Dispos.* 2017;45(10):1049-1059. doi: 10.1124/dmd.116.074567

Ho L, Crabtree GR. Chromatin remodelling during development. *Nature.* 2010;463(7280):474-484. doi: 10.1038/nature08911

Hodges C, Kirkland JG, Crabtree GR. The many roles of BAF (mSWI/SNF) and PBAF complexes in cancer. *Cold Spring Harb Perspect Med.* 2016;6(8). doi: 10.1101/cshperspect.a026930

Kadoch C, Hargreaves DC, Hodges C, et al. Proteomic and bioinformatic analysis of mammalian SWI/SNF complexes identifies extensive roles in human malignancy. *Nat Genet.* 2013;45(6):592-601. doi: 10.1038/ng.2628

Kantarjian H, Kadia T, DiNardo C, et al. Acute myeloid leukemia: current progress and future directions. *Blood Cancer Journal.* 2021;11(2):41. doi: 10.1038/s41408-021-00425-3

Kantarjian HM, Thomas XG, Dmoszynska A, et al. Multicenter, randomized, open-label, phase III trial of decitabine versus patient choice, with physician advice, of either supportive care or low-dose cytarabine for the treatment of older patients with newly diagnosed acute myeloid leukemia. *J Clin Oncol.* 2012;30(21):2670-2677. doi: 10.1200/jco.2011.38.9429

Kayser S, Levis MJ. Updates on targeted therapies for acute myeloid leukaemia. *British Journal of Haematology.* 2022;196(2):316-328. doi: 10.1111/bjh.17746

Lai C, Doucette K, Norsworthy K. Recent drug approvals for acute myeloid leukemia. *J Hematol Oncol.* 2019;12(1):100. doi: 10.1186/s13045-019-0774-x

Lee SH, Erber WN, Porwit A, Tomonaga M, Peterson LC, International Council for Standardization In H. ICSH guidelines for the standardization of bone marrow specimens and reports. *Int J Lab Hematol.* 2008;30(5):349-364. doi: 10.1111/j.1751-553X.2008.01100.x

Lessard JA, Crabtree GR. Chromatin regulatory mechanisms in pluripotency. *Annu Rev Cell Dev Biol.* 2010;26:503-532. doi: 10.1146/annurev-cellbio-051809-102012

McKenzie MD, Ghisi M, Oxley EP, et al. Interconversion between tumorigenic and differentiated states in acute myeloid leukemia. *Cell Stem Cell.* 2019;25(2):258-272.e259. doi: 10.1016/j.stem.2019.07.001

Meyers RM, Bryan JG, McFarland JM, et al. Computational correction of copy number effect improves specificity of CRISPR-Cas9 essentiality screens in cancer cells. *Nat Genet.* 2017;49(12):1779-1784. doi: 10.1038/ng.3984

Montesinos P, Bergua JM, Vellenga E, et al. Differentiation syndrome in patients with acute promyelocytic leukemia treated with all-trans retinoic acid and anthracycline chemotherapy: characteristics, outcome, and prognostic factors. *Blood.* 2009;113(4):775-783. doi: 10.1182/blood-2008-07-168617

Muthuswami R, Bailey L, Rakesh R, Imbalzano AN, Nickerson JA, Hockensmith JW. BRG1 is a prognostic indicator and a potential therapeutic target for prostate cancer. *J Cell Physiol.* 2019. doi: 10.1002/jcp.28161

NCCN. NCCN Clinical Practice Guidelines in Oncology (NCCN Guidelines), Myelodysplastic Syndromes. National Comprehensive Cancer Network; September 2022.

NCCN. NCCN Clinical Practice Guidelines in Oncology (NCCN Guidelines), Acute Myeloid Leukemia. Version 1.2023. National Comprehensive Cancer Network; March 2023.

Norsworthy KJ, Mulkey F, Scott EC, et al. Differentiation syndrome with ivosidenib and enasidenib treatment in patients with relapsed or refractory IDH-mutated AML: A U.S. Food and Drug Administration systematic analysis. *Clin Cancer Res.* 2020;26(16):4280-4288. doi: 10.1158/1078-0432.Ccr-20-0834

Oken MM, Creech RH, Tormey DC, et al. Toxicity and response criteria of the Eastern Cooperative Oncology Group. *Am J Clin Oncol.* 1982;5(6):649-655

Onida F, Barosi G, Leone G, et al. Management recommendations for chronic myelomonocytic leukemia: consensus statements from the SIE, SIES, GITMO groups. *Haematologica.* 2013;98(9):1344-1352. doi: 10.3324/haematol.2013.084020

Padron E, Tinsley-Vance SM, Dezern A, et al. Efficacy and safety of ruxolitinib for treatmetn of symptomatic chronic myelomonocytic leukemia (CMML): results of a multicenter Phase II clinical trial. Oral presentation presented at 64th American Society of Hematology (ASH) Annual Meeting and Exposition; 10-13 Dec 2022; New Orleans, LA.

Rago F, Elliott G, Li A, et al. The discovery of SWI/SNF chromatin remodeling activity as a novel and targetable dependency in uveal melanoma. *Mol Cancer Ther.* 2020;19(10):2186-2195. doi: 10.1158/1535-7163.MCT-19-1013

SEER. Chronic myelomonocytic leukemia. National Cancer Institute Surveillance, Epidemiology, and End Results (SEER) Program. Accessed 06 March 2023. https://seer.cancer.gov/seertools/hemelymph/51f6cf57e3e27c3994bd5396/

SEER. Cancer Stat Facts: Leukemia — Acute Myeloid Leukemia (AML). National Cancer Institute Surveillance, Epidemiology, and End Results (SEER) Program. Accessed 11 November 2020. https://seer.cancer.gov/statfacts/html/amyl.html

Shi J, Whyte WA, Zepeda-Mendoza CJ, et al. Role of SWI/SNF in acute leukemia maintenance and enhancer-mediated Myc regulation. *Genes Dev.* 2013;27(24):2648-2662. doi: 10.1101/gad.232710.113

Stahl M, Tallman MS. Differentiation syndrome in acute promyelocytic leukaemia. *British Journal of Haematology.* 2019;187(2):157-162. doi: 10.1111/bjh.16151

Tsherniak A, Vazquez F, Montgomery PG, et al. Defining a cancer dependency map. *Cell.* 2017;170(3):564-576 e516. doi: 10.1016/j.cell.2017.06.010

Venclexta (venetoclax tablets) for oral use. Package Insert. AbbVie; June 2022.

Wu Q, Madany P, Dobson JR, et al. The BRG1 chromatin remodeling enzyme links cancer cell metabolism and proliferation. *Oncotarget.* 2016;7(25):38270-38281. doi: 10.18632/oncotarget.9505

### 15. APPENDICES

#### 15.1. National Cancer Institute Common Terminology Criteria for Adverse Events Version 5.0

The NCI CTCAE, version 5.0, can be accessed using the following link:

https://ctep.cancer.gov/protocolDevelopment/electronic_applications/docs/CTCAE_v5_Quick_Reference_8.5x11.pdf

#### 15.2. Schedules of Assessments for 1-Week-On/1-Week Off Dosing Regimens (Parts I and II [FHD-286 Monotherapy] *[Closed to Enrollment]* Only)

The following tables present the schedules of assessments to be used when FHD-286 is administered according to an alternative dosing regimen (eg, intermittent dosing).

- One week of dosing followed by 1 week without dosing (1-week-on/1-week-off regimen)
  - Dosing regimen: Table 20
  - Schedule of Assessments: Table 21
  - Timing of safety, PK, PD, and translational/exploratory sampling and ECGs: Table 22

**Table 20: Parts I and II *(Closed to Enrollment)*: FHD-286 Monotherapy 1-Week-On/1-Week-Off Dosing Regimen**

| Cycle | Week | Days | Dosing |
|---|---|---|---|
| All cycles | Week 1 | Days 1-7 | Dosing |
| | Week 2 | Days 8-14 | No dosing |
| | Week 3 | Days 15-21 | Dosing |
| | Week 4 | Days 22-28 | No dosing |

**Table 21: Parts I and II *(Closed to Enrollment)*: FHD-286 Monotherapy 1-Week-On/1-Week-Off Dosing Regimen: Schedule of Assessments**

| Visit/Cycle | Screening[a] | Cycle 1, 2, & 3 | | | | Cycle 4+ | | EOT Visit[b] | Safety Follow-up | Long-term Follow-up[c] |
|---|---|---|---|---|---|---|---|---|---|---|
| Study Day | | 1[d] | 3 or 4 (Cycle 1 only) | 7 (Cycles 1 & 3) or 8 (Cycle 2) | 15 | 1 | 15[e] | | +28[f] | |
| Window (days) | | NA (Cycle 1) or +2 (Cycles 2 and 3) | NA | −2 (Cycles 1 & 3) or ±2 (Cycle 2) | ±2 | +2 | ±2 | +5 | ±7 | ±14 |
| Informed consent/assent | X | | | | | | | | | |
| Study eligibility | X | | | | | | | | | |
| Demographics | X | | | | | | | | | |
| Medical & surgical history | X | | | | | | | | | |
| Physical examination[g] | X | X | | X | X | X | X | X | X | |
| Vital signs | X | X | | X | X | X | X | X | X | |
| ECOG PS | X | X | | X | X | X | X | X | X | |
| Clinical laboratory tests (local)[h] | See Table 22. | | | | | | | | | |
| ECHO (or other means) for LVEF | X | X (either C3D1 or C4D1 [±3 days]) | | | | | | X | | |
| Electrocardiogram[i] | See Table 22. | | | | | | | | | |
| PK/PD assessments[j] | See Table 22. | | | | | | | | | |
| Bone marrow biopsy and/or aspirate[k] | X | Starting on C2D1, bone marrow biopsy and/or aspirate will be collected approximately every 4 weeks for the first 24 weeks, then approximately every 8 weeks for the next 48 weeks of treatment, and as clinically indicated thereafter | | | | | | X | | |

**Table 21: Parts I and II *(Closed to Enrollment)*: FHD-286 Monotherapy 1-Week-On/1-Week-Off Dosing Regimen: Schedule of Assessments**

| Visit/Cycle | Screening[a] | Cycle 1, 2, & 3 | | | | Cycle 4+ | | EOT Visit[b] | Safety Follow-up | Long-term Follow-up[c] |
|---|---|---|---|---|---|---|---|---|---|---|
| Study Day | | 1[d] | 3 or 4 (Cycle 1 only) | 7 (Cycles 1 & 3) or 8 (Cycle 2) | 15 | 1 | 15[e] | | +28[f] | |
| Window (days) | | NA (Cycle 1) or +2 (Cycles 2 and 3) | NA | −2 (Cycles 1 & 3) or ±2 (Cycle 2) | ±2 | +2 | ±2 | +5 | ±7 | ±14 |
| Evaluate extent of disease and response to treatment[l] | | Evaluation will occur in accordance with bone marrow biopsy and/or aspirate collection | | | | | | X | | |
| Hepatitis panel (A, B, C) | X | | | | | | | | | |
| Serum HIV | X | | | | | | | | | |
| Pregnancy testing[m] | X | X | | | | X | | | X | |
| Dispense study medication[n] | | X | | X | X | X | X | | | |
| Concomitant medications/ procedures | X | X | | X | X | X | X | X | X | |
| AE assessment | X | X | | X | X | X | X | X | X | |
| Survival status | | | | | | | | | X | X |

Abbreviations: AE = adverse event; C1D1 = Cycle 1, Day 1; C4D15 = Cycle 4, Day 15; ECG = electrocardiogram; ECHO = echocardiogram; ECOG = Eastern Cooperative Oncology Group; EOT = End of Treatment; hCG = human chorionic gonadotropin; HIV = human immunodeficiency virus; IWG = International Working Group; PD = pharmacodynamics; PK = pharmacokinetics; PS = Performance Status.

[a] Within 28 days prior to first dose of FHD-286 (C1D1).

[b] EOT Visit will take place within 5 days after treatment ends (treatment end is defined as the date on which the Investigator decided to discontinue FHD-286 treatment and is not required to be the date of the last FHD-286 dose). If a subject's dose is interrupted for 28 days and then the subject discontinues study participation, the EOT Visit will serve as the Safety Follow-up Visit.

[c] After subjects have either experienced documented disease progression/treatment failure or discontinued study treatment, whichever occurs later, they will be contacted by telephone approximately every 2 months to assess survival status, document receipt and type of subsequent anticancer therapy, and document disease status (until subject experiences disease progression/treatment failure or starts a new anticancer therapy).

**Table 21: Parts I and II *(Closed to Enrollment)*: FHD-286 Monotherapy 1-Week-On/1-Week-Off Dosing Regimen: Schedule of Assessments**

| Visit/Cycle | Screening[a] | Cycle 1, 2, & 3 | | | | Cycle 4+ | | EOT Visit[b] | Safety Follow-up | Long-term Follow-up[c] |
|---|---|---|---|---|---|---|---|---|---|---|
| Study Day | | 1[d] | 3 or 4 (Cycle 1 only) | 7 (Cycles 1 & 3) or 8 (Cycle 2) | 15 | 1 | 15[e] | | +28[f] | |
| Window (days) | | NA (Cycle 1) or +2 (Cycles 2 and 3) | NA | −2 (Cycles 1 & 3) or ±2 (Cycle 2) | ±2 | +2 | ±2 | +5 | ±7 | ±14 |

d Assessments on C1D1 will be performed predose, except for PK/PD samples and ECGs which will be collected according to Table 22.
e At C4D15 and beyond, samples for laboratory assessments will be collected on site; other assessments may be made via telemedicine.
f Safety Follow-up Visit will occur approximately 28 days after the EOT Visit.
g A complete physical examination, including height and weight, will be performed during Screening. Physical examinations may be abbreviated (ie, partial) at all other protocol-specified visits. Weight should be collected on Day 1 of each cycle.
h Clinical laboratory tests include hematology, serum chemistry, and coagulation studies. Samples for analysis will be collected as specified in Table 22. Specific analytes are outlined in Table 14.
I 12-lead ECGs will be collected in triplicate prior to each PK time point as described in Table 22 (starting after approximately 3 minutes of recumbency or semi-recumbency, with each ECG performed approximately 2 minutes apart). There is no associated PK sample collection for the Screening ECG; as such, the 12-lead Screening ECG may be performed in triplicate at any time during the Screening window.
j PK/PD and translational/exploratory assessments will be collected as specified in Table 22. Drug should be administered on site on days that PK/PD assessments will take place.
k A bone marrow biopsy and/or aspirate should be conducted if progression of disease is suspected. If additional bone marrow biopsies and/or aspirates are performed during the study at the Investigator's discretion, study samples will be collected.
l Disease response will be based upon modified IWG criteria as detailed in Section 10.6.
m Female subjects of reproductive potential only. Serum hCG testing is to be conducted at the Screening Visit. Serum or urine hCG testing will be conducted predose on Day 1 of every cycle.
n Subjects will be dispensed the appropriate number of Sponsor-packaged, labeled bottles to allow for dosing for a full cycle or until the next scheduled visit.

Note: All doses of FHD-286 will be administered under fasted conditions as described in Section 9.5.1.1.

**Table 22: Parts I and II *(Closed to Enrollment)*: FHD-286 Monotherapy 1-Week-On/1-Week-Off Dosing Regimen: Timing of Safety, Pharmacokinetic, Pharmacodynamic, and Exploratory Sampling and Electrocardiograms**

| Cycle | Study Day | Hour[a] | Safety Blood | PK Blood | PD Blood | Exploratory Blood | ECG[b] |
|---|---|---|---|---|---|---|---|
| Screen-ing | -28[c] | | X | | | X | X |
| 1 | 1 | 0 | X | X | X | | X |
| | | 0.5 | | X | | | |
| | | 1 | | X | | | |
| | | 2 | | X | | | X |
| | | 4 | | X | X | | X |
| | | 6 | | X | | | X |
| | | 8 | | X | X | | X |
| | 2 | 0 | | X | X | | X |
| | 3 or 4 | | X[d] | | | | |
| | 7 | 0 | X | X | X | X | X |
| | | 0.5 | | X | | | |
| | | 1 | | X | | | |
| | | 2 | | X | | | X |
| | | 4 | | X | X | | X |
| | | 6 | | X | | | X |
| | | 8 | | X | X | | X |
| | 8 | 0 | | X | X | | X |
| | 15 | 0 | X | X | | | |
| 2 | 1 | 0 | X | X | X | | X |
| | | 2 | | X | | | X |
| | 8 | 0 | X | X | | | |
| | 15 | 0 | X | X | | | |
| 3 | 1 | 0 | X | X | | | X |
| | | 4 | | X | | | X |
| | | 8 | | X | | | |
| | 7 | 0 | X | | X | X | |
| | | 4 | | | X | | |
| | | 8 | | | X | | |
| | 15 | 0 | X | X | | | |

**Table 22: Parts I and II *(Closed to Enrollment)*: FHD-286 Monotherapy 1-Week-On/1-Week-Off Dosing Regimen: Timing of Safety, Pharmacokinetic, Pharmacodynamic, and Exploratory Sampling and Electrocardiograms**

| Cycle | Study Day | Hour[a] | Safety Blood | PK Blood | PD Blood | Exploratory Blood | ECG[b] |
|---|---|---|---|---|---|---|---|
| 4+ | 1 | 0 | X | X | X[e] | X[e] | |
| EOT[f] | | | X | X | X | X | |

Abbreviations: ECG = electrocardiogram; EOT = End of Treatment; PD = pharmacodynamics; PK = pharmacokinetics.
[a] Hour 0 samples to be collected within 30 minutes before dose (or, on non-dosing days, approximately 24 hours after the previous day's dose). Samples at all other hours to be collected within ±10 minutes of specified time. ECGs should be collected within approximately 10 minutes prior to PK sample collection.
[b] 12-lead ECGs will be collected in triplicate prior to PK sample collection (starting after approximately 3 minutes of recumbency or semi-recumbency; each ECG performed approximately 2 minutes apart). There is no associated PK sample collection for the Screening ECG; as such, the 12-lead Screening ECG may be performed in triplicate at any time during the Screening window.
[c] Within 28 days prior to first dose of FHD-286 (C1D1).
[d] In Cycle 1 only, serum chemistry will be collected on either Day 3 or Day 4.
[e] After Cycle 3, PD and translational/exploratory blood will be collected on D7 of every odd-numbered cycle (Cycle 5, 7, etc.) for the first 48 weeks.
[f] EOT Visit will take place within 5 days after treatment ends (treatment end is defined as the date on which the Investigator decided to discontinue FHD-286 treatment and is not required to be equal to the date of the last FHD-286 dose).

Notes:
1. Drug should be administered on site on days that PK and/or PD assessments will take place.
2. Samples for PK may also be used for other exploratory analysis, including but not limited to metabolite profiling and additional biomarker analysis.
3. Subjects may be requested to undergo unscheduled PK/PD/safety assessments and ECGs if a safety event occurs.

#### 15.3. New York Heart Association Classification

| Class | Symptomatology |
|---|---|
| I | No limitation of physical activity. Ordinary physical activity does not cause undue fatigue, palpitation, dyspnea, or anginal pain. |
| II | Slight limitation of physical activity. Comfortable at rest, but ordinary physical activity results in fatigue, palpitation, dyspnea, or anginal pain. |
| III | Marked limitation of physical activity. Comfortable at rest, but less than ordinary activity results in fatigue, palpitation, dyspnea, or anginal pain. |
| IV | Unable to carry on any physical activity without discomfort. Symptoms at rest. If any physical activity is undertaken, discomfort is increased. |

Source: The Criteria Committee of the New York Heart Association. Nomenclature and Criteria for Diagnosis of Diseases of the Heart and Great Vessels. 9th ed. Boston, Mass: Little, Brown & Co; 1994:253-256.

#### 15.4. Cytochrome P450 3A Substrates, Inhibitors, and Inducers

The tables in this appendix include examples from the Food and Drug Administration (FDA). An additional comprehensive resource is the Indiana University Drug Interactions Flockhart Table, accessible at https://drug-interactions.medicine.iu.edu/MainTable.aspx. If any questions arise, discuss with the Sponsor.

| CYP Enzyme | Sensitive Substrates |
|---|---|
| CYP3A | alfentanil, avanafil, budesonide, buspirone, conivaptan, darifenacin, darunavir(a), dasatinib, dronedarone, ebastine, eletriptan, eplerenone, everolimus, felodipine, ibrutinib, indinavir(a), lomitapide, lovastatin(b), lurasidone, midazolam, maraviroc, naloxegol, nisoldipine, quetiapine, saquinavir(a), sildenafil, simvastatin(b), sirolimus, tacrolimus, ticagrelor, tipranavir(a), tolvaptan, triazolam, vardenafil |

Source: FDA DDI Website: Table 3-1. Examples of Clinical Substrates for P450-mediated Metabolism (12/03/2019).
Abbreviations: AUC = area under the concentration-time curve; CYP = cytochrome P450; DDI = drug-drug interaction; EM = extensive metabolizer; FDA = Food and Drug Administration; OATP1B1 = organic anion transporting polypeptide 1B1.
Note: For an updated list, see the following link: https://www.fda.gov/drugs/drug-interactions-labeling/drug-development-and-drug-interactions-table-substrates-inhibitors-and-inducers.
Note from the FDA source: Sensitive substrates are drugs that demonstrate an increase in AUC of ≥5-fold with strong index inhibitors of a given metabolic pathway in clinical DDI studies. Sensitive substrates of CYP3A with ≥10-fold increase in AUC by co-administration of strong index inhibitors are shown above the dashed line. Other elimination pathways may also contribute to the elimination of the substrates listed in the table above and should be considered when assessing the drug interaction potential.
Note from the FDA source: This table is prepared to provide examples of clinical substrates and not intended to be an exhaustive list. DDI data were collected based on a search of the University of Washington Metabolism and Transport Drug Interaction Database [Hachad et al. (2010), Hum Genomics, 5(1):61].
[a] Usually administered to patients in combination with ritonavir, a strong CYP3A inhibitor.
[b] Acid form is an OATP1B1 substrate.

| CYP Enzyme | Strong Inhibitors | Moderate Inhibitors |
|---|---|---|
| CYP3A4 | boceprevir, clarithromycin[(c)], cobicistat[(c)], danoprevir and ritonavir[(d)], elvitegravir and ritonavir[(d)], grapefruit juice[(e)], idelalisib, indinavir and ritonavir[(d)], itraconazole[(c)], ketoconazole, lopinavir and ritonavir[(c,d)], nefazodone, nelfinavir[(c)], paritaprevir and ritonavir and (ombitasvir and/or dasabuvir)[(d)], posaconazole, ritonavir[(c,d)], saquinavir and ritonavir[(c,d)], telaprevir[(c)], tipranavir and ritonavir[(c,d)], telithromycin, troleandomycin, voriconazole | aprepitant, ciprofloxacin, conivaptan[(f)], crizotinib, cyclosporine, diltiazem[(g)], dronedarone[(c)], erythromycin, fluconazole[(b)], fluvoxamine[(a)], imatinib, tofisopam, verapamil[(c)] |

Source: FDA DDI Website: Table 3-2. Examples of Clinical Inhibitors for P450-mediated Metabolism (03/06/2020).
Abbreviations: AUC = area under the concentration-time curve; CYP = cytochrome P450; DDI = drug-drug interaction; FDA = Food and Drug Administration; HIV = human immunodeficiency virus; HCV = hepatitis C virus; P-gp = permeability glycoprotein.
Note: For an updated list, see the following link: https://www.fda.gov/drugs/drug-interactions-labeling/drug-development-and-drug-interactions-table-substrates-inhibitors-and-inducers.
Note from the FDA source: Strong, moderate, and weak inhibitors are drugs that increase the AUC of sensitive index substrates of a given metabolic pathway ≥5-fold, ≥2 to <5-fold, and ≥1.25 to <2-fold, respectively. Strong inhibitors of CYP3A causing ≥10-fold increase in AUC of sensitive index substrate(s) are shown above the dashed line.
Note from the FDA source: This table is prepared to provide examples of clinical inhibitors and is not intended to be an exhaustive list. DDI data were collected based on a search of the University of Washington Metabolism and Transport Drug Interaction Database [Hachad et al. (2010), Hum Genomics, 5(1):61].
[a] Strong inhibitor of CYP1A2 and CYP2C19. Moderate inhibitor of CYP3A and weak inhibitor of CYP2D6.
[b] Strong inhibitor of CYP2C19 and moderate inhibitor of CYP2C9 and CYP3A.
[c] Inhibitor of P-gp (defined as those increasing AUC of digoxin to ≥1.25-fold).
[d] Ritonavir is usually given in combination with other anti-HIV or anti-HCV drugs in clinical practice. Caution should be used when extrapolating the observed effect of ritonavir alone to the effect of combination regimens on CYP3A activities.
[e] The effect of grapefruit juice varies widely among brands and is concentration-, dose-, and preparation-dependent. Studies have shown that it can be classified as a “strong CYP3A inhibitor” when a certain preparation was used (eg, high dose, double strength) or as a “moderate CYP3A inhibitor” when another preparation was used (eg, low dose, single strength).
[f] The classification is based on studies conducted with intravenously administered conivaptan.
[g] Diltiazem increased AUC of certain sensitive CYP3A substrates (eg, buspirone) more than 5-fold.

| CYP Enzyme | Strong Inducers | Moderate Inducers |
|---|---|---|
| CYP3A4 | apalutamide, carbamazepine[(c)], enzalutamide[(e)], mitotane, phenytoin[(b)], rifampin[(a)], St. John's wort[(f)] | bosentan, efavirenz[(d)], etravirine, phenobarbital, primidone |

Source: FDA DDI Website: Table 3-3. Examples of Clinical Inducers for P450-mediated Metabolism (12/03/2019)
Abbreviations: AUC = area under the concentration-time curve; CYP = cytochrome P450; DDI = drug-drug interaction; FDA = Food and Drug Administration.
Note: For an updated list, see the following link: https://www.fda.gov/drugs/drug-interactions-labeling/drug-development-and-drug-interactions-table-substrates-inhibitors-and-inducers.
Note from the FDA source: Strong, moderate, and weak inducers are drugs that decreases the AUC of sensitive index substrates of a given metabolic pathway by ≥80%, ≥50% to <80%, and ≥20% to <50%, respectively.
Note from the FDA source: This table is prepared to provide examples of clinical index inducers and not intended to be an exhaustive list. DDI data were collected based on a search of the University of Washington Metabolism and Transport Drug Interaction Database [Hachad et al. (2010), Hum Genomics, 5(1):61].
[a] Strong inducer of CYP3A and moderate inducer of CYP1A2, CYP2C19.
[b] Strong inducer of CYP2C19, CYP3A, and moderate inducer of CYP1A2, CYP2B6, CYP2C8, CYP2C9.
[c] Strong inducer of CYP2B6, CYP3A, and weak inducer of CYP2C9.
[d] Moderate inducer of CYP2B6, CYP2C19, and CYP3A.
[e] Strong inducer of CYP3A and moderate inducer of CYP2C9, and CYP2C19.
[f] The effect of St. John's wort varies widely and is preparation dependent.

#### 15.5. Transporter-Sensitive Substrates

The table in this appendix includes examples from the FDA. If any questions arise, discuss with the Sponsor.

| Transporter | Gene | Substrate |
|---|---|---|
| P-gp | *ABCB1* | dabigatran etexilate, digoxin, fexofenadine |
| BCRP | *ABCG2* | rosuvastatin, sulfasalazine |
| MATE1 | *SLC47A1* | metformin |

Source: FDA DDI Website: Table 5-1. Examples of Clinical Substrates for Transporters (12/03/2019)
Abbreviations: AUC = area under the plasma concentration-time curve; BCRP = breast cancer resistance protein; DDI = drug-drug interaction; FDA = Food and Drug Administration; MATE1 = multidrug and toxin extrusion transporter 1; P-gp = permeability glycoprotein.
Note: For an updated list, see the following link: https://www.fda.gov/drugs/drug-interactions-labeling/drug-development-and-drug-interactions-table-substrates-inhibitors-and-inducers.
Note: Criteria for selecting clinical substrates are as follows:
P-gp: (1) AUC fold-increase ≥2 with verapamil or quinidine co-administration and (2) in vitro transport by P-gp expression systems, but not extensively metabolized.
BCRP: (1) AUC fold-increase ≥2 with pharmacogenetic alteration of ABCG2 (421C>A) and (2) in vitro transport by BCRP expression systems.
This table is prepared to provide examples of clinical substrates for various transporters and not intended to be an exhaustive list. DDI data were collected based on a search of the University of Washington Metabolism and Transport Drug Interaction Database [Hachad et al. (2010), Hum Genomics, 5(1):61].

#### 15.6. Clinical Inhibitors for Transporters

The table in this appendix includes examples from the FDA. If any questions arise, discuss with the Sponsor.

| Transporter | Gene | Inhibitor |
|---|---|---|
| P-gp | *ABCB1* | amiodarone, carvedilol, clarithromycin, dronedarone, itraconazole, lapatinib, lopinavir and ritonavir, propafenone, quinidine, ranolazine, ritonavir, saquinavir and ritonavir, telaprevir, tipranavir and ritonavir, verapamil |
| BCRP | *ABCG2* | curcumin, cyclosporine A, eltrombopag |

Source: FDA DDI Website: Table 5-2. Examples of Clinical Inhibitors for Transporters (09/26/2016)
Abbreviations: AUC = area under the plasma concentration-time curve; BCRP = breast cancer resistance protein; DDI = drug-drug interaction; FDA = Food and Drug Administration; P-gp = permeability glycoprotein.
Note: For an updated list, see the following link: https://www.fda.gov/drugs/drug-interactions-labeling/drug-development-and-drug-interactions-table-substrates-inhibitors-and-inducers.
Note: Criteria for selecting clinical substrates are as follows:
P-gp: (1) AUC fold-increase of digoxin ≥2 with co-administration and (2) in vitro inhibitor.
BCRP: (1) AUC fold-increase of sulfasalazine ≥1.5 with co-administration and (2) in vitro inhibitor. Cyclosporine A and eltrombopag were also included, although the available DDI information was with rosuvastatin, where inhibition of both BCRP and OATPs may have contributed to the observed interaction.
This table is prepared to provide examples of clinical inhibitors for various transporters and not intended to be an exhaustive list. DDI data were collected based on a search of the University of Washington Metabolism and Transport Drug Interaction Database [Hachad et al. (2010), Hum Genomics, 5(1):61].

#### 15.7. Eastern Cooperative Oncology Group Performance Status Scoring

| Grade | Symptomatology |
|---|---|
| 0 | Fully active, able to carry on all pre-disease performance without restriction |
| 1 | Restricted in physically strenuous activity but ambulatory and able to carry out work of a light or sedentary nature, eg, light housework, office work |
| 2 | Ambulatory and capable of all self-care but unable to carry out any work activities. Up and about more than 50% of waking hours |
| 3 | Capable of only limited self-care, confined to bed or chair more than 50% of waking hours |
| 4 | Completely disabled. Cannot carry on any self-care. Totally confined to bed or chair |
| 5 | Dead |

Source: Oken, et al (1982).

#### 15.8. Additional Guidance on Potential Signs and Symptoms of Differentiation Syndrome

The following criteria are provided as additional guidance in identifying potential signs and symptoms of differentiation syndrome.

| Category | Adverse event, lab, or vital sign abnormality |
|---|---|
| Dyspnea | Acute respiratory failure, cardiopulmonary failure, cardiorespiratory distress, cough, dyspnea, respiratory arrest, respiratory failure, respiratory distress |
| Fever | Pyrexia, febrile neutropenia, or temperature ≥38.3ºC |
| Weight gain | Capillary leak syndrome, fluid overload, fluid retention, generalized edema, hydremia, hypervolemia, oedema, oedema peripheral, or weight gain >5 kg from baseline |
| Hypotension | Hypotension or systolic blood pressure <90 mmHg |
| Acute renal failure | Acute kidney injury, anuria, cardiorenal syndrome, hepatorenal failure, prerenal failure, renal failure, renal failure acute, renal impairment, renal injury, or creatinine (µmol/L) >26.52 µmol/L above baseline or 1.5 × above baseline |
| Pulmonary infiltrates or pleuropericardial effusion | Acute pulmonary edema, acute respiratory distress syndrome, non-cardiogenic pulmonary edema, pulmonary congestion, pulmonary edema, pleural effusion, pericardial effusion, acute interstitial pneumonitis, acute lung injury, atypical pneumonia, lower respiratory tract infection, lower respiratory tract inflammation, lung infection, lung infiltration, pneumonia, pneumonitis, pulmonary toxicity |
| Multiple organ dysfunction | Multiple organ dysfunction syndrome or multi-organ failure |

Source: Montesinos, et al (2009); Norsworthy, et al (2020); Stahl and Tallman (2019).

#### 15.9. Drugs With a Risk of QT Interval Prolongation and/or Torsades de Pointes/Ventricular Fibrillation

A list of drugs with a risk of QT interval prolongation and/or torsades de pointes/ventricular fibrillation is provided below. An additional comprehensive resource is https://www.crediblemeds.org/.

| | | |
|---|---|---|
| Alfuzosin | Fexinidazole | Pentamidine |
| Amantadine | Fingolimod | Pimavanserin |
| Amiodarone | Flecainide | Pimozide |
| Amisulpride | Fluconazole | Pitolisant |
| Amitriptyline | Fluoxetine | Posaconazole |
| Apalutamide | Fostemsavir | Probucol |
| Aripiprazole | Galantamine | Procainamide |
| Arsenic trioxide | Gatifloxacin | Quetiapine |
| Asenapine | Glasdegib | Quinidine |
| Astemizole | Grepafloxacin | Ranolazine |

| | | |
|---|---|---|
| Atazanavir | Halofantrine | Remimazolam |
| Azithromycin | Haloperidol | Ribociclib |
| Bedaquiline | Ibogaine | Rilpivirine |
| Bepridil | Ibutilide | Risperidone |
| Berotralstat | Iloperidone | Roxithromycin |
| Betrixaban | Indapamide | Rucaparib |
| Cabozantinib | Ivosidenib | Saquinavir |
| Ceritinib | Ketoconazole | Selpercatinib |
| Chloroquine | Lapatinib | Sertindole |
| Chlorpheniramine | Lefamulin | Sibutramine |
| Chlorpromazine | Levofloxacin | Siponimod |
| Ciprofloxacin | Levomethadyl acetate | Sorafenib |
| Cisapride | Lithium | Sotalol |
| Citalopram | Lofexidine | Sparfloxacin |
| Clarithromycin | Loperamide | Sunitinib |
| Clomipramine | Lumateperone | Tacrolimus |
| Clozapine | Macimorelin | Tamoxifen |
| Crizotinib | Methadone | Telavancin |
| Dasatinib | Mifepristone | Telithromycin |
| Deutetrabenazine | Mobocertinib | Terfenadine |
| Disopyramide | Moxifloxacin | Tetrabenazine |
| Dofetilide | Nilotinib | Thioridazine |
| Dolasetron | Ofloxacin | Thioridazine 2-sulfoxide (mesoridazine) |
| Domperidone | Oliceridine | Trilaciclib |
| Dronedarone | Olodaterol | Valbenazine |
| Droperidol | Ondansetron | Vandetanib |
| Eliglustat | Osilodrostat | Vardenafil |
| Encorafenib | Osimertinib | Vemurafenib |
| Entrectinib | Pacritinib | Venlafaxine |
| Eribulin | Paliperidone (9-hydroxyrisperidone) | Voclosporin |
| Erythromycin | Panobinostat | Vonoprazan and amoxicillin and clarithromycin |
| Etelcalcetide | Pasireotide | Voriconazole |
| Ezogabine (retigabine) | Pazopanib | Ziprasidone |
