## Supplemental Materials for "Mechanism of response to FHD-286 and decitabine combination in patients with advanced myeloid malignancies"

**Figure S1. CONSORT diagram of combination portion of Study FHD-286-C-002.** Patients who were enrolled and received ≥1 dose of study treatment were analyzed for safety and clinical activity. Patients who received ≥1 dose of FHD-286 and had ≥1 blood sample providing evaluable pharmacokinetic data for FHD-286 were analyzed for pharmacokinetics.

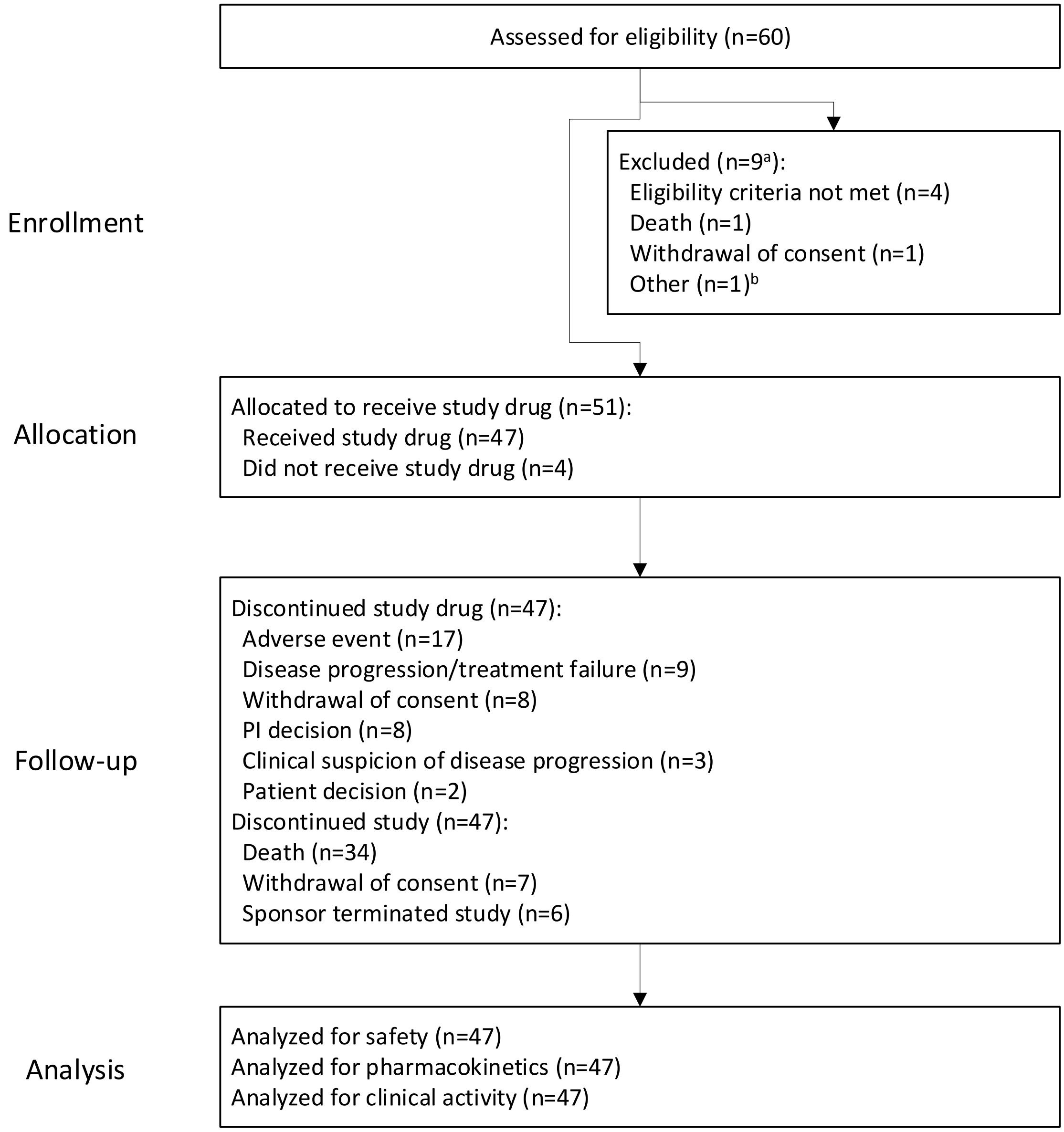


[a] Reasons for screen failure were available for 7 of the 9 subjects who failed screening.
[b] Physician and patient decided to keep patient on current therapy.

**Figure S2. Schema of Study FHD-286-C-002.** CYP, cytochrome P450; IV, intravenous; LDAC, low-dose cytarabine; QD, once daily; SC, subcutaneous.

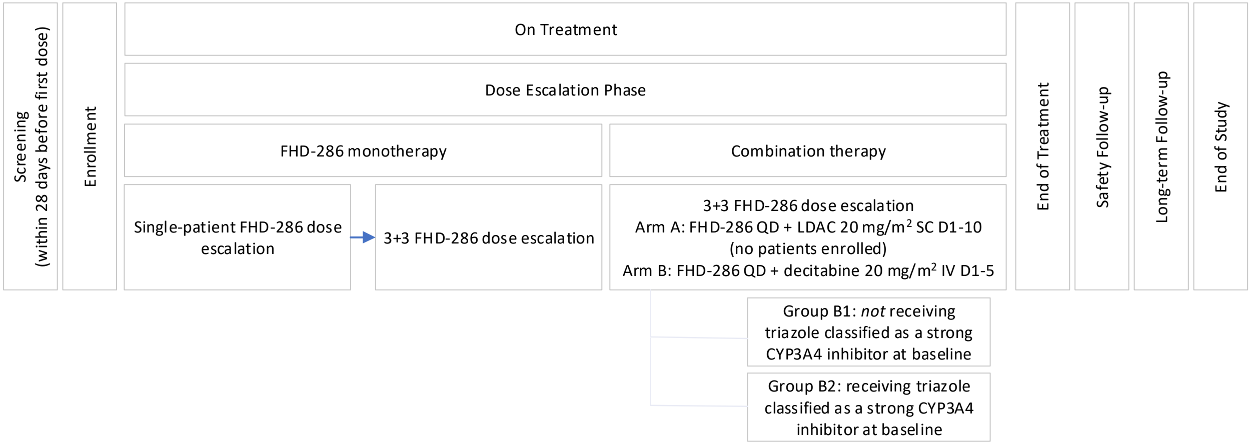

**Figure S3: Mean (SD) FHD-286 Plasma Concentration After Daily Oral Administration of FHD-286 2.5 mg in Combination With Decitabine on C1D1 and C1D15.** Group B1: Subjects *not* receiving a triazole antifungal agent classified as a strong CYP3A inhibitor at baseline. Group B2: Subjects receiving a triazole antifungal agent classified as a strong CYP3A inhibitor at baseline. CxDy, Cycle x Day y.

**C1D1** **C1D15**

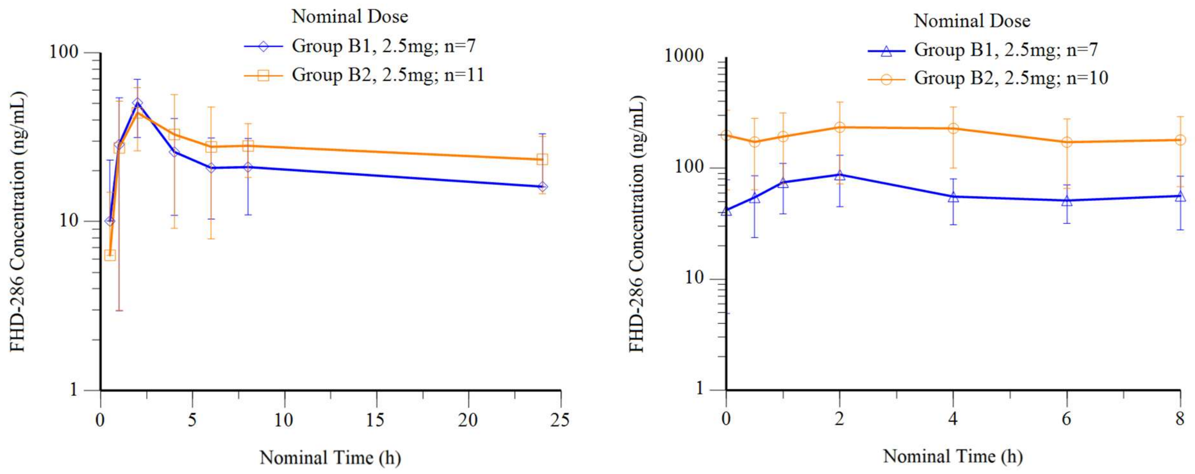

**Figure S4: Swim Lane Plots of Time on Treatment With FHD-286 and Decitabine in Study FHD-286-C-002.** All subjects had acute myeloid leukemia, except * (myelodysplastic syndrome) and ** (chronic myelomonocytic leukemia). B1: Subjects *not* receiving a triazole antifungal agent classified as a strong CYP3A inhibitor at the start of study treatment. B2: Subjects receiving a triazole antifungal agent classified as a strong CYP3A inhibitor at the start of study treatment. Responses per modified International Working Group criteria *(10, 11)*. Treatment failure includes resistant disease, death in aplasia, death from indeterminate cause, relapse, and disease progression. CRp, complete remission with incomplete platelet recovery; MLFS, morphologic leukemia-free state; PR, partial remission; QD, once daily; SD, stable disease; TF, treatment failure.

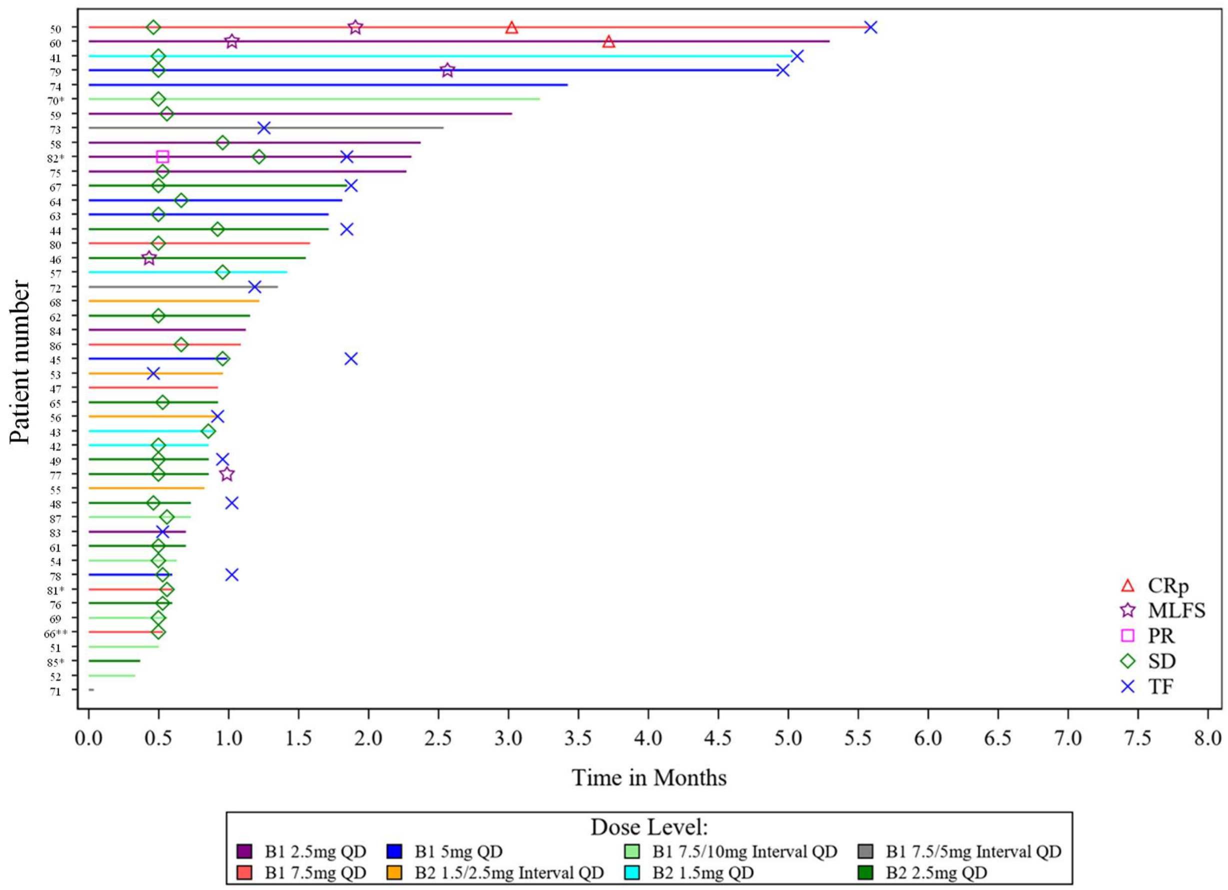

**Figure S5. Cell type identification using FACS.**
(**A**) t-SNE plots colored by scaled fluorescence intensity for the indicated markers.
(**B**) Box plots showing scaled median fluorescence intensity for indicated markers within each cluster. Each point represents one sample. Boxes represent interquartile range (IQR) with median indicated by center line, and whiskers extending to 1.5x IQR.

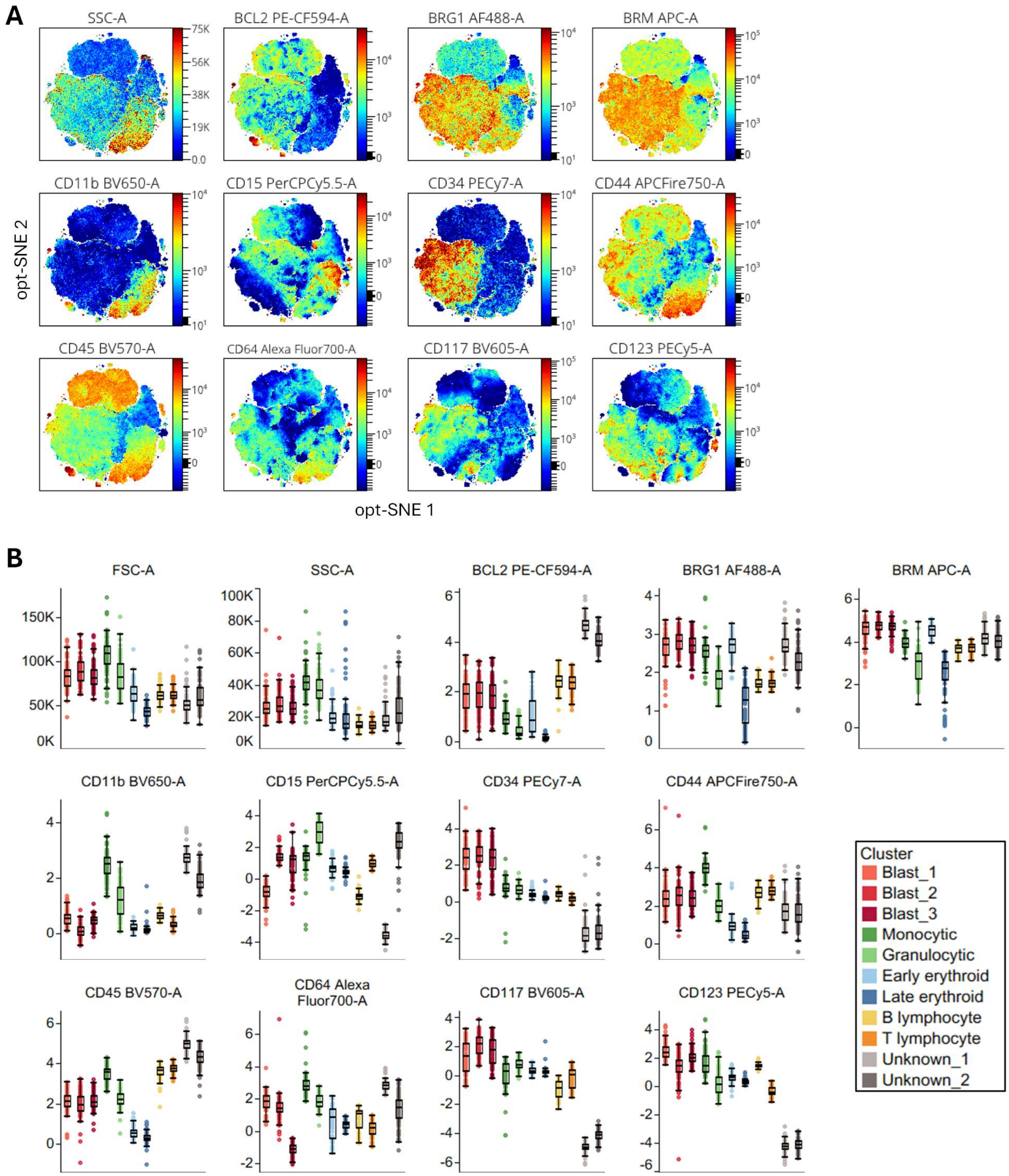

**Figure S6. Single cell genomics – dataset annotation and validation.**
(**A**) Dot plot displaying genes enriched within each annotated cell type cluster.
(**B**) Stacked bar plot showing relative proportion of each patient's contribution to individual cell type clusters. Multiple longitudinal samples from same patient were aggregated together.
(**C**) Stacked bar plot showing relative proportion of each cell type in patient samples. Multiple longitudinal samples from same patient were aggregated together.
(**D**) UMAP plots showing relatively uniform distribution of cells from combination and monotherapy trials.
(**E**) UMAP plots showing relatively uniform distribution of cells from samples collected at screening and after treatment.

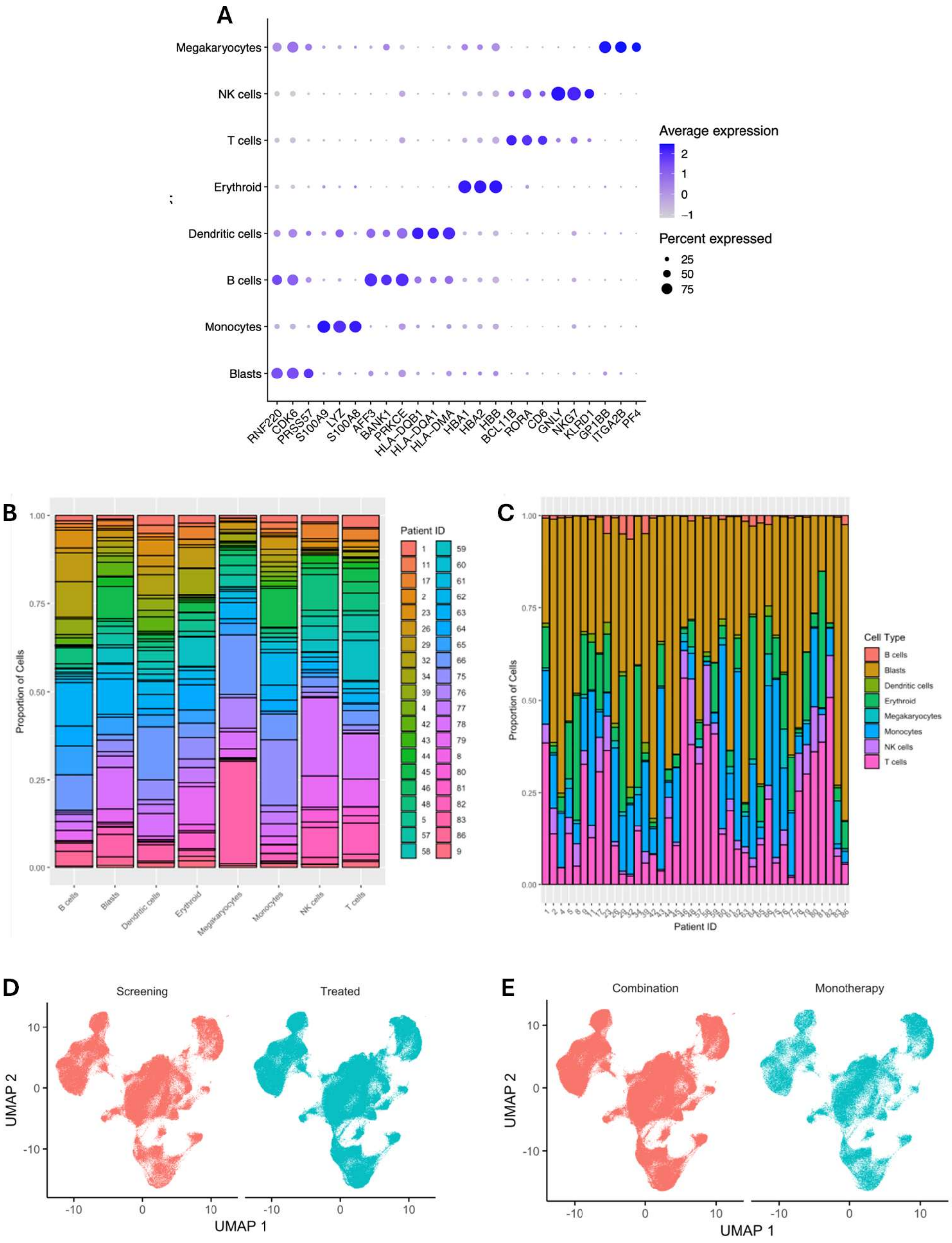

**Figure S7. Single cell genomics – blast compartment identification and validation.**
(**A**) Heatmap showing proportion of cells in each cell type (columns) that match each blast identification measure (rows).
(**B**) UMAP plot showing distribution of Ucell score from AML blast signature for each cell.
(**C**) UMAP plot showing distribution of cells labeled as "aneuploid" by CopyKAT algorithm.
(**D**) Scatter plot showing correlation between blast percentages determined morphologically at clinical sites (x-axis) and proportion of blast cells measured by scRNA-seq (y-axis) in individual BMA samples.

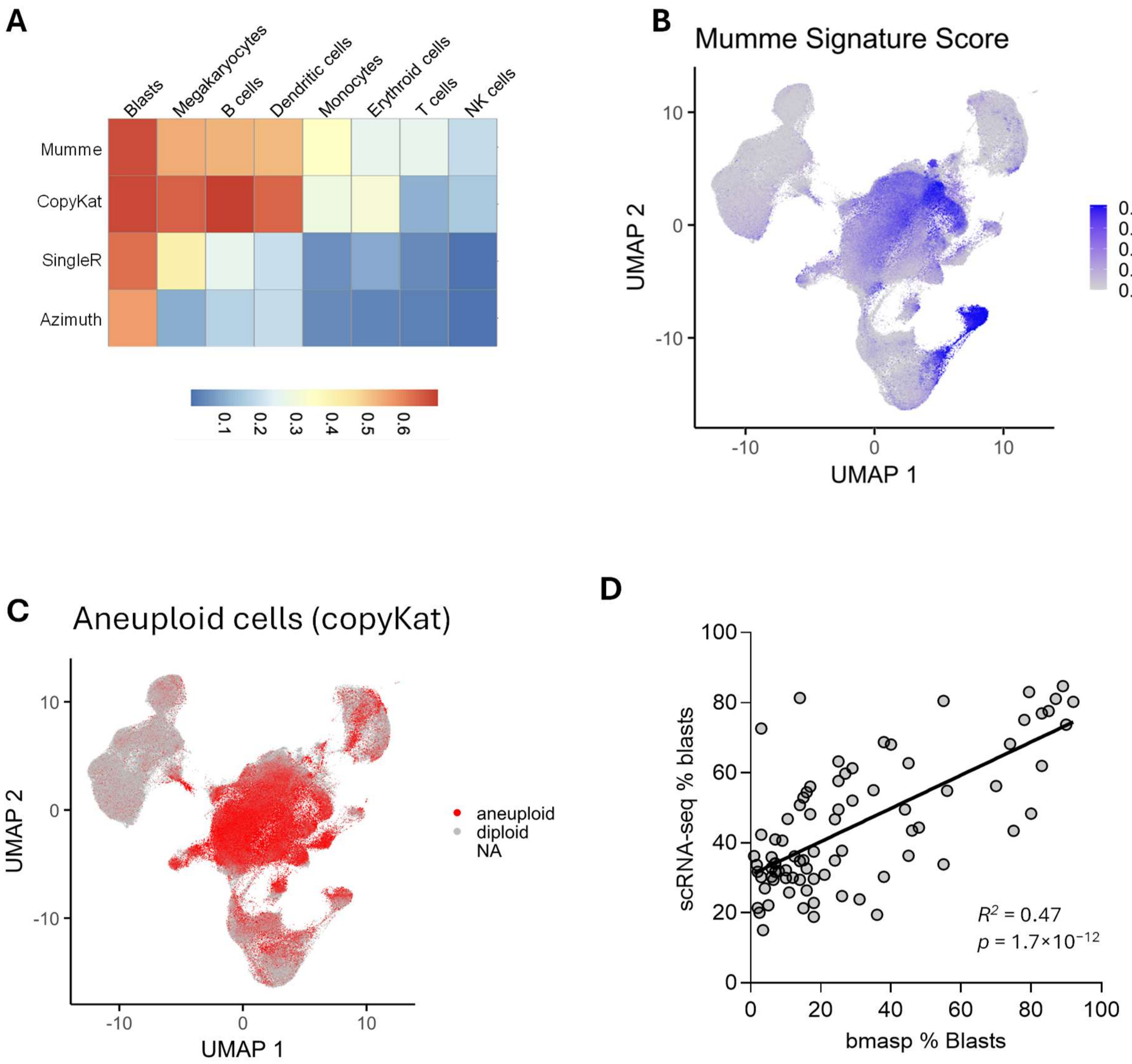

**Figure S8. Reanalysis of single cell genomics dataset of decitabine-treated patients.**
(**A**) UMAP plot showing distribution of cells for 14 patients after dataset integration.
(**B**) UMAP plot showing all 157,042 cells with their correspondingly labeled cell types.
(**C**) UMAP plot showing distribution of Ucell score from AML blast signature for each cell.
(**D**) UMAP plot showing distribution of cells labeled as "aneuploid" by CopyKAT algorithm.

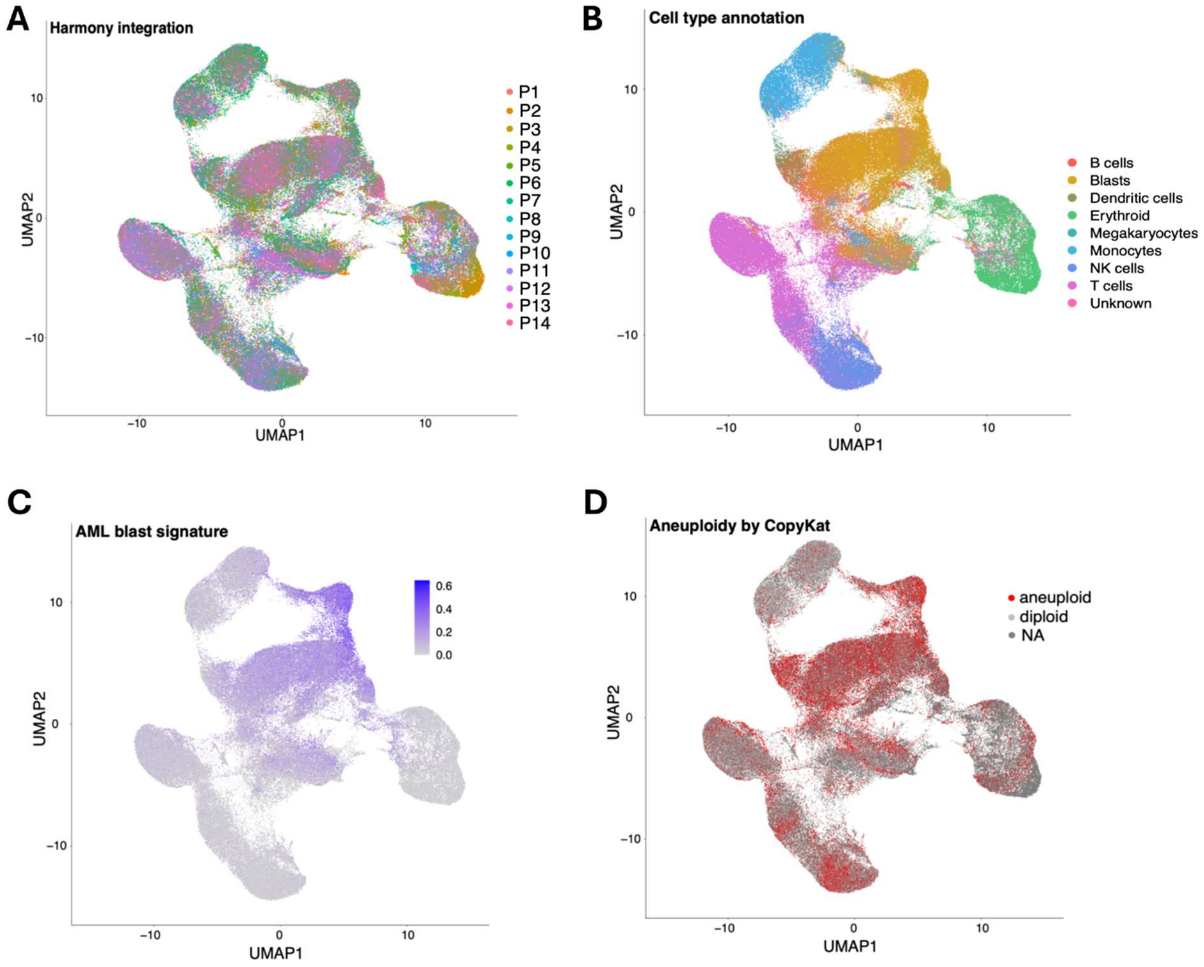

**Figure S9. Identification of a decitabine-regulated transcriptional signature.**
(**A**) Volcano plot of differential gene expression (DGE) comparing decitabine-treated vs untreated (at screening) patient samples. Decitabine (DAC)-induced genes (n=134) are labeled in red, and decitabine-repressed genes (n=10) are labeled in blue. Vertical dashed lines indicate the absolute $\log_2$ fold change cutoff of 1, while the horizontal dashed line indicates adjusted p-value cutoff of 0.05.
(**B**) Pathway analysis of DGE for (A). Top upregulated and downregulated pathways are labeled in red and blue, respectively.
(**C**) Gene set expression analysis of decitabine-modulated gene signature (decitabine-induced genes in upper panels and decitabine-repressed genes in lower panels) applied to DGE in monotherapy, combination therapy, and responder patients.

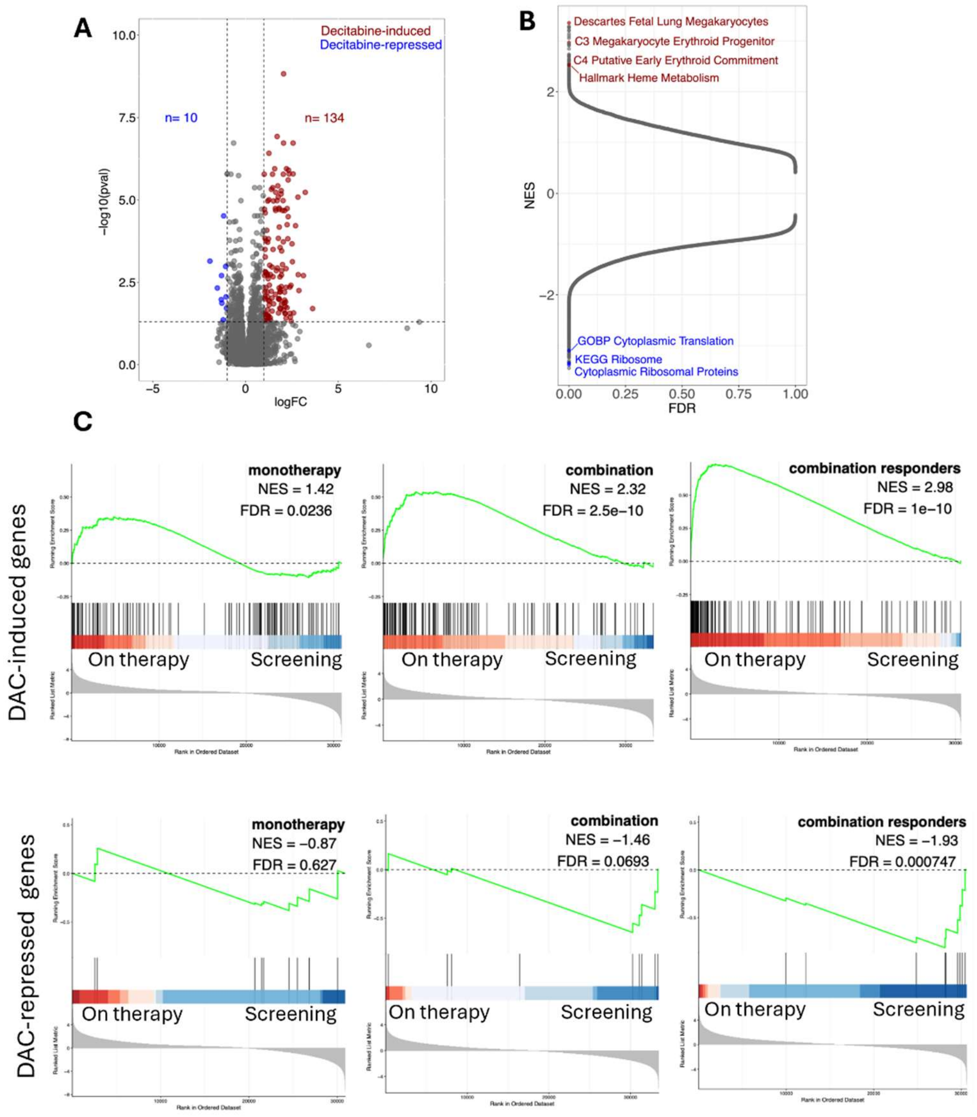

**Figure S10. Validation of Numbat tumor cell annotation.**
(**A**) Confusion matrix showing agreement between cytogenetic alterations identified by Numbat and clinical cytogenetic results.
(**B**) Heatmap of percentage-point changes from screening to treatment (early time point, C1D15 or C2D1) in individual tumor clones that underwent overall statistically significant change, within each cell type on a per-patient basis. Color indicates percentage-point abundance change, size indicates statistical significance.

**A**

| | Cytogenetics – no alteration | Cytogenetics – altered |
|---|---|---|
| Numbat – no alteration | 944 | 50 |
| Numbat – altered | 192 | 90 |

Fisher's exact test $p=1 \times 10^{-30}$

**B**

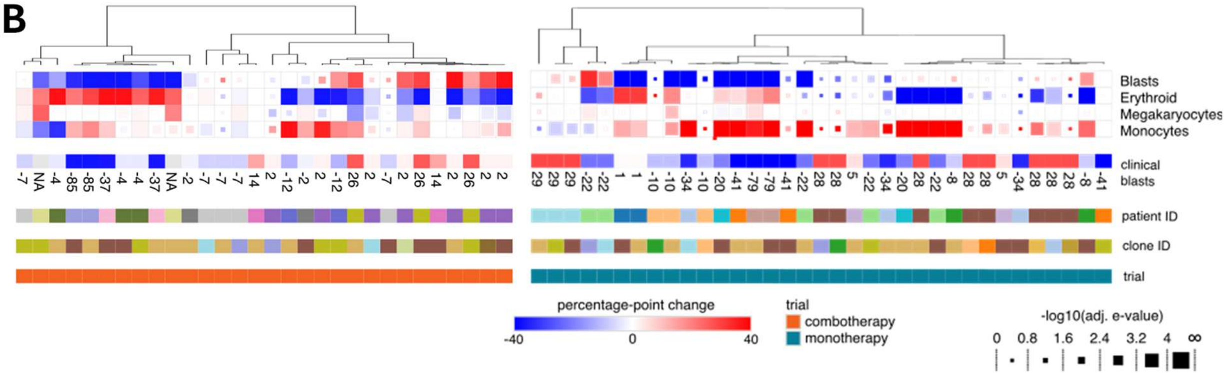

**Figure S11. Responders display transcriptional similarity to *CEBPA*-mutant AML patients, additional datasets.**
(**A-B**) Heatmaps of pairwise Pearson correlation (corr) coefficients between transcriptional profiles of FHD286+DAC responders and TARGET-AML patients (n=2613) (A) or TCGA-LAML patients (n=149) (B), represented individually in rows and columns. One sample represents a composite profile generated for all FHD286+DAC responders (n=4). The *CEBPA*-mutant AML subcluster is highlighted with a dashed outlined and further examined in a zoomed-in heatmap at the right. Position of responders is highlighted in zoomed-in heatmap. Color represents correlation strength.
(**C-D**) Corresponding box plots showing distribution of pairwise Pearson correlation coefficients from (A-B). Each point represents Pearson correlation coefficient for comparison between combination responders and TARGET-AML patients (C) or TCGA-LAML (D). Patients with *CEBPA*-mutant AML are represented in orange and all other AML subtypes are represented in gray. Statistical significance between two groups was assessed using a Wilcoxon rank test and is represented by asterisk (*), corresponding to Wilcoxon p-value thresholds. **** is $P \leq 0.0001$, *** is $P \leq 0.001$. Boxes represent interquartile range (IQR) with median indicated by the center line, and whiskers extending to 1.5x IQR.

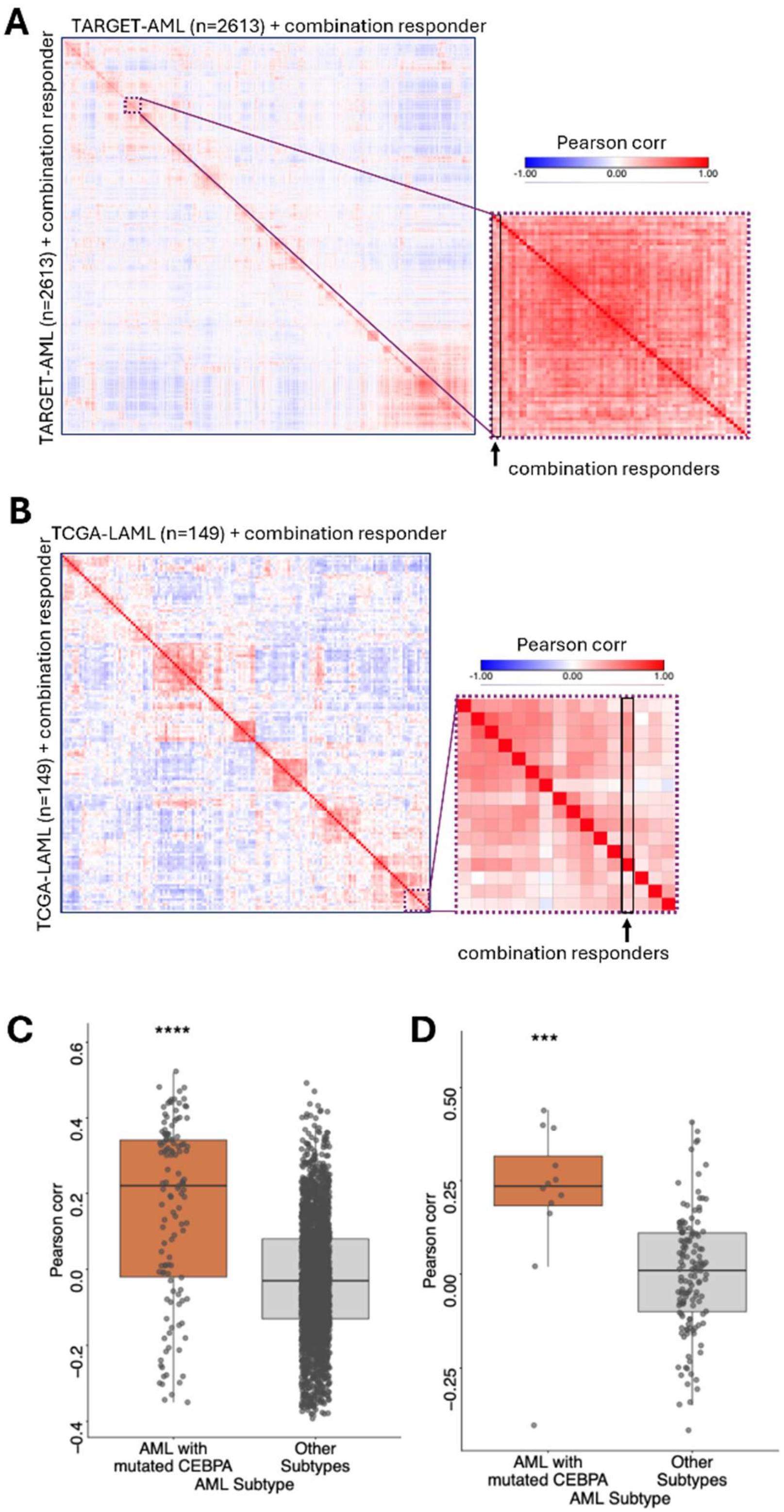
A
TARGET-AML (n=2613) + combination responder
TARGET-AML (n=2613) + combination responder
Pearson corr
-1.00
0.00
1.00
combination responders
B
TCGA-LAML (n=149) + combination responder
TCGA-LAML (n=149) + combination responder
Pearson corr
-1.00
0.00
1.00
combination responders
C
****
Pearson corr
0.6
0.4
0.2
0.0
-0.2
-0.4
AML with mutated CEBPA
Other Subtypes
AML Subtype
D
***
Pearson corr
0.50
0.25
0.00
-0.25
AML with mutated CEBPA
Other Subtypes
AML Subtype

**Table S1. Baseline Demographics and Disease Characteristics of Patients in the Combination Portion of Study FHD-286-C-002**

| Parameter | FHD-286 dose level (mg QD) (in combination with decitabine 20 mg/m² QD Days 1-5) | | | | | | | | | | Total (N=47) |
|---|---|---|---|---|---|---|---|---|---|---|---|
| | Group B1 | | | | | | Group B2 | | | | |
| | 2.5 (N=7) | 5 (N=6) | 7.5 (N=6) | 7.5/5 (N=3) | 7.5/10 (N=6) | Total (N=28) | 1.5 (N=4) | 1.5/2.5 (N=4) | 2.5 (N=11) | Total (N=19) | |
| **Sex, n (%)** | | | | | | | | | | | |
| Male | 3 (42.9) | 3 (50.0) | 4 (66.7) | 1 (33.3) | 4 (66.7) | 15 (53.6) | 2 (50.0) | 2 (50.0) | 5 (45.5) | 9 (47.4) | 24 (51.1) |
| Female | 4 (57.1) | 3 (50.0) | 2 (33.3) | 2 (66.7) | 2 (33.3) | 13 (46.4) | 2 (50.0) | 2 (50.0) | 6 (54.5) | 10 (52.6) | 23 (48.9) |
| **Age (years), median (min, max)** | 78 (27, 84) | 51.5 (23, 77) | 68.5 (32, 80) | 56 (30, 70) | 68.5 (66, 74) | 66.5 (23, 84) | 40.5 (19, 71) | 63.5 (41, 81) | 67 (35, 81) | 65 (19, 81) | 66 (19, 84) |
| **Cytogenetics at screening, n (%)** | | | | | | | | | | | |
| Abnormal karyotype | 5 (71.4) | 5 (83.3) | 4 (66.7) | 2 (66.7) | 5 (83.3) | 21 (75.0) | 4 (100) | 3 (75.0) | 11 (100) | 18 (94.7) | 39 (83.0) |
| Normal karyotype | 0 | 0 | 2 (33.3) | 0 | 0 | 2 (7.1) | 0 | 0 | 0 | 0 | 2 (4.3) |
| Missing | 2 (28.6) | 1 (16.7) | 0 | 1 (33.3) | 1 (16.7) | 5 (17.9) | 0 | 1 (25.0) | 0 | 1 (5.3) | 6 (12.8) |
| **Risk stratification by genetics at screening, n (%)** | | | | | | | | | | | |
| Favorable | 0 | 1 (16.7) | 0 | 1 (33.3) | 0 | 2 (7.1) | 0 | 0 | 0 | 0 | 2 (4.3) |
| Intermediate | 0 | 0 | 2 (33.3) | 1 (33.3) | 0 | 3 (10.7) | 0 | 0 | 4 (36.4) | 4 (21.1) | 7 (14.9) |
| Adverse | 5 (71.4) | 4 (66.7) | 4 (66.7) | 0 | 5 (83.3) | 18 (64.3) | 4 (100) | 3 (75.0) | 7 (63.6) | 14 (73.7) | 32 (68.1) |
| Unknown | 0 | 0 | 0 | 0 | 0 | 0 | 0 | 0 | 0 | 0 | 0 |
| Missing | 2 (28.6) | 1 (16.7) | 0 | 1 (33.3) | 1 (16.7) | 5 (17.9) | 0 | 1 (25.0) | 0 | 1 (5.3) | 6 (12.8) |
| **Time since initial cancer diagnosis[a] (years), median (min, max)** | 0.950 (0.320, 2.497) | 1.459 (0.920, 3.625) | 0.958 (0.361, 2.628) | 2.541 (0.038, 3.493) | 0.600 (0.156, 1.342) | 1.010 (0.038, 3.625) | 1.031 (0.564, 1.572) | 0.658 (0.044, 1.763) | 1.232 (0.309, 2.806) | 1.032 (0.044, 2.806) | 1.032 (0.038, 3.625) |

**Table S1. Baseline Demographics and Disease Characteristics of Patients in the Combination Portion of Study FHD-286-C-002**

| | | | | | | | | | | | |
|---|---|---|---|---|---|---|---|---|---|---|---|
| **Type of cancer, n (%)** | | | | | | | | | | | |
| AML | 6 (85.7) | 6 (100) | 4 (66.7) | 3 (100) | 5 (83.3) | 24 (85.7) | 4 (100) | 4 (100) | 10 (90.9) | 18 (94.7) | 42 (89.4) |
| MDS | 1 (14.3) | 0 | 1 (16.7) | 0 | 1 (16.7) | 3 (10.7) | 0 | 0 | 1 (9.1) | 1 (5.3) | 4 (8.5) |
| CMML | 0 | 0 | 1 (16.7) | 0 | 0 | 1 (3.6) | 0 | 0 | 0 | 0 | 1 (2.1) |
| **Type of AML, n (%)** | | | | | | | | | | | |
| De novo | 2 (28.6) | 5 (83.3) | 4 (66.7) | 2 (66.7) | 5 (83.3) | 18 (64.3) | 2 (50.0) | 3 (75.0) | 5 (45.5) | 10 (52.6) | 28 (59.6) |
| Secondary AML | 4 (57.1) | 1 (16.7) | 0 | 1 (33.3) | 0 | 6 (21.4) | 2 (50.0) | 1 (25.0) | 5 (45.5) | 8 (42.1) | 14 (29.8) |

Abbreviations: AML, acute myeloid leukemia; CMML, chronic myelomonocytic leukemia; max, maximum; MDS, myelodysplastic syndromes; min, minimum; QD, once daily.
Note: Group B1: Subjects *not* receiving a triazole antifungal agent classified as a strong CYP3A inhibitor at baseline. Group B2: Subjects receiving a triazole antifungal agent classified as a strong CYP3A inhibitor at baseline.
[a] Time since initial cancer diagnosis is calculated as (date of first dose of study treatment−date of initial diagnosis+1)/365.25.

**Table S2: Representativeness of the Study Population**

| | |
|---|---|
| Cancer type(s)/subtype(s)/stage(s)/condition | Relapsed or refractory (R/R) acute myeloid leukemia (AML)<br>R/R myelodysplastic syndrome (MDS)<br>R/R chronic myelomonocytic leukemia (CMML)<br>Because only 8 patients with R/R MDS and 1 patient with R/R CMML were treated, no specific statements on representativeness can be made for these indications. |
| **Considerations related to:** | |
| Sex | AML is diagnosed slightly more frequently in males than females. Based on data from the US SEER database from 2019 through 2022, the age-adjusted incidence of AML was 5.2 per 100,000 men and 3.7 per 100,000 women (*54*). |
| Age | For AML, based on data from the US SEER database from 2019 through 2023, the median age at diagnosis is approximately 70 years (*54*). |
| Race/ethnicity | AML is diagnosed more frequently among non-Hispanic White individuals (*54*). Based on data from the US SEER database from 2017 through 2021, the age-adjusted incidence of AML was 5.4 (males)/3.6 (females), 4.5/3.4, 4.2/3.4, 4.2/3.0, and 4.0/3.1 per 100,000 non-Hispanic White, non-Hispanic American Indian/Alaska Native, non-Hispanic Black, non-Hispanic Asian/Pacific Islander, and Hispanic individuals, respectively (*55*). A single-center retrospective analysis in Baltimore, Maryland, US, found that of 548 patients diagnosed with AML from 2000 through 2009, 73% were White, 19% were Black, 4% were Hispanic, and 4% were Asian (*56*). |
| Geography | This was a US-based study, and as such it is not representative of the global geography of AML. Published data on the geographic distribution of AML within the United States are limited. |
| Other considerations | None |
| Overall representativeness of the study | Overall, the study was generally representative of the population of patients with AML in the US.<br>The study enrolled nearly equal percentages of men (50.6%) and women (49.4%).<br>The median age of participants in the study was 66 years.<br>78.2% of participants identified as White, and 83.9% identified as “not Hispanic or Latino.” |

Abbreviations: SEER, Surveillance, Epidemiology, and End Results Program.

**Table S3: Baseline Demographics, Disease Characteristics, and Cancer-Related Medical History for Patients in the Monotherapy and Combination Therapy Portions of Study FHD-286-C-002**

| Parameter | Monotherapy (n=40) | Combination therapy (n=47) | Total (n=87) |
|---|---|---|---|
| **Age, median (min, max)** | 65.5 (25, 84) | 66.0 (19, 84) | 66.0 (19, 84) |
| **Risk stratification by genetics at screening, n (%)** | | | |
| Favorable | 1 (2.5) | 2 (4.3) | 3 (3.4) |
| Intermediate | 3 (7.5) | 7 (14.9) | 10 (11.5) |
| Adverse | 34 (85.0) | 31 (66.0) | 65 (74.7) |
| Unknown | 1 (2.5) | 0 | 1 (1.1) |
| Missing | 1 (2.5) | 7 (14.9) | 8 (9.2) |
| **Hematologic parameters at screening, median (min, max)** | | | |
| Leukocytes ($\times 10^9$/L) | 2.1 (0.2, 21.9) | 2.3 (0, 17.9) | 2.2 (0, 21.9) |
| Absolute neutrophil count ($\times 10^9$/L) | 0.4 (0, 4.15)<br>n=39 | 0.32 (0, 4.13)<br>n=46 | 0.39 (0,4.15)<br>n=85 |
| Platelets ($\times 10^9$/L) | 21.5 (2, 1328) | 25 (4, 323) | 24 (2, 1328) |
| Hemoglobin (g/L) | 83.5 (58, 112) | 78 (66, 116) | 80 (58, 116) |
| **Transfusion dependent[a], n (%)** | 31 (77.5) | 42 (91.3)<br>n=46[b] | 73 (84.9)<br>n=86 |
| **Prior anticancer therapy** | | | |
| Number of prior lines of systemic anticancer therapy for AML/MDS/other AHD, median (min, max) | 3.0 (1, 7) | 3.0 (1, 9) | 3.0 (1,9) |
| **Prior HMAs, n (%)** | | | |
| Any prior HMA | 37 (92.5) | 45 (95.7) | 82 (94.3) |
| Prior azacitidine | 26 (65.0) | 30 (63.8) | 56 (64.4) |
| Prior decitabine | 17 (42.5) | 21 (44.7) | 38 (43.7) |

| | | | |
|---|---|---|---|
| Prior cedazuridine/decitabine | 3 (7.5) | 6 (12.8) | 9 (10.3) |
| Prior venetoclax-based regimen, n (%) | 36 (90.0) | 45 (95.7) | 81 (93.1) |
| Cytarabine-based regimens, n (%) | 24 (60.0) | 32 (68.1) | 56 (64.4) |
| Prior HSCT, n (%) | 12 (30.0) | 13 (27.7) | 25 (28.7) |

Abbreviations: AHD, antecedent hematologic disorder; AML, acute myeloid leukemia; CMML, chronic myelomonocytic leukemia; HSCT, hematopoietic stem cell transplant; max, maximum; MDS, myelodysplastic syndromes; min, minimum; QD, once daily.

[a] AML: Receipt of any red blood cell (RBC) or platelet transfusions within at least 28 days before the start of study treatment. MDS: Either low or high transfusion burden. Low transfusion burden: 3-7 RBC transfusions within 16 weeks, in ≥2 transfusion episodes. High transfusion burden: ≥8 RBC transfusions within 16 weeks or ≥4 RBC transfusions within 8 weeks, in ≥2 transfusion episodes.

[b] Transfusion dependence was not available for the 1 patient with chronic myelomonocytic leukemia.

**Table S4: Cancer-Related Medical History for Patients in the Combination Portion of Study FHD-286-C-002**

| Parameter | FHD-286 dose level (mg QD) (in combination with decitabine 20 mg/m² QD Days 1-5) | | | | | | | | | | Total (N=47) |
|---|---|---|---|---|---|---|---|---|---|---|---|
| | Group B1 | | | | | | Group B2 | | | | |
| | 2.5 (N=7) | 5 (N=6) | 7.5 (N=6) | 7.5/5 (N=3) | 7.5/10 (N=6) | Total (N=28) | 1.5 (N=4) | 1.5/2.5 (N=4) | 2.5 (N=11) | Total (N=19) | |
| **Prior anticancer therapy, n (%)** | | | | | | | | | | | |
| Yes | 7 (100) | 6 (100) | 6 (100) | 3 (100) | 6 (100) | 28 (100) | 4 (100) | 4 (100) | 11 (100) | 19 (100) | 47 (100) |
| Prior systemic anticancer therapy for AML, MDS, or other AHD | 7 (100) | 6 (100) | 6 (100) | 3 (100) | 6 (100) | 28 (100) | 4 (100) | 4 (100) | 11 (100) | 19 (100) | 47 (100) |
| Prior HMAs | 7 (100) | 6 (100) | 6 (100) | 3 (100) | 5 (83.3) | 27 (96.4) | 4 (100) | 4 (100) | 10 (90.9) | 18 (94.7) | 45 (95.7) |
| Prior azacitidine | 5 (71.4) | 4 (66.7) | 4 (66.7) | 2 (66.7) | 1 (16.7) | 16 (57.1) | 4 (100) | 2 (50.0) | 8 (72.7) | 14 (73.7) | 30 (63.8) |
| Prior decitabine | 2 (28.6) | 3 (50.0) | 2 (33.3) | 1 (33.3) | 4 (66.7) | 12 (42.9) | 3 (75.0) | 2 (50.0) | 4 (36.4) | 9 (47.4) | 21 (44.7) |
| Prior cedazuridine/ decitabine | 2 (28.6) | 0 | 1 (16.7) | 0 | 0 | 3 (10.7) | 0 | 1 (25.0) | 2 (18.2) | 3 (15.8) | 6 (12.8) |
| Prior venetoclax-based regimen | 7 (100) | 6 (100) | 5 (83.3) | 3 (100) | 6 (100) | 27 (96.4) | 4 (100) | 4 (100) | 10 (90.9) | 18 (94.7) | 45 (95.7) |
| Prior cytarabine-based regimens | 3 (42.9) | 5 (83.3) | 4 (66.7) | 2 (66.7) | 4 (66.7) | 18 (64.3) | 4 (100) | 2 (50.0) | 8 (72.7) | 14 (73.7) | 32 (68.1) |
| Prior HSCT[a] | 2 (28.6) | 3 (50.0) | 1 (16.7) | 3 (100) | 0 | 9 (32.1) | 1 (25.0) | 0 | 3 (27.3) | 4 (21.1) | 13 (27.7) |
| Prior menin inhibitor | 0 | 0 | 1 (16.7) | 1 (33.3) | 0 | 2 (7.1) | 0 | 0 | 0 | 0 | 2 (4.3) |
| Prior CAR T-cell therapy | 0 | 0 | 0 | 0 | 0 | 0 | 0 | 0 | 1 (9.1) | 1 (5.3) | 1 (2.1) |
| Prior NK cell therapy | 0 | 0 | 0 | 0 | 0 | 0 | 1 (25.0) | 0 | 0 | 1 (5.3) | 1 (2.1) |

**Table S4: Cancer-Related Medical History for Patients in the Combination Portion of Study FHD-286-C-002**

| Parameter | FHD-286 dose level (mg QD) (in combination with decitabine 20 mg/m$^2$ QD Days 1-5) | | | | | | | | | | Total (N=47) |
|---|---|---|---|---|---|---|---|---|---|---|---|
| | Group B1 | | | | | | Group B2 | | | | |
| | 2.5 (N=7) | 5 (N=6) | 7.5 (N=6) | 7.5/5 (N=3) | 7.5/10 (N=6) | Total (N=28) | 1.5 (N=4) | 1.5/2.5 (N=4) | 2.5 (N=11) | Total (N=19) | |
| **Prior anticancer therapy, n (%)** | | | | | | | | | | | |
| Radiotherapy | 2 (28.6) | 0 | 0 | 0 | 0 | 2 (7.1) | 0 | 0 | 0 | 0 | 2 (4.3) |
| Surgery | 0 | 0 | 1 (16.7) | 0 | 0 | 1 (3.6) | 0 | 0 | 0 | 0 | 1 (2.1) |
| Prior systemic anticancer therapy for other prior cancer(s) | 4 (57.1) | 1 (16.7) | 0 | 0 | 1 (16.7) | 6 (21.4) | 3 (75.0) | 0 | 2 (18.2) | 5 (26.3) | 11 (23.4) |
| **Number of prior lines of systemic anticancer therapy for AML/MDS/other AHD** | | | | | | | | | | | |
| 1, n (%) | 2 (28.6) | 1 (16.7) | 2 (33.3) | 0 | 3 (50.0) | 8 (28.6) | 0 | 2 (50.0) | 1 (9.1) | 3 (15.8) | 11 (23.4) |
| 2, n (%) | 2 (28.6) | 1 (16.7) | 3 (50.0) | 0 | 0 | 6 (21.4) | 0 | 0 | 3 (27.3) | 3 (15.8) | 9 (19.1) |
| ≥3, n (%) | 3 (42.9) | 4 (66.7) | 1 (16.7) | 3 (100) | 3 (50.0) | 14 (50.0) | 4 (100) | 2 (50.0) | 7 (63.6) | 13 (68.4) | 27 (57.4) |
| Median (min, max) | 2 (1, 5) | 3.5 (1, 4) | 2 (1, 5) | 4 (3, 7) | 2 (1, 4) | 2.5 (1, 7) | 5 (3, 6) | 2 (1, 6) | 3 (1, 9) | 3 (1, 9) | 3 (1, 9) |

Abbreviations: AHD=antecedent hematologic disorder; AML=acute myeloid leukemia; max=maximum; CAR=chimeric antigen receptor; MDS=myelodysplastic syndromes; min=minimum; NK=natural killer; HMA=hypomethylating agent; HSCT=hematopoietic stem cell transplant; QD=once daily.
Note: Group B1: Subjects *not* receiving a triazole antifungal agent classified as a strong CYP3A inhibitor at baseline. Group B2: Subjects receiving a triazole antifungal agent classified as a strong CYP3A inhibitor at baseline.
[a] All patients in this category had had 1 prior HSCT for AML, MDS, or another AHD.

**Table S5: Summary of TEAEs Reported in ≥20% of Patients Overall, Treatment-Related TEAEs Reported in ≥10% of Patients Overall, and Actions Taken in Response to a TEAE in the Combination Portion of Study FHD-286-C-002**

| TEAE | FHD-286 dose level (mg QD) (in combination with decitabine 20 mg/m$^2$ QD Days 1-5), n (%) | | | | | | | | | | Total (N=47) n (%) |
|---|---|---|---|---|---|---|---|---|---|---|---|
| | Group B1 | | | | | | Group B2 | | | | |
| | 2.5 (N=7) | 5 (N=6) | 7.5 (N=6) | 7.5/5 (N=3) | 7.5/10 (N=6) | Total (N=28) | 1.5 (N=4) | 1.5/2.5 (N=4) | 2.5 (N=11) | Total (N=19) | |
| **Any TEAE** | **7 (100)** | **6 (100)** | **6 (100)** | **3 (100)** | **6 (100)** | **28 (100)** | **4 (100)** | **4 (100)** | **11 (100)** | **19 (100)** | **47 (100)** |
| **Any treatment-related TEAE** | **6 (85.7)** | **3 (50.0)** | **5 (83.3)** | **1 (33.3)** | **3 (50.0)** | **18 (64.3)** | **2 (50.0)** | **3 (75.0)** | **8 (72.7)** | **13 (68.4)** | **31 (66.0)** |
| Hypokalemia | 2 (28.6) | 4 (66.7) | 3 (50.0) | 2 (66.7) | 3 (50.0) | 14 (50.0) | 3 (75.0) | 2 (50.0) | 6 (54.5) | 11 (57.9) | 25 (53.2) |
| Fatigue | 6 (85.7) | 3 (50.0) | 2 (33.3) | 0 | 1 (16.7) | 12 (42.9) | 2 (50.0) | 2 (50.0) | 5 (45.5) | 9 (47.4) | 21 (44.7) |
| Treatment-related fatigue | 2 (28.6) | 2 (33.3) | 0 | 0 | 1 (16.7) | 5 (17.9) | 0 | 0 | 0 | 0 | 5 (10.6) |
| Decreased WBC count[a] | 3 (42.9) | 4 (66.7) | 3 (50.0) | 1 (33.3) | 3 (50.0) | 14 (50.0) | 2 (50.0) | 1 (25.0) | 3 (27.3) | 6 (31.6) | 20 (42.6) |
| Treatment-related decreased WBC count | 2 (28.6) | 1 (16.7) | 1 (16.7) | 0 | 0 | 4 (14.3) | 0 | 0 | 1 (9.1) | 1 (5.3) | 5 (10.6) |
| Dyspnea | 3 (42.9) | 1 (16.7) | 4 (66.7) | 0 | 3 (50.0) | 11 (39.3) | 3 (75.0) | 2 (50.0) | 4 (36.4) | 9 (47.4) | 20 (42.6) |
| Nausea/vomiting[b] | 5 (71.4) | 3 (50.0) | 3 (50.0) | 1 (33.3) | 2 (33.3) | 14 (50.0) | 1 (25.0) | 2 (50.0) | 3 (27.3) | 6 (31.6) | 20 (42.6) |
| Treatment-related nausea/vomiting | 4 (57.1) | 1 (16.7) | 2 (33.3) | 0 | 1 (16.7) | 8 (28.6) | 0 | 1 (25.0) | 0 | 1 (5.3) | 9 (19.1) |
| Cough | 2 (28.6) | 0 | 3 (50.0) | 2 (66.7) | 3 (50.0) | 10 (35.7) | 3 (75.0) | 1 (25.0) | 4 (36.4) | 8 (42.1) | 18 (38.3) |
| Decreased neutrophil count[c] | 4 (57.1) | 3 (50.0) | 3 (50.0) | 1 (33.3) | 3 (50.0) | 14 (50.0) | 1 (25.0) | 1 (25.0) | 1 (9.1) | 3 (15.8) | 17 (36.2) |
| Treatment-related decreased neutrophil count | 3 (42.9) | 2 (33.3) | 2 (33.3) | 0 | 0 | 7 (25.0) | 0 | 0 | 0 | 0 | 7 (14.9) |
| Diarrhea | 1 (14.3) | 2 (33.3) | 3 (50.0) | 0 | 4 (66.7) | 10 (35.7) | 4 (100) | 1 (25.0) | 2 (18.2) | 7 (36.8) | 17 (36.2) |

**Table S5: Summary of TEAEs Reported in ≥20% of Patients Overall, Treatment-Related TEAEs Reported in ≥10% of Patients Overall, and Actions Taken in Response to a TEAE in the Combination Portion of Study FHD-286-C-002**

| TEAE | FHD-286 dose level (mg QD) (in combination with decitabine 20 mg/m$^2$ QD Days 1-5), n (%) | | | | | | | | | | Total (N=47) n (%) |
|---|---|---|---|---|---|---|---|---|---|---|---|
| | Group B1 | | | | | | Group B2 | | | | |
| | 2.5 (N=7) | 5 (N=6) | 7.5 (N=6) | 7.5/5 (N=3) | 7.5/10 (N=6) | Total (N=28) | 1.5 (N=4) | 1.5/2.5 (N=4) | 2.5 (N=11) | Total (N=19) | |
| Increased blood glucose[d] | 2 (28.6) | 1 (16.7) | 4 (66.7) | 0 | 3 (50.0) | 10 (35.7) | 1 (25.0) | 3 (75.0) | 3 (27.3) | 7 (36.8) | 17 (36.2) |
| Febrile neutropenia | 2 (28.6) | 2 (33.3) | 2 (33.3) | 2 (66.7) | 2 (33.3) | 10 (35.7) | 2 (50.0) | 3 (75.0) | 1 (9.1) | 6 (31.6) | 16 (34.0) |
| Increased blood bilirubin[e] | 2 (28.6) | 1 (16.7) | 3 (50.0) | 1 (33.3) | 3 (50.0) | 10 (35.7) | 0 | 2 (50.0) | 4 (36.4) | 6 (31.6) | 16 (34.0) |
| Treatment-related increased blood bilirubin | 0 | 0 | 2 (33.3) | 0 | 1 (16.7) | 3 (10.7) | 0 | 0 | 2 (18.2) | 2 (10.5) | 5 (10.6) |
| Pneumonia | 3 (42.9) | 2 (33.3) | 4 (66.7) | 1 (33.3) | 1 (16.7) | 11 (39.3) | 1 (25.0) | 0 | 4 (36.4) | 5 (26.3) | 16 (34.0) |
| Stomatitis | 1 (14.3) | 2 (33.3) | 4 (66.7) | 1 (33.3) | 0 | 8 (28.6) | 0 | 2 (50.0) | 6 (54.5) | 8 (42.1) | 16 (34.0) |
| Treatment-related stomatitis | 0 | 1 (16.7) | 4 (66.7) | 0 | 0 | 5 (17.9) | 0 | 0 | 2 (18.2) | 2 (10.5) | 7 (14.9) |
| Anemia | 5 (71.4) | 2 (33.3) | 2 (33.3) | 1 (33.3) | 2 (33.3) | 12 (42.9) | 0 | 0 | 3 (27.3) | 3 (15.8) | 15 (31.9) |
| Constipation | 3 (42.9) | 2 (33.3) | 1 (16.7) | 0 | 2 (33.3) | 8 (28.6) | 0 | 3 (75.0) | 3 (27.3) | 6 (31.6) | 14 (29.8) |
| Decreased appetite | 1 (14.3) | 1 (16.7) | 1 (16.7) | 0 | 3 (50.0) | 6 (21.4) | 1 (25.0) | 3 (75.0) | 4 (36.4) | 8 (42.1) | 14 (29.8) |
| Treatment-related decreased appetite | 1 (14.3) | 1 (16.7) | 0 | 0 | 1 (16.7) | 3 (10.7) | 0 | 0 | 2 (18.2) | 2 (10.5) | 5 (10.6) |
| Decreased platelet count[f] | 5 (71.4) | 3 (50.0) | 1 (16.7) | 0 | 3 (50.0) | 12 (42.9) | 0 | 0 | 2 (18.2) | 2 (10.5) | 14 (29.8) |
| Muscular weakness | 3 (42.9) | 1 (16.7) | 3 (50.0) | 1 (33.3) | 1 (16.7) | 9 (32.1) | 1 (25.0) | 1 (25.0) | 3 (27.3) | 5 (26.3) | 14 (29.8) |
| Rash[g] | 0 | 3 (50.0) | 4 (66.7) | 1 (33.3) | 1 (16.7) | 9 (32.1) | 0 | 2 (50.0) | 3 (27.3) | 5 (26.3) | 14 (29.8) |

**Table S5: Summary of TEAEs Reported in ≥20% of Patients Overall, Treatment-Related TEAEs Reported in ≥10% of Patients Overall, and Actions Taken in Response to a TEAE in the Combination Portion of Study FHD-286-C-002**

| TEAE | FHD-286 dose level (mg QD) (in combination with decitabine 20 mg/m$^2$ QD Days 1-5), n (%) | | | | | | | | | | Total (N=47) n (%) |
|---|---|---|---|---|---|---|---|---|---|---|---|
| | Group B1 | | | | | | Group B2 | | | | |
| | 2.5 (N=7) | 5 (N=6) | 7.5 (N=6) | 7.5/5 (N=3) | 7.5/10 (N=6) | Total (N=28) | 1.5 (N=4) | 1.5/2.5 (N=4) | 2.5 (N=11) | Total (N=19) | |
| Treatment-related rash | 0 | 1 (16.7) | 4 (66.7) | 0 | 1 (16.7) | 6 (21.4) | 0 | 1 (25.0) | 2 (18.2) | 3 (15.8) | 9 (19.1) |
| Increased AST | 0 | 2 (33.3) | 1 (16.7) | 2 (66.7) | 1 (16.7) | 6 (21.4) | 2 (50.0) | 2 (50.0) | 3 (27.3) | 7 (36.8) | 13 (27.7) |
| Epistaxis | 1 (14.3) | 2 (33.3) | 2 (33.3) | 2 (66.7) | 1 (16.7) | 8 (28.6) | 2 (50.0) | 0 | 3 (27.3) | 5 (26.3) | 13 (27.7) |
| Pyrexia | 4 (57.1) | 2 (33.3) | 1 (16.7) | 0 | 2 (33.3) | 9 (32.1) | 2 (50.0) | 0 | 2 (18.2) | 4 (21.1) | 13 (27.7) |
| Dizziness | 3 (42.9) | 1 (16.7) | 2 (33.3) | 1 (33.3) | 1 (16.7) | 8 (28.6) | 1 (25.0) | 0 | 3 (27.3) | 4 (21.1) | 12 (25.5) |
| Hyperkalemia | 1 (14.3) | 1 (16.7) | 4 (66.7) | 1 (33.3) | 3 (50.0) | 10 (35.7) | 0 | 1 (25.0) | 1 (9.1) | 2 (10.5) | 12 (25.5) |
| Hypotension | 3 (42.9) | 2 (33.3) | 2 (33.3) | 0 | 1 (16.7) | 8 (28.6) | 0 | 1 (25.0) | 3 (27.3) | 4 (21.1) | 12 (25.5) |
| Increased cardiac troponin[h] | 2 (28.6) | 0 | 2 (33.3) | 1 (33.3) | 2 (33.3) | 7 (25.0) | 0 | 3 (75.0) | 2 (18.2) | 5 (26.3) | 12 (25.5) |
| Treatment-related increased cardiac troponin | 2 (28.6) | 0 | 1 (16.7) | 0 | 1 (16.7) | 4 (14.3) | 0 | 2 (50.0) | 1 (9.1) | 3 (15.8) | 7 (14.9) |
| Peripheral edema | 1 (14.3) | 2 (33.3) | 2 (33.3) | 0 | 1 (16.7) | 6 (21.4) | 0 | 1 (25.0) | 4 (36.4) | 5 (26.3) | 11 (23.4) |
| Oropharyngeal pain | 3 (42.9) | 2 (33.3) | 1 (16.7) | 2 (66.7) | 2 (33.3) | 10 (35.7) | 1 (25.0) | 0 | 0 | 1 (5.3) | 11 (23.4) |
| Increased ALP | 2 (28.6) | 1 (16.7) | 0 | 0 | 2 (33.3) | 5 (17.9) | 1 (25.0) | 1 (25.0) | 3 (27.3) | 5 (26.3) | 10 (21.3) |
| Confusional state | 1 (14.3) | 2 (33.3) | 3 (50.0) | 0 | 1 (16.7) | 7 (25.0) | 1 (25.0) | 1 (25.0) | 1 (9.1) | 3 (15.8) | 10 (21.3) |
| Hyperuricemia | 1 (14.3) | 2 (33.3) | 4 (66.7) | 1 (33.3) | 1 (16.7) | 9 (32.1) | 0 | 0 | 1 (9.1) | 1 (5.3) | 10 (21.3) |
| Hypophosphatemia | 1 (14.3) | 0 | 3 (50.0) | 1 (33.3) | 2 (33.3) | 7 (25.0) | 1 (25.0) | 1 (25.0) | 1 (9.1) | 3 (15.8) | 10 (21.3) |
| Hypoxia | 0 | 0 | 2 (33.3) | 1 (33.3) | 1 (16.7) | 4 (14.3) | 1 (25.0) | 2 (50.0) | 3 (27.3) | 6 (31.6) | 10 (21.3) |

**Table S5: Summary of TEAEs Reported in ≥20% of Patients Overall, Treatment-Related TEAEs Reported in ≥10% of Patients Overall, and Actions Taken in Response to a TEAE in the Combination Portion of Study FHD-286-C-002**

| | FHD-286 dose level (mg QD) (in combination with decitabine 20 mg/m$^2$ QD Days 1-5), n (%) | | | | | | | | | | |
|---|---|---|---|---|---|---|---|---|---|---|---|
| | **Group B1** | | | | | | **Group B2** | | | | **Total (N=47) n (%)** |
| **TEAE** | **2.5 (N=7)** | **5 (N=6)** | **7.5 (N=6)** | **7.5/5 (N=3)** | **7.5/10 (N=6)** | **Total (N=28)** | **1.5 (N=4)** | **1.5/2.5 (N=4)** | **2.5 (N=11)** | **Total (N=19)** | |
| Treatment-related differentiation syndrome | 1 (14.3) | 0 | 1 (16.7) | 0 | 2 (33.3) | 4 (14.3) | 2 (50.0) | 1 (25.0) | 1 (9.1) | 4 (21.1) | 8 (17.0) |
| Treatment-related dysgeusia | 1 (14.3) | 2 (33.3) | 0 | 0 | 0 | 3 (10.7) | 0 | 0 | 2 (18.2) | 2 (10.5) | 5 (10.6) |

Abbreviations: ALP, alkaline phosphatase; AST, aspartate aminotransferase; QD, once daily; TEAE, treatment-emergent adverse event; WBC, white blood cell.
Note: Group B1: Patients *not* receiving a triazole antifungal agent classified as a strong CYP3A inhibitor at baseline. Group B2: Patients receiving a triazole antifungal agent classified as a strong CYP3A inhibitor at baseline. Percentages are based on the number of subjects in each dose group/total.
A TEAE is any adverse event either reported for the first time or that represents a worsening of a preexisting event after the first dose of study treatment and within 28 days of the last dose of study treatment.
A patient is counted only once for multiple events of the same name.
[a] Decreased WBC count includes the leukopenia and decreased WBC count.
[b] Nausea/vomiting includes the nausea, vomiting, and retching.
[c] Decreased neutrophil count includes neutropenia and decreased neutrophil count.
[d] Increased blood glucose includes hyperglycemia and increased blood glucose.
[e] Increased blood bilirubin includes increased blood bilirubin and hyperbilirubinemia.
[f] Decreased platelet count includes thrombocytopenia and decreased platelet count.
[g] Rash includes acute febrile neutrophilic dermatosis; drug eruption; skin erosion and exfoliation; rash in general and exfoliative, macular, erythematous, generalized, maculopapular, papular, pruritic, pustular, vesicular, and butterfly rash; allergic and bullous dermatitis; and urticaria.
[h] Increased cardiac troponin includes increased troponin, troponin I, and troponin T.

**Table S6: Summary of SAEs Reported in ≥5% of Patients Overall and Treatment-Related SAEs Reported in ≥1 Patient Overall in the Combination Portion of Study FHD-286-C-002**

| SAE | FHD-286 dose level (mg QD) (in combination with decitabine 20 mg/m$^2$ QD Days 1-5), n (%) | | | | | | | | | | Total (N=47) n (%) |
|---|---|---|---|---|---|---|---|---|---|---|---|
| | **Group B1** | | | | | | **Group B2** | | | | |
| | **2.5 (N=7)** | **5 (N=6)** | **7.5 (N=6)** | **7.5/5 (N=3)** | **7.5/10 (N=6)** | **Total (N=28)** | **1.5 (N=4)** | **1.5/2.5 (N=4)** | **2.5 (N=11)** | **Total (N=19)** | |
| **Any SAE** | **7 (100)** | **5 (83.3)** | **6 (100)** | **3 (100)** | **6 (100)** | **27 (96.4)** | **4 (100)** | **4 (100)** | **10 (90.9)** | **18 (94.7)** | **45 (95.7)** |
| **Any treatment-related SAE** | **3 (42.9)** | **2 (33.3)** | **4 (66.7)** | **0** | **2 (33.3)** | **11 (39.3)** | **2 (50.0)** | **3 (75.0)** | **5 (45.5)** | **10 (52.6)** | **21 (44.7)** |
| Febrile neutropenia | 2 (28.6) | 2 (33.3) | 1 (16.7) | 2 (66.7) | 1 (16.7) | 8 (28.6) | 2 (50.0) | 2 (50.0) | 1 (9.1) | 5 (26.3) | 13 (27.7) |
| Pneumonia | 3 (42.9) | 2 (33.3) | 2 (33.3) | 1 (33.3) | 1 (16.7) | 9 (32.1) | 0 | 0 | 4 (36.4) | 4 (21.1) | 13 (27.7) |
| DS | 1 (14.3) | 0 | 1 (16.7) | 0 | 2 (33.3) | 4 (14.3) | 2 (50.0) | 1 (25.0) | 2 (18.2) | 5 (26.3) | 9 (19.1) |
| Treatment-related DS | 1 (14.3) | 0 | 1 (16.7) | 0 | 2 (33.3) | 4 (14.3) | 2 (50.0) | 1 (25.0) | 1 (9.1) | 4 (21.1) | 8 (17.0) |
| Acute kidney injury | 1 (14.3) | 1 (16.7) | 1 (16.7) | 0 | 0 | 3 (10.7) | 0 | 1 (25.0) | 2 (18.2) | 3 (15.8) | 6 (12.8) |
| Increased cardiac troponin[a] | 1 (14.3) | 0 | 2 (33.3) | 0 | 0 | 3 (10.7) | 0 | 1 (25.0) | 1 (9.1) | 2 (10.5) | 5 (10.6) |
| Treatment-related increased cardiac troponin | 1 (14.3) | 0 | 1 (16.7) | 0 | 0 | 2 (7.1) | 0 | 1 (25.0) | 1 (9.1) | 2 (10.5) | 4 (8.5) |
| Rash[b] | 0 | 0 | 2 (33.3) | 0 | 0 | 2 (7.1) | 0 | 1 (25.0) | 2 (18.2) | 3 (15.8) | 5 (10.6) |

**Table S6: Summary of SAEs Reported in ≥5% of Patients Overall and Treatment-Related SAEs Reported in ≥1 Patient Overall in the Combination Portion of Study FHD-286-C-002**

| SAE | FHD-286 dose level (mg QD) (in combination with decitabine 20 mg/m$^2$ QD Days 1-5), n (%) | | | | | | | | | | Total (N=47) n (%) |
|---|---|---|---|---|---|---|---|---|---|---|---|
| | Group B1 | | | | | | Group B2 | | | | |
| | 2.5 (N=7) | 5 (N=6) | 7.5 (N=6) | 7.5/5 (N=3) | 7.5/10 (N=6) | Total (N=28) | 1.5 (N=4) | 1.5/2.5 (N=4) | 2.5 (N=11) | Total (N=19) | |
| Treatment-related rash | 0 | 0 | 2 (33.3) | 0 | 0 | 2 (7.1) | 0 | 1 (25.0) | 2 (18.2) | 3 (15.8) | 5 (10.6) |
| Sepsis | 0 | 0 | 3 (50.0) | 0 | 1 (16.7) | 4 (14.3) | 0 | 1 (25.0) | 0 | 1 (5.3) | 5 (10.6) |
| Treatment-related sepsis | 0 | 0 | 1 (16.7) | 0 | 0 | 1 (3.6) | 0 | 0 | 0 | 0 | 1 (2.1) |
| Anemia | 2 (28.6) | 0 | 0 | 0 | 0 | 2 (7.1) | 0 | 0 | 1 (9.1) | 1 (5.3) | 3 (6.4) |
| Atrial fibrillation | 0 | 0 | 0 | 0 | 1 (16.7) | 1 (3.6) | 0 | 1 (25.0) | 1 (9.1) | 2 (10.5) | 3 (6.4) |
| Increased blood bilirubin[c] | 0 | 0 | 1 (16.7) | 0 | 1 (16.7) | 2 (7.1) | 0 | 0 | 1 (9.1) | 1 (5.3) | 3 (6.4) |
| Treatment-related increased blood bilirubin | 0 | 0 | 1 (16.7) | 0 | 1 (16.7) | 2 (7.1) | 0 | 0 | 1 (9.1) | 1 (5.3) | 3 (6.4) |
| Peripheral edema | 0 | 1 (16.7) | 0 | 0 | 0 | 1 (3.6) | 0 | 1 (25.0) | 1 (9.1) | 2 (10.5) | 3 (6.4) |
| Sinus bradycardia | 0 | 1 (16.7) | 1 (16.7) | 0 | 0 | 2 (7.1) | 0 | 0 | 1 (9.1) | 1 (5.3) | 3 (6.4) |
| Treatment-related sinus bradycardia | 0 | 0 | 1 (16.7) | 0 | 0 | 1 (3.6) | 0 | 0 | 1 (9.1) | 1 (5.3) | 2 (4.3) |
| Treatment-related increased ALT | 0 | 0 | 0 | 0 | 1 (16.7) | 1 (3.6) | 0 | 0 | 0 | 0 | 1 (2.1) |

**Table S6: Summary of SAEs Reported in ≥5% of Patients Overall and Treatment-Related SAEs Reported in ≥1 Patient Overall in the Combination Portion of Study FHD-286-C-002**

| SAE | FHD-286 dose level (mg QD) (in combination with decitabine 20 mg/m$^2$ QD Days 1-5), n (%) | | | | | | | | | | Total (N=47) n (%) |
|---|---|---|---|---|---|---|---|---|---|---|---|
| | Group B1 | | | | | | Group B2 | | | | |
| | 2.5 (N=7) | 5 (N=6) | 7.5 (N=6) | 7.5/5 (N=3) | 7.5/10 (N=6) | Total (N=28) | 1.5 (N=4) | 1.5/2.5 (N=4) | 2.5 (N=11) | Total (N=19) | |
| Treatment-related increased AST | 0 | 0 | 0 | 0 | 1 (16.7) | 1 (3.6) | 0 | 0 | 0 | 0 | 1 (2.1) |
| Treatment-related bradycardia | 0 | 0 | 0 | 0 | 0 | 0 | 0 | 1 (25.0) | 0 | 1 (5.3) | 1 (2.1) |
| Treatment-related cholangitis | 0 | 0 | 0 | 0 | 0 | 0 | 0 | 0 | 1 (9.1) | 1 (5.3) | 1 (2.1) |
| Treatment-related conduction disorder | 0 | 0 | 0 | 0 | 0 | 0 | 0 | 1 (25.0) | 0 | 1 (5.3) | 1 (2.1) |
| Treatment-related decreased platelet count[d] | 1 (14.3) | 0 | 0 | 0 | 0 | 1 (3.6) | 0 | 0 | 0 | 0 | 1 (2.1) |
| Treatment-related diarrhea | 0 | 0 | 0 | 0 | 1 (16.7) | 1 (3.6) | 0 | 0 | 0 | 0 | 1 (2.1) |
| Treatment-related dry mouth | 0 | 0 | 0 | 0 | 0 | 0 | 0 | 0 | 1 (9.1) | 1 (5.3) | 1 (2.1) |
| Treatment-related epistaxis | 0 | 1 (16.7) | 0 | 0 | 0 | 1 (3.6) | 0 | 0 | 0 | 0 | 1 (2.1) |
| Treatment-related gastric hemorrhage | 1 (14.3) | 0 | 0 | 0 | 0 | 1 (3.6) | 0 | 0 | 0 | 0 | 1 (2.1) |

**Table S6: Summary of SAEs Reported in ≥5% of Patients Overall and Treatment-Related SAEs Reported in ≥1 Patient Overall in the Combination Portion of Study FHD-286-C-002**

| SAE | FHD-286 dose level (mg QD) (in combination with decitabine 20 mg/m$^2$ QD Days 1-5), n (%) | | | | | | | | | | Total (N=47) n (%) |
|---|---|---|---|---|---|---|---|---|---|---|---|
| | Group B1 | | | | | | Group B2 | | | | |
| | 2.5 (N=7) | 5 (N=6) | 7.5 (N=6) | 7.5/5 (N=3) | 7.5/10 (N=6) | Total (N=28) | 1.5 (N=4) | 1.5/2.5 (N=4) | 2.5 (N=11) | Total (N=19) | |
| Treatment-related increased blood glucose[e] | 0 | 0 | 0 | 0 | 0 | 0 | 0 | 0 | 1 (9.1) | 1 (5.3) | 1 (2.1) |
| Treatment-related nausea/vomiting[f] | 0 | 0 | 1 (16.7) | 0 | 0 | 1 (3.6) | 0 | 0 | 0 | 0 | 1 (2.1) |
| Treatment-related pancytopenia | 0 | 1 (16.7) | 0 | 0 | 0 | 1 (3.6) | 0 | 0 | 0 | 0 | 1 (2.1) |
| Treatment-related presyncope | 0 | 0 | 0 | 0 | 0 | 0 | 0 | 1 (25.0) | 0 | 1 (5.3) | 1 (2.1) |
| Treatment-related streptococcal bacteraemia | 0 | 1 (16.7) | 0 | 0 | 0 | 1 (3.6) | 0 | 0 | 0 | 0 | 1 (2.1) |

Abbreviations: ALT, alanine aminotransferase; AST, alanine aminotransferase; DS, differentiation syndrome; QD, once daily; SAE, serious treatment-emergent adverse event.
Note: Group B1: Subjects *not* receiving a triazole antifungal agent classified as a strong CYP3A inhibitor at baseline. Group B2: Subjects receiving a triazole antifungal agent classified as a strong CYP3A inhibitor at baseline.
Percentages are based on the number of patients within each dose group.
A TEAE is any adverse event either reported for the first time or worsening of a preexisting event after the first dose of study treatment and within 28 days of the last administration of study treatment.
A patient is counted only once for multiple events of the same name.
[a] Increased cardiac troponin includes increased troponin, troponin I, and troponin T.
[b] Rash includes acute febrile neutrophilic dermatosis; drug eruption; skin erosion and exfoliation; rash in general and exfoliative, macular, erythematous, generalized, maculopapular, papular, pruritic, pustular, vesicular, and butterfly rash; allergic or bullous dermatitis; and urticaria.
[c] Increased blood bilirubin includes increased blood bilirubin and hyperbilirubinemia.
[d] Decreased platelet count includes thrombocytopenia and decreased platelet count.
[e] Increased blood glucose includes hyperglycemia and increased blood glucose.
[f] Nausea/vomiting includes nausea, vomiting, and retching.

**Table S7: Antibodies used for FACS**

| **Monotherapy panel** | | | | |
|---|---|---|---|---|
| **Marker** | **Clone** | **Fluorophore** | **Vendor** | **Catalog #** |
| CD45 | 2D1 | APCCy7 | BioLegend | 368516 |
| CD14 | M5E2 | FITC | BioLegend | 301804 |
| CD34 | 561 | PE | BioLegend | 343606 |
| CD11c | 3.9 | PE-Dazzle594 | BioLegend | 301642 |
| HLADR | L243 | PECy5 | BioLegend | 307608 |
| CD15 | W6D3 | PerCPCy5.5 | BioLegend | 323020 |
| CD33 | WM53 | PECy7 | BioLegend | 303434 |
| CD38 | HIT2 | APC | BioLegend | 305310 |
| CD64 | 10.1 | AlexaFluor700 | BioLegend | 305040 |
| CD117 | 104D2 | BV510 | BioLegend | 313220 |
| CD13 | WM15 | BV605 | BioLegend | 301728 |
| CD11b | ICRF44 | BV650 | BioLegend | 301336 |
| **Combination surface panel** | | | | |
| **Marker** | **Clone** | **Fluorophore** | **Vendor** | **Catalog #** |
| CD45 | HI30 | BV570 | BioLegend | 304034 |
| CD117 | 104D2 | BV605 | BioLegend | 313218 |
| CD11b | ICRF44 | BV650 | BioLegend | 301336 |
| CD123 | 6H6 | PECy5 | BioLegend | 306008 |
| CD15 | W6D3 | PerCPCy5.5 | BioLegend | 323020 |
| CD34 | 561 | PECy7 | BioLegend | 343616 |
| CD64 | 10.1 | AlexaFluor700 | BioLegend | 305040 |
| CD44 | BJ18 | APCFire750 | BioLegend | 338818 |
| **Combination intracellular panel** | | | | |
| **Marker** | **Clone** | **Fluorophore** | **Vendor** | **Catalog #** |
| Cleaved Caspase 3 | C92-605 | V450 | BD Biosciences | 560627 |
| BRG1 | EPNCIR111A | AlexaFluor488 | Abcam | ab196314 |
| gH2A.X | N1-431 | PE | BD Biosciences | 562377 |
| BCL2 | Bcl-2/100 | PE-CF594 | BD Biosciences | 563601 |
| BRM | EPR23102-44 | APC | Abcam | ab305747 |
